# Effect of a spatial repellent on malaria incidence in Mali: a cluster-randomized, controlled trial

**DOI:** 10.1101/2025.08.01.25332474

**Authors:** Issaka Sagara, Alassane Dicko, Ismaila Thera, Mamadou Coulibaly, Daman Sylla, Momar Mbodji, Mamadou Diango Traore, Ghislain Ismael Nana, Suzanne Van Hulle, Ashley Hudson, Jared Hendrickson, Fang Liu, Nicole L. Achee, John P. Grieco, AEGIS Mali Working Group

## Abstract

**Background:** Spatial repellents (SR) are a novel vector control intervention class that release volatile chemicals that disperse in the air creating a non-conducive environment for mosquitoes leading to movement away from a structure, inhibition of host attraction and/or feeding inhibition thereby reducing risk of human-vector contact and aiding in the prevention of pathogen transmission. SRs are under evaluation as a complementary strategy for malaria vector control. The AEGIS consortium conducted two cluster-randomized trials, one in Mali, reported here and one in Kenya(1), reported elsewhere.

**Methods:** The study was a double blinded, parallel-group, cluster-randomized controlled trial with 30 clusters per treatment arm (SR intervention and placebo), a 6-month baseline, and a post cluster randomization follow-up period of 24□months with intervention. The trial was conducted in Kolondieba District, Sikasso Region, Mali, a location that has some of the highest malaria transmission rates in the country, with widespread vector resistance. Two seasonal malaria chemoprevention campaigns and one mass distribution of ITNs were conducted during the trial. The main study objective was to demonstrate and quantify protective efficacy (PE) of SRs in reducing first-time malaria infection, as measured by microscopy, in a cohort of children aged 6 months to <10 years. The rate of adverse events and severe adverse events was captured in subjects and residents of enrolled structures.

**Results:** A total of 1,907 participants were enrolled across 60 clusters. A total of 12,246 positive malaria infections were detected during a 24 months intervention period (Apr 2022-Mar 2024). Most infections were identified as *Plasmodium falciparum* (95%). The estimated PE of SRs against the first-time malaria infection relative to placebo was -3.10% (95% 2-sided CI -32.24%, 19.62%, p-value 0.810). Overall, the PE effect in younger children (under 5 years old) was larger numerically, but not statistically significant and results do not suggest statistically significant effects of SR by season (rainy vs dry) or by year (Year 2 vs Year 1) compared to placebo. No statistically significant effects of SRs, compared to placebo, were demonstrated on entomological endpoints. No reported AE/SAEs were deemed associated with the SR intervention, patterns were consistent with disease frequency expected in study clusters, and safety assessments by the DSMB supported no significant safety concerns.

**Conclusions:** Our trial in Mali did not demonstrate SR efficacy. We propose several factors to have contributed to this lack of demonstrative effect. However, results have enhanced the knowledge-base for the SR intervention class regarding efficacy, coverage, durability, and safety. There remains a gap in evidence for SR health impact against malaria which will be required before a public health policy for inclusion of SRs in vector control programs can be fully endorsed.

## Introduction

Despite the scale-up of vector control tools like insecticide treated nets (ITNs) and indoor residual spraying (IRS), combined with other malaria control efforts, malaria remains one of the primary causes of morbidity and mortality in sub-Saharan Africa(2). The World Health Organization (WHO) African Region accounted for roughly 94% of malaria cases and 95% of malaria deaths globally in 2023.(2) New tools, such as a spatial repellent (SR), are needed to complement existing strategies in addressing gaps in protection for prevention of malaria transmission and sustaining the drive to elimination.(3)

Products in the SR intervention class release volatile chemicals that disperse in the air creating a non-conducive environment for mosquitoes leading to movement away from a structure, inhibition of host attraction and/or feeding inhibition thereby reducing risk of human-vector contact and aiding in the prevention of pathogen transmission.(4)(5)(6) The inclusion of SR in malaria control programs is dependent on evidence of epidemiological impact (protective efficacy (PE)) from two clinical trials against a specific disease as assessed by the WHO Vector Control Advisory Group (VCAG) for an endorsement of a WHO SR public health policy recommendation.(7)

Previous studies have demonstrated the protective effect of SRs. In China, transfluthrin coils reduced up to 77% of overall *Plasmodium falciparum* malaria infections.(8) In Kenya, a passive transfluthrin emanator demonstrated 33.4% protective efficacy against first-time infections (FTI).(1) In Peru, a different transfluthrin passive emanator reduced *Aedes*-borne virus infections by 34.1%.(9) A fourth trial in Indonesia demonstrated a reduction in malaria infection incidence (27.7%), although the difference was not statistically significant.(10) While the Kenya trial satisfied the evidence of epidemiological impact against malaria for one trial, evidence gaps remain.

As part of the Advancing Evidence for the Global Implementation of Spatial Repellents (AEGIS) consortium,(11) the Mali cRCT was designed to generate evidence on the PE and safety of a SR in reducing malaria infection incidence to assess the public health value of SR in a west African ecological setting.

## Materials and Methods

The study was conducted from July 2021 to March 2024 (Figure 1). The study was registered in clinicaltrials.gov (NCT04795648). Detailed descriptions of study design, methods, and statistical analyses have been published previously(11) and are reported following CONSORT guidelines (S1 - Consort Checklist).

**Figure 1.**
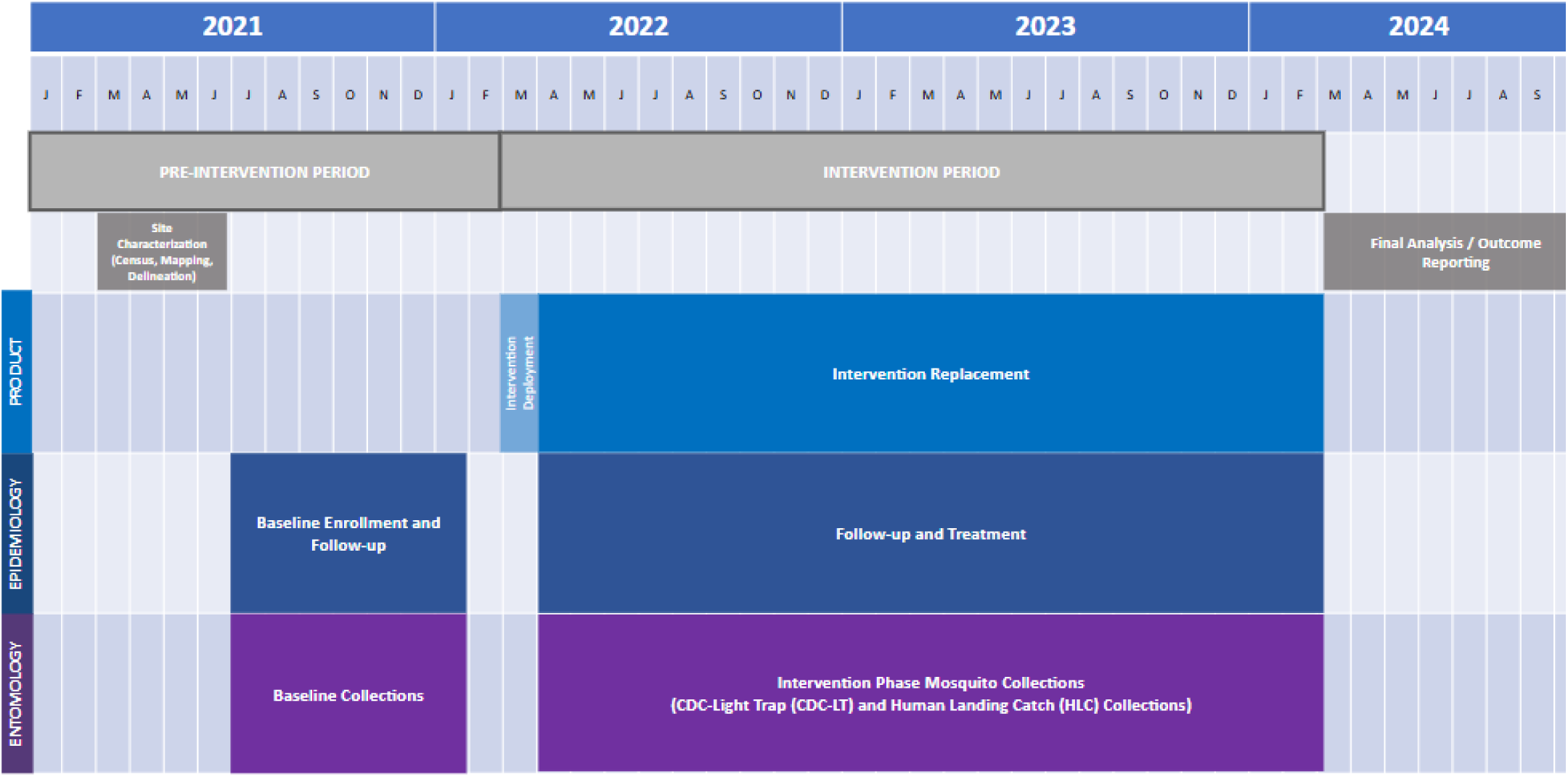
Study activity timeline

### Ethical statement

Ethical approval for the study protocol was granted by the Institutional Review Boards of the University of Sciences, Techniques and Technologies of Bamako, Mali (protocol #2020/252/CE/FMOS/FAPH), the University of Notre Dame (protocol #20-10-6245) and WHO Ethical Review Committee (0003449). Informed consent was obtained from parents or legal guardians of study participants for cohort participation, mosquito collectors conducting human landing catch (HLC) collections, and head of households for product placement.

### Study setting

The trial was conducted in Kolondieba District, Sikasso Region, Mali (S2. Supplemental Information, Figure 1) which has 161 villages. Based on a buffer distance of 5□km between villages to avoid overlap between clusters, 60 clusters were selected. There are a total of 17 health facilities (16 at the rural level and 1 in the peri-urban town of Kolondieba). Kolondieba has some of the highest malaria transmission rates in the country with 205 cases/1000 population in 2018 with one malaria transmission peak period from July to November (12). The main malaria vectors in this setting are *Anopheles gambiae, An. funestus,* and *An. coluzzii*(13).

Wide-spread vector resistance to pyrethroids, organochlorines, carbamates, and organophosphates has been documented, particularly, to pyrethroids in the *An. gambiae* complex which could be explained by an extensive use of insecticides for agricultural purposes.(14–16) The study area has mixed ethnicities including Bambara, Senoufo, and Fulani. Households are predominately traditional mud walls with grass or iron sheet roofing.

Two seasonal malaria chemoprevention (SMC; >3-59 months) campaigns, each with four cycles, were conducted during the trial: one July to October 2022 and another July to October 2023. A mass distribution of ITNs (Olyset-PBO) was conducted by Mali’s national malaria control program (NMCP) in June 2023.

### Trial design

The study was a double blinded, parallel-group, cluster-randomized, controlled trial (cRCT) with 30 clusters per treatment arm (SR intervention and placebo), with a 6-month baseline follow-up period, and a post cluster randomization follow-up period of 24□months with intervention. Baseline subject recruitment, screening, and enrollment commenced July 2021 (Figure 1). The 6-month baseline occurred between July 2021 and January 2022 (Figure 1).

Participants enrolled in baseline were re-enrolled for the intervention phase. Intervention deployment commenced March 2022, two weeks prior to parasite clearance. The intervention phase occurred between April 2022 and March 2024. Study staff distributed deltamethrin ITNs in July 2021 (2 ITNs per household, 1 for the participant, 1 for the parents). Clinical trial oversight was conducted throughout the trial (June and September 2022, March, June, and October 2023, March 2024).

The main study objective was to demonstrate and quantify the PE of SRs in reducing first-time malaria infection (defined in S3. Study Protocol), as measured by microscopy, in a cohort of children aged 6 months to <10 years. Secondary epidemiology endpoints were 1) overall new malaria infections (number of infections averted); and 2) first-time clinical malaria infections. Entomology endpoints were adult female anopheline: 1) blood-fed status (proxy for human-biting rates); 2) parity status (proxy for age-structure), 3) density, and 4) sporozoite positivity. Additional endpoints are outlined in clinicaltrials.gov (NCT04795648).

Sample size and power calculations for the primary endpoint are also published.(11) The study’s primary hypothesis is H0: SR does not have a protective effect against the first-time malaria infection compared to placebo vs. H1: SR has a protective effect against the first-time malaria infection compared to placebo. In brief, the number of clusters per treatment arm and the number of subjects per cluster were determined to detect a 30% reduction in first-time malaria infection (event) hazard rate by SR compared to placebo (primary endpoint). With assumptions of a baseline FTI hazard rate of 1.0 per person-year and a between-cluster coefficient of variation (CV) of 47%, a total of 788 events would be required to yield 80% power for establishing the primary hypothesis SRs reduce the first-time malaria hazard rate compared to placebo at a 5% one-sided Type-I error rate. To obtain 1056 events in the two-year study with a 35% loss to follow-up (LTFU) rate, each treatment during the intervention period had 30 clusters with 26 households (one subject per household) per cluster. The study was not powered to detect a statistically significant reduction by SRs in entomological endpoints as entomological endpoints are subject to high variability and detecting a meaningful reduction in the human landing rate (HLR) via HLC would have required too large of a volume of clusters and households per cluster to be accommodated within this study due to logistical and resource considerations.

### Participants

A total of 1907 participants were enrolled (Figure 2; 1920 sample size target) across 60 clusters. Enrollment was conducted by the study team. Participation included: 1) census, 2) malaria surveillance (bi-weekly blood sampling based on fever history), 3) monthly blood draws, 4) entomological surveys, and 5) intervention household placement. From compounds comprising multiple households (S2. Supplemental Information, Figure 2) in study clusters, one child, aged 6 months up to 10 years at time of recruitment, from a single household were provided the opportunity to enroll as subjects in the study. Additional eligibility requirements included having slept in the study clusters >90% of nights during any given month, no intention of extended travel exceeding 5 weeks outside the study cluster, maintaining hemoglobin levels > 7 g/dL, not participating in another clinical trial or being on regular malaria prophylaxis, and parents or guardians providing written consent.

**Figure 2.**
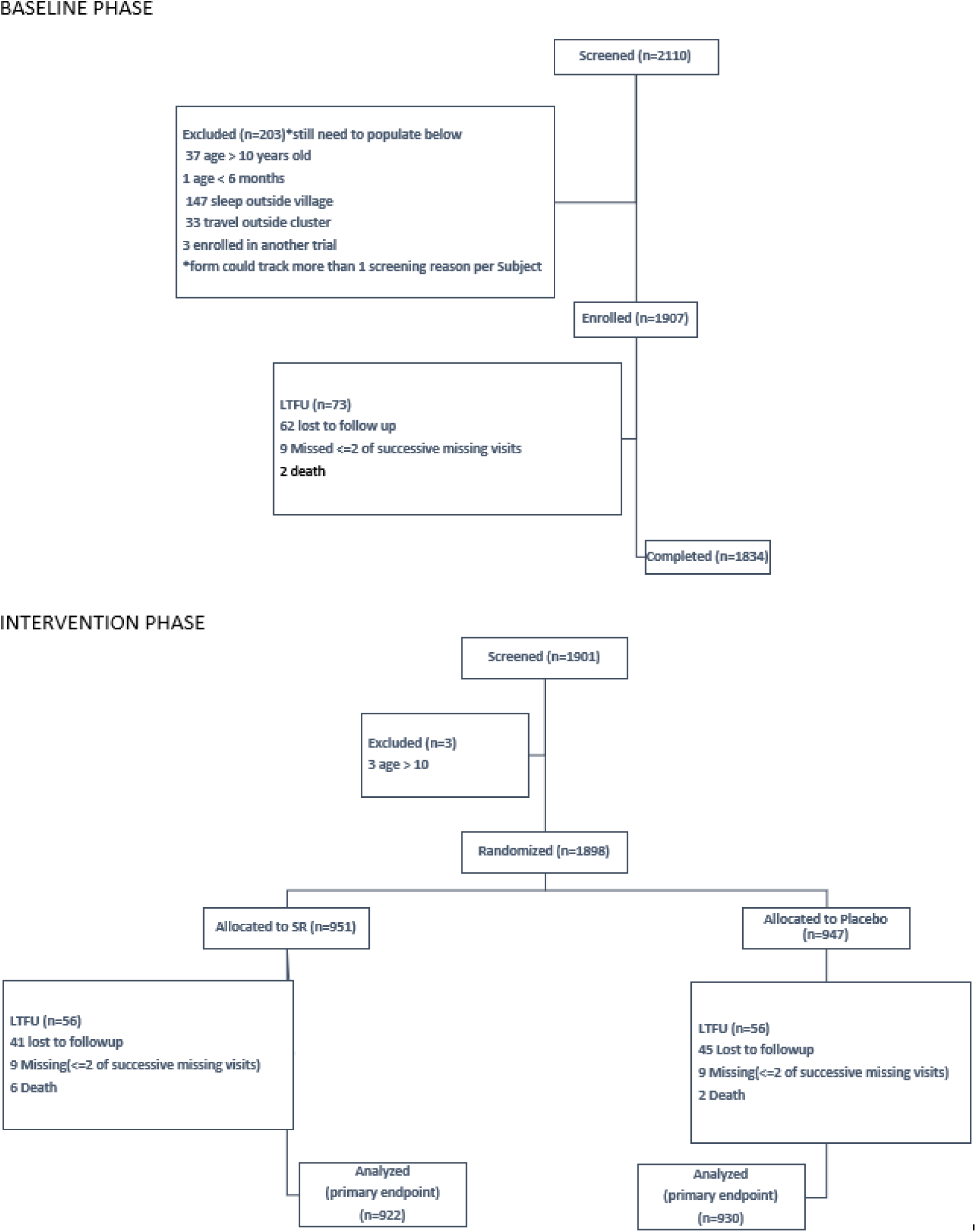
Trial Participant Flow Chart

### Intervention

The SR intervention was a transfluthrin-based passive emanator (SC Johnson SC Johnson Mosquito Shield ™) designed and produced by SC Johnson & Son, Inc (Racine, WI, USA), as described previously.(11)

Placebo and SR products had identical packaging and were deployed in all households of compounds where subjects resided by trained study personnel using a blinded coding scheme. Intervention placement (2 units/9 m^2^) and replacement schedule (28-day, with -2 to +14-day window) followed manufacturer specifications for indoor use conditions. During each product placement and replacement cycle, study personnel recorded placement and attrition (based on the number of products placed and removed from each structure) (S2. Supplemental Information, Figure 3).

**Figure 3.**
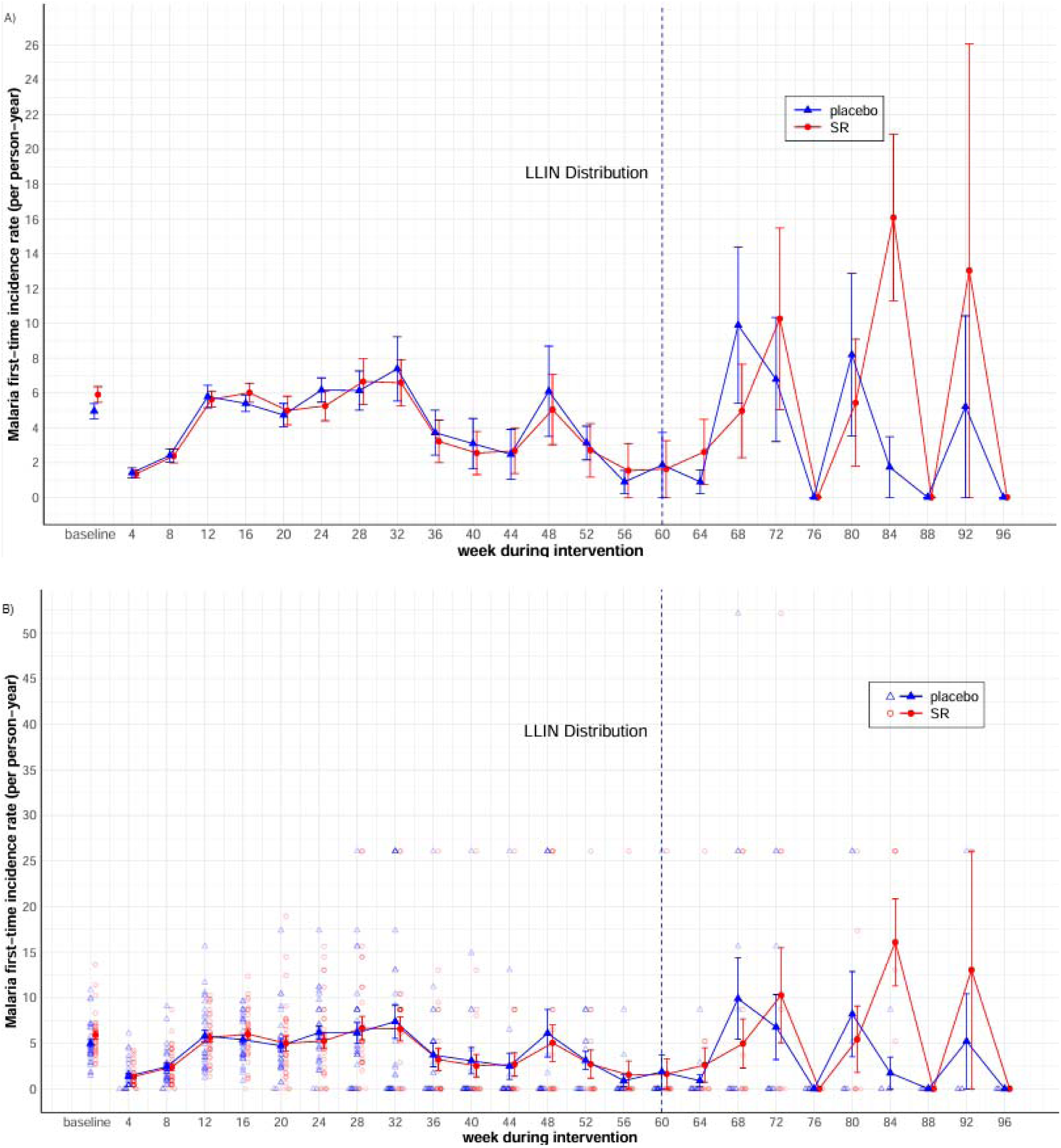
Malaria incidence rates. (A) Mean (+/- SE) incidence rate of the first-time malaria infection (primary endpoint). (B) The cluster-level incidence rates for the 30 SR and 30 placebo clusters.

On-site storage conditions of temperature and humidity were monitored by the study team and verified independently by SCJ and fhiClinical. Quality control analysis was performed in 2024 by Ross Laboratories, India on used (removed from study structures) and unused (in storage) emanators to verify the amount of transfluthrin in active emanators and the absence of transfluthrin in placebos. At the conclusion of the trial, used, unused, and expired emanators were disposed of according to manufacturer guidelines and Mali regulations on disposal requirements.

Compounds with an enrolled participant were eligible to receive product, while compounds without an enrolled participant were excluded from product placement (due to logistical and budgetary constraints) (S2. Supplemental Information, Figure 2). Standard-of-care for clinical management of malaria or vector control interventions (e.g., ITNs, IRS) was provided by study staff or was available through routine channels to participants in both treatment arms.

### Randomization

Cluster randomization and intervention allocation occurred February 2022. All randomization was conducted by an external statistician using the sample () function from R Studio 2022.02 (or R-3.6.3.). Investigators, staff, and study participants were blinded to cluster allocation. The Mali intervention administrator coordinated distribution of blinded active or placebo products to enrolled households within each cluster corresponding to the pre-labeled package code. A total of 60 clusters (average 197 resident households per cluster with approximately 142 enrolled households per cluster; ∼74% average cluster coverage) were randomized to ensure: 1) allocation of 60 clusters (30 SR, 30 placebo); 2) allocation of 20 clusters (10 SR, 10 placebo) for monitoring indoor adult mosquito density using Centers for Disease Control and Prevention (CDC) light traps (LTs); and 3) allocation of 12 clusters (6 SR, 6 placebo) for monitoring indoor and outdoor adult mosquito diversity, density, and HLR using HLC.

### Procedures

Prior to enrollment, all structures in the study area were mapped using GPS coordinates and a census was administered. Data from census and mapping were used for cluster delineation.

#### Epidemiology

Participants were randomly selected from the master list of households obtained during census and mapping to achieve the desired sample size. Following consent, a cohort of children aged 6□months to <□10□years of age were screened for inclusion and exclusion criteria and provided malaria parasite clearance therapy. The enrollment and baseline period was conducted over a 6-month period (Figure 1), during which time incidence of malaria infection was measured (rapid diagnostic tests and confirmation by microscopy). At the start of the intervention phase, participants were re-consented and cleared of malaria parasites, and households were enrolled for product placement. The first scheduled follow-up visit commenced two weeks after parasite clearance and continued for 2.5yrs. Follow-up visits included fever history and blood sampling for microscopy diagnosis (primary endpoint) at scheduled monthly, scheduled bi-weekly, and unscheduled sick visits. Participants who tested positive were treated with first-line artemisinin-based combination therapy (ACT). Malaria infection incidence within the cluster was used to assess SR efficacy on FTIs (primary endpoint), and overall new infections and first-time clinical infections (secondary endpoints).

#### Entomology

CDC-LT and HLC sampling were conducted to measure entomological endpoints (S2. Supplemental Information, Figure 4). For CDC-LT collections, ten randomly selected houses in each of 20 clusters (10 SR, 10 placebo) were sampled on a monthly basis (1-night collection; 6pm to 6am). For HLC collections, four randomly selected houses (total 48 houses across both arms) in each of 12 clusters (6 SR, 6 placebo) were sampled on a quarterly basis (2 nights collection; 6pm-6am). Houses sampled by HLC remained fixed throughout the intervention phase. Species of CDC-LT and HLC collected mosquitoes were identified using morphological keys(17) and confirmed by PCR.(18) Blood-fed status of anophelines collected by CDC-LT was measured by direct observation using a stereoscope. Samples were tested for the presence of sporozoite with ELISA assays.(19) Standard WHO and CDC insecticide susceptibility assays(20) were conducted with wild-type anopheline vectors (*Anopheles gambiae* s.l.) collected within 8 study clusters once during intervention (August 2022) and again post-intervention phase (November 2023) against deltamethrin and transfluthrin (S2. Supplemental Information, Figure 5).

#### Adverse event and serious adverse event monitoring

Participants were monitored for adverse events (AEs) or serious adverse events (SAEs) at each bi-weekly and monthly scheduled follow-up. Summary reports of both AEs and SAEs were submitted to a Data Safety Monitoring Board (DSMB) on a quarterly basis for safety assessment.

#### Data management and verification

Data collection was designed around a tablet-based platform, CommCare^TM^ (Dimagi Inc, MA, USA), linked to a custom-built database and web portal. The portal was constructed using Django+PostgreSQL for the back end and VueJS for the front end. Docker was used for containerization and deployment. Python scripts were developed for data cleaning and analysis. Data was cleaned according to the protocol and statistical analysis plan (SAP) to ensure data integrity. Data was cross-checked, identifying missing, incomplete, or suspect data submissions. Queries on study data were relayed to the study site data manager for resolution, after which data was verified, locked, and extracted for final study analyses.

### Statistical analyses

The detailed analytical approach is provided in the SAP (S4. Statistical Analysis Plan). Data analyses were conducted using SAS Studio 3.8.1 (Enterprise edition) and R Studio 2023.06.1+524 or higher.

#### Epidemiology

The primary hypothesis on PE of SRs against first-time malaria infection was tested by comparing the event hazard rates between SR and placebo in the intention-to-treat (ITT) population. The complementary-log-log (cloglog) regression was used to model interval-censored time to event data (collected every 2 weeks) and estimate the hazard ratio between SR and placebo with a proportional hazard assumption. Besides the treatment arm (SR vs placebo) and visit (biweekly), the cloglog model also contained individual-level, household-level, and cluster-level covariates as fixed effects (age, sex, number of doors, number of windows, open or closed eaves, roof type, baseline malaria incidence rate, cluster population size), and two independent zero-mean normally distributed random effects of cluster and of subject nested within cluster. The null hypothesis of PE is 0% against the alternative hypothesis PE is greater than 0% was tested by the lower one-sided Wald test.

The secondary hypothesis on PE against overall new malaria infections was tested by comparing hazard rates of the overall malaria infection in core areas between SR and placebo in the ITT population using a similar cloglog model as above. When determining which positive malaria infection in a subject was new or a carry-over case from a preceding malaria infection, we applied rules such as whether and when a subject with positive malaria diagnosis received anti-malarial treatment and whether two positive diagnoses were separated by a negative diagnosis, and factored in a 14-day protection (risk-free) period following anti-malarial treatment.

For both the primary and secondary hypotheses above, while pre-specified as one-sided, two-sided 95% CIs and p-values were provided, per the request of the reviewers, alongside the pre-specified one-sided 95% CIs and p-values.

In addition, we estimated SR PE against first-time and new infections by removing all fixed-effect baseline covariates from cloglog models. We also calculated the PE of SR against first-time and overall infections by age group (<6 years old vs >=6 years old at enrollment) via similar cloglog models.

In addition, a cloglog model similar to the primary first-time malaria infection endpoint was run to compare first-time clinical malaria infection hazard rates between SR and placebo in the ITT population, where clinical malaria case is defined as (axillary temperature ≥ 37.5°C OR history of fever within the last 48 hours and a positive microscopy read).

#### Entomology

Summary statistics were provided for all the pre-specified entomological endpoints, including CDC-LT anopheline density, anopheline HLR, sporozoite positivity rate, entomological inoculation rate (EIR), anopheline female parity rate, blood fed rate. Generalized Linear mixed models that accommodated different types of endpoints (e.g. count data for anopheline density and anopheline HLR, binary data for blood fed status) and the longitudinal nature of the collected data were also performed, as applicable, to examine the entomological effects of SR compared to placebo. For insecticide resistance characterization, anopheline mosquito larvae were reared to 3-5 day-old adults before exposure to deltamethrin or transfluthrin. Deltamethrin exposures were conducted using standard WHO tube assays(21), while CDC bottle assays(20) were utilized in testing transfluthrin. Evaluations were conducted mid-intervention phase (Aug 2022) and post-intervention (Nov 2023).

#### Safety

Total numbers of AEs and SAEs by symptom, frequency, and percentage of subjects that experienced at least one episode of AE or SAE, were summarized for both baseline and intervention periods and by treatment (SR, placebo), respectively.

## Results

### Study population

Subject retention rate was 94.1% in both the SR and placebo arms. (Figure 2). Baseline participant characteristics (primary endpoint) were balanced between SR and placebo at individual, household, cluster levels (Table 1). ITN usage, based on the question ‘How many bed nets were used for sleeping in the household last night’ was balanced between treatment arms as well (SR 93.0%, placebo 90.3%).

**Table 1.**
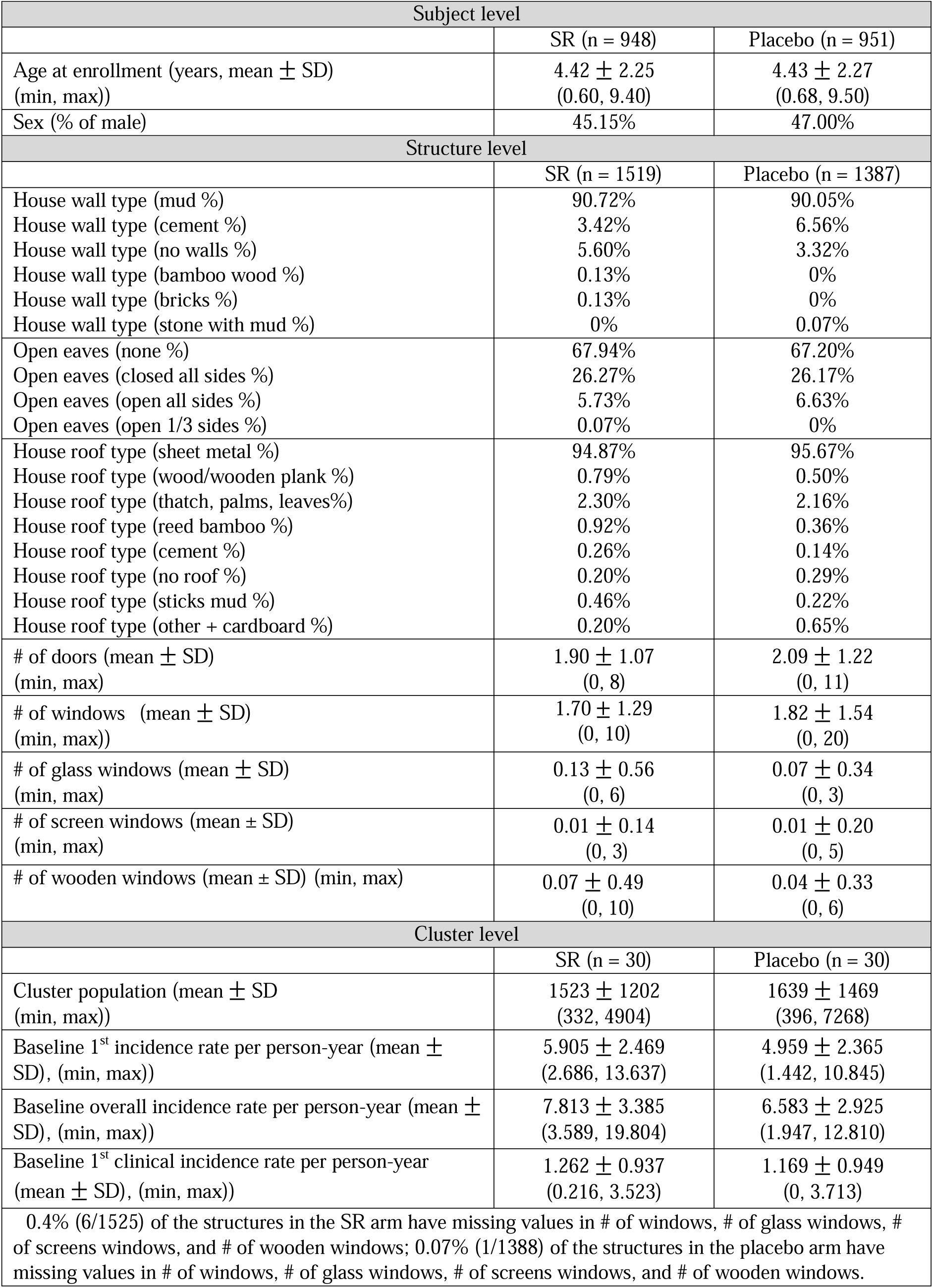
Baseline characteristics.

### Epidemiology

A total of 12,246 positive malaria infections were detected during 24 month intervention period (April 2022 to March 2024). Most infections were identified as *Plasmodium falciparum* (95%). In the SR arm, a total of 913 first-time; 6159 overall; and 409 clinical infections were reported with a total of 913 first-time; 6087 overall; and 413 clinical infections reported from the placebo arm. No statistically significant protective effect of SR against FTI, overall new infection, and first time-time clinical infection were observed compared to placebo (Figure 3, Table 2, and S2. Supplemental Information, Tables 1-7).

**Table 2.** Summary of protective efficacy estimates.

| First-time infection |  |  |  |
| --- | --- | --- | --- |
| Treatment | baseline incidence rate per person-year (mean ± SD) | # of subjects | # of 1<sup>st</sup>-time infections (person-years) |
| SR | 5.905 ± 2.469 | 922 | 913 (280.15) |
| Placebo | 4.959 ± 2.365 | 930 | 913 (283.04) |
| SR vs Placebo | Hazard ratio (1-sided 95% CI)<br>(2-sided 95% CI) | PE (%) (1-sided 95% CI)<br>(2-sided 95% CI) | 1-sided p-value<br>2-sided p-value |
|  | 1.031 (0, 1.270)<br>(0.804, 1.322) | -3.10 (-27.05, 100)<br>(-32.24, 19.62) | 0.405<br>0.810 |
| Baseline coefficient of variation (CV) of incidence rate: 45% |  |  |  |
| Overall new infections |  |  |  |
| Treatment | baseline incidence rate per person-year (mean ± SD) | # of subjects | # of new infection |
| SR | 7.813 ± 3.385 | 922 | 6159 |
| Placebo | 6.583 ± 2.925 | 930 | 6087 |
| SR vs Placebo | Hazard ratio (1-sided 95% CI)<br>(2-sided 95% CI) | PE (%) (1-sided 95% CI)<br>(2-sided 95% CI) | 1-sided p-value<br>2-sided p-value |
|  | 0.997 (0, 1.099)<br>(0.889, 1.119) | 0.26 (-9.85, 100)<br>(-11.90, 11.10) | 0.482<br>0.965 |
| Baseline coefficient of variation (CV) of incidence rate: 44% |  |  |  |
| 1<sup>st</sup>-time clinical infection |  |  |  |
| Treatment | baseline incidence rate per person-year (mean ± SD) | # of subjects | # of 1<sup>st</sup>-time clinical infection (person-years) |
| SR | 1.262 ± 0.937 | 922 | 409 (1241.76) |
| Placebo | 1.169 ± 0.949 | 930 | 413 (1240.63) |
| SR vs Placebo | Hazard ratio (95% CI) | PE (%) (95% CI) | 2-sided p-value |
|  | 1.082 (0.779, 1.502) | -8.19 (-50.18, 22.06) | 0.638 |
| Baseline coefficient of variation (CV) of incidence rate: 77% |  |  |  |

The estimated PE of SR against the first-time malaria infection relative to placebo was - 3.10% (95% 1-sided CI (-27.05%, 100%), 1-sided p-value 0.405, 95% 2-sided CI (-32.24%, 19.62%), 2-sided p-value 0.810). The PE of SR against overall new malaria infections relative to placebo was 0.26% (95% 1-sided CI (-9.85%, 100%), 1-sided p-value 0.482; 95% 2-sided CI (- 11.90%, 11.10%), 2-sided p-value 0.965). The estimated PE at the end of the intervention period of SR against first-time clinical malaria infection relative to placebo was -8.19% (95% 1- sided CI (-42.47%, 100%),1-sided p-value 0.319; 95% 2-sided CI (-50.18%, 22.06%), 2-sided p-value 0.638). Overall, the PE effects in younger children (under 5 years old) were slightly larger numerically, but not statistically significant and results do not suggest statistically significant effects of SR by season (rainy vs dry) or by year (Year 2 vs Year 1) compared to placebo (S2. Supplemental Information, Table 8-12).

SMC (>3-59 mo) coverage rate from July to October 2022 was 49.9%, 62.4%, 66.2% and 75.8%, respectively, while during July to October 2023 was 60.4%, 67.9%, 76.6% and 80.5%, respectively. Mass distribution of ITNs (Olyset-PBO) by the NMCP in June 2023 resulted in a reported district-level access/ownership rate of 93.2%. Community-level coverage of ITNs was not measured; however, study participant ITN usage measured during trial was 93.0% in the SR arm and 90.3% in the placebo arm (S2. Supplemental Information, Table 13).

### Entomology

Throughout both baseline and intervention phase, a total of 18,877 and 11,263 mosquitoes were collected by CDC-LT and HLC, respectively, of which most (56.3% and 80.9%, respectively) were *An. gambiae s.l.*. Overall, results do not indicate statistically significant effects of SR, compared to placebo, on the HLR (a proxy for human biting rate), parity rate, sporozoite positivity rate, EIRs, anopheline density, or blood fed rate (S2. Supplemental Information, Tables 14-24, Figures 6-7). Insecticide resistance characterization using the CDC-bottle assay indicated wild-type mosquitoes were sensitive (100% mortality at diagnostic time) to transfluthrin in six of 8 clusters with resistance suggested in two study clusters (70-90% mortality at diagnostic time). Mosquitoes were resistant (range 5% - 98% mortality at diagnostic time) to deltamethrin using WHO tube assays in all clusters where the test was carried out.

### Intervention coverage

Consent was comprehensive and applied to all relevant structures of the compound. There were no cases where a structure owner only consented to installing the SR product in part of the house.

Two types of ‘coverage’ measurements were assessed: 1) product placement rate. The percentage of products applied inside a household during each 28-day cycle in relation to the number of products required for the household based on manufacturer specifications, room area, and product rounding rules (S2. Supplemental Information, Figure 8); 2) product attrition rate. The percentage of products removed from a household during each 28-day cycle compared to the number of products required for the household.

#### 1. Product placement rate

Across all enrolled households throughout the entire intervention period, 85% of households in the SR arm had 100% placement rate, 14.1% had 0% placement rate, with <1% in between (0%, 100%). In the placebo arm, 82.3% had 100%, 17.0 % had 0%, with <1 % in between (0%, 100%) (S2. Supplemental Information, Table 25 and Figure 9).

#### 2. Product attrition rate

Across all 60 clusters, the average 28-day product attrition rate was ∼41% (0.01%, 82.4%) (S2. Supplemental Information, Table 26). Throughout the 24-mo follow up period, the average cluster-level product attrition rate fluctuated from 0% to 100% in the SR and placebo arms (S2. Supplemental Information, Figure 9). Across all enrolled households throughout the study, in the SR arm, on average, there were less than 52.2% of the target number of products in the household during the product retrieval process. In the placebo arm, on average, there were less than 57.4% of the target number of products in the household during the product retrieval process. (S2. Supplemental Information, Table 25 and Figure 10).

### Safety

During the baseline period and intervention phase, fever and vomiting were the top 2 symptoms in both occurrences and percentage of subjects who experienced at least one episode of the symptoms. Other observed symptoms include cough, headache, abdominal pain, loss of appetite, diarrhea, nausea, vomiting, chills, and others. Similar patterns of symptoms were observed across treatment arms, however, placebo clusters had more occurrences of AE symptoms than the SR clusters during the intervention phase. A total of 12 participant deaths occurred. No reported AEs or SAEs were deemed associated with SRs (S5. Safety Annex), patterns were consistent with disease frequency expected in study clusters, and safety assessments by the DSMB supported no significant safety concerns of the SR intervention.

### Intervention Quality Control

A total of 149 products were assessed by chemical analysis. Of these, 62 represented unused (from on-site storage) and 87 used products, the latter taken from a sub-sample of study households from both arms at time of replacement. All unused placebo products (n= 29) tested negative for transfluthrin. The average transfluthrin quantity from unused, SR active products (n= 33) was 101.2 ± 2.2 mg, within the manufacturer specification range (93.5 mg – 126.5 mg), and that of used, active SR products (n=87) 42.9 ± 28.9 mg (median 35.0mg, range 0.0mg-99.5mg). There are no specifications for used product. A total of 5 (out of 87) used, active SR products were within the specification for unused products. A total of 4 (out of 87) used, active SR products had no detectable residual transfluthrin.

## Discussion

Spatial repellents are designed to protect enclosed or semi-enclosed spaces from mosquitoes, and are being evaluated in large-scale clinical trials. The SC Johnson SC Johnson Mosquito Shield™ intervention used in the Mali trial is a passive emanator, meaning it doesn’t require electricity or a flame to work, and is designed to be placed on interior walls of enclosed spaces and is meant to serve as a complementary tool to ITNs for malaria control. Products in the SR intervention class are anticipated to be suitable for public health use in semi-permanent homes, clinics, temporary internally displaced person camps, and rapid response settings.

In the 2014 meeting of the WHO VCAG, the panel outlined five key evidence gaps for determining SR public health value and operational use guidance.(22) This included 1) whether SR could provide protection as a stand-alone and/or integrated tool; 2) the product coverage required for impact; 3) whether there is diversion or a community PE; 4) how efficacy may vary with setting (transmission dynamics); and 5) whether a pyrethroid-based SR may be affected by underlying resistance in the vector population. Since then, the underlying premise of new data generation for the intervention class has been to address these points to guide public health policy and operational programs on contextual conditions where SR interventions are expected to provide benefit, or not. The AEGIS program supported the clinical trial implemented by Ochomo et al which conclusively demonstrated reduced first-time malaria infection, and benefit to surrounding homes, using the SC Johnson Mosquito Shield ^TM^ in Busia County, Kenya.(1) Here we discuss findings from our Mali clinical trial in relation to VCAG data requirements.

Our trial was appropriately powered on the primary endpoint (first-time malaria incidence). However, no statistically significant or conclusive PE was demonstrated for the SR intervention against epidemiological or entomological endpoints compared to placebo. We propose several factors to have contributed to this lack of demonstrative effect.

### Population density and malaria incidence

The censused population density per study cluster reached up to 7,000 persons. As reference, this is two times greater than the reported cluster density in the Kenya SR trial(1). Such a relatively high population density is expected to influence the pool of individuals available to serve as infectious reservoirs and thus potentially increase the overall risk for malaria transmission to study participants residing in the study area. Any effect of the intervention, as deployed in this trial, may have been overwhelmed. To this point, the Kolondieba District historically has, and remains, to be one of the highest burdened districts in Mali.(23)In fact, baseline malaria incidence among Mali study participants was found to be 1.5 times greater (∼5 per person per year) compared to that of the Kenya context (∼3 per person per year).

### Presence of other vector control strategies

The NMCP conducted mass deployment of PBO ITNs across all households in our study clusters. However, this occurred almost a full year from the start of SR intervention (June 2023). There were no other vector control strategies implemented in the study area during the intervention period. Given the number of first-time malaria events required for the primary endpoint analyses was captured by May 2023, the PE outcome from our trial essentially reflects SR efficacy as a ‘stand-alone’ vector control strategy, complementary to SMC. Given the high malaria incidence rate reported during intervention, the SC Johnson Mosquito Shield ^TM^ may have been overwhelmed as deployed in this contextual setting suggesting that ITNs may be essential to reduce transmission before SRs are effective. In the Kenya trial, the NMCP-led mass PBO ITN campaign occurred one month prior to the start of the intervention phase, in which case the SR product was serving as a complementary vector control strategy and demonstrated effect reflecting SR added benefit to ITNs.

### Cluster Coverage and Product Placement

We achieved an average of 74% cluster household coverage, with ∼85% product placement rate in enrolled households over the intervention period, suggesting good coverage; however, only households within compounds with an enrolled participant were eligible and recruited to receive SR products. This approach may have resulted in diminishing a protective effect on non-participants in the community who may have served as infectious reservoirs and thus reducing the probability of SR effect on malaria transmission overall. This is particularly true in the context of the Kolondieba District which traditionally has experienced high transmission rates. In alignment with WHO VCAG recommendations, future trials should aim for implementation of cluster-level coverage of households.

The number of products placed by trained study teams were based on manufacturer specifications [2 units / 9m^2^] following rounding rules for structure design per the study standard operating protocol. However, enrolled households may not have met target placement rate throughout each 28-day period which may have resulted in incomplete protection. While we did not experience high cases of rejections from households to receive and hang the product for the full month, there were partial, or complete, compounds that experienced interrupted deployment due to study participant refusal. Those cases occurred one year after the trial initiation and typically were a result of inaccessibility by study staff due to travel by the head of household whom granted permission to receive the product. The team resolved those cases through homeowner education, raising awareness and explaining the scope and interest of the study and its objective which depended on SR placement.

Additionally, calculations of product attrition rate took into consideration the number of products removed from a structure at time of scheduled replacement in relation to the number of products that were required for that structure. Data showed ’0’, ’N/A’ and ’missing’ data points recorded at time of product replacement throughout the trial. For ’0’ data (indicating no product to remove), the root cause for the observed attrition is unclear. Additional activities related to the social/behavioral practices are necessary to better understand why products were missing after placement by study staff. For ’N/A’ data, these were treated as ’0’ during analyses to be overly conservative in assumptions of continual protection in the previous 28-day period. For ’missing’ data, it is uncertain whether a product was available for removal, or not. The root cause of ’N/A’ can be partially explained by the fact that staff reported some product’s barcodes were unscannable (e.g., air bubbles) when products were replaced. These data management reflections offer insights for future large-scale, SR studies.

### Product Durability and Homeowner Use

Product specifications for transfluthrin dosing of active SR products, and lack thereof in placebo products, was verified at time of manufacturing and supported by independent chemical analyses upon study completion. Results from product assessments indicated a proportion of used samples (5 of 87) removed from study households after 28 days of deployment had residual transfluthrin levels within specification for unopened products and 4 samples (out of 87) that had no residual transfluthrin. Both outcomes lead to uncertainty in post-placement durability (chemical emanation being absent and/or waning), either from environmental factors and/or homeowner use, between product placement periods and thus uncertainty in probability for continual protection. Future studies should include social science monitoring activities to assist in human behavior monitoring, as conducted in the AEGIS Kenya trial.(1)

### Other contextual factors

The lack of demonstrated entomological effect was not entirely unexpected given the trial was not powered to detect an effect of the SR intervention on vector-related endpoints. Mosquito endpoints are known to be associated with large variabilities and thus the CDC-LT and HLC sample size and/or sampling credence was likely not sufficient to generate robust datasets to detect a statistically significant difference between SR and placebo, even if such a difference indeed exists. Cluster coverage, product placement and attrition rates, and uncertainties on product emanation and use may have impacted the detection of any effect of SRs on entomology as well. Lastly, the mode of action of SRs includes reduced entry, reduced host-seeking and/or reduced human biting. Current field methodologies employed to measure these indicators may not be the most appropriate due to other mosquito behaviors that may be affected such as confusion and feeding inhibition. Lastly, although not measured during the trial, anecdotally, human nighttime behavior included a large amount of time spent outdoors late at night, thus decreasing time under protection.

In conclusion, while efficacy findings from our Mali study do not align with outcomes from previous SR clinical trials(8–10), particularly the AEGIS Kenya study using the same SR product (SC Johnson Mosquito Shield^TM^)(1), results have enhanced the knowledge-base for the SR intervention class regarding efficacy, coverage, durability, and safety. According to current WHO VCAG standards and processes, there remains a gap in evidence for SR health impact against malaria which will be required before a public health policy can be fully endorsed.

## Supporting information

Consort Checklist

Study Protocol

Statistical Analysis Plan

Safety Data Annex

## Data Availability

De-identified analytical datasets, including de-identified participant data, data dictionaries, and analytical codes, will be made available with publication on request to at.

## Ethics Approval

Ethical approval for the study protocol was granted by the Institutional Review Boards of the University of Sciences, Techniques and Technologies of Bamako, Mali (protocol #2020/252/CE/FMOS/FAPH), the University of Notre Dame (protocol #20-10-6245) and World Health Organization Ethical Review Committee (0003449). Informed consent was obtained from parents or legal guardians of study participants for cohort participation, mosquito collectors conducting human landing catch collections, and head of households for product placement.

## Clinical Trial Registration

The study was registered in clinicaltrials.gov (NCT04795648).

## Acknowledgments

We extend our sincere appreciation to Evercita Eugenio for her support in serving as external statistician on the DSMB. We thank additional DSMB members: Dennis Shanks, Chris Drakeley, and Neal Alexander for assurances on data integrity and safety assessments. We recognize the efforts of Noel Recla and Shaun Whitfield of the University of Notre Dame, Center for Research Computing for development of field and laboratory data entry systems and management of trial data. We thank Sofia Li-Harezlak for assistance with the safety summary. We also acknowledge Alex Perkins and Sean Moore of the University of Notre Dame for ongoing efforts in modeling impact projections. Our gratitude is extended to the WHO Vector Control Advisory Group for their comments during project assessments. We also express our appreciation to fhiClinical for trial oversight and monitoring of protocol compliance. The following reagent was obtained through BEI Resources, NIAID, NIH: *Plasmodium falciparum* Sporozoite ELISA Reagent Kit, MRA-890, contributed by Robert A. Wirtz.”

We express our thanks to Unitaid program management for their long-term support of SR product evaluation and development, especially Rachel Evans and Jan Kolaczinski. We are deeply grateful to SC Johnson & Son, Inc for providing integral industry and product expertise including the independent funding of the development, manufacturing, delivery, and shipment of the intervention (both SR and placebo) used in the trial.

We thank the Kolondieba District residents for their support and participation in this study and willingness to allow this study to be conducted in their community. We greatly appreciate the support of Dr Lassana Sissoko, the chief of Kolondieba district physician, who facilitated our work in Kolondieba. We thank MRTC and CRS leadership and staff who provided institutional support for implementation of the trial including Professor Abdoulaye Djimde, Director of PMRTC, USTTB, Bamako, Mali. We thank community leaders without whom the recruitment of study households and participants would not have been possible.

## Data Sharing

De-identified analytical datasets, including de-identified participant data, data dictionaries, and analytical codes, will be made available with publication on request to FL (at).

Authors’ current addresses: including affiliation, city, country, and email address

Malaria Research and Training Center (MRTC), Faculty of Medicine, Dentistry and Pharmacy at the University of Sciences, Techniques and Technologies of Bamako (USTTB), Bamako, Mali

Issaka Sagara

Catholic Relief Services, Baltimore, MD, USA

Suzanne

Catholic Relief Services, Dakar, Senegal

Catholic Relief Services, Bamako, Mali

Department of Biological Sciences, Eck Institute for Global Health, University of Notre Dame, Notre Dame, IN, USA.

Nicole L Achee

John P Grieco.

Ashley Hudson

Center for Research Computing, University of Notre Dame, Notre Dame, IN, USA.

Bradley Sandberg

Jared Hendrickson

Department of Applied and Computational Mathematics and Statistics, University of Notre Dame, Notre Dame, IN, USA.

Fang Liu

Ruyu Zhou

Xiaoan Lang

