## Supplementary material for "Effect of a spatial repellent on malaria incidence in Mali: a cluster-randomized, controlled trial": Consort Checklist

### S1. CONSORT Checklist

### Reporting checklist for randomised trial.

Based on the CONSORT guidelines.

#### Instructions to authors

Complete this checklist by entering the page numbers from your manuscript where readers will find each of the items listed below.

Your article may not currently address all the items on the checklist. Please modify your text to include the missing information. If you are certain that an item does not apply, please write "n/a" and provide a short explanation.

Upload your completed checklist as an extra file when you submit to a journal.

In your methods section, say that you used the CONSORTreporting guidelines, and cite them as:

Schulz KF, Altman DG, Moher D, for the CONSORT Group. CONSORT 2010 Statement: updated guidelines for reporting parallel group randomised trials

| | | Reporting Item | Page Number |
|---|---|---|---|
| **Title and Abstract** | | | |
| Title | #1a | Identification as a randomized trial in the title. | 1 |
| Abstract | #1b | Structured summary of trial design, methods, results, and conclusions | Abstract pgs. 3-4 |
| **Introduction** | | | |
| Background and objectives | #2a | Scientific background and explanation of rationale | Introduction pgs. 6-7 |
| Background and objectives | #2b | Specific objectives or hypothesis | Trial Design pg. 8 |
| **Methods** | | | |

| | | | |
|---|---|---|---|
| Trial design | #3a | Description of trial design (such as parallel, factorial) including allocation ratio. | Trial Design pg. 8 |
| Trial design | #3b | Important changes to methods after trial commencement (such as eligibility criteria), with reasons | n/a – no changes made |
| Participants | #4a | Eligibility criteria for participants | Participants pg. 10 |
| Participants | #4b | Settings and locations where the data were collected | Trial Design pgs. 7-8 |
| Interventions | #5 | The experimental and control interventions for each group with sufficient details to allow replication, including how and when they were actually administered | Randomization/Intervention pg. 11 |
| Outcomes | #6a | Completely defined prespecified primary and secondary outcome measures, including how and when they were assessed | Procedures pgs. 11-13<br><br>Statistical Analysis pgs. 14-15 |
| Outcomes | #6b | Any changes to trial outcomes after the trial commenced, with reasons | n/a – no changes made |
| Sample size | #7a | How sample size was determined. | Trial Design pgs. 9 |
| Sample size | #7b | When applicable, explanation of any interim analyses and stopping guidelines | n/a – no interim analysis or stopping guidelines |
| Randomization - Sequence generation | #8a | Method used to generate the random allocation sequence. | Randomization/Intervention pgs. 10-11 |

| | | | |
|---|---|---|---|
| Randomization - Sequence generation | #8b | Type of randomization; details of any restriction (such as blocking and block size) | Randomization/Intervention pgs. 10-11 |
| Randomization - Allocation concealment mechanism | #9 | Mechanism used to implement the random allocation sequence (such as sequentially numbered containers), describing any steps taken to conceal the sequence until interventions were assigned | Randomization/Intervention pg. 10-11 |
| Randomization - Implementation | #10 | Who generated the allocation sequence, who enrolled participants, and who assigned participants to interventions | Randomization/Intervention pgs. 10-11<br>Participants pgs. 9-10 |
| Blinding | #11a | If done, who was blinded after assignment to interventions (for example, participants, care providers, those assessing outcomes) and how. | Randomization/Intervention<br>pgs. 10-11 |
| Blinding | #11b | If relevant, description of the similarity of interventions | Randomization/Intervention<br>pgs. 10-11 |
| Statistical methods | #12a | Statistical methods used to compare groups for primary and secondary outcomes | Methods pg. 7<br>Statistical Analysis pgs. 14-15 |
| Statistical methods | #12b | Methods for additional analyses, such as subgroup analyses and adjusted analyses | Methods pg. 7<br>Statistical Analysis pgs. 14-15 |

**Results**

| | | | |
|---|---|---|---|
| Participant flow diagram (strongly recommended) | #13a | For each group, the numbers of participants who were randomly assigned, received intended treatment, and were analysed for the primary outcome | Figure 2. CONSORT Flow Diagram<br><br>Trial Design Pg. 8 |
| Participant flow | #13b | For each group, losses and exclusions after randomization, together with reason | Figure 2. CONSORT Flow Diagram |
| Recruitment | #14a | Dates defining the periods of recruitment and follow-up | Figure 1. Timeline<br><br>Trial Design pg. 8 |
| Recruitment | #14b | Why the trial ended or was stopped | Results pg. 16 |
| Baseline data | #15 | A table showing baseline demographic and clinical characteristics for each group | Table 1<br><br>Results pg. 16 |
| Numbers analysed | #16 | For each group, number of participants (denominator) included in each analysis and whether the analysis was by original assigned groups | Figure 1. CONSORT FlowDiagram<br><br>Table 2<br><br>Results pg. 16 |
| Outcomes and estimation | #17a | For each primary and secondary outcome, results for each group, and the estimated effect size and its precision (such as 95% confidence interval) | Table 2<br><br>Results pgs. 15-17<br><br>S3. Supplementary Appendix |
| Outcomes and estimation | #17b | For binary outcomes, presentation of both absolute and relative effect sizes is recommended | Table 2<br><br>Results pgs. 16-19<br><br>S3. Supplementary Appendix |
| Ancillary analyses | #18 | Results of any other analyses performed, including subgroup analyses and adjusted analyses, | Results pgs. 16-19<br><br>S3. Supplementary Appendix |

| | | | |
|---|---|---|---|
| | | distinguishing pre-specified from exploratory | |
| Harms | #19 | All important harms or unintended effects in each group (For specific guidance see CONSORT for harms) | Results pgs. 18-19<br>S5. Supplementary Appendix |
| **Discussion** | | | |
| Limitations | #20 | Trial limitations, addressing sources of potential bias, imprecision, and, if relevant, multiplicity of analyses | Discussion pgs.19-24 |
| Generalisability | #21 | Generalisability (external validity, applicability) of the trial findings | Discussion pgs. 19-24 |
| Interpretation | #22 | Interpretation consistent with results, balancing benefits and harms, and considering other relevant evidence | Discussion pgs. 19-24 |
| Registration | #23 | Registration number and name of trial registry | Methods pg.7 |
| **Other information** | | | |
| Interpretation | #22 | Interpretation consistent with results, balancing benefits and harms, and considering other relevant evidence | Discussion pgs. 19-24 |
| Registration | #23 | Registration number and name of trial registry | Methods pg.7 |
| Protocol | #24 | Where the full trial protocol can be accessed, if available | Methods pg.7<br>S3. Study Protocol |
| Funding | #25 | Sources of funding and other support (such as supply of drugs), role of funders | Financial Disclosure Statement<br>pgs. 26-27 |

None The CONSORT checklist is distributed under the terms of the Creative Commons Attribution License CC-BY. This checklist can be completed online using https://www.goodreports.org/, a tool made by the EQUATOR Network in collaboration with Penelope.ai
