## Supplemental Information for "Effect of a spatial repellent on malaria incidence in Mali: a cluster-randomized, controlled trial"

### S2. Supplemental Information

This appendix has been provided by the authors to give readers additional information about their work.

Figure 1. Location of 60 study clusters which were monitored for malaria incidence in Kolondieba District, Sikasso Region, Mali.

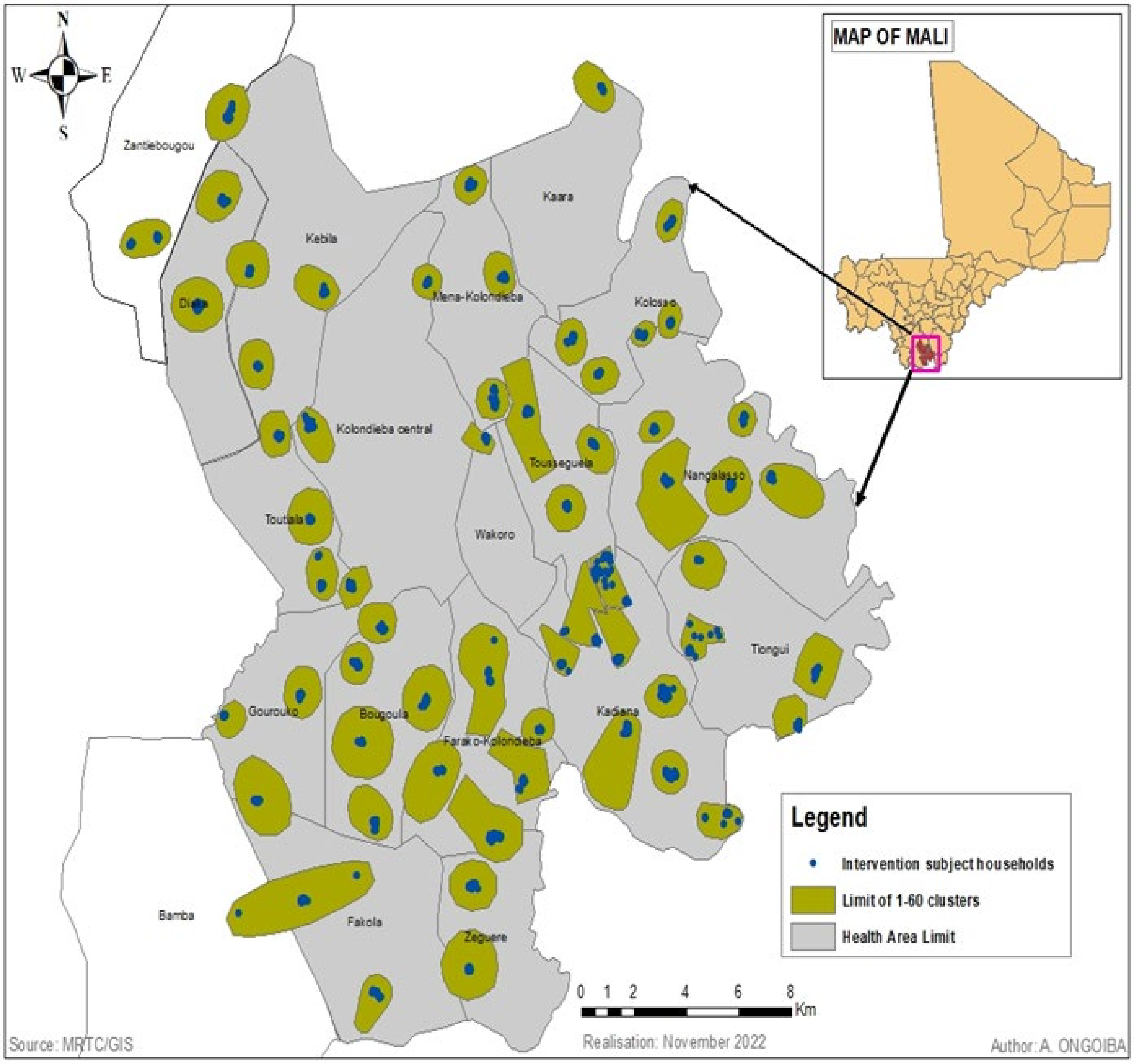

Figure 2. Mali cRCT Cluster Context and Terminology

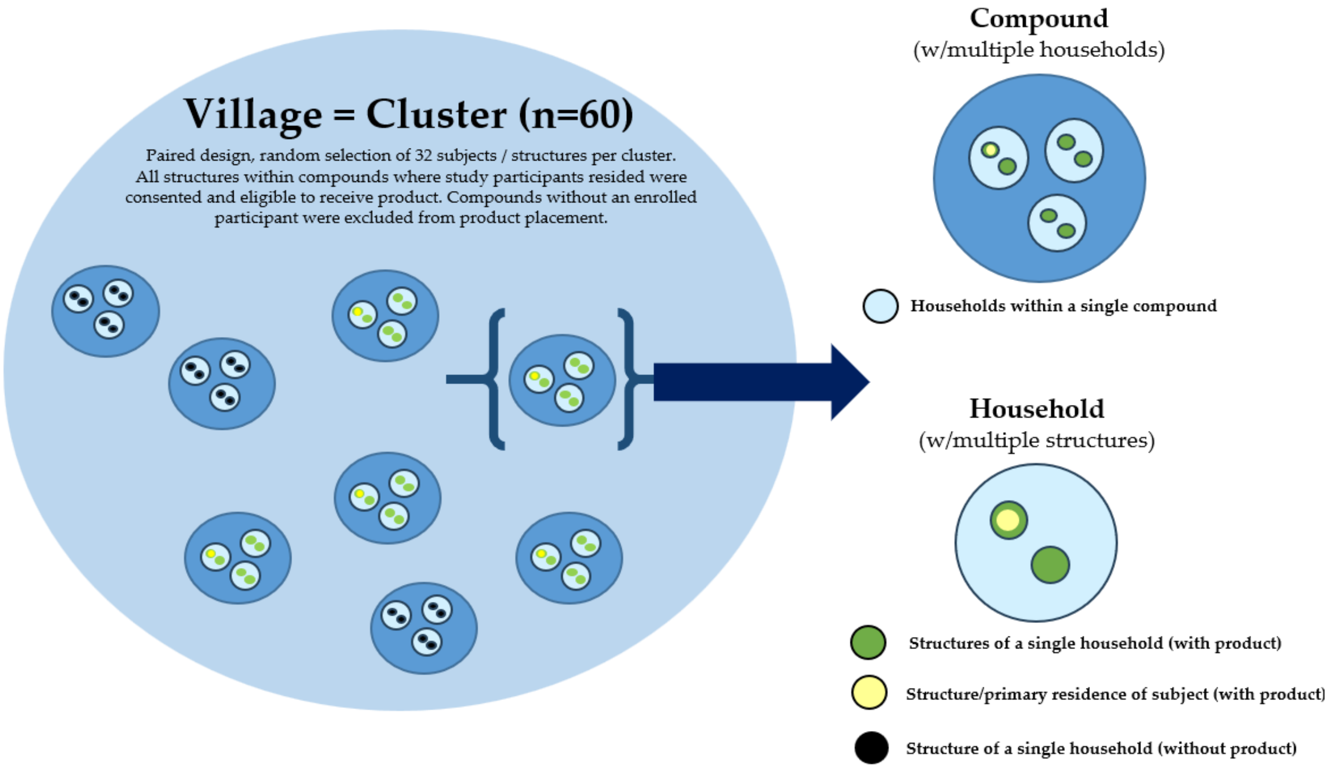


Figure 3. Mali cRCT Product Placement and Replacement Scheme

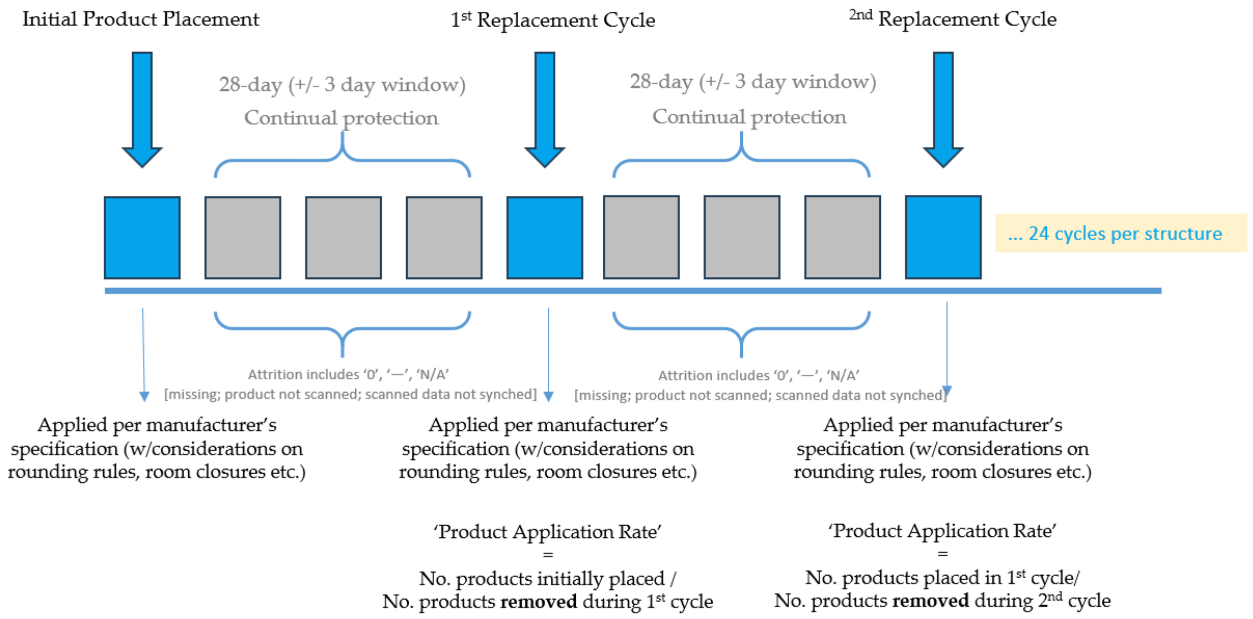

Figure 4. Mali cRCT Entomology Clusters

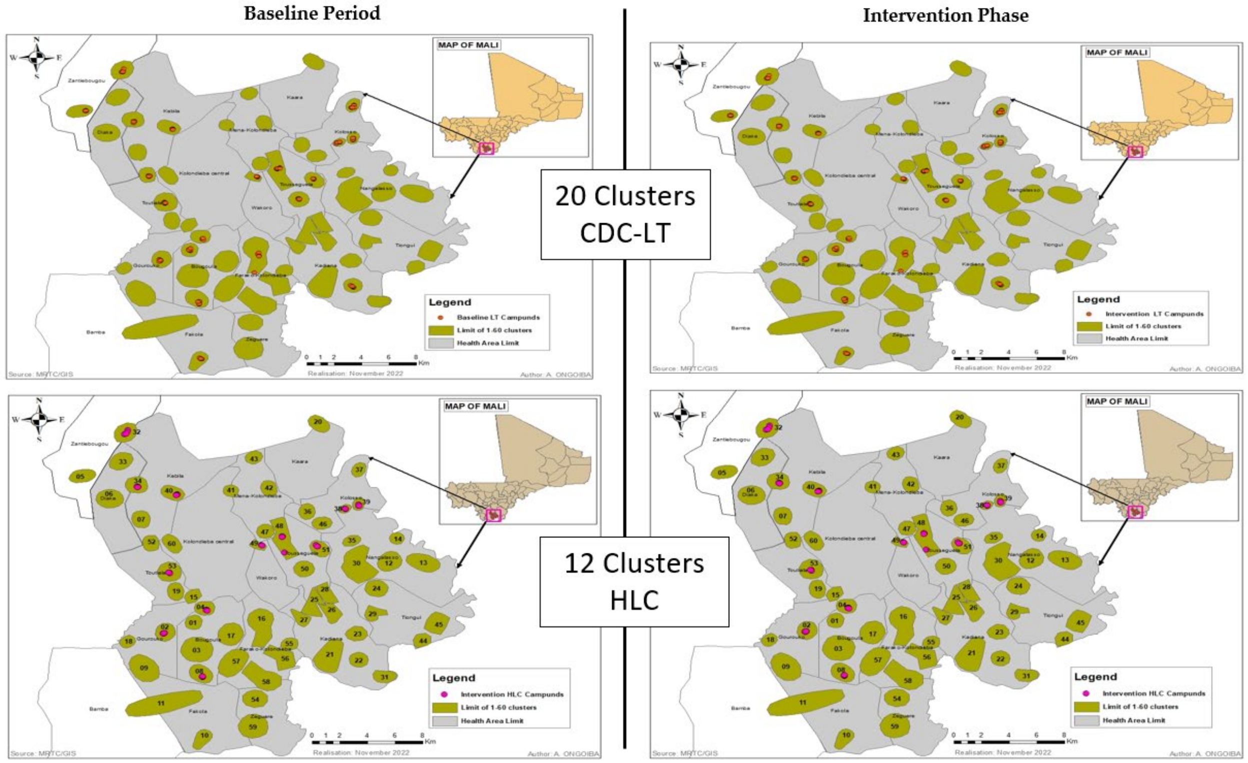


Figure 5. Mali cRCT Insecticide Resistance Clusters

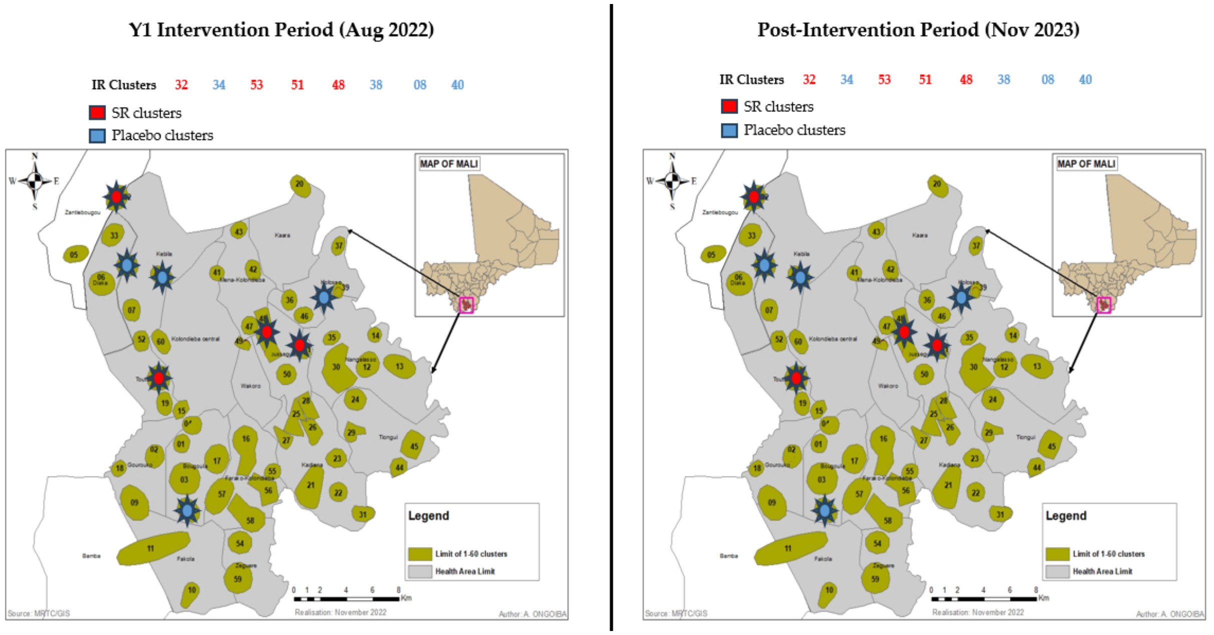

Table 1. Number of subjects at risk for the first-time infection during the intervention period

| Treatment | Week during intervention | | | | | | | | | | | | | | | | | | | | | | | |
|---|---|---|---|---|---|---|---|---|---|---|---|---|---|---|---|---|---|---|---|---|---|---|---|---|
| Number of subjects at risk for the first-time infection | | | | | | | | | | | | | | | | | | | | | | | | |
| | **4** | **8** | **12** | **16** | **20** | **24** | **28** | **32** | **36** | **40** | **44** | **48** | **52** | **56** | **60** | **64** | **68** | **72** | **76** | **80** | **84** | **88** | **92** | **96** |
| Placebo | 930 | 836 | 701 | 461 | 309 | 227 | 147 | 97 | 68 | 55 | 47 | 41 | 35 | 27 | 25 | 24 | 22 | 15 | 10 | 9 | 6 | 5 | 5 | 4 |
| SR | 922 | 833 | 706 | 461 | 302 | 218 | 150 | 100 | 67 | 55 | 48 | 40 | 32 | 28 | 27 | 25 | 21 | 15 | 11 | 10 | 8 | 3 | 3 | 2 |
| Number of clusters with subjects at risk for the first-time infection | | | | | | | | | | | | | | | | | | | | | | | | |
| | **4** | **8** | **12** | **16** | **20** | **24** | **28** | **32** | **36** | **40** | **44** | **48** | **52** | **56** | **60** | **64** | **68** | **72** | **76** | **80** | **84** | **88** | **92** | **96** |
| Placebo | 30 | 30 | 30 | 30 | 30 | 30 | 30 | 29 | 22 | 21 | 20 | 18 | 14 | 14 | 14 | 13 | 13 | 10 | 8 | 7 | 5 | 5 | 5 | 4 |
| SR | 30 | 30 | 30 | 30 | 30 | 30 | 29 | 27 | 25 | 24 | 22 | 21 | 18 | 17 | 16 | 14 | 13 | 11 | 8 | 8 | 6 | 2 | 2 | 1 |
| There were 17 censored subjects in the Placebo arm, and 9 censored subjects under the SR at various time points before the end of their follow-up. | | | | | | | | | | | | | | | | | | | | | | | | |

Table 2. Covariate effect on the hazard rate of 1st-time infection

| **Covariate** | **hazard ratio** | **95% CI** | **2-sided p-value↑↑** |
|---|---|---|---|
| **Age (year)** | **1.05** | **(1.02, 1.08)** | **0.0013** |
| Gender (F vs. M) | 0.96 | (0.85, 1.09) | 0.5197 |
| Number of doors | 0.94 | (0.88, 1.01) | 0.0802 |
| Number of windows | 1.05 | (0.98, 1.11) | 0.1429 |
| Number of glass windows | 1.10 | (0.95, 1.28) | 0.2096 |
| Number of screens windows | 0.86 | (0.62, 1.19) | 0.3680 |
| Number of wooden windows | 0.99 | (0.87, 1.13) | 0.8503 |
| Open eaves (closed vs. open) ↑ | 0.88 | (0.63, 1.23) | 0.4553 |
| Open eaves (none vs. open) ↑ | 1.03 | (0.77, 1.38) | 0.8398 |
| Wall type (mud/bamboo vs. no walls) * | 1.28 | (0.84, 1.96) | 0.2464 |
| Wall type (bricks/cement vs. no walls) ** | 0.87 | (0.54, 1.41) | 0.5714 |
| Roof type (cement vs. other) | 0.51 | (0.13, 2.05) | 0.3451 |
| Roof type (metal vs. other) | 0.87 | (0.35, 2.16) | 0.7586 |
| Roof type (mud vs. other) | 0.71 | (0.18, 2.71) | 0.6143 |
| Roof type (no roof vs. other) | 0.70 | (0.13, 3.65) | 0.6721 |
| Roof type (wood vs. other) # | 0.66 | (0.25, 1.77) | 0.4126 |
| Cluster population (increase by 1 unit on log scale) | 0.86 | (0.72, 1.03) | 0.1057 |
| Baseline incidence rate | 1.03 | (0.98, 1.09) | 0.1935 |

↑↑**raw p-values without multiplicity adjustment (multiple hypothesis testing).**
↑ "open 1 to 3 sides" and "open all sides" are collapsed into "open".
* "mud", "stone with mud", "adobe uncoated", "bamboo/wood", "cane/sticks/bamboo/reed" collapsed into "mud/bamboo".
* * "bricks", and "cement" are collapsed into "bricks/cement".
### "mat", "reed bamboo", "wooden planks", "wood" and "thatch/palms/leaves" are collapsed into "wood"; "cardboard" and "other" categories are collapsed into "other".

Table 3. Covariate effect on the hazard rate of overall new infection

| Covariate | hazard ratio | 95% CI | 2-sided p-value[↑↑] |
|---|---|---|---|
| **Age (year)** | **1.06** | **(1.04, 1.07)** | **<0.0001** |
| **Gender (F vs. M)** | **0.94** | **(0.90, 0.99)** | **0.0114** |
| Number of doors | 0.99 | (0.97, 1.02) | 0.4871 |
| Number of windows | 1.00 | (0.98, 1.03) | 0.8014 |
| Number of glass windows | 1.02 | (0.97, 1.08) | 0.4444 |
| Number of screens windows | 0.94 | (0.82, 1.08) | 0.3731 |
| Number of wooden windows | 0.95 | (0.90, 1.00) | 0.0681 |
| Open eaves (closed vs. open)[↑] | 1.12 | (0.99, 1.27) | 0.0617 |
| Open eaves (none vs. open)[↑] | 1.07 | (0.96, 1.20) | 0.2055 |
| Wall type (mud/bamboo vs. no walls)[*] | 1.08 | (0.91, 1.27) | 0.3803 |
| Wall type (bricks/cement vs. no walls)[**] | 0.98 | (0.81, 1.19) | 0.8379 |
| Roof type (cement vs. other) | 1.07 | (0.60, 1.93) | 0.8156 |
| Roof type (metal vs. other) | 1.32 | (0.92, 1.91) | 0.1368 |
| Roof type (mud vs. other) | 1.18 | (0.68, 2.04) | 0.5587 |
| Roof type (no roof vs. other) | 1.52 | (0.81, 2.86) | 0.1883 |
| Roof type (wood vs. other)[#] | 1.28 | (0.86, 1.89) | 0.2200 |
| Cluster population (increase by 1 unit on the log scale) | 0.96 | (0.89, 1.05) | 0.3867 |
| **Baseline incidence rate** | **1.03** | **(1.01, 1.04)** | **0.0062** |

[↑↑]**The p-values are raw p-values without multiplicity adjustment (multiple hypothesis testing).**
[↑] "open 1 to 3 sides" and "open all sides" are collapsed into "open".
[*] "mud", "stone with mud", "adobe uncoated", "bamboo/wood", and "cane/sticks/bamboo/reed" are collapsed into "mud/bamboo".
[**] "bricks", and "cement" are collapsed into "bricks/cement".
[#] "mat", "reed bamboo", "wooden planks", "wood" and "thatch/palms/leaves" are collapsed into "wood"; "cardboard" and "other" categories are collapsed into "other".

Table 4. Covariate effect on the hazard rate of 1st-time clinical infection

| **Covariate** | **hazard ratio** | **95% CI** | **2-sided p-value**[↑↑] |
|---|---|---|---|
| **Age (year)** | **0.96** | **(0.93, 0.99)** | **0.0202** |
| Gender (F vs. M) | 1.00 | (0.86, 1.15) | 0.9527 |
| Number of doors | 1.08 | (0.99, 1.18) | 0.0666 |
| Number of windows | 0.96 | (0.89, 1.03) | 0.2713 |
| Number of glass windows | 1.04 | (0.85, 1.26) | 0.7207 |
| Number of screens windows | 0.79 | (0.46, 1.34) | 0.3806 |
| Number of wooden windows | 1.03 | (0.89, 1.20) | 0.6891 |
| Open eaves (closed vs. open)[↑] | 0.90 | (0.60, 1.36) | 0.6250 |
| Open eaves (none vs. open)[↑] | 1.20 | (0.83, 1.73) | 0.3280 |
| Wall type (mud/bamboo vs. no walls)[*] | 0.74 | (0.47, 1.16) | 0.1839 |
| Wall type (bricks/cement vs. no walls)[**] | 0.92 | (0.55, 1.54) | 0.7464 |
| Roof type (cement vs. other) | 4.69 | (0.70, 31.5) | 0.1120 |
| Roof type (metal vs. other) | 2.05 | (0.48, 8.71) | 0.3314 |
| Roof type (mud vs. other) | 2.11 | (0.18, 24.8) | 0.5515 |
| Roof type (no roof vs. other) | 4.89 | (0.70, 33.9) | 0.1085 |
| Roof type (wood vs. other)[#] | 1.86 | (0.41, 8.43) | 0.4235 |
| Cluster population (increase by 1 unit on the log scale) | 0.94 | (0.74, 1.19) | 0.6136 |
| **Baseline incidence rate** | **1.39** | **(1.17, 1.66)** | **0.0002** |

[↑↑]**The p-values are raw p-values without multiplicity adjustment (multiple hypothesis testing).**
[↑] "open 1 to 3 sides" and "open all sides" are collapsed into "open".
[*] "mud", "stone with mud", "adobe uncoated", "bamboo/wood", and "cane/sticks/bamboo/reed" are collapsed into "mud/bamboo".
[**] "bricks", and "cement" are collapsed into "bricks/cement".
[#] "mat", "reed bamboo", "wooden planks", "wood" and "thatch/palms/leaves" are collapsed into "wood"; "cardboard" and "other" categories are collapsed into "other".

Table 5. PE of SR against 1st-time infection estimated by model without covariates

| **Treatment** | **baseline incidence rate per person-year (mean ± SD)** | **# of subjects** | **# of 1st-time infection (sum of person-years)** |
|---|---|---|---|
| SR | 5.905 ± 2.469 | 922 | 913 (280.15) |
| Placebo | 4.959 ± 2.365 | 930 | 913 (283.04) |
| Comparison | **Hazard ratio (2-sided 95% CI)** | **PE (%) (2-sided 95% CI)** | **2-sided p-value** |
| | (0.830, 1.393) | -7.53 (-39.31, 16.99) | 0.582 |
| Baseline coefficient of variation (CV) of incidence rate: 45% | | | |

Table 6. PE of SR against overall new infection estimated by model without covariates

| **Treatment** | **baseline incidence rate per person-year (mean ± SD)** | **# of subjects** | **# of new infection** |
|---|---|---|---|
| SR | 7.813 ± 3.385 | 922 | 6159 |
| Placebo | 6.583 ± 2.925 | 930 | 6087 |
| Comparison | **Hazard ratio (95% CI)** | **PE (%) (95% CI)** | **2-sided p-value** |
| | 1.028 (0.908, 1.164) | -2.84 (-16.43, 9.16) | 0.658 |

| Baseline coefficient of variation (CV) of incidence rate: 44% |
|---|

Table 7. PE of SR against the 1st-time clinical infection estimated by model without covariates

| Treatment | baseline incidence rate per person-year (mean ± SD) | # of subjects | # of 1st-time clinical infection (person-years) |
|---|---|---|---|
| SR | 1.262 ± 0.937 | 922 | 409 (1241.76) |
| Placebo | 1.169 ± 0.949 | 930 | 413 (1240.63) |
| Comparison | **Hazard ratio (95% CI)** | **PE (%) (95% CI)** | **2-sided p-value** |
| | 1.134 (0.776, 1.657) | -13.39 (-65.70, 22.41) | 0.516 |
| Baseline coefficient of variation (CV) of incidence rate: 77% | | | |

Table 8. PE of SR against 1st-time infection by age group

| Treatment | # of subjects | # of 1st-time infection | Hazard ratio (95% CI) | PE (%) (95% CI) |
|---|---|---|---|---|
| | Age group [16 months, 6 years) | | | |
| SR | 633 | 627 | 0.959 (0.741, 1.231) | 4.11 (-23.05, 25.87) |
| Placebo | 626 | 611 | | |
| | Age group [6 years, 10.1 years) | | | |
| SR | 289 | 286 | 1.177 (0.856, 1.618) | -17.65 (-61.78, 14.44) |
| Placebo | 304 | 302 | | |

Table 9. PE of SR against overall new infections by age group

| Treatment | # of subjects | # of new infections | Hazard ratio (95% CI) | PE (%) (95% CI) |
|---|---|---|---|---|
| | Age group [16 months, 6 years) | | | |
| SR | 633 | 4088 | 0.985 (0.871, 1.113) | 1.54 (-11.26, 12.87) |
| Placebo | 626 | 3892 | | |
| | Age group [6 years, 10.1 years) | | | |
| SR | 289 | 2071 | 0.991 (0.866, 1.133) | 0.897 (-13.35, 13.35) |
| Placebo | 304 | 2195 | | |

Table 10. PE of SR against 1st-time clinical infection by age group

| Treatment | # of subjects | # of 1st-time clinical infection | Hazard ratio (95% CI) | PE (%) (95% CI) |
|---|---|---|---|---|
| | Age group [16 months, 6 years) | | | |
| SR | 633 | 299 | 1.101 (0.794, 1.525) | -10.07 (-52.51, 20.57) |
| Placebo | 626 | 283 | | |
| | Age group [6 years, 10.1 years) | | | |
| SR | 289 | 110 | 1.114 (0.659, 1.882) | -11.40 (-88.17, 34.05) |
| Placebo | 304 | 130 | | |

Table 11. PE by seasonality (rainy vs dry) and by year on three malaria infection endpoints

| infection | PE (%) in rainy season (95% CI) | PE (%) in dry season (95% CI) | 2-sided p-value (interaction between season and treatment) |
|---|---|---|---|
| first time | 0.66 (-46.76, 32.76) | 6.73 (-43.32, 39.30) | 0.6392 |
| Overall | 0.49 (-11.90, 11.51) | -0.09 (-14.03, 12.14) | 0.8895 |
| first-time clinical | -11.89 (-55.85, 19.67) | 9.29 (-53.90, 46.53) | 0.3723 |
| **infection** | **PE (%) in Year 1 (95% CI)** | **PE (%) in Year 2 (95% CI)** | **2-sided p-value (interaction between year and treatment)** |
| first time | -1.49 (-31.83, 21.87) | 8.71 (-74.85, 52.33) | 0.7354 |
| Overall | 1.24 (-11.61, 12.61) | -0.86 (-14.02, 10.78) | 0.5700 |
| first-time clinical | 5.39 (-39.41, 35.79) | -7.28 (-63.55, 29.63) | 0.4121 |

Table 12. PE by seasonality (rainy vs dry), assuming a one-month hysteresis effect in the seasonality effect on malaria infection, and by year on three malaria infection endpoints

| infection | PE (%) in rainy season (95% CI) | PE (%) in dry season (95% CI) | 2-sided p-value (interaction between season and treatment) |
|---|---|---|---|
| first time | 3.04 (-44.15, 34.78) | 1.94 (-46.38, 34.31) | 0.9174 |
| Overall | 0.51 (-11.88, 11.53) | -0.84 (-15.13, 11.68) | 0.7568 |
| first-time clinical | -10.65 (-54.38, 20.70) | -1.07 (-70.68, 40.15) | 0.6995 |
| | **PE (%) in Year 2 (95% CI)** | **PE (%) in Year 1 (95% CI)** | **2-sided p-value (interaction between year and treatment)** |
| first time | 8.07 (-75.84, 51.94) | -3.42 (-32.71, 19.40) | 0.7064 |
| Overall | -1.39 (-14.75, 10.42) | 1.05 (-11.85, 12.46) | 0.5112 |
| first-time clinical | -13.38 (-71.57, 25.07) | 1.36 (-45.84, 33.29) | 0.3614 |

Table 13. summary of bednet usage among the enrolled subjects during intervention

| Treatment | Yes | No | Not asked | Number of follow ups in enrolled subjects |
|---|---|---|---|---|
| SR | 93.02% | 2.82% | 4.23% | 22712 |
| Placebo | 90.33% | 4.97% | 3.78% | 22485 |

Figure 6. Mean (SD) HLR by treatment in the HLC collection (averaged over 6 cluster per treatment with roughly 4 households per cluster)

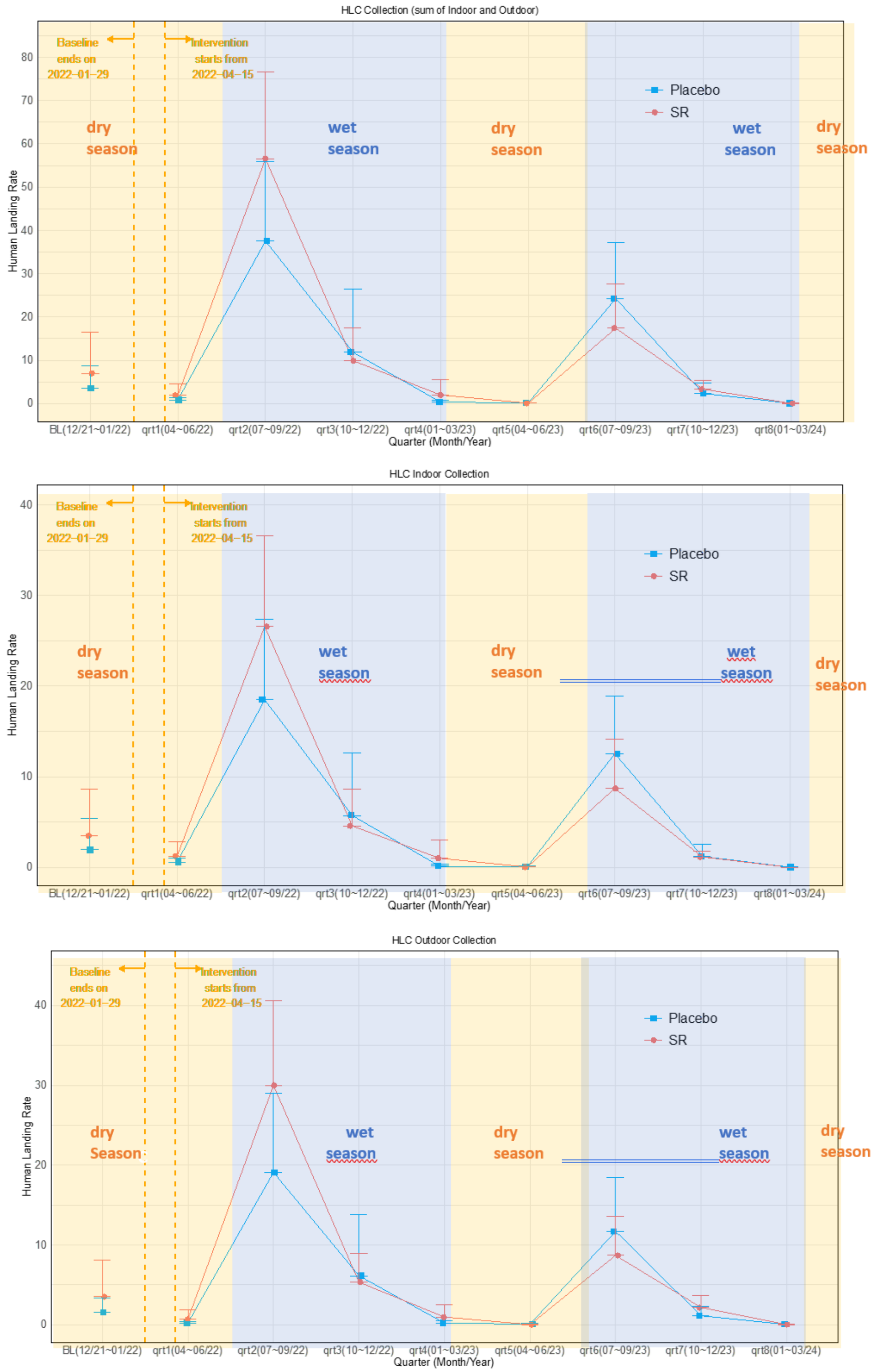

Table 14. Summary statistics of HLR in the HLC collection (n is the number of clusters)

| | | mean (± SD) HLR per household-day (min, max) | |
|---|---|---|---|
| **study phase** | **location** | **SR (n = 6)** | **placebo (n = 6)** |
| baseline | Indoor | 3.49 ± 5.12 (0, 13.56) | 1.95 ± 3.48 (0.19, 9.00) |
| | outdoor | 3.56 ± 4.52 (0.04, 11.38) | 1.58 ± 1.81 (0.25, 5.06) |
| intervention | Indoor | 5.41 ± 2.30 (3.66, 9.66) | 4.84 ± 2.44 (2.27, 9.19) |
| | outdoor | 5.84 ± 2.56 (3.28, 9.22) | 4.83 ± 2.84 (2.15, 10.06) |

Table 15. Effects of SR and covariates on HLR in the HLC collection

| Comparison | % Change (95% CI) | 2-sided p-value |
|---|---|---|
| Indoor: SR vs placebo | -5.85 (-51.95, 84.49) | 0.824 |
| Outdoor: SR vs placebo | 11.64 (-44.07, 122.87) | 0.692 |
| qrt2(07~09/22) vs qrt1(04~06/22) | 2252.63 (1658.69, 3047.15) | <0.001 |
| qrt3(10~12/22) vs qrt1(04~06/22) | 460.25 (311.79, 662.24) | <0.001 |
| qrt4(01~03/23) vs qrt1(04~06/22) | -37.03 (-59.58, -1.90) | 0.041 |
| qrt5(04~06/23) vs qrt1(04~06/22) | -82.93 (-91.33, -66.39) | <0.001 |
| qrt6(07~09/23) vs qrt1(04~06/22) | 970.65 (695.86, 1340.31) | <0.001 |
| qrt7(10~12/23) vs qrt1(04~06/22) | 108.03 (48.19 192.02) | <0.001 |
| qrt8(01~03/24) vs qrt1(04~06/22) | -94.78 (-98.37, -83.27) | <0.001 |
| Cluster population (increased by 1 unit on the log-scale) | 26.34 (-19.83, 99.12) | 0.314 |
| Baseline HLR (increase by 1 unit on the log-scale) | 3.65 (-6.16, 14.50) | 0.480 |

Table 16. Summary statistics of anopheline parity in the HLC collection (n = number of clusters)

| Treatment | Study Phase | Location | Parous | Nulliparous | Parity Rate |
|---|---|---|---|---|---|
| SR | Baseline | In | 206 | 0 | 100.00% |
| | | Out | 209 | 2 | 99.05% |
| | Intervention | In | 1699 | 27 | 98.44% |
| | | Out | 1867 | 24 | 98.73% |
| Placebo | Baseline | In | 125 | 0 | 100.00% |
| | | Out | 70 | 1 | 98.59% |
| | Intervention | In | 1637 | 19 | 98.73% |
| | | Out | 1628 | 21 | 98.73% |

Figure 7. Mean (SD) anopheline density by treatment of the LT collection (averaged over 10 clusters per treatment with roughly 10 households per cluster)

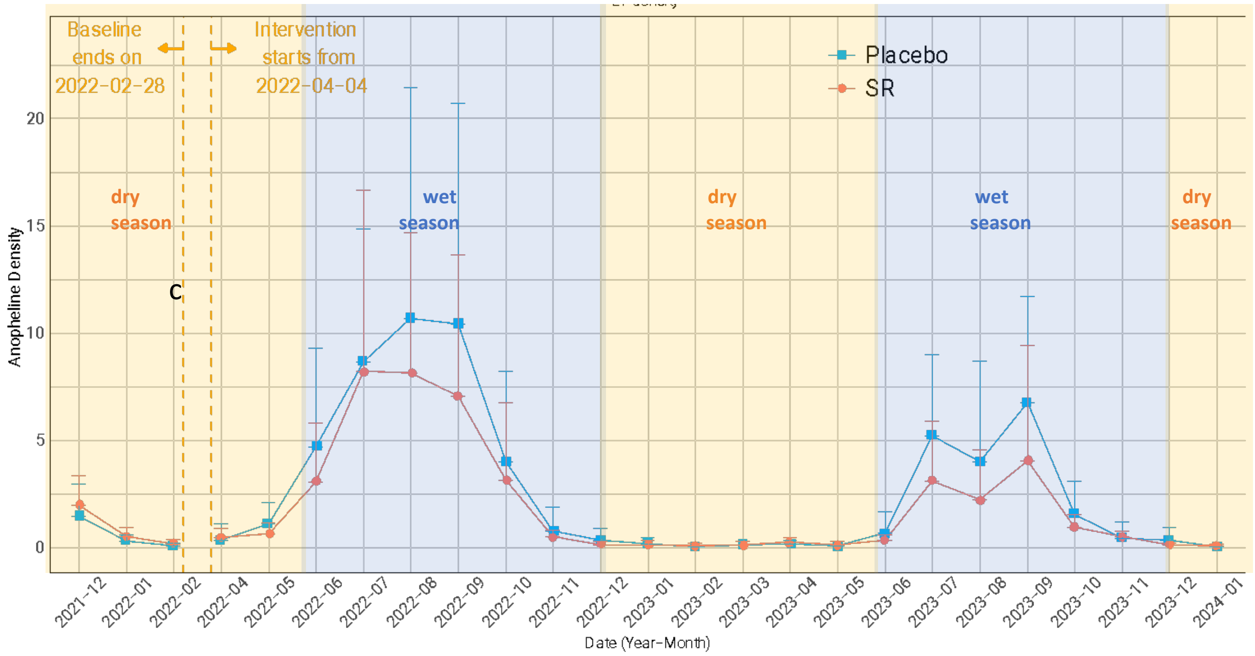


Table 17. Summary statistics on anopheline density of the LT collection (n is the number of clusters)

| | SR (n = 10) | placebo (n = 10) |
|---|---|---|
| Baseline anopheline count per household-day (mean ± SD, (min, max)) | 0.89 ± 0.54 (0.23, 1.83) | 0.62 ± 0.55 (0.08, 1.62) |
| Intervention anopheline count per household-day (mean ± SD, (min, max)) | 1.97 ± 1.23 (0.58, 3.89) | 2.75 ± 1.24 (0.81, 4.40) |

Table 18. Effects of SR and other covariates on anopheline density in the LT collection

| Comparison | % Change (95% CI) | 2-sided p-value |
|---|---|---|
| SR vs placebo | -0.85 (-22.82, 27.40) | 0.947 |
| 2022-05 vs 2022-04 | 103.52 (40.62, 194.58) | <0.001 |
| 2022-06 vs 2022-04 | 388.73 (248.13, 586.12) | <0.001 |
| 2022-07 vs 2022-04 | 719.00 (488.04, 1040.66) | <0.001 |
| 2022-08 vs 2022-04 | 766.95 (521.78, 1108.80) | <0.001 |
| 2022-09 vs 2022-04 | 721.00 (489.27, 1043.86) | <0.001 |
| 2022-10 vs 2022-04 | 379.20 (241.38, 572.64) | <0.001 |
| 2022-11 vs 2022-04 | 70.22 (16.00, 149.78) | 0.007 |
| 2022-12 vs 2022-04 | -24.36 (-52.45, 20.32) | 0.238 |
| 2023-01 vs 2022-04 | -59.42 (-77.10, -28.08) | 0.002 |
| 2023-02 vs 2022-04 | -80.54 (-90.84, -58.65) | <0.001 |
| 2023-03 vs 2022-04 | -57.46 (-75.70, -25.54) | 0.003 |
| 2023-04 vs 2022-04 | -14.28 (-45.21 34.13) | 0.500 |
| 2023-05 vs 2022-04 | -62.95 (-79.39, -33.43) | <0.001 |
| 2023-06 vs 2022-04 | 16.93 (-22.98, 77.52) | 0.463 |
| 2023-07 vs 2022-04 | 423.90 (273.59, 634.68) | <0.001 |
| 2023-08 vs 2022-04 | 342.04 (213.91, 522.46) | <0.001 |
| 2023-09 vs 2022-04 | 450.53 (291.04, 675.06) | <0.001 |
| 2023-10 vs 2022-04 | 129.72 (58.95, 232.02) | <0.001 |
| 2023-11 vs 2022-04 | 4.43 (-31.94, 60.22) | 0.843 |
| 2023-12 vs 2022-04 | -45.45 (-67.56, -8.28) | 0.022 |
| 2024-01 vs 2022-04 | -80.35 (-90.75, -58.26) | <0.001 |
| Cluster population (every 1 unit increase on the log-scale) | -32.09 (-45.30, -15.70) | <0.001 |
| Baseline density (every 1 unit increase on the log-scale) | 10.54 (-3.73, 26.92) | 0.155 |

Table 19. Frequency and percentage of female anopheline abdominal status in the LT collection

| abdominal status | baseline | | intervention | | Total |
|---|---|---|---|---|---|
| | SR | placebo | SR | placebo | |
| Fed | 20 (8.33%) | 17 (10.62%) | 537 (12.46%) | 578 (9.68%) | 1152 |
| Gravid | 35 (14.58%) | 29 (18.12%) | 202 (4.69%) | 293 (4.91%) | 559 |
| Half Gravid | 12 (5.00%) | 9 (5.62%) | 138 (3.20%) | 122 (2.04%) | 281 |
| Unfed | 173 (72.08%) | 105 (65.62%) | 3432 (79.65%) | 4979 (83.37%) | 8689 |
| Total | 240 (100%) | 160 (100%) | 4309 (100%) | 5972 (100%) | 10681 |

Table 20. Summary statistics on female anopheline blood fed rate (fed, gravid, and half-gravid) in the LT collection (n is the number of clusters)

| Study phase | **SR (n = 10)** | **placebo (n = 10)** |
|---|---|---|
| Baseline (mean ± SD, (min, max)) | 31.1% ± 21.6% (5.4%, 63.5%) | 40.5% ± 31.4% (0.0%, 100.0%) |
| Intervention (mean ± SD, (min, max)) | 25.7% ±9.4% (12.8%, 43.8%) | 24.4% ± 6.3% (15.8%, 38.6%) |

Table 21. Effects of covariates on female anopheline blood fed rate (fed, gravid, and half-gravid) in the LT collection

| **Comparison** | **Odds ratio (95% CI)** | **2-sided p-value** |
|---|---|---|
| SR vs placebo | 0.98 (0.69, 1.39) | 0.902 |
| 2022-05 vs 2022-04 | 0.89 (0.46, 1.72) | 0.722 |
| 2022-06 vs 2022-04 | 0.26 (0.14, 0.45) | <0.001 |
| 2022-07 vs 2022-04 | 0.20 (0.11, 0.35) | <0.001 |
| 2022-08 vs 2022-04 | 0.17 (0.10, 0.30) | <0.001 |
| 2022-09 vs 2022-04 | 0.15 (0.08, 0.26) | <0.001 |
| 2022-10 vs 2022-04 | 0.12 (0.07, 0.23) | <0.001 |
| 2022-11 vs 2022-04 | 0.11 (0.05, 0.23) | <0.001 |
| 2022-12 vs 2022-04 | 0.09 (0.03, 0.27) | <0.001 |
| 2023-01 vs 2022-04 | 0.21 (0.07, 0.62) | 0.005 |
| 2023-02 vs 2022-04 | 0.15 (0.03, 0.83) | 0.029 |
| 2023-03 vs 2022-04 | 0.31 (0.10, 0.94) | 0.038 |
| 2023-04 vs 2022-04 | 0.20 (0.07, 0.55) | 0.002 |
| 2023-05 vs 2022-04 | 0.46 (0.13, 1.63) | 0.231 |
| 2023-06 vs 2022-04 | 0.28 (0.14, 0.58) | 0.001 |
| 2023-07 vs 2022-04 | 0.21 (0.12, 0.37) | <0.001 |
| 2023-08 vs 2022-04 | 0.18 (0.10, 0.33) | <0.001 |
| 2023-09 vs 2022-04 | 0.12 (0.07, 0.22) | <0.001 |
| 2023-10 vs 2022-04 | 0.14 (0.07, 0.27) | <0.001 |
| 2023-11 vs 2022-04 | 0.15 (0.07, 0.33) | <0.001 |
| 2023-12 vs 2022-04 | 0.03 (0.01, 0.15) | <0.001 |
| 2024-01 vs 2022-04 | 0.00 (Inf, 0.00) | 0.991 |
| Cluster population (increased by 1 unit on the log-scale) | 1.51 (1.08, 2.10) | 0.015 |
| Baseline blood fed rate (increased by 1%) | 0.92 (0.46, 1.84) | 0.807 |

Table 22. Summary statistics on sporozoite positivity in the HLC collection

| Location | Treatment | Positive | Negative | Sporozoite Positivity Rate |
|---|---|---|---|---|
| **baseline** | | | | |
| Indoor | SR | 20 | 309 | 6.08% |
| | placebo | 3 | 182 | 1.62% |
| Outdoor | SR | 22 | 299 | 6.85% |
| | placebo | 3 | 147 | 2.00% |
| **intervention** | | | | |
| Indoor | SR | 34 | 2031 | 1.65% |
| | placebo | 32 | 1842 | 1.71% |
| Outdoor | SR | 29 | 2258 | 1.27% |
| | placebo | 34 | 1820 | 1.83% |

Table 23. Summary statistics of EIR in the HLC collection

| Location | Treatment | Baseline | Intervention |
|---|---|---|---|
| Indoor | SR | 0.3864 | 0.1609 |
| | placebo | 0.0685 | 0.1718 |
| Outdoor | SR | 0.4565 | 0.1240 |
| | placebo | 0.0410 | 0.1740 |

Table 24. Summary statistics on sporozoite positivity in the LT collection

| Treatment | Positive | Negative | Sporozoite Positivity Rate |
|---|---|---|---|
| **Baseline** | | | |
| SR | 4 | 203 | 1.93% |
| placebo | 4 | 136 | 2.86% |
| **Intervention** | | | |
| SR | 49 | 4134 | 1.17% |
| placebo | 88 | 5804 | 1.49% |

Figure 8. Product Volume and Placement Scheme

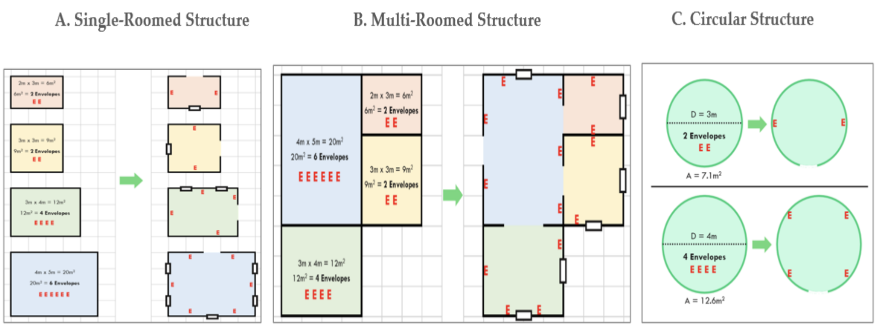


NOTES:

1. Formula for product volume requirements: (**room_area**/9), rounded up to nearest whole number, x 2
2. A small structure with multiple rooms may require a greater product volume than a larger structure with only one room.

Table 25. Percentage of household-level product placement and retrieval rates based on product application requirement* overall and by treatment throughout the intervention period (categorized).

| | 0% | (0, 25]% | (25, 50]% | (50, 75]% | (75, 100)% | 100% |
|---|---|---|---|---|---|---|
| | SR | | | | | |
| placement | 14.14 | 0.04 | 0.53 | 0.21 | 0.07 | 85.00 |
| retrieval | 50.00 | 0.19 | 1.12 | 0.53 | 0.36 | 47.81 |
| | placebo | | | | | |
| placement | 16.96 | 0.05 | 0.37 | 0.29 | 0.06 | 82.27 |
| retrieval | 53.93 | 0.22 | 1.30 | 1.11 | 0.82 | 42.62 |
| The product placement rate in the SR arm was 85% and that in the placebo arm was 82.27%, providing evidence on the team's effort in product placement during the study. *Manufacturer specification: 2 units / $9m^2$ | | | | | | |

Figure 9. Time profile of mean (the error bar stands for SD) product placement rate (0 to 100%) by treatment group

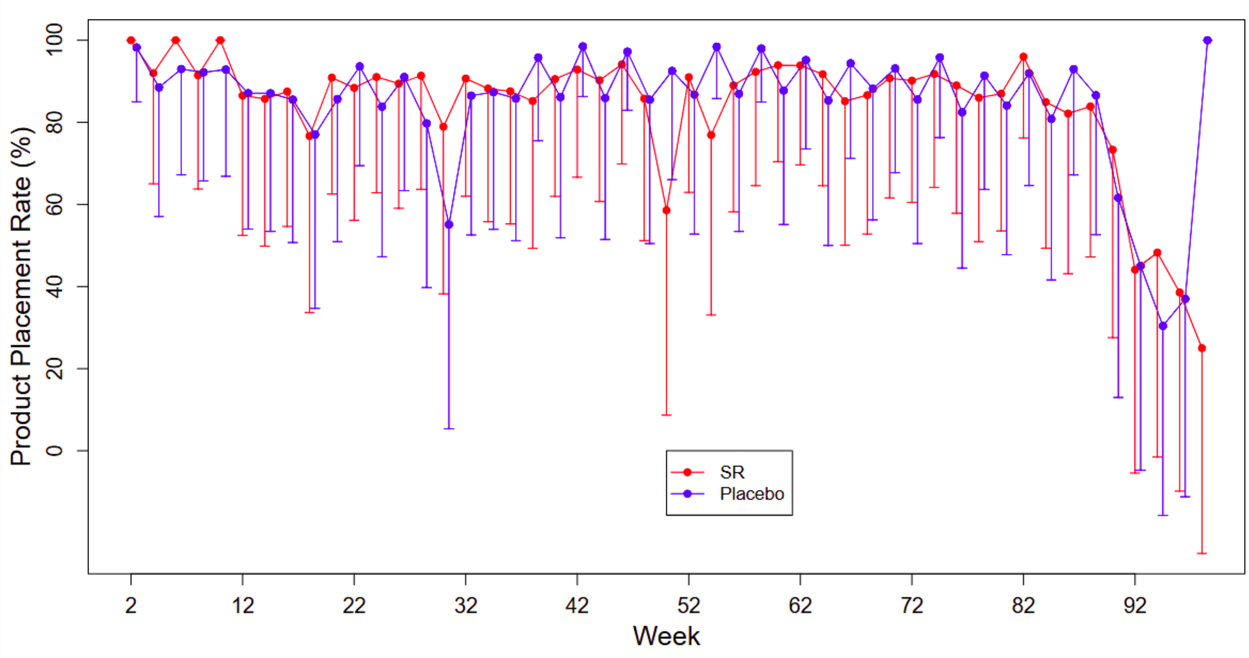

Figure 10. Time profile of **mean (the error bar stands for SD)** product attrition rate (0 to 100%) by treatment group

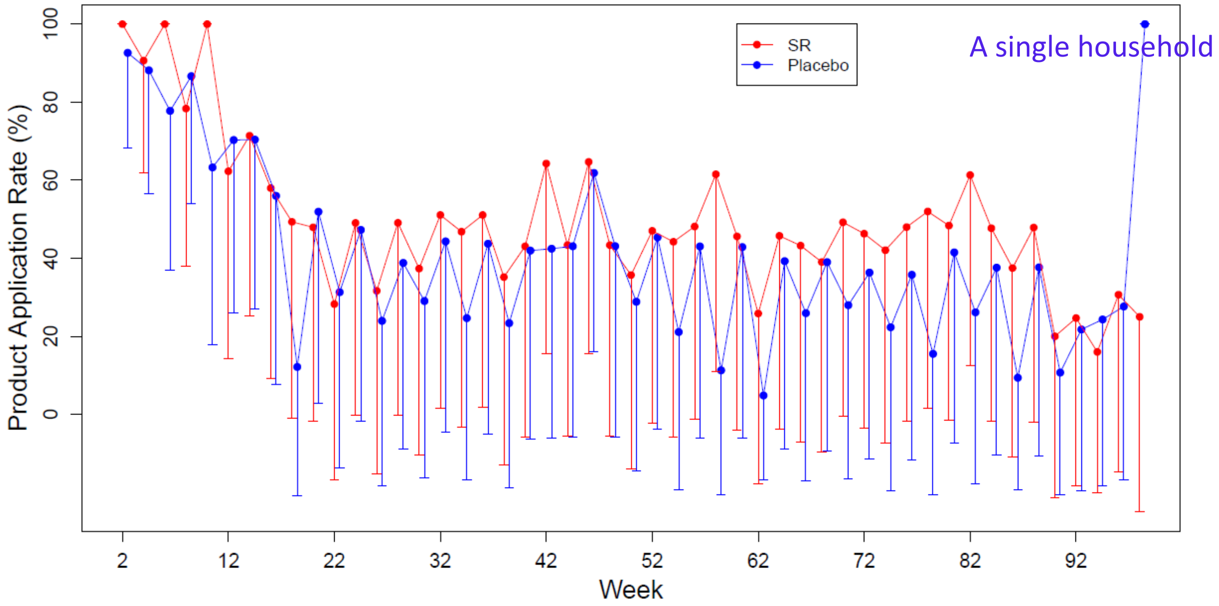


Table 26. Household Coverage and Attrition Rates

| Measure | Mali[2] | Formula |
|---|---|---|
| A. Household [Structure] Coverage | Avg. ~74% (41.1%, 98.9%) | No. enrolled structures with >1 product / No. structures in cluster (with + without participants) |
| B. Product Attrition Rate[3] | Avg. ~59% (17.6%, 99.9%) | No. products **removed** from each structure with product / No. products required in each structure every 28-days (with + without participants) |
| C. Coverage based on Product Attrition Rate | | |
| at 100% | Avg. ~24% (0%, 97.2%) | No. structures at 100% of req. product application rate / No. structures receiving product in cluster (with + without participants) |
| >75%[4] | Avg. ~44% (0%, 100%) | No. structures at >75% of req. product application rate / No. structures receiving product in cluster (with + without participants) |
| D. Coverage based on Product Attrition Rate | | |
| at 100% | Avg. ~19% (0%, 85.1%) | No. structures at 100% of req. product application rate / No. structures in cluster (with + without participants) |
| >75% | Avg. ~33% (0%, 92.6%) | No. structures at >75% of req. product application rate / No. structures in cluster (with + without participants) |

[1] Post-hoc analyses
[2] **SR distribution in Mali cRCT occurred only within compounds of an enrolled study participant. Remaining compounds without enrolled study participants were intentionally excluded from receiving product.**
[3] Dependent on room design and/or homeowners refusing product placement in spaces
[4] >75% threshold based on SR Peru cRCT analytical approach (Morrison et al. PNAS 2023)
