## Supplementary material for "Effect of a spatial repellent on malaria incidence in Mali: a cluster-randomized, controlled trial": Study Protocol

### S3. Study Protocol

This appendix has been provided by the authors to give readers additional information about their work.

CATHOLIC RELIEF SERVICES

### CLINICAL TRIAL PROTOCOL

**Title:** Spatial Repellent Products for Control of Vector Borne Diseases

**Protocol No: TBD**

| | |
|---|---|
| **University Notre Dame No.:** | **20-10-6245** |
| **Clinical Trials Registration No.:** | **NCT04795648** |
| **Brief Title:** | Spatial Repellents for Control of Vector Control |
| **Protocol Version – Date:** | February 2, 2022 v8.4 |
| **Sponsor:** | University of Notre Dame, Notre Dame, IN 46556 |
| **Principal Investigator:** | Suzanne Van Hulle |

### 1. SIGNATURE PAGE

The clinical trial will be carried out in accordance with the protocol, the DMID Good Clinical Practice Resource Guide and in accordance to local legal and regulatory requirements.

| Principal Investigator: | Signature: | Date: |
|---|---|---|
| ____________________ | ____________________ | ____________________ |
| Sponsor’s Representative: | Signature: | Date: |
| ____________________ | ____________________ | ____________________ |

#### 2. PROTOCOL AMENDMENT(S)

**Amendment #2:**

| Principal Investigator: | Signature: | Date: |
|---|---|---|
| ____________________ | ____________________ | ____________________ |
| Principal Investigator: | Signature: | Date: |
| ____________________ | ____________________ | ____________________ |

| Spatial Repellent Products for Control of Vector Borne Diseases | | |
|---|---|---|
| **Version** | **Date** | **Notes** |
| 8.1 | 29 Oct 2020 | VCAG Approved version – WHO ERC Peer reviewer comments integrated |
| 8.2 | 24 Jan 2021 | WHO ERC and Univ. of Bamako comments integrated |
| 8.3 | 10 Dec 2021 | Amendment submitted with WHO ERC Continuing Review – Updated product name from Envelope to Mosquito Shield™ |
| 8.4 | 01 Feb 2022 | Amendment for malaria infection treatment and hemoglobin testing during follow up. |

#### 4. KEY ROLES

For questions regarding this protocol, contact Suzanne Van Hulle,.

**Author(s):** Suzanne Van Hulle, Catholic Relief Services and John P. Grieco, Nicole L. Achee, Fang Liu, University of Notre Dame

**Sponsor's Representative:**
Eric Felde[3]
*Director of Research Compliance*
Notre Dame Research
940 Grace Hall University of Notre Dame
Notre Dame, IN 46556
E:
P: +1.574.631.1461

**Site Principal Investigator:** Issaka Sagara, MD, PhD, Malaria Research and Training Center (MRTC), Faculty of Medicine, Dentistry and Pharmacy at the University, of Sciences, Techniques and Technologies of Bamako (USTTB), Mali

**Sub-Investigator(s):**
Suzanne Van Hulle, Catholic Relief Services, Headquarters
Momar Mbodji, MD, MPH, Catholic Relief Services, Mali
Ghislain Ismael Nana, Catholic Relief Services, Headquarters
Mamadou Coulibaly, PharmD, PhD, Malaria Research and Training Center (MRTC), Faculty of Medicine, Dentistry and Pharmacy at the University, of Sciences, Techniques and Technologies of Bamako (USTTB), Mali
Alassane Dicko, MD, PhD, Malaria Research and Training Center (MRTC), Faculty of Medicine, Dentistry and Pharmacy at the University, of Sciences, Techniques and Technologies of Bamako (USTTB), Mali
John Hembling, Catholic Relief Services, Headquarters

**Sponsor Principal Investigator:** John P. Grieco **Sponsor Scientific Director:** Nicole L. Achee
**Sponsor's Medical Expert:** TBD
**Trial monitor(s):** fhiClinical **Local Safety Monitor:** TBD **Chair of DMC/DSMB:** TBD
**Statistician:** Fang Liu, University of Notre Dame
**External Adviser:** TBD
**Clinical Laboratory:** TBD

**Local Ethics Committee:** Ethical Committee of the Faculty of Medicine, Odonto- Stomatology and Pharmacy

#### 5. ABBREVIATIONS

| | |
|---|---|
| ACT | Artemisinin-combination treatment |
| AE | Adverse events |
| AI | Active ingredient |
| AL | Arthemether - Luméfantrine |
| CDC-LT | Centers for Disease Control and Prevention Light Traps Community |
| CHW | Health Workers/hygiene promoters |
| CI | Confidence interval |
| CNS | Central nervous system |
| CRF | Case Report Form |
| CRS | Catholic Relief Services |
| CV | Coefficient of variance |
| DSMB | Data safety management board |
| FMOS-FAPH | Faculté de Médecine et d'Odonto Stomatologie – Faculté de Pharmacie |
| HH | Households |
| HLC | Human Landing Catches |
| HR | Hazard ratio |
| ICF | Informed consent form |
| IRS | Indoor residual spraying |
| ITN | Insecticide treated nets |
| LLIN | Long-lasting insecticide nets |
| LTFU | Loss to follow up |
| MRTC | Malaria Research and Training Center |
| PE | Protective efficacy |
| RCT | Randomized Control Trial |
| RDT | Rapid diagnostic test |
| SAE | Serious adverse events |
| SR | Spatial repellents |
| SS | Sample size |
| TBD | To be determined |
| WHO | World Health Organization |

### 6. PROTOCOL SUMMARY

**Title:** Advancing Spatial Repellents for Vector-borne Disease Control

**Phase:** III

**Population:** Residents in selected households in Mali

**Number of participants:** 32 subjects and 30 clusters per arm in Mali (1,920 total subjects)

**Number of sites:** 1

**Location of site:** Kolondieba District, Sikasso Region, Mali

**Trial Duration:** 30 months
- **Clinical Phase:** 24 months
- **Whole Trial:** 30 months

**Duration for Participants:** 6-month baseline and 24-month follow up.

**Description of Investigational Products:** The SR will be a new formulation of transfluthrin. This active ingredient (AI) is widely used in mosquito coils and other household pest control products. The new formulation is a passive emanator that will release the AI over a period of up to four weeks. The emanator will consist of a pre-treated piece of cellulose acetate, which will be positioned within consenting households according to manufacturer specifications. The SRs and placebos for this study will be designed and manufactured by SC Johnson, Inc. USA.

**Objectives:** The primary objective of the study is to demonstrate and quantify the protective efficacy of a single SR product, in reducing malaria infection in a human cohort. The study design will be a prospective cluster Randomized Control Trial (RCT).

**Endpoints:** The primary epidemiological outcome measure of the SR trial will be microscopy confirmed malaria infection incidence (first-time infection). The secondary epidemiological outcome measure will be overall (first and subsequent) infection. The primary entomological measure will be human landing catch rate. Secondary entomological measures include sporozoite rate (to calculate EIR), and parity rate to determine if there are entomological correlates of SR efficacy that may be useful for the evaluation of new SRs.

**Description of Study Design:** Children ≥ 6 months to < 10 years of age will be enrolled in a single cohort across 30 clusters. The cohort will be followed for 6 months for baseline covariate measurements and 24 months with intervention. Blood samples will be taken once every 4 weeks from all cohort subjects to test for malaria infection and whenever a subject reports a recent history of fever (within previous 48h). RDTs will be used for point-of-care diagnosis of malaria infection with microscopy used to confirm infection status. All positive malaria infections as indicated by either RDT or microscopy, clinical and asymptomatic, will be treated throughout intervention follow up. Entomological measurements of SR effect will be conducted from a randomly selected subset of 12 clusters. To estimate the impact of the SR on mosquito density, light traps will be deployed next to a person sleeping under a net. To measure the impact of the SR on mosquito-human contact, human landing catches (HLC) will be performed once during the baseline phase and then quarterly during follow up with intervention. Mosquito collectors will be tested for malaria if they develop symptoms and treated if the result is positive.

#### 7. TITLE OF THE PROJECT

Advancing Spatial Repellents for Vector-borne Disease Control

#### 8. INVESTIGATORS AND INSTITUTIONAL AFFILIATIONS

Issaka Sagara, MD, PhD, Malaria Research and Training Center (MRTC), Faculty of Medicine, Dentistry and Pharmacy at the University, of Sciences, Techniques and Technologies of Bamako (USTTB), Mali
Suzanne Van Hulle, Catholic Relief Services, Headquarters Momar Mbodji, MD, MPH, Catholic Relief Services, Mali Ghislain Ismael Nana, Catholic Relief Services, Headquarters
Mamadou Coulibaly, PharmD, PhD, Malaria Research and Training Center (MRTC), Faculty of Medicine, Dentistry and Pharmacy at the University, of Sciences, Techniques and Technologies of Bamako (USTTB), Mali
Alassane Dicko, MD, PhD, Malaria Research and Training Center (MRTC), Faculty of Medicine, Dentistry and Pharmacy at the University, of Sciences, Techniques and Technologies of Bamako (USTTB), Mali
John Hembling, Catholic Relief Services, Headquarters

#### 9. ABSTRACT

Spatial repellents (SRs) have been widely used for the prevention of mosquito bites but their efficacy in reducing mosquito borne diseases has never been evaluated in Africa. Additionally, SRs have the potential of being critical tools in the prevention of mosquito borne diseases in contexts where typical vectors control efforts such as long-lasting insecticide nets (LLINs) and indoor residual spray (IRS) are inaccessible or underutilized such as among displaced peoples or in emergency relief settings. To address this knowledge gap, Kolondieba District, Sikasso Region, Mali was selected as a site to estimate the impact of the Mosquito Shield™, an SR that incorporates transfluthrin on a plastic sheet, on malaria related outcomes. Over the past decade, the Region of Sikasso, Health districts of Kadiolo, Yorosso and Kolondieba have remained among the most afflicted, characterized by an annual parasite incidence of more than 116 cases per 1000 population (SLIS, 2018) and a *Plasmodium falciparum* prevalence rate of 29.7% (DHS 2018). Children ≥ 6 months and adolescents < 10 years of age will be enrolled into a single cohort which will be followed for two years (24 months). A total of 1,920 subjects will be enrolled. While there is a risk of the SR product not providing any protective effect, some protection is expected based on previous studies in Peru and Indonesia using the same intervention in a 2-week formulation, and transfluthrin active ingredient. Based on this evidence, it is anticipated that protection will benefit all household members from both genders.

#### 10.INTRODUCTION/BACKGROUND

SRs are a promising new vector control paradigm that could add to the existing armamentarium for malaria prevention (Achee et al. 2012, Ogoma et al. 2012). SRs such as mosquito coils have been shown to reduce mosquito biting (Lucas et al. 2007, Kawada et al. 2008) and, in studies conducted in Indonesia (Syaffrudin, 2014) and China (Hill et al. 2014), reduce malaria transmission in human populations. Evidence is required, however, to show the effectiveness of SRs across a range of malaria transmission endemicities, a range of mosquito vector species, and across different contexts of insecticide treated net (ITN) coverage before SRs can be recommended as a tool for malaria prevention by health authorities. Despite evidence of SR efficacy in some regions, they have never been thoroughly evaluated in sub-Saharan Africa where the primary malaria vectors are strongly anthropophilic and the burden of malaria is highest.

Mali is one of the most under developed countries in the world, with nearly 65 percent of its inhabitants living in poverty. Although tremendous gains in malaria control have been made over the years, malaria still remains a major public health concern in the country with a number of regions still registering high transmission [DHS 2018]. Throughout the country, malaria remains the leading cause of morbidity and mortality, particularly among children under the age of five. Almost two and a half million clinical cases of malaria were reported in health facilities in 2015, accounting for more than a third of all outpatient visits for all age groups (DHS 2015). During the period of peak malaria transmission, the prevalence of malaria parasitemia among children aged 6-59 months was 32% based on rapid diagnostic tests (RDTs), and 36% based on microscopy (DHS 2015). Because of the high burden of malaria and the history of the CRS efforts in this region, the site was selected to evaluate the efficacy of a new SR product formulated to last up to four weeks for the prevention of malaria. This will be a cluster randomized control trial (RCT), whereby children ≥ 6 months to < 10 years of age will be enrolled into a single cohort and followed for 24 months with intervention. While there is a risk of the SR product not providing any protective effect, some protection is expected based on previous studies in Peru and Indonesia using the same intervention in a 2-week formulation, and transfluthrin active ingredient. Based on this evidence, it is anticipated that protection in this study will benefit all household members from both genders. All blood samples will be taken for microscopic confirmation of malaria infection for the measure of time to first-infection as well as the measure for overall, subsequent new infections. RDTs will be used for point-of-care diagnosis of malaria infection with microscopy used to confirm infection status. All positive malaria infections as indicated by either RDT or microscopy, clinical and asymptomatic, will be treated throughout intervention follow up. This evaluation will serve as an efficacy trial of SR products for sub-Saharan Africa. Findings will be submitted to the World Health Organization Vector Control Advisory Group (WHO VCAG) for assessment of whether SRs have "public health value." Entomological outcomes will also be measured as proxies of malaria transmission to help develop guidelines for the evaluation of future SR products.

### 11.JUSTIFICATION OF THE STUDY

Malaria is the primary cause of morbidity and mortality in Mali, particularly among children under five years of age. According to the National Health Statistic (SLIS) of 2018, the Mali health facilities have registered 2 345 481 cases of malaria among them 1001 deaths [SLIS 2018]. Malaria places a tremendous burden on Mali's health system as it accounts for about 37% of the motivation of the health facility consultation in Mali. Currently, the main malaria control interventions used in Mali are: malaria case management, use of the Insecticide Treated Net (ITN), Indoor Residual Spraying (although only in specific regions of the country and not in the proposed RCT site), Seasonal Malaria Chemoprevention (SMC) for children 3-59 months, and Intermittent Preventive Treatment in pregnant women (IPTp). Additional control measures are therefore needed to complement current interventions.

Children ≥ 6 months to < 10 years of age will be enrolled in a single cohort across 60 clusters (30 clusters per treatment arm). The cohort will be followed for 6 months for baseline covariate measurements and 24 months with intervention. Blood samples will be taken once every 4 weeks from all cohort subjects to test for malaria infection and whenever a subject reports a recent history of fever (within previous 48 hours). All blood samples will be taken for microscopic confirmation of malaria infection. RDTs will be used for point-of-care diagnosis of malaria infection with microscopy used to confirm infection status. All positive malaria infections as indicated by either RDT or microscopy, clinical and asymptomatic, will be treated throughout intervention follow up. Microscopy sample diagnosis will be used as the primary measurement for the primary (first-time infection) and secondary endpoint (overall new infections) analyses of malaria incidence. The incidence of malaria infection as measured by microscopy will be estimated and compared between treatment arms to determine the benefit of using an SR in an area with high, seasonal transmission of malaria. Entomological endpoints of exposure risk to mosquitoes will also be measured to identify entomological correlates of SR efficacy that may be useful for the evaluation of new SR products.

Despite the scale up of effective tools for the prevention and control of malaria, this disease remains one of the primary causes of morbidity and mortality in sub-Saharan Africa. New tools are needed to address the threat of insecticide resistance and outdoor biting vectors and to sustain the drive to elimination. SR products have shown promise as a tool to reduce biting by mosquitoes and the WHO Pesticide Evaluation Scheme has developed methods to evaluate the efficacy of new SR products. However, proof of principle of SRs as tools to reduce disease burden has only been shown under small-scale study designs in two locations, Indonesia (Syaffrudin, 2014) and China (Hill et al. 2014). In addition, assessments of an SR intervention delivered under programmatic capacity of health services has not been evaluated. It is therefore unlikely that efficacy estimates derived from tightly controlled phase III trials will be realized in programme settings. This study will address the knowledge gap of whether or not SRs are effective in reducing human malaria disease in humanitarian assistance and emergency response settings in sub-Saharan Africa where underlying transmission rates are high at baseline, ITNs may or may not be widely deployed, and will inform the policy makers on whether to recommend SRs as a means to further reduce malaria transmission.

#### 12.MAIN OBJECTIVE

The main objective of the study is to demonstrate and quantify the protective efficacy (PE) of a single SR product, in reducing malaria infection in children ≥ 6mo to < 10yrs of age in Mali under controlled conditions. The null hypothesis (H0) is that there is no difference in malaria incidence between intervention and control arms.

#### 13.SPECIFIC OBJECTIVES AND HYPOTHESES TO BE TESTED

**Primary Objective:**
To evaluate the PE of SRs against the first-time malaria infection in children ≥ 6mo to <10yrs of age in Mali.

**Secondary Objectives:**
Secondary objectives will address key issues related to the optimization and application of SR products for public health and confirm the range of contexts within which SR PE can be achieved. Secondary objectives are:

1. Confirming and measuring the entomological correlates of reduced infection (e.g., a reduction in vector densities, mosquito infection, biting and parity rates) to set benchmark thresholds and streamline future intervention trials against other SR products by measuring only those endpoints that are correlated to PE;
2. Quantifying the efficacy in an epidemiological setting with insecticide resistant vectors;

3. Quantifying the total number of infections averted using the spatial repellent intervention;

**Supplementary Objectives:**

1. To assess the relationship between the reduction in the malaria hazard rate and the decrease in mosquito CDC-LT density per night and other entomological endpoints (e.g., HLC, parity rate, and sporozoite positivity rate).
2. To evaluate the PE of SR for overall (total new) malaria infection, both first-time and recurrents.

### 14.PRIMARY OUTCOME MEASURE

The primary outcome measure will be microscopy confirmed malaria infection incidence from blood smears.

### 15.DESIGN AND METHODOLOGY

The study design will be a prospective cluster, randomized control trial (RCT)

*(a) Study site and population*

The trial will be conducted in Kolondieba District, Sikasso Region, Mali (**Figure 1**). Kolondieba district has some of the highest malaria transmission rates in the country with 205/1000 population in 2018 with one malaria transmission peak period from July to November (DHIS2 2018).

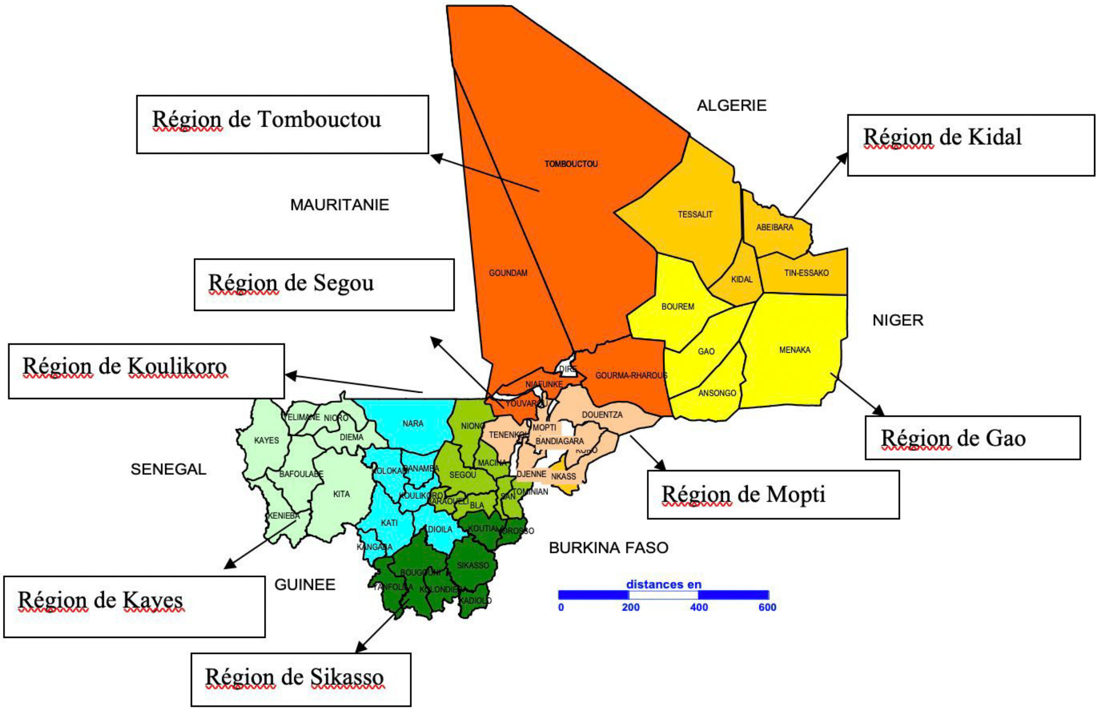

**Figure 1**:Map showing different districts and regions of Mali

The selected district of Kolondieba has 161 villages whose GIS mapping is available through Mali's INSTAT (collected in 2009). Based on a distance of 5km between villages to avoid clusters overlapping each other, a delineation of 60 clusters has been explored (**Figure 2**). There are a total of 16 health facilities in the study area with average distance to village/cluster groupings of 10km (Figure 1).

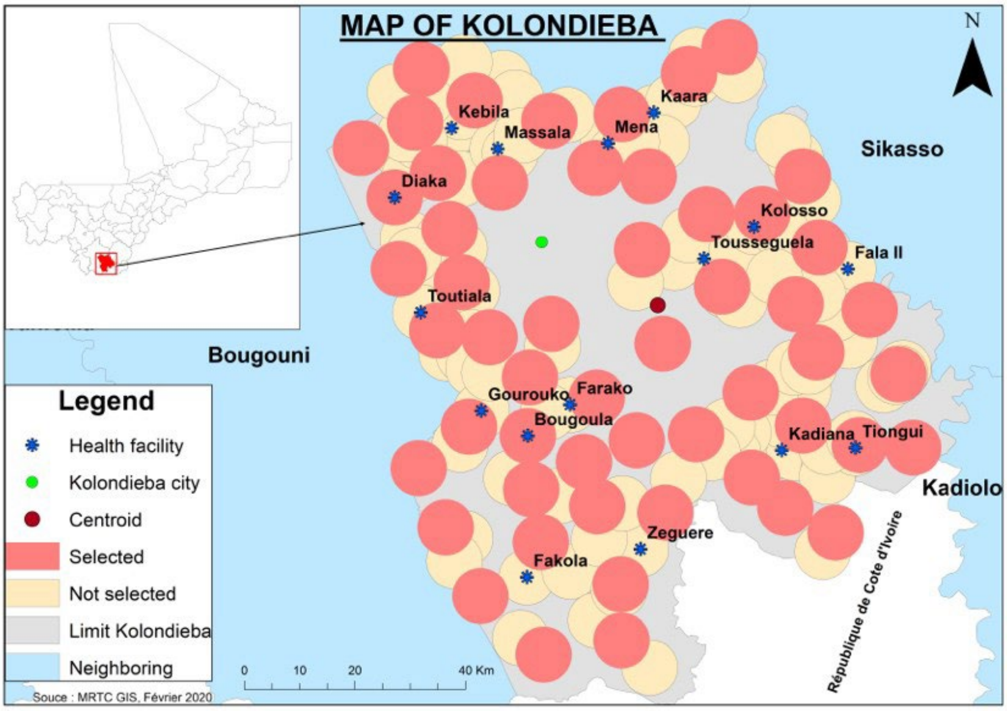

**Figure 2.** Tentative distribution of clusters and location of health facilities in the study area.

The investigative team will monitor ongoing malaria control interventions in the clusters during the trial period to help inform in potential biases across study clusters as well as potential augmented benefit of SR intervention to these strategies. Note too that all health interventions in the study area are under the responsibility of the District Chief Medical Officer who is involved in this research project. Thus, the research team will be aware of all the health interventions taking place in the area and will be able to harmonize data collection methods, since the same actors are involved.

Favorable conditions for vector propagation exist in terms of temperature, vegetation and surface water distribution with members from the 3 main malaria vector species complexes present (*An. gambiae*, *An. funestus*, and *An. coluzzii*). Wide-spread resistance to pyrethroid, organochlorines, carbamates and organophosphates has been indicated, particularly, to pyrethroids in the *An. gambiae* complex (Cisse et al, 2016, Keita et al, 2017, Fanello et al, 2003). In the general area considered for this study, resistance has been described (Tripet et al, 2007). This could be explained by the intensive use of insecticides in agriculture. The Sikasso Region has mixed ethnicities with mainly Bambara, Senoufo and Fulani. Housing construction is made predominately of traditional mud walls with grass or iron sheet roofing. There are limited interventions with main-stay malaria vector control methods (mainly bed net use) due to various health system challenges. Currently, there is no presence/planned engagement from other MOH partner Malaria control programs/projects therefore there are no competing trials that are planned for this district of Kolondieba.

*(b) Cohort Study Inclusion/Exclusion criteria*

Children ≥ 6 months to < 10 years of age who report sleeping in selected study clusters >90% of the nights of each month and do not have any plans to travel outside the study area will be eligible for inclusion in the study. Children who have a measured hemoglobin at baseline screening/enrollment, or re-screening/re-enrollment for intervention phase of ≤7 g/dL, or with known chronic disease or who have signs of clinical decompensation will also be excluded from the study. During follow up of enrolled subjects, study clinicians will have the option to conduct a Hb test for enrolled subjects when they may present signs of anemia to see if they might need additional treatment beyond malaria ACTs (if malaria infection is indicated).

Persons who are participating in another clinical trial investigating a drug, vaccine, medical device or procedure will also be excluded from the study. The written consent of the parent or legal guardian of each child is required for inclusion in the study.

Inclusion/exclusion criteria are below:

| Inclusion Criteria | Exclusion Criteria |
|---|---|
| Children ≥ 6 months to < 10 years of age | Children ≤ 6 months to > 10 years of age |
| Children with Hb ≥ 7g/dL and no signs of known chronic disease or other serious illness | Children with Hb ≤ 7g/dL with signs of known chronic disease or other serious illness, or Hb <6g/dL with signs of clinical decompensation |
| Sleeps in cluster >90% of nights during any given month | Sleeps in cluster <90% of nights during any given month |
| Not participating in another clinical trial investigating a vaccine, drug, medical device, or a medical procedure during the trial | Participating or planned participation in another clinical trial investigating a vaccine, drug, medical device, or a medical procedure during the trial |
| Provision of informed consent form (ICF) signed by the parent(s) or guardian | No provision of ICF signed by the parent(s) or guardian |

*(c) Randomization and Blinding*

The unit of randomization for the intervention and placebo will be a cluster. The study statistician will analyse data from the baseline follow-up period to inform on potential stratification requirements prior to randomization. Criteria for stratification will be baseline malaria incidence levels and/or adult entomological endpoints. Following stratification (as needed), clusters will be allocated to receive either active or placebo treatment using a random number generator (https://www.random.org). Following cluster treatment allocation, a simple random sample will be completed for the selection of sub-clusters assigned for entomology collection. The cluster allocation code will be made available from the intervention manufacturer to the Data Safety Monitoring Board (DSMB) for use in safety assessments. The site database manager will assign a unique identification number to each household (HIN) and the site intervention administrator will coordinate distribution of blinded active or placebo to enrolled households within each cluster corresponding to the pre-labeled package code. Unblinded assignments will be shared with a site administrator in a sealed envelope placed in a secure location for purposes of emergency unblinding related to adverse and/or serious adverse events. Thus, the investigators, research team, study subjects, and residents will be blinded as to which cluster receives active versus placebo devices until after completion of the study.

*(d) Data Safety Monitoring Board (DSMB)*

The DSMB will be responsible for the review of safety data from the RCT on an ongoing basis in order to monitor and rapidly identify any accumulating safety issues from across the program. The

DSMB will provide additional credibility about study quality, by reviewing regular (summary) reports from Principal Investigators during baseline and intervention periods and making recommendations as needed about study adjustment for study quality reasons. The DSMB can recommend to the Primary UND to stop or alter the trials in the event there is evidence of harm. As appropriate, the DSMB will provide updates to the WHO. The DSMB will be comprised of Biostatistician, a Clinician and an Epidemiologist with clinical trial experience.

Specific areas of review include:

a) Safety data should be reviewed routinely and regularly by the DSMB Medical Monitor. If significant concern is raised, he/she can engage with the committee, which can recommend that the trial be stopped either at a particular site or altogether.
b) Summary of adverse events (AE), severe adverse events (SAE) and death reports observed during the studies should be reviewed by the entire committee at pre-determined checks (quarterly). This should include comparison of the rate of AE and SAE in the two study arms and at the individual study site. The DSMB will be notified of any SAEs that are 'at least possibly related' to the research study as they are reported to Principal Investigators.
c) Unblinded efficacy data will be analyzed according to a pre-defined Statistical Analyses Plan (SAP) by an Independent Statistician when pre-defined enrolment targets have been achieved. The role of the DSMB statistician will be to agree to what information will be reviewed (see below) and then review and interpret this information with the other DSMB members, perhaps to request further analysis, for example. The DSMB statistician will contribute input to program Principal Investigators as to what subset of the SAP is to be presented at different meetings.

*(e) Adverse Events (AE) and Severe Adverse Events (SAE) definitions and reporting*

An adverse event includes any noxious, pathological or unintended change in anatomical, physiological or metabolic functions as indicated by physical signs, symptoms and/or laboratory detected changes occurring in any phase of the clinical study whether associated with the study intervention or placebo. This definition includes an exacerbation of pre-existing conditions or events, intercurrent illnesses. A serious adverse event is any untoward medical occurrence that results in death, is life threatening, results in persistent or significant disability/incapacity, requires in-patient hospitalization or prolongation of existing hospitalization or is a congenital anomaly/birth defect in the offspring of a study subject. In addition, important medical events that may jeopardize the participant or may require intervention to prevent one of the other outcomes listed above will be considered serious.

All serious adverse events will be:

- Recorded on the appropriate serious adverse event case report form
- Followed through resolution by a study physician
- Reviewed by a study physician

Symptomatic uncomplicated or severe malaria infection will be coded as adverse events as will any other acute illness, throughout the 24 months of the study. Clinical malaria will be recorded in the same fashion as other adverse events. In general, uncomplicated malaria will constitute an AE and severe malaria will constitute an SAE. Anticipated day-to-day fluctuations of pre-existing conditions that do not represent a clinically significant exacerbation need not be considered adverse events. Discrete episodes of chronic conditions occurring during a study period will be reported as AEs to assess changes in frequency or severity. Pre-existing conditions or signs and/or symptoms (including any that are not recognized at study entry but are recognized during the study period) present in a participant prior to the start of the study will be recorded on the participant's Case Report Form (CRF).

AEs will be documented in terms of a medical diagnosis. When it is not possible to make a specific medical diagnosis, the adverse event will be documented in terms of signs and/or symptoms observed by the investigator or reported by the subject at each study visit. Any hospitalization will be considered a serious adverse event.

AEs to be recorded as endpoints will be pre-defined based on SC Johnson, Inc. toxicology reports of 'probable', 'possible', 'plausible' and 'unlikely' **(Appendix 0. Investigator's Brochure):**

- **Probable**: sensory irritation (oral and dermal)
- **Possible**: nausea/vomiting (oral), skin irritation/rash (dermal), runny nose (inhalation)
- **Plausible**: salivation
- **Unlikely**: eye irritation, headache

AEs and SAEs will be recorded throughout the study. Serious adverse events and the study reports will also be provided to Mali's Faculté de Médecine et d'Odonto Stomatologie - Faculté de Pharmacie (FMOS-FAPH) ethical committee as required by their current Standard Operating Procedures. Study teams will record AEs for every 2-week surveillance period at time of house visit (during active malaria detection) and for the 30- day surveillance period at time of clinic visits (during passive malaria detection). SAEs will be recorded at time of death, hospitalization. Assessment of SAE relatedness to intervention will be conducted by the study clinician in-country with confirmation/verification by the DSMB Medical Monitor based on the study clinician SAE report and trends in other SAE datasets.

*(f) Procedures*

<u>Community sensitization and informed consent</u>

Information regarding the study will be distributed through study personnel and CHWs, and other MRTC/CRS staff, targeting parent(s)/ guardian(s) of potential participants in the community, community leaders, and organizations working in the district. Information will be provided through some of the following means:

- Community meetings will be held at a local venue and study staff will present the study and have discussions with the community;
- Radio spots

The study will be explained and consent form read in the local language to the head of household or spouse (for SR or placebo application in the home) (**Appendix 3(i): Informed Consent to Recieve SR Intervention**), or to the parent/guardian of children eligible to participate in the cohort (**Appendix 1 and 2: Informed Consent for Cohort Participation**). The study will be explained in local dialect and time allowed for questions to be answered. Once all concerns are addressed, ICFs written in French (the official language in Mali) will be signed or a thumb print taken (either on paper or electronically).

- Informed consent will take place with two study populations. The study will be conducted at the cluster scale (first population) and all households (HHs) within the selected clusters will be eligible to receive the SR product (second population). Therefore, all heads of HHs in the selected clusters will be approached and be read an ICF. Before being asked to sign the ICF, they will be asked to confirm their understanding of the ICF, to have the SR product placed in their houses and replaced every 4 weeks **(Appendix 3(i). Informed Consent to Receive SR Intervention).** The ICF will also ask that they report to the study clinic if they experience any adverse events (AEs) or serious adverse events (SAEs) of interest. AEs and SAEs of interest will be described verbally and will be provided in written form. Secondly, a cohort of children and adolscents will be enrolled to estimate the impact of the SR product on the incidence of malaria disease (SR and placebo clusters). Parents/guardians of eligible children selected to participate in the cohort will be asked to provide consent. An attempt to obtain written, informed consent from both parents of the child will be made, but consent from only one parent will be required for participation (national protocols will be observed).

- Separately, parent(s) and guardian(s) of eligible children aged 6 months to 10 years will be asked to provide consent to participate in the following cohorts:
    - Baseline cohort (6 months duration)
    - Follow up cohorts (24 months in duration for each)
- Children aged between 6 months to 10 years old in identified households will be approached by the study team, to explain the study and ask them to voluntarily participate in the study.
- If subjects' consent to participate, the study team will screen eligible children.
- After screening children for eligibility, the children will be enrolled
- Eligible children will then be tested and treated for malaria free of charge during the course of the study.

Parent(s)/guardian(s) who cannot sign their names will provide a thumb print on the consent form for documentation of willingness to participate, and a witness not associated with the study will sign the consent form indicating that the ICF was read, and that participation and thumb- printing were given willingly without coercion. Time will be granted to those parent(s)/ guardian(s) who would wish to make consultations with their family members before signing. It will be stressed to all parents/guardians approached that their children's entry into the study is voluntary and they may withdraw from the study at any time for any reason without any penalty. In HHs with multiple children that meet the study inclusion criteria, all eligible children will be invited to participate. Consented subjects will be assigned a subject identity code (or ID card) on enrollment. Consented participants will be screened for inclusion/exclusion criteria.

For participants who are illiterate, the following procedures will be conducted:

- The staff member will ensure whenever an illiterate potential subject is consented, a literate impartial witness (a family or community member of the parent/guardians' choice) who is not affiliated with the study is present during the informed consent process.
    - The witness must be present during the entire consent process
    - The language of ICF used should be the one best understood by both potential subject or guardian/Legally Authorized Representative (LAR) and the witness.
- The staff member will read the informed consent form, pausing frequently to allow the potential subject, guardian/LAR or witness to ask questions.
- The witness will be asked to verify to the best of his/her knowledge, the potential subject understood and agreed with the information provided during the consenting process and all the questions or concerns were addressed satisfactorily.
- The potential subject or guardian/LAR will be directed by the staff on how to put his/her thumbprint on the appropriate box (both copies). The witness will be verbally instructed

to write the name and date for the participant. The witness will then be instructed to write his/her name, sign and date the consent forms in the appropriate spaces provided for the witness.

Mapping of the study area and baseline census measurements

Prior to enrollment, all structures in the study area will be mapped using GPS coordinates and assigned a unique Household Identification Number. A baseline questionnaire will be administered at the same time as the mapping exercise to measure housing structure and household demographic characteristics. Profiles of enrolled houses that could potentially confound effect on mosquitoes will be generated. This includes house construction, socioeconomic status, number of inhabitants and their age, current household method(s) used to prevent mosquito bites, including ITNs.

Villages will be considered as the unit of randomization. According to Mali's 2009 General Population Census (le *Recensement General de la Population et de l'Habitat du Mali (RGPH)* 2009, the circle of Kolondieba is comprised of 11 rural communes with a population size of 201,456 inhabitants. This population is divided between 30,079 households and 14,730 family compounds, where a compound is defined as a family unit with one or more structures. A typical family compound has 5 household members.

Clusters will be randomly allocated to intervention and control using standard statistical software (e.g. SAS, STATA or R). All households in each village will be invited to participate in the study by allowing study staff to place SR or placebos in their household. SR or placebo will be placed in each household that consents.

Within each cluster, individual compounds will be enumerated and then randomly selected for inclusion in each cohort.

Screening and enrollment of the cohort

A cohort of children aged ≥ 6 months to < 10 years of age will be screened and enrolled within study clusters to assess the PE effect of the SR product against placebo control. HHs will be randomly selected from the master list of HHs obtained during baseline mapping to achieve the desired sample size (**Appendix 4. Statistical Analyses Plan**).

Selected HHs based on cluster maps will be visited and if a child is present, the study will be explained and consent obtained from the child's primary caregiver (as needed) before screening the child to verify inclusion criteria for enrollment into the study. All eligible children in a selected household will be invited to participate.

At screening for baseline, and re-screening for intervention phase, the age and gender of the child will be recorded and the parent/guardian (as needed) will be asked about involvement in other clinical trials, travel outside the cluster area and the use of ITNs and anti-malarial drugs. Contact information, as available, will be captured. A fingerstick blood sample will be taken for a malaria RDT, a blood smear, and a measurement of hemoglobin.

All children successfully meeting screening inclusion criteria and enrolled into the cohort will be provided a long-lasting insecticidal net (LLIN). This will allow us to measure the added benefit of SR product above that provided by currently recommended preventive measures. In addition, at baseline, and start of intervention phase subjects enrolled into the cohort will be provided a treatment dose of artemisinin-combination treatment (ACT), artemether and lumefantrine, according to national protocol, to clear any prepatent or patent malaria parasites.

RDTs will be used for point-of-care diagnosis of malaria infection with microscopy used to confirm infection status. All positive malaria infections as indicated by either RDT or microscopy, clinical and asymptomatic, will be treated. If a subject has a RDT negative outcome but a positive microscopy diagnosis, follow up treatment for the malaria infection will be provided to the subject within 72hrs of the microscopy read.

The enrollment and pre-product introduction period (baseline) will take approximately 6 months, during which time incidence of malaria infection will be measured prior to the deployment of the study product.

Follow up of the cohort

All efforts to overlay commencement of baseline follow-up with start of malaria transmission season will occur; however, programmatic timelines may not allow for perfect alignment. The primary goal during baseline will be to ensure capturing incidence across a range of anticipated transmission timing in order to inform on sample size assumptions prior to intervention.

At enrollment, children will be cleared of parasitemia with a treatment dose of ACT, artemether and lumefantrine (AL), just after deployment of the study product. This will be done so that incidence of malaria parasitemia following study product deployment can be measured. The distribution of the SR will occur after the completion of the baseline period. At this time, cohort participants will be presumptively cleared of parasites with a treatment dose of AL unless they have recently been treated (within the last 2 weeks). This will be done so that incidence of malaria infection following study product deployment can be measured.

During the entire study period, each subject will be asked to come to the nearest study health facility for a total of 30 scheduled clinic follow-up visits for passive malaria case detection. Scheduled clinic visits will occur one time per month during each the 6-month baseline period and 24-month period with intervention. Subjects will be visited at their home if they cannot reach the clinic for the scheduled follow-up. In addition, each subject will have scheduled household visits conducted by study staff for active malaria case detection where blood sampling will occur only for those subjects reporting fever history. Scheduled household visits will occur one time per month during each the 6-month baseline period and 24-month period with intervention. Lastly, subjects will be informed to make an unscheduled clinic visit any time during the trial should they experience malaria-related symptoms.

At each visit, the parent/guardian will be asked about recent use of ITNs and other vector control interventions as well as recent history of child's illness, recent travels outside the household, and recent use of anti-malarial drugs. At every other visit (on a monthly basis), a blood sample will be taken for malaria RDT, and a blood smear for confirming malaria diagnosis. At the intervening visits, a blood sample will only be taken if the child has a recent history of fever. Total blood volume for samples at each visit will not exceed 500 microliters.

During follow up of enrolled subjects, study clinicians will have the option to conduct a Hb test for enrolled subjects when they may present signs of anemia to see if they might need additional treatment beyond malaria ACTs (if malaria infection is indicated).

RDTs will be used for point-of-care diagnosis of malaria infection with microscopy used to confirm infection status. All positive malaria infections as indicated by either RDT or microscopy, clinical and asymptomatic, will be treated. If a subject has a RDT negative outcome but a positive microscopy diagnosis, follow up treatment for the malaria infection will be provided to the subject within 72hrs of the microscopy read.

Cohort subjects who test positive for malaria by either RDT or microscopy, symptomatic or asymptomatic, during both scheduled and unscheduled visits will be treated with ACTs free of charge according to national treatment guidelines. Hospital fees will not be paid by the study unless the illness or injury is due to study product or procedures as determined by a study clinician. If there is illness or injury due to study product or procedures, fees will be paid for care at the government clinic or District or Provincial Hospital according to the clinical trial insurance policy of the trial.

Enrollment to receive the SR product

The study product will be explained to the head of household, as described in the main protocol, and consent for participation will be obtained prior to participation. The head of household may be asked questions about their perceptions and acceptability of the product after the product has been deployed. No routine blood samples will be sought.

Application of the SR

The SR intervention will be a new formulation of transfluthrin. This active ingredient (AI) is widely used in mosquito coils and other household pest control products. The new formulation is a passive emanator that will release the AI over a period of up to four weeks. The emanator will consist of a pre-treated piece of cardstock, cellulose acetate or other medium, which will be positioned within consenting HHs by dedicated, trained study personnel according to manufacturer specifications (**Appendix 0. Investigator Brochure**). More than one emanator may be placed in a single structure depending on the size of the structure.

The SR product will be replaced by paid study personnel throughout the intervention period. Both participants and study staff will be blinded as to whether the product has the SR or is a placebo. Product will be replaced every four-weeks as recommended by the product manufacturer. Study teams will be employed to ensure proper storage of unused product at site and supply management and coordination for the timely replacement of products. Study teams will also perform periodic, unannounced spot checks in a random 10% of enrolled HHs during intervention on a quarterly basis to monitor SR product compliance (installation according to manufacturer specifications) and LLIN usage, if available. Each SR product and placebo will have a unique code, which will be recorded at the time of installation and the time of replacement. Both participants and study staff will be blinded as to whether the product has the SR or is a placebo.

Entomological monitoring

Twenty clusters (10 SR, 10 placebo) will be randomly selected to estimate the impact of the SR on entomological measures of malaria transmission. Within each cluster, light trap collections will be conducted monthly in 10 randomly selected households to assess the impact of SRs on the density of Anopheles mosquitoes indoors. The light traps will be deployed next to a person sleeping under a net and will run from approximately 5pm to 7am. The 10 SR and 10 placebo clusters will be randomly selected at the beginning of the study and will remain fixed throughout the study. Households will be sampled with replacement and therefore will be eligible for repeated sampling throughout the study. Households enrolled in the cohort study will not be eligible for mosquito sampling.

Human landing catches will be done indoors and outdoors in 6 intervention and 6 control clusters (the 12 clusters will remain fixed throughout the study) in four houses (randomly selected) in each cluster for the period of 2 nights (total of 48 houses across both arms) once every quarter (3 months) to determine the effect of SR on the host seeking behavior of mosquitoes. The same clusters and the houses will be sampled each sampling period. SR and placebo clusters will be selected in pairs to reduce the degree of heterogeneity between two arms. Sentinel houses will be chosen such that they are balanced as to house design and inhabitants to reduce bias due to mosquito density heterogeneity (i.e., some houses may be more attractive to mosquitoes than others). Households with children enrolled in the cohort are not eligible for the human landing catches. One pair of clusters will be sampled on any given night. Collections will start at 5pm each night with one person inside and one person outside. The teams will collect for 45 minutes each hour and then take a 15-minute break before starting the next hour of collection. At midnight, a new team will begin collecting and will continue until 7am. Mosquitoes will be sorted by site, date and time of collection. All mosquitoes will be identified to species and all female anopheline mosquitoes will be tested for the presence of sporozoites by ELISA and/or PCR. A subset of up to 30 per cluster per sampling period will be dissected for parity.

A team of 16 collectors will be hired to conduct the indoor/outdoor landing catches in 4 houses in each of 12 clusters (6 intervention and 6 control) every quarter following informed consent (**Appendix 5: Informed Consent for Human Landing Catches**). Mosquito collectors will be tested for malaria if they develop symptoms and treated according to national policy if the result is positive.

Monitoring for insecticide resistance will be done once during pre-intervention/baseline, mid-intervention (12 months) and post-intervention (within 3 months following last subject follow- up) in 4 intervention and 4 control clusters (selected by the independent statistician, blinded to investigators) spread across the study area. These data will not be used to assess the impact insecticide resistance would have on the intervention but rather to inform on the general status of insecticide resistance in the area. In each site, larvae or adults will be collected and reared in the insectary at a MRTC facility. Unfed adult females that are 2-5 days old will be tested in the WHO tube bioassay technique against permethrin to characterize resistance status. CDC bottle bioassays will be conducted, as appropriate, to assess transfluthrin susceptibility as well as permethrin resistance intensity at 1X, 2X, 5X and 10X diagnostic dose.

Temperature and relative humidity will be recorded from inside and outdoors of households enrolled in entomological sampling using Hobo data logging devices. Precipitation (rainfall) data from national weather stations will also be explored for clusters within the entomology sampling frame. Aggregation of precipitation data for entomology clusters will be conducted by treatment arm, as needed, based on limitations of geographical availability.

Monitoring for AEs. AEs and SAEs of interest (**Appendix 0. Investigators Brochure**) of enrolled subjects in the cohort and on other household members who receive the study product will be collected through prospective active surveillance at time of removal of SR product, or every 4 weeks. Anyone experiencing AEs of interest will be encouraged to seek care for at the study clinic for clinical assessment. Note that pregnancy in itself will not be regarded as an AE.

AEs of interest considered at least “possibly related” to the product will be reported to the University of Notre Dame and the Data Safety Management Board (DSMB) on a quarterly basis.

Unexpected SAEs determined to be at least "possibly related" to the product will be reported to the University of Notre Dame and the DSMB, and to local Ethical Review Committee(s) within 24 hours of the site PI becoming aware of the SAE, or according to each IRB reporting requirements. The initial report will be a short description by email, which will be followed within 7 days by a more detailed description of the SAE.

Monitoring staff
The primary sponsor (University of Notre Dame) has contracted for clinical trial oversight by fhiClinical.

Medical staff
Study participants will be followed up at home by study personnel. Care will be provided at the study clinics by physicians or clinical officers as patients routinely receive care from clinical officers at health facilities and hospitals.

Laboratory procedures
Blood samples will be processed at study clinics using RDTs for point of care (SD Bioline, antigen detection dipstick assays).

*(g) Statistical Analysis Plan*

The statistical methods are outlined in the statistical analyses plan (**Appendix 4. Statistical Analyses Plan)**.

Primary objective
The primary objective on the quantification of PE will be examined by comparing the hazard rates of the first-time malaria infection between SR and control. The complementary log-log (cloglog) model $\log(-\log(1-\theta_{kjit})) = \beta_{0t} + x_{kji}^T\beta_1 + z_k + z_{j(k)}$. $\theta_{kjit}$ is the discrete time hazard rate of subject $i$ from household $j$ in cluster $k$ at time $t$, and $x_{kji}$ contains time (as a categorical variable), the individual-, household- and cluster-level factors that include the treatment group (SR and control) and other relevant covariates (e.g., age, gender, household type, etc). First-order interactions terms will also be included in the model if deemed scientifically or statistically relevant. $z_k \sim N(0,\sigma_1^2)$ and $z_{j(k)} \sim N(0,\sigma_2^2)$ are
1 2 3
the random effects at the cluster, household and individual levels respectively. $z_{k(k)}$ will be only necessary if some HHs contribute more than one individuals to the study.

The null hypothesis of PE = 0% is equivalent to $\beta = 0$, which will be tested by the Wald's test $z = \hat{\beta}/s$ where $ss$ is the estimated standard error of $\hat{\beta}$. PE is estimated $1 - exp\ (\hat{\beta})$ ) ×100%, where $\hat{\beta}$ is the estimated regression coefficient associated with SR-reference compared to the control, and $\exp(\hat{\beta})$ is the estimated hazard ratio (HR) between SR-reference and the control. The Wald-type 95% confidence interval (CI) will also be provided along with the point estimate (1 – $\exp(\hat{\beta})$) ×100%.

Secondary objectives
The same model as outlined above for the primary objective will be also applied to analyze the PE of SR against overall new malaria infections (all malaria events from each individual are included, not just the first-time infection).

The mosquito density per night is defined as the average number of mosquitos captured in a CDC-LT dring a 12-hour period). An appropriate statistical model will be identified after examining the distributional characteristics of the data, which is likely to be a (zero-inflated) Poisson distribution, a (zero-inflated) negative binomial distribution if there is over-dispersion, or a log-normal distribution. The covariates will include the fixed effects of treatment group, time, relevant cluster-level and household-level information, and a random effect for cluster. Statistically significant and relevant interaction terms will also be included in the model. The ratio between SR and blank in CDC-LT density will be estimated, and the reduction in CDC- LT by SR is given by (1-CDC-LT ratio)×100%.

To explore the relationship between the epidemiological and entomological endpoints, a similar model as the cloglog model as the cloglog models used to address the primary objective on the first-time malaria infection will be applied to the epidemiological and entomological data in the clusters from which the entomological data are collected. The random effects and the individual-level and, household-level covariates will the same as the cloglog models specified above, and cluster-level covariate will include the baseline incidence rate, cluster population size, and the log-transformed average CDC-LT density in a cluster (averaged across the examined HHs at each visit time point). The regression coefficient associated with the log transformed CDC-LT quantifies the change in the hazard rate on the log scale, given one unit decrease in CDC-LT on the log-scale. Relationship between the malaria hazard rate and other entomological endpoints (e.g. parity rate, sporozoite positivity rate) will also be investigated.

Interim analysis

No formal interim analysis will be performed in this study.

*(h) Sample size dermination*

The sample size determination is based on the following parameter specifications: 1-sided type- I error rate = 5%, true PE = 30%, a between-cluster coefficient of variance (CV) of hazard rate = 47% (based on the historical data collected from Mali) then 788 independent first-time malaria events will need to be observed to reach 80% power in testing the primary hypothesis on PE.

With a baseline first-time malaria infection hazard rate of 1.0 per person-year (ppy), 30 clusters per treatment, 32 households per cluster (factoring in a loss to follow-up rate at 35%) in each treatment arm post randomization are expected to yield 788 independent first-time malaria events within 24 months follow-up period per cohort post randomization to yield 80% power.

***Note:*** *the SS might be adjusted based on the data collected during the 6-month baseline period. The SS recalculation is only affected by the baseline data (baseline incidence and CV), there is no risk of Type-I error rate inflation.*

#### 16.DATA MANAGEMENT

A combination of standardized paper-based or digital forms (under Android tablets) will be used across countries. Sharing common data models and protocols (indicators and variable definitions) across countries and mapping country-specific codes, enables the capability of cross-referencing and aggregating data for interim and final analysis.

CRS and UND/CRC will work together to develop the quantitative forms to be uploaded on Android phones with input from MRTC. All data issuing from the electronic data collection system will follow the same data collection processes outlined in the paragraph below. Any changes to quantitative data forms will need to be agreed by the RCT overall CRS PI (in consultation with CRS' technical advisors), the country level MRTC and CRS PIs, and UND's co-PI. In the event that a consensus on proposed changes cannot be achieved, UND's PI will make the final decision.

Any data collected on paper forms (including consent forms) will be scanned and transferred to binders for storage in a secure and locked restricted access area, while all electronic captured data will be archived with a documented history of changes or corrections at the local study site. Using CommCare, MRTC (with technical support from CRS) will collect data which will be securely stored on Android devices and then synchronized to CommCare cloud at least every 14 days. MRTC will download the data from CommCare and conduct initial data cleaning and de-identification, in collaboration with CRS, before syncing it to UND's central study database. The downloaded data will be stored on password-protected computers at the MRTC office to ensure confidentiality. Data from MRTC to University of Notre Dame will be transferred through a dedicated secure sFTP server with password protect access from CRS.

A password protected central study database warehousing data from all sites will be developed and managed by the University of Notre Dame and serve as a data repository and utilized for safe and confidential data storage, extraction, integration and analysis. The data warehouse and file repository will be backed up weekly at the local server level to ease recovery as needed. In addition, data is stored and backed-up on CommCare cloud. Access to study data is controlled through centralized administration and access will be granted only through the PI's permission. Research records for all study subjects including history and physical findings, and results of consultations are to be maintained by the local site PI in a secure storage facility and by the University of Notre Dame and CRS, for a minimum of 3 years after the end of the projects per national guidance or until notified by grantee. Data and samples can be destroyed at any given time after those 3 years.

#### 17.PROJECT MANAGEMENT

**CRS and MRTC** field teams are responsible for in-country study implementation and local oversight. They will work hand in hand to conduct this RCT in Kolondieba District. The **University of Notre Dame** is the study sponsor responsible for overall program management and reporting as well as data and statistical analysis and **FHI Clinical** provides the clinical trial oversight. The **DSMB** provides safety monitoring and oversight and **SC Johnson** oversees product registration, manufacturing, packaging, and shipping.

<u>CRS will have the following staff working either partial or full time on the RCT:</u>
**Principal Investigator / Senior Malaria Advisor**, 20% LOE, Washington DC, USA
The Principal Investigator for CRS will work closely with the CRS Project Director in Mali and MRTC staff. S/he is responsible for ensuring that the RCT is conducted respecting the approved protocol. S/he provides both technical and management support to the Project Director and MRTC PI, as needed.

**Malaria Technical Advisor**, 25% LOE, Washington DC, USA Specialized in monitoring evaluation and research, the malaria technical advisor provides technical support to the CRS team in day to day activities pertaining to ensuring application of protocol and research activities.

**Senior Technical Advisor – Health Evaluation & Research**, 10% LOE, Chicago, USA Provides oversight and research support to PI and Project Director in Mali, as requested.

**Project Director**, 70% LOE, based in Bamako, Mali
The Project Director will be the main point of contact in CRS Mali country office, liaising with MRTC and has overall responsibility for the success of the RCT. S/he will be responsible for day to day management of all aspects of the project in Mali. S/he will manage and coordinate the implementation of all activities throughout the study - work plan design, project start-up, implementation and close-out - to enable efficient and effective implementation in line with CRS quality principles and standards, donor requirements and good practices.

**MEAL Manager**, 100% LOE, based in Bamako, Mali
Reporting to the Project Director, the Monitoring Evaluation, Accountability & Learning (MEAL) Manager will lead the overall monitoring and evaluation component of the project and will work with the MRTC Data Manager to ensure consistency with the global performance monitoring system. S/he will facilitate the achievement of the Project objectives by coordinating all the MEAL activities of the project, providing technical directives and advice related to MEAL to the staff and to the implementing partner

**Product Supply chain officer**, 100% LOE, based in Bamako, Mali
The Product supply chain officer is in charge to coordinate and support consistency and compliance of warehousing facilities, commodity movements and distribution practices. S/he will effectively monitor commodity activities and identify deficiencies and means for improvement.

**Three (3) Field supervisors,** 100% LOE, based in Bamako, Mali
Reporting to the Project Director, the field supervisors (3) will support implementation of activities in the field. They will lead efforts to ensure availability of all items needed in the implementation areas, support placement of SR in houses, monitor implementation at field level, as well as the development of social- behavior change communication (SBCC) activities. 3 field supervisors to be recruited; each field supervisor will cover 5 health facilities (Cscoms) and roughly 20 clusters.

<u>**MRTC** will have the following staff working either partial or full time on the RCT:</u>

**Principal Investigator, 25% LOE, based in Bamako, Mali**
The principal investigator will be responsible for the overall management of all aspects of the project. S/he is accountable for a high-quality program delivery by supporting activities and implementers in the field through regular coordination and relationship with all stakeholders including MOH, NMCP, WHO. S/he is responsible for ensuring that the clinical investigation is conducted according to the signed agreement, the investigational plan, and regulations in Mali. S/he will make sure that the rights, safety, and welfare of subjects under the investigation are protected according to standard norms. S/he is also responsible for controlling the Mosquito Shield™ (SR) under investigation.

**Study coordinator, 100% LOE, based in Bamako, Mali**
The study coordinator will be responsible for day to day management of all aspects of the project in the field.

**Senior Epidemiologist, 20% LOE, based in Bamako, Mali**
The epidemiologist will provide day to day oversight on study design, implementation and

management of epidemiological aspects of the study.

**Senior Entomologist, 25% LOE, based in Bamako, Mali**
The entomologist will provide day to day oversight on study design, implementation and management of entomological and epidemiological aspects of the study.

**Four (4) Entomology technicians, 25% LOE, based in Bamako, Mali**
The entomology technicians will be responsible for the collection and processing of mosquitoes.

**Data Manager, 20% LOE, based in Bamako, Mali**
The data manager will be responsible for designing data entry screens, cleaning data and providing interim data summaries.

**Six (6) Data collectors, 100% LOE, based in Kolondieba, Mali**
Data collectors will be collecting data via electronic devices during baseline and 24 months of follow-up.

**Fifteen (15) Lab technicians, 20% LOE, based in Kolondieba, Mali**
The lab technicians will be responsible for receiving, cataloging and maintaining samples from **the field, including blood smears and filter papers.**

**Microscopist, 20% LOE, based in Bamako, Mali**
The microscopist will be solely responsible for reading blood smears collected as part of the study.

**Fifteen (15) Clinical officers, 25% LOE, based in Kolondieba, Mali**
Clinical officers will participate in enrollment of subjects and will be the primary interviewers at routine follow ups. They will also be available to study participants for clinical consultation during the cohort study.

#### 18.TIME FRAME/DURATION OF THE PROJECT

The trial is expected to begin in mid 2021 and continue for 2 years (2023).

#### 19.ETHICAL CONSIDERATIONS

a. Ethical committee reviews

The protocol and relevant documents will be submitted and approved by FMOS-FAPH ethical committee and then submitted to the Mali Ministry of Health to obtain authorization to conduct the study, followed by the product import permit. Additionally, this protocol and additional documents will be submitted to WHO’s Vector Control Advisory Group and WHO’s Ethical Review Committee for approval.

b. Consent

Community sensitization will occur before consent is obtained. The study will be presented to Chiefs, religious leaders, camp leaders, ministry of health representatives, and community gatherings. Once each community has agreed to participate in the study, informed consent of household heads and parents or guardians of cohort participants and individual mosquito collectors will be obtained. Consent will be obtained from 3 populations. First, consent will be sought from the heads of all HHs to receive the SR product in their house, for study staff to replace the SR product every 4 weeks and for study staff to perform periodic spot checks on the placement of the SR product. Secondly consent will be sought from the parents/guardians of children selected to participate in the cohort study. In all cases, the study will be explained in the local language, including French and Bamabara, and study staff will ensure that all questions have been addressed before the consent from is signed. For a household to receive the SR product, the head of house or his/her spouse will sign the form. The mosquito collectors will be required to give consent for themselves. In all cases, written consent will be obtained. If the participant or guardian is not able to write, a witness not associated with the study will be asked to sign to confirm that the person has consented to participate, alternatively a thumb print will be asked.

All parents/guardians whose child may die at home in the course of study will be visited at home as soon as possible for a detailed verbal autopsy interview using the standardized World Health Organization's questionnaires (WHO | Verbal autopsy standards: ascertaining and attributing causes of death) that will be used to categorize the potential cause of death. The verbal autopsy is done as a death report with causation might not be available. If a child or member of a household with intervention dies during the study, a verbal autopsy will be used to obtain causal information for reporting to the study DSMB for safety assessments.

c. Risks to study participants

There is a risk of the SR product not providing any protective effect, however, some protection is expected based on previous studies in Peru and Indonesia. The SR product contains transfluthrin which has undergone human safety testing and is currently registered in many countries throughout the world and used in currently available household products for mosquito control in the form of mosquito coils or other household pesticide products (**Appendix 0: Investigators Brochure**). However, it may cause mild eye and skin irritation, so household owners will be advised to avoid contact with eye, skin and clothing. These effects are usually transient and disappear after time. Inhalation of transfluthrin may cause central nervous system (CNS) effects, so household owners will also be advised to avoid breathing vapors directly. However, CNS effects are unlikely at the low doses at which the product will be deployed in houses. The product may be harmful if chewed on or swallowed, so recipients will be advised to keep it away from children. The SR product will be fixed at a position that is out of the reach of children and it will be monitored at replacement to ensure that it has not been moved. If a product is found to have been removed from its position in the household, study staff will discuss with the household owner to determine why it was removed and if there was any problem that led to its removal. Study staff will also reiterate safety precautions that should be taken in regard to the SR product.

Children enrolled in the cohort will be asked to provide blood at every scheduled monthly visit, every interim visit where there is a history of fever, and every sick visit for an RDT and blood smear. Participants may experience mild pain at the site of the finger stick and there is a risk of infection although our staff will use procedures to minimize this risk.

As with all medication, there is a risk of adverse event during intake of ACTs. However, the study will conform to Mali's national treatment protocols in terms of the dosage and type of ACTs to be provided in our targeted age group of children 6 months to <10 years. All adverse events will be recording through the national pharmacovigilance system and through the study's procedures.

d. Benefits to study participants

Cohort participants will be provided regular testing for malaria using RDTs and microscopy and will be treated when they test positive by either diagnosis. The testing and treatment will be provided without cost. Lastly, those clusters that will receive SRs which have been shown to reduce mosquito biting in many settings, may experience reduce malaria transmission and malaria illness.

The clearance dose will support the elimination of residual malaria parasite in the target population, which is the most vulnerable population to malaria infection. Additionally, the subjects will be followed closely throughout 30 months (6 months of baseline and 24 months of follow-up) benefiting from free malaria testing and treatment.

#### 20.EXPECTED APPLICATION OF THE RESULTS

Data from the Mali study site will be submitted to the WHO Vector Control Advisory Group which will use the data to determine whether the WHO Global Malaria Programme should recommend SRs as a new product category for vector control. If the SR product is effective, it may be deployed in malarious areas to complement other vector control interventions such as ITNs to help mitigate the problem of insecticide resistance, outdoor vector biting where ITNs may be ineffective and to further drive malaria towards elimination.

#### 21.REFERENCES:

Achee, N. L., M. J. Bangs, R. Farlow, G. F. Killeen, S. Lindsay, J. G. Logan, S. J. Moore, M. Rowland, K. Sweeney, S. J. Torr, L. J. Zwiebel, and J. P. Grieco. 2012. Spatial repellents: from discovery and development to evidence-based validation. Malaria

Annuaire Statistique 2018 du Systeme Local d'Information Sanitaire (SLIS) du Mali, 2018 Cisse,

M. B., C. Keita, A. Dicko, D. Dengela, J. Coleman, B. Lucas, J. Mihigo, A. Sadou, A. Belemvire, K. George, C. Fornadel and R. Beach "Characterizing the insecticide resistance of Anopheles gambiae in Mali." Malar J 14: 327.

Demographic Health Survey, Mali, 2018 Demographic Health Survey, Mali, 2015

Fanello, C., V. Petrarca, A. della Torre, F. Santolamazza, G. Dolo, M. Coulibaly, A. Alloueche, C. F. Curtis, Y. T. Toure and M. Coluzzi (2003). "The pyrethroid knock-down resistance gene in the Anopheles gambiae complex in Mali and further indication of incipient speciation within An. gambiae s.s." Insect Mol Biol 12(3):

Health Management Information System, DHIS2, 2018

Hill, N., H. N. Zhou, P. Wang, X. Guo, I. Carneiro, and S. J. Moore. 2014. A household randomized, controlled trial of the efficacy of 0.03% transfluthrin coils alone and in combination with long-lasting insecticidal nets on the incidence of Plasmodium falciparum and Plasmodium vivax malaria in Western Yunnan Province, China. Malar J 13: 208.

Kawada, H., E. A. Temu, J. N. Minjas, O. Matsumoto, T. Iwasaki, and M. Takagi. 2008. Field evaluation of spatial repellency of metofluthrin-impregnated plastic strips against Anopheles gambiae complex in Bagamoyo, coastal Tanzania. Journal of the American Mosquito Control Association 24: 404-409.

Keita, M., S. Traore, N. Sogoba, A. M. Dicko, B. Coulibaly, A. Sacko, S. Doumbia and S. F. Traore "[Susceptibility status of Anopheles gambiae sensu lato to insecticides commonly used for malaria control in Mali]." Bull Soc Pathol Exot 109(1): 39-45.

Lucas, J. R., Y. Shono, T. Iwasaki, T. Ishiwatari, N. Spero, and G. Benzon. 2007. U.S. laboratory and field trials of metofluthrin (SUMIONE®) emanators for reducing mosquito biting outdoors. Journal of the American Mosquito Control Association 23: 47-54.

Ogoma, S. B., S. J. Moore, and M. F. Maia. 2012. A systematic review of mosquito coils and passive emanators: defining recommendations for spatial repellency testing methodologies. Parasites and Vectors 5: 28

Recensement General de la Population et de l'Habitat du Mali (RGPH) 2009

Syafruddin, D., Bangs, M.J., Sidik, D., Elyazar, I., Asih, P.B.S., Chan, K. Nurleila, S., Nixon, C., Hendarto, J., Wahid, I., Ishak, H., Bogh, C., Grieco, J.P., Achee, N.L., Baird, J.K. 2014. Impact of a spatial repellent on a malaria incidence in two villages in Sumba, Indonesia. Am J Trop Med and Hyg. 91: 6

Tripet, F., J. Wright, A. Cornel, A. Fofana, R. McAbee, C. Meneses, L. Reimer, M. Slotman, T. Thiemann, G. Dolo, S. Traore and G. Lanzaro (2007). "Longitudinal survey of knockdown resistance to pyrethroid in Mali, West Africa, and evidence of its emergence in the Bamako form of Anopheles gambiae s.s." Am J Trop Med Hyg 76(1): 81-87.

#### 22.APPENDICES

| | |
|---|---|
| **Appendix 0:** | **Investigator's Brochure** |
| **Appendix 1a(i):** | **Informed Consent for Cohort Participation – Baseline** |
| **Appendix 1b(i):** | **Informed Consent for Cohort Participation - 24-month follow-up** |
| **Appendix 2(i):** | **Informed consent for human landing catches** |
| **Appendix 2a (i):** | **Informed consent for conducting human landing catches in a community household's room** |
| **Appendix 3(i):** | **Informed Consent to Receive SR Intervention** |
| **Appendix 4:** | **Statistical Analysis Plan** |
| **Appendix 5:** | **Informed Consent for Human Landing Catches** |
