## Supplementary material for "Effect of a spatial repellent on malaria incidence in Mali: a cluster-randomized, controlled trial": Statistical Analysis Plan

### S4. Statistical Analysis Plan

This appendix has been provided by the authors to give readers additional information about their work.

### Spatial Repellent Products for Control of Vector Borne Diseases
#### Mali
Statistical Analysis Plan

**Version 2.7**
**Nov 25, 2024**

**Prepared by**
**Fang Liu**


#### Summary of changes in the Mali study SAP

| Version | Date | Summary of Changes |
|---|---|---|
| V1.0 | Sep 24, 2019 | The first draft |
| V2.0 | Feb 13, 2020 | Revised based on the feedback from VCAG |
| V2.1 | Jan 6, 2021 | 1) Removed pregnancy testing from the study procedure and limit the age of enrolled subjects to <10yrs, per comments from WHO ERC.<br>2) Revised sample size estimation based on the updated study population and baseline first-time infection incidence rate, which is 0.7 per-person-year down from 1 per-person-year previously. |
| V2.2 | Jan 19, 2021 | Corrected of a typo ("overall new Pf malaria infection" was misplaced under "primary endpoint" in Sec 3 and should be under "secondary endpoints" in Sec 3). |
| V2.3 | March 4, 2021 | Modified the subsection (7.4) on how missing data will be handled; adjusted the definition of new infection and protection period after malaria clearance treatment in Sec 6. |
| V2.4 | Oct 7, 2021 | Corrected a few typos and clarified some wording based on the feedback during the review of the draft manuscript submitted to the journal of TRIALS. |
| V2.5 | Nov 11, 2021 | 1) Revised the safety analysis plan per the request of the UNITAID DSMB; 2) added the formula for calculating EIR |
| V2.6 | Nov 23, 2021 | Add a secondary endpoint on symptomatic (clinical) malaria |
| V2.6.1 | April 17, 2022 | Provide clear and precise definition on the endpoints in the primary and secondary hypotheses. |
| V2.7 | Nov 25, 2024 | Removed the analysis that are not performed in this study: 1) PE and entomological analysis by anopheline species (consistent with the Kenya and Indonesia trials); 2) relationship and epidemiological and entomological endpoints (deemed insignificant and uninsightful based on the results of the same type of analysis in other studies); 3) insecticide resistance (if data were collected, they would be summarized by the local study team); 4) human-behavior analysis adjusted PE and entomological data analysis (no social science data were collected in the study). Updated the sample figures and tables in the appendix (some of them were included by mistakes and do not apply to the study; the KM curves were replaced by a statistically more meaningful plots for cluster-randomized trials, to be consistent with the Kenya manuscript accepted by the Lancet. |

### Table of Contents

Summary of changes in the Mali study SAP 2
1 Objectives 4
2 Hypotheses 4
3 Endpoints 4
4 Study Design 5
5 Population for analysis 5
6 Definition of new malaria infection 6
7 Statistical Methods 7
7.1 Primary endpoint (ITT Population) 7
7.2 Secondary endpoints (ITT Population) 7
7.3 Supplementary analysis 10
7.4 Handling of missing data 10
7.5 Analysis of baseline 10
7.6 Interim analysis 11
8. Software 11
9. Sample Size Determination 11
Reference 12
Appendix 13
I. Sample size under different assumptions of baseline incidence rate 13
II. Mock Tables and Figures 14
III. Some sample SAS and R procedures used in the analysis 17

**Remark 1:** Unless otherwise stated, the term "first-time infection" in the primary hypothesis refers to the first-time malaria infection per a positive diagnosis either independently for Pf, Pv, Po, Pm, or any combinations of the four; and the term "overall infection" in the secondary hypothesis refers to the overall malaria infections per a positive diagnosis independently either for Pf, Pv, Po, Pm, or any combinations of the four.

### 1 Objectives

**Primary Objective**

1. To evaluate the protective efficacy (PE) of spatial repellent (SR) against the first-time malaria infection in Mali.

**Secondary Objectives**

1. To evaluate the PE of SR against overall new malaria infections
2. To evaluate the PE of SR against first-time clinical malaria infections.
3. To evaluate the PE of SR against first-time overall new infections by two age subgroups (6-month to 71-month old, and 6-year to 10-year old).
4. To evaluate the effect of SR on anopheline-human contact using human biting rate (HBR) as an indicator indoors for all anopheline.
5. To evaluate the effect of SR on anopheline survival and population age structure using parity rate as an indicator for all anopheline.
6. To evaluate the effect of SR on anopheline infectivity using sporozoite rate as an indicator for all anopheline.
7. To evaluate the effect of SR on anopheline infectivity using entomological inoculation rate (EIR = HBR ×sporozoite rate) as an indicator for all anopheline.
8. To evaluate the effect of SR on light trap indoor density for for all anophelines.
9. To assess the safety of SR.

### 2 Hypotheses

**Primary Hypothesis**

$H_0$: SR does not reduce the first-time malaria hazard rate compared to placebo in Mali.

$H_1$: SR reduces the first-time malaria hazard rate compared to placebo (malaria hazard ratio between SR and placebo is $< 1$; the expected hazard ratio is 70% or PE is 30%)

**Secondary Hypothesis**

$H_0$: SR does not reduce the overall new malaria hazard rate compared to placebo in Mali.

$H_1$: SR reduces the overall new malaria hazard rate compared to placebo (overall new malaria hazard ratio between SR and placebo is $< 1$; the expected hazard ratio is 70% or PE is 30%)

### 3 Endpoints

The primary endpoint is the first-time malaria infection from the intervention follow-up period. The second endpoints include:

1. Overall new malaria infections during the intervention follow-up period.
2. The first-time and overall new malaria infections by two age groups (6-month to 71monthold; and 6 year to 10- year old).

3. First-time new clinical (symptomatic) malaria infections during the intervention follow-up period
4. Anopheline-human contact using human biting rate (HBR) as an indicator indoors for all anopheline.
5. Anopheline survival and population age structure using parity rate as an indicator for all anopheline.
6. Anopheline infectivity using sporozoite rate as an indicator for all anopheline.
7. Anopheline infectivity using EIR as an indicator for all anopheline.
8. Light trap indoor density for all anophelines.
9. Adverse events and serious adverse events.

### 4 Study Design

The study design is a cluster randomized trial with 30 clusters per intervention arm (SR and Placebo), a 6-month baseline follow-up plus a 2-week baseline evaluation period, and a post-randomization intervention follow-up period of 24 months. For the evaluation of the primary objective, 26 households are recruited with each cluster into the intervention period (factoring in a 35% loss-to-follow-up rate). The same number of households (26) will be recruited into the baseline period. It is expected that 80% of the baseline households will continue to participate the intervention period. At least one kid aged from 6-month to 10-year old from each household is recruited for the biweekly (every 2 weeks) malaria check-up (scheduled and passive) during the intervention follow-up period.

Twenty clusters (10 SR, 10 placebo) will be randomly selected to estimate the impact of the SR on entomological measures of malaria transmission using light trap. Within each cluster, light trap collections will be conducted in 10 randomly selected houses every month to assess the impact of SRs on the density of Anopheles mosquitoes indoors. The indoor human landing catches (HLC) will be conducted in 4 randomly selected houses every quarter in 12 randomly selected clusters (6 SR, 6 placebo).

### 5 Population for analysis

The intention to treat (ITT) analysis is the primary analysis approach for both the primary and secondary objectives. The ITT population includes the first recruited kid from each recruited household that receives at least one SR product or placebo per the cluster randomization schedule. If a recruited subject comes from a household used for entomological data collection, that subject will be not used in the ITT analysis. The per-protocol (PP) analysis is included as a supplementary analysis for the primary and secondary objectives. The PP population includes the subjects from the ITT population that are treated following the specifications of the study protocol without major protocol deviations.

#### *5.1 Subjects who moves to a new house during the intervention follow-up period*

- For a subject who moves to a different house within the same cluster, that subject will be included in both the ITT and PP analyses. The household characteristics will be updated at the time the subjects moved.
- For a subject who moves to a different house in a different cluster, the data from the subject before the subject moves will be included in the ITT analysis. All data from the subject will be

included in the PP analysis, both the treatment information and the household characteristics will be updated at the time the subjects moved.

*5.2 Subjects who are hospitalized for serious complicated illness (e.g. chronic illness), die, drop out, or miss scheduled visits due to reasons not related to the malaria outcome or intervention during the follow-up period*

For subjects that fall under this category, the available data from the subjects (up to the time point when the subjects are hospitalized, die, or drop out; data from the scheduled visits that the subjects did not miss) will be included in both the ITT and PP analyses as the missing or absent data can be ignored (see Section 6.4 of the SAP for more details).

*5.3 Subjects who do not receive (complete) intervention due to travelling outside, mis-application or partial application of the product*

For the ITT analyses, these subjects will be included as is. For the PP analysis, “travel outside” (Y or N; an individual-level covariate) and the product application rate in each household (expected to be close to 100%) will be included as covariates if the data are not overly imbalanced between the Y and N categories for “travel outside”, and there is practically/clinically meaningful variation in the product application rate across households and clusters.

#### 6 Definition of new malaria infection

Following a positive malaria diagnosis, whether the next positive malaria diagnosis, either during the active or passive screening periods, is a new infection or just a remnant or a carryover from the previous malaria infection depends on the time lapse between the two malaria infections, and whether and when the first malaria infection is treated.

The treatment for malaria infection last for 3 days. Denote the first day of treatment by Day 1. A diagnosis test of malaria will take place on Day 15 +/- 3 days (that is, Day 12 to Day 18). If a positive malaria infection is detected during the diagnosis test, then the malaria infection will not be regarded as a new infection but rather a carryover and a second round of 3-day treatment will be conducted. The number of subjects that will be infected but not treated is expected to be minimal (to be confirmed at interim and final analyses of the baseline data). If there is no treatment for an infection, the subsequent positive will be regarded as a new infection only with one negative blood slide between the two positives.

The positive diagnosis that cannot be treated as a new infection will be re-coded as negative before any of the following statistical analysis on malaria infection (baseline, first-time infection, overall malaria infection) is applied. Correspondingly, for the overall incidence rate calculation, (the number of days from Day 1 treating a malaria infection and the post-treatment diagnosis test on Day 15 +/- 3 days).

Also noted is that the active screening of malaria occurs every 4 weeks and the passive screening occurs in between two active screenings only when a subject experiences a fever. It is possible that there are only a few passive screenings upon the completion of data, leading to data imbalance between the odd-numbered visits (the active screening) and the even-numbered visits (the passive screening). To deal with this problem if it occurs, we will apply the following approach. If the passive screening in a visit is negative on malaria, then that data point will be removed as it contains no additional info on malaria or time at risk on top of the active screenings before and after it. If the passive screening is positive, then the passive positive will be assigned to either the active screening visit immediately before the passive screening or after, whichever is closer to the passive screening in time. The same approach will be applied to the analysis of first-time overall malaria infections in Sec 7.1 and 7.2.

### 7 Statistical Methods

The statistical analysis and results reporting will follow the CONSORT guidelines for CRTs[1].

#### 7.1 Primary endpoint (ITT Population)

The baseline characteristics of the enrolled subjects, households, and clusters will be summarized by treatment arm. Specifically, we will examine subject age and gender at the individual level, wall type and roof type, floor height, house open eaves, # of window, # of doors at the household levels, and cluster population and baseline overall infection incidence at the cluster level.

The primary hypothesis on PE against the first-time malaria infection will be tested by comparing the hazard rates of first-time malaria infection between SR and placebo upon the completion of the study in the ITT population. The cloglog regression model $\log(-\log(1-\theta_{kjit})) = \beta_{0t} + x_{kji}^T\beta_1 + z_k + z_{j(k)}$ will be applied[2-7]. $\theta_{kjit}$ is the discrete time hazard rate of subject $i$ from household $j$ in cluster $k$ at time $t$, and $x_{kji}$ contains the individual-level (age, gender), household-level (number of doors, number of windows, open eaves Y or N, wall type, roof type), and cluster-level (baseline incidence rate, cluster population size, intervention group) covariates. If the data are extremely unbalanced in a categorical covariate (not in terms of the distributions between the treatment arms, but the marginal distribution of the variable itself; e.g., 99% households have the same type of walls vs 1% do not) or if a non- ignorable portion of the subjects have missing values on a covariate (due to MAR or MCAR), that covariate may be excluded in the model. $z_k \sim N(0, \sigma_1^2)$ and $z_{j(k)} \sim N(0, \sigma_2^2)$ are the random effects at the cluster and household levels respectively. The cloglog model is a proportional hazard model and thus HR does not depend on time $t$. The null hypothesis of PE = 0% is equivalent to $\beta = 0$, where $\beta$ is the log(hazard ratio) between SR and placebo. The hypothesis will be tested by the Wald's test $z = \hat{\beta}/s$, where $s$ is the estimated standard error of the estimated $\hat{\beta}$. PE is estimated by $(1 - exp\ (\hat{\beta})\ )$ ×100%, along with a 90% 2-sided CI. The lower bound of the 90% CI corresponds to the lower-bound of the 95% 1-sided CI (the hypothesis is one-sided).

#### 7.2 Secondary endpoints (ITT Population)

<u>PE of SR against the overall (both first-time and recurrent) malaria infection</u>

The secondary hypothesis on PE against the overall new malaria infections, as defined in Sec 6, will be tested by comparing the hazard rates of the overall malaria infection between SR and placebo in the ITT population. The complementary log-log (cloglog) regression model $log\ (-\ log\ (1-\theta_{kjit})\ ) = \beta_{0t} + x_{kji}^T\beta_1 + z_k + z_{j(k)} + z_{i(jk)}$ will be applied[2-7]. $\theta_{kjit}$ is the discrete time hazard rate of subject $i$ from household $j$ in cluster $k$ at time $t$, and $x_{kji}$ contains the individual-level (age, gender), household-level (number of doors, number of windows, open eaves Y or N, wall type, roof type), and cluster-level (baseline incidence rate, cluster population size, intervention group) covariates. If the data are extremely unbalanced in a categorical covariate (not in terms of the distributions between the treatment arms, but the marginal distribution of the variable itself; e.g., 99% households had the same type of walls vs 1% that did not) or if an non-ignorable portion of the subjects have missing values on a covariate (due to MAR or MCAR), that covariate may be excluded in the model. $z_k \sim N(0, \sigma_1^2)$, $z_{j(k)} \sim N(0, \sigma_2^2)$, and $z_{i(jk)} \sim N(0, \sigma_3^2)$ are the random effects at the cluster, household, and individual levels respectively. The cloglog model is a proportional hazard model and thus HR does not depend on time $t$. The null hypothesis of PE = 0% is equivalent to $\beta = 0$, where $\beta$ is the log(hazard ratio) between SR and placebo. The hypothesis will be tested by the Wald's test $z = \hat{\beta}/s$, where $s$ is the estimated standard error of the estimated $\hat{\beta}$. A one-sided p-value (the hypothesis is one-sided)and 90% 2-sided confidence interval (CI) for PE, estimated by $(1 - exp\,(\hat{\beta})$, will be provided, the lower bound of which corresponds to the lower-bound of the 95% 1-sided CI. If p-value < 0.05, we reject the null hypothesis, claiming SR reduces the overall malaria hazard rate compared to placebo in Mali at the significance level of 5%; otherwise, we fail to reject the null hypothesis, claiming SR does not reduce the overall malaria hazard rate compared to placebo in Mali.

<u>PE of SR against the first-time clinical (symptomatic) malaria infections</u>

To assess the PE of SR against the first-time symptomatic malaria infections, which will include clinical diagnosis from the scheduled visits (passive and active screening alternating every 2 weeks) and unscheduled visits if a subject experiences symptom, a similar cloglog model as used in the malaria infection above will be used. PE is estimated by $(1 - \exp{(\hat{\beta})}) \times 100\%$, where $\exp{(\hat{\beta})}$ is the estimated HR between SR and placebo, along with a 90% 2-sided CI.

<u>Subgroup PE analysis by age group</u>

The above analysis of the first-time and overall malaria infections in the examination of the PE of SR will be based on all the subjects aged 6 months to 10 years. The same set of analysis will also be performed by two age subgroups: 6-month to 71-month old, and 6-year to 10-year old to examine if the PE of SR differs by two age groups.

<u>PE analysis without baseline covariates</u>

A PE analysis on the first-time and the overall infections will be also performed by removing all the baseline covariates from the cloglog models presented Sec 7.1 and 7.2 and only keeping "intervention group" as the only covariate (in addition to visit, as a categorical predictor per the model assumptions and set-up). The hazard ratios between SR and placebo will be provided, along with 2-sided 90% CIs.

<u>Incidence rate</u>

The first-time and overall malaria incidence rates per person-year during the whole intervention follow-up will be calculated by cluster for the SR and the placebo arms respectively. The first-time malaria incidence rate is defined as the ratio of the number of first-time malaria cases during the whole study vs sum of the time to event/time at risk (in year) across the individuals within the same cluster, and the overall malaria incidence rate is defined as the ratio of the number of new malaria cases during the whole study vs sum of the time to event/time at risk (in year) for each of the new cases across the individuals within the same cluster.

Since the active screenings of malaria incidences are either every 4 weeks (active screening) with passive screening taken between two active screenings if fever is reported, the actual time for contracting malaria is unknown (interval censored). Therefore, the mid-point between two consecutive screenings will be used as the time at risk for a malaria event that occurs in the latter screening. The average per-person-year first-time and overall malaria incidence rates in the SR and placebo arms, and the incidence ratios between the two will be calculated, together with the coefficients of variation in both arms on both incidence endpoints.

<u>Effects SR on entomological endpoints</u>

The endpoints in the entomological analysis include the HBR (number of anopheline- caught during the 12-hr interval overnight), anopheline parity rate, anopheline sporozoite rate, and anopheline EIR, and the anopheline indoor density collected by light-trap.

We will report the frequencies and proportions of anopheline collected using HLC and light trap methods across clusters and treatment arm. In addition, the following analysis will be performed for each major vector and for all anophelines.

The time profile plots of each aggregated entomological endpoints will be obtained over the baseline and intervention period. An appropriate statistical model for the HBR will be identified after examining the distributional characteristics of the HBR data, which are likely to follow (zero-inflated) Poisson distribution, or (zero-inflated) negative binomial distribution if there is over-dispersion. The covariates in the models will include the fixed effects of intervention, time, cluster population size, number of houses in a cluster, location (inside or outside), location and intervention interaction; and a random effect for cluster. The ratio between SR and placebo in HBR will be estimated, and the %change in HBR by SR is given by (1-HBR ratio)·100%.

The time profile plots of each of light trap indoor density will be obtained over the baseline and intervention period. An appropriate statistical model will be identified after examining the distributional characteristics of the light trap indoor density which are likely to follow (zero-inflated) Poisson distribution, or (zero-inflated) negative binomial distribution if there is over-dispersion. The covariates in the models will include the fixed effects of intervention, time, cluster population size, and a random effect for cluster. The ratio between SR and placebo on light trap indoor density will be estimated.

The model for parity rate will be the (zero-inflated) Poisson distribution or a (zero-inflated) negative binomial distribution with the daily porous mosquitos as the outcome and the daily HBR as the offset, and the same set of covariates as those used in the model for analyzing HBR. The model for blood fed rate based on abdominal status analysis will be similar, but with the LT density as the off and the same set of covariates as those used in the model for analyzing the LT density. The model for the sporozoite rate will be similar to the parity rate with the change of outcome variable to daily mosquitos with positive sporozoite. Note that if the data on parity, fed rate, and sporozoite positivity are highly unbalanced (not in terms of the distributions between the treatment arms, but the marginal distribution of the variable itself; e.g., 99% nulliparous or 99% negative sporozoite), then the model might lead to unstable estimates or the model might not even converge. In such cases, only summary statistics will be provided.

EIR is defined as the number of infective mosquito bites a person receives per unit time (typically annually) and is calculated as HBR $\times$ sporozoite rate $= \frac{\text{# of mosquitos collected}}{\text{# of capture nights}} \times \frac{\text{# of sporozoite positive mosquitos}}{\text{# of mosquitos tested}}$. We will calculate the EIR calculation per household per cluster per unit time and provide summary statistics at baseline and per year during the intervention period by treatment group.

<u>Safety assessment</u>

Summary of symptom-based adverse events (AE), severe adverse events (SAE), and death reports observed during the studies will be reviewed by the trial DSMB at predetermined checks (quarterly). The AE/SAE will be labelled “Probable”, “Possible”, “Plausible”, “Unlikely” due to SR. Summary about AE/SAE, including mean, minimum and maximum frequencies and percentages across clusters among enrolled subjects, will be provided by the treatment arm. Statistical comparisons of the AE/SAE rates between the two arms will be conducted upon the completion of the study. Two sets of statistical analysis will be run. One set will compare the proportion of having at least one occurrence in each symptom-based AE/SAE during the whole study between the two arms, and the other will compare the total number of occurrences for each AE/SAE between the two study arms. If the data collected permits meaningful statistical hypothesis testing, p-values from the treatment comparisons will be reported, with multiplicity correction via the FDR approach [8].

##### 7.3 Supplementary analysis

Temporality of PE effects

It is expected malaria incidence changes by seasonality (rainy vs dry) and year. To examine the temporality of malaria incidence rates and the PE effect, a supplementary analysis will performed by adding the seasonality (Jun-Dec/wet/peak) and Jan-May/dry/low) and year (1 and 2) and their interaction with intervention to the covariate list in the cloglog models used for analyzing the first-time and overall infections. The PE will be estimated by seasonality and year.

Per-protocol analysis

If the PP sample set differs from the ITT sample set, per the criteria listed in Sec 5, the primary analysis on the first-time infection and the secondary analysis on the overall infections as listed in Sec 7.1 and 7.2 will also performed in the PP sample set.

##### 7.4 Handling of missing data

Significant effort will be made to avoid having missing values on outcome (malaria infection status and visit dates, and entomological endpoints). When missing values occur for an outcome for reasons not related to the outcome, reasons for missingness and the missing fraction by treatment arm and cluster will be reported. Per protocol, the subjects are screened actively on their malaria status (the outcome) every four weeks.

- If a subject misses one or more scheduled visits due to reasons not related to the SR product or the outcome, the subject will have missing values on the outcome that can be regarded as ignorable missingness (MAR or MCAR) [9].
- If a subject drops out study due to reasons unrelated to the SR product and/or malaria infection, then the missing observations from the subject can be regarded as ignorable missingness (MAR or MCAR).

In both cases, all the available data from the subject will be included in the primary and secondary analysis, without employing any specific missing data analysis techniques, due to the ignorability of the missing mechanisms.

Missing baseline covariates (individual-level, household-level, and cluster-level) that are a part of the regression models for the outcome of interest will be imputed using simple hot-deck imputation methods if the missing fraction for the covariate is <5%> If the missing fraction for a covariable are ⩾5%, appropriate multiple imputation approaches will be applied. If a non-ignorable portion of the subjects have missing values on a covariate (due to missing at random or missing completely at random), that covariate may be excluded in the model.

##### 7.5 Analysis of baseline

The per-person-year first-time and overall malaria incidence rate from the 60 recruited clusters will be calculated. Since the malaria incidences are collected on a biweekly basis, the mid-point between two visits will be imputed as the time at risk for a malaria event. The average incidence rate will be calculated, together with the coefficients of variation. The baseline analysis will occur the end of Month 3 and Month 6, respectively, during the baseline period internally.

##### 7.6 Interim analysis

No interim analysis will be performed on the malaria and entomological data collected from the intervention period post randomization. The baseline data will be analyzed at the mid-point of the baseline period.

#### 8. Software

Software used will be SAS for Windows, Version 9.4 or higher (SAS Institute, Cary, NC, USA) and RStudio Version 1.0.143 or higher (RStudio, Inc, Boston, MA, USA).

#### 9. Sample Size Determination

The sample sizes below might be adjusted based on the data collected during the 4-month baseline period. Since the adjustment will only utilize the baseline data (baseline incidence and CV) with intervention information, the Type-I error rate will not be inflated.

Primary hypothesis on first-time malaria infection

The sample size determination on the required number of households per cluster for testing the primary hypothesis on PE is based on the hazard rate comparison in the proportional hazards regression model [10-11]. With the following specifications: 1-sided type-I error rate = 5% (because the primary hypothesis is one-sided as SR is very unlikely to increase the hazard rate of malaria infection compared to placebo), true PE = 30%, a between-cluster coefficient of variance (CV) of hazard rate = 47% (based on the historical data collected from Mali) then **788 independent first-time** infections (as defined in Remark 1)- will need to be observed to reach 80% power in testing the primary hypothesis on PE.

With a baseline first-time malaria infection hazard rate of 1.0 per person-year (ppy), 30 clusters per treatment, **26 households per cluster** (factoring in a loss to follow-up rate at 35%) **in each treatment arm post randomization** are expected to yield 788 independent first-time malaria events within 24 months follow-up period per cohort post randomization to yield 80% power. If, by the end of the 2-year study, 788 independent malaria events are not reached, the study may extend until 788 events are collected without inflating the type I error rate in the testing of the primary hypothesis. Appendix I provides the sample size under different assumptions of baseline incidence rate (1.5, 1, 0.7, 0.5 and 0.25 ppy).

The same number of households (26) will be recruited into the baseline period. It is expected that 80% of the baseline households will continue to participate the intervention period.

Secondary hypothesis on overall malaria infection

The sample size calculated to yield 85% power for establishing the primary hypothesis on first-time infection PE also leads to at least 85% power when it comes to the testing of the secondary hypothesis on the overall malaria infection. This is because that the baseline overall malaria incidence rate is likely to be no lower than 0.7 per person-year (the first-time incidence rate), and there is no interim analysis on the second hypothesis.

**Note**: Since the sample sizes for PE evaluations already factors in the LTFU rate, there is no need for replacement subjects unless the LTFU is larger than assumed. If replacement subjects are to be recruited, they should not have been exposed to the intervention until the time they are considered for replacement.

#### Appendix

##### I. Sample size under different assumptions of baseline incidence rate

The table below shows, with a 47% between-cluster coefficient of variation (CV) of hazard rate 30 cluster per treatment arm, the required number of households to yield a total number of first-time independent events of 788 within a two-year intervention period for different baseline incidence rate. The calculation factors in a 35% LTFU rate for establishing the primary hypothesis on PE with 80% power under the 1-sided type-I error rate = 5% with true PE = 30%.

| baseline | # HHs per cluster | Total # of HHs (Intervention period) | # of HHs per cluster (Intervention + baseline) * | Total # of HHs (Intervention + baseline) * |
|---|---|---|---|---|
| 1.5 | 20 | 1200 | 24 | 1440 |
| 1 | 23 | 1380 | 28 | 1680 |
| 0.7 | 26 | 1560 | 32 | 1920 |
| 0.5 | 32 | 1920 | 39 | 2340 |
| 0.25 | 52 | 3120 | 63 | 3780 |
| * Assuming 80% HHs in baseline will continue to participate in the intervention period | | | | |

#### II. Mock Tables and Figures

Figure A1: flow diagram of progress of clusters and individuals (From Campbell (2010): *Consort 2010 statement: extension to cluster randomized trials*)

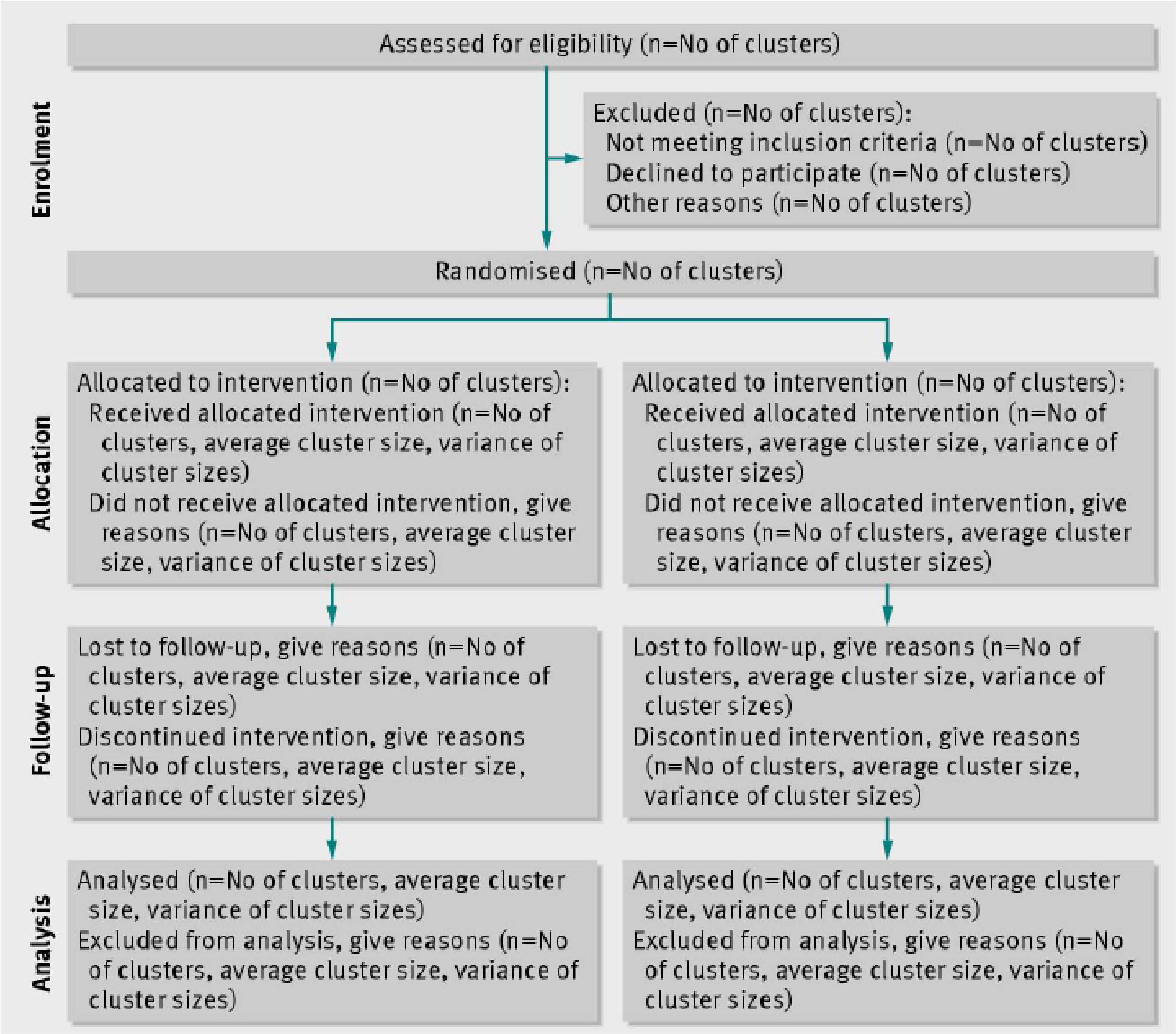

Table A1: Summary on baseline covariates

| **Individual level** | | |
|---|---|---|
| | **SR** | **Placebo** |
| Age in months (mean ± SD, (min, max))<br>Gender (% of boys) | | |
| **Household level** | | |
| | **SR** | **Placebo** |
| house wall type (%, n)<br>house roof type (%, n)<br>Floor height (mean ± SD, n)<br>house open eaves (%, n)<br># of windows (mean± SD, n)<br># of doors (mean± SD, n) | | |
| **Cluster level** | | |
| | **SR** | **Placebo** |
| Cluster population (mean ± SD, (min, max)) | | |
| Baseline overall infection incidence per person-year (mean ± SD, (min, max)) | | |

Table A2: Protective Efficacy (PE) of SR against all infections

| Treatment | Baseline incidence rate | # of households | # of infections | hazard ratio (95% CI) | PE (95% CI) | p-value |
|---|---|---|---|---|---|---|
| SR | | | | | | |
| placebo | | | | | | |

Baseline coefficient of variation (CV) of incidence rate: xxx%

Similar tables will be provided on the 1) 1st-time malaria infections, 2) by-age group analysis on PE; and 3) the supplementary PE analysis for 1st-time, overall, and by-age group infections with no covariates included in the cloglog model.

Table A3: Effects of SR compared to placebo on the entomological endpoints

| | Mean (95% CI) | | Ratio (95% CI) |
|---|---|---|---|
| Endpoint | SR | Placebo | SR vs. placebo |
| HBR | | | |
| Parity rate | | | |
| sporozoite positivity rate | | | |
| Indoor density (light trap) | | | |

Replaced by a time profile of incidence rate over time by treatment arm.

Figure A3: time profile of estimated HBR (adjusted for baseline. The time unit is every 2 weeks)

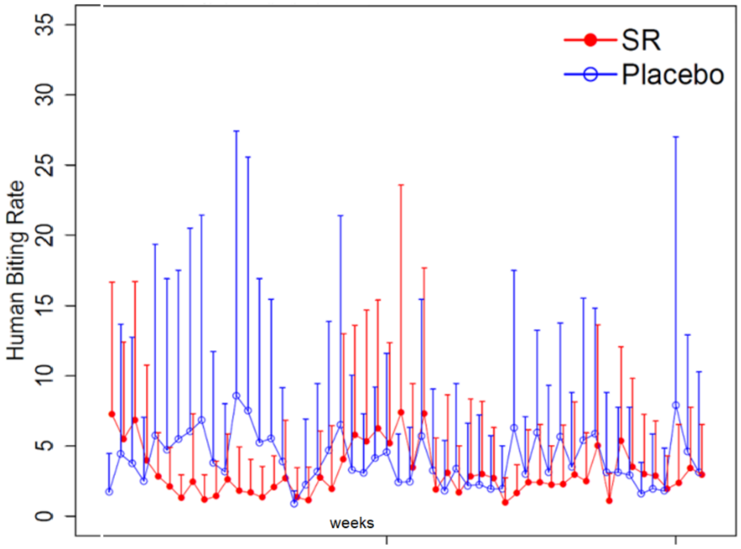

#### III. Some sample SAS and R procedures used in the analysis

Note the final codes for KM curves and estimation if PE could differ slightly from the sample codes below, which are meant to demonstrate the main procedures/commands in R and SAS to run the those two types of analyses rather than to be followed strictly.

**a)** KM curves for each cluster. Some sample codes are given below.

```
library(interval)
fit<-icfit(Surv(left,right,type="interval2")~treatment, data=malaria)
plot(fit)
```

**b)** For estimating the PE of SR against first-time and overall malaria infection SAS procedure `PROC glimmix` with the `cloglog` link. Each subject will have multiple rows, one for each visit. The statement `random` will be included to take account of the dependency of the subjects from the same cluster. Some sample codes are given below.

```
proc glimmix data=WORK.IMPORT NOCLPRINT method=LAPLACE;
class Subject_ID Cluster Final_Diagnosis Treatment_Allocation Gender
Eaves_Open wallwood recoded_visit;
model Final_Diagnosis (event='POS')=
   Treatment_Allocation
   Gender
   Eaves_Open
   wallwood
   age_scaled
   Number_of_Doors
   clusterpop_scaled
   BaselineIncidence
    recoded_visit/ dist=binary solution link=cloglog;
random int / subject=Cluster;
random int / subject=subject_ID(Cluster);

estimate 'Treatment' Treatment_Allocation 1 -1 /alpha=0.1 cl exp;
ods output estimates=treatest; run;
```
