## Supplementary material for "Effect of a spatial repellent on malaria incidence in Mali: a cluster-randomized, controlled trial": Safety Data Annex

### S5. Safety Data

This appendix has been provided by the authors to give readers additional information about their work.

Safety Summary in the Mali study, obtained by baseline and intervention periods from the analysis listed in the table below.

| | Analysis | |
|---|---|---|
| 1 | Total number of AE (adverse events) and SAE (Serious AEs) by symptom in each cluster | secondary |
| 2 | Total number of AEs (adverse events) and SAEs (Serious AEs) by symptom by treatment (spatial repellent/SR vs placebo) | secondary |
| 3 | Frequency and percentage of subjects that experienced at least one episode of AE by symptom in each cluster | secondary |
| 4 | Frequency and percentage of subjects that experienced at least one episode of SAE by symptom in each cluster | secondary |
| 5 | Histograms of the total number of AEs per cluster by treatment | secondary |
| The analysis was performed on the baseline data and intervention data, respectively. | | |

Datasets: The safety datasets contain only the subjects who had experienced at least one episode of any AE or SAE in the study. The enrolled subjects who did not have any AE or SAE during the study are not part of the safety data sets. The symptoms associated AE were defined as those the status of which were either NA or resolve; the symptoms associated SAE were defined as those the status of which falls within the categories listed in Table 1 (e.g. death, hospitalization)

Summary of the baseline period (564 subjects with at least one SAE or AE symptom episodes)

**B.1** Among the observed symptoms, fever and vomiting were the top 2 symptoms in total occurrences (361 and 259, respectively), followed by cough (182), headache (100), loss of appetite (66), diarrhea (64), nausea/vomiting (46), and abdominal pain (33). Severe malaria-related symptoms had 86 occurrences across the 60 clusters. In addition, there were 71 occurrences of "other" symptoms (Tables B1 and B5). Broken down by SR/placebo, similar patterns across different symptoms were observed. The SR clusters have more occurrences of both AE symptoms than the placebo clusters. There was significant variation in the total numbers of AE across the clusters for both SR and placebo at baseline (Figures 1 and 2; Tables B1 to B3).

**B.2** Among the observed AE symptoms, fever and vomiting were the top 2 symptoms with the highest cluster-level average prevalence rate (% percentage of subjects in a cluster with at least one AE symptom episode) across the 60 clusters at 16.54% and 12.42%, respectively. The prevalence rate was 8.3% for cough, 5.24% for headache, 3.37% for loss of appetite, 3.19% for diarrhea, and 2.38% for nausea/vomiting; prevalence rate was < 2% for each of the remaining symptoms (Table B4).

**B.3** There were 13 occurrences of SAEs in the baseline for SR and placebo combined (Table 1) including 1 death, 11 hospitalizations, and 10 life threatening events. Broken down by SR vs placebo, there were 5 occurrences in placebo clusters and 8 occurrences. There were also 33 occurrences of different symptoms for SR and placebo combined (Tables B5 to B7) with 2 Abdominal Pain, 2 Anorexia, 1 Chills, 1 Convulsions, 4 Fever, 2 Loss of Appetite, 1 Scratch on Face, 1 Severe Malaria - Prostration / Very weak, unable to stand, 1 Severe Malaria - Severe Pallor, 3 Tired, 1 Unable to eat, drink or suckle, 4 Vomiting, 1 Weak and unable to stand/move, and 9 Other. Broken down by SR vs placebo, there were 13 occurrences in 4 placebo clusters and 20 occurrences in 8 SR clusters.

**B.4** Regarding the prevalence of the symptoms associated with the SAEs at the cluster level (% percentage of subjects with at least one SAE occurrence in a cluster), the largest are 1.04% associated with both fever and vomiting (Table B8).

<u>Results of the intervention period (1112 subjects with at least one SAE or AE episodes)</u>

**I.1** Among the observed symptoms, fever and vomiting were the top 2 symptoms in both total occurrences (1762 and 741, respectively), followed by cough (623), headache (551), abdominal pain (214), loss of appetite (189), diarrhea (98), and chills (58). Severe malaria-related symptoms had 127 occurrences across the 60 clusters during intervention of the SR and placebo arms combined. In addition, there were 342 occurrences of "other" symptoms (Tables I1 and I5). Broken down by SR/placebo, similar patterns across symptoms were observed. The placebo clusters had more occurrences of AE symptoms than the SR clusters. There was significant variation in the total numbers of AE across the clusters for both SR and placebo during intervention (Figures 3 and 4; Tables I1 to I3).

**I.2** Among the observed AE symptoms, fever and vomiting were the top 2 symptoms with the highest cluster-level average prevalence rate (% percentage of subjects in a cluster with at least one AE symptom episode) across the 60 clusters at 41.39% and 26.82%, respectively. The prevalence rate was 21.21% for headache, 20.05% for cough, 8.69% for abdominal pain, 7.22% for loss of appetite, 3.97% for diarrhea, and 2.57% for chills; prevalence rate was < 2% for each of the remaining symptoms (Table I4).

**I.3** There were 13 occurrences of SAEs during the intervention for SR and placebo combined (Table 1) including 10 deaths, 2 hospitalizations, and 2 life threatening events. Broken down by SR/placebo, there were 4 occurrences in placebo clusters and 9 occurrences. There were also 45 occurrences of SAE symptoms during the intervention for SR and placebo combined (Tables I5 to I7), with 1 Abdominal Pain, 1 Convulsions, 1 Cough, 3 Diarrhea, 1 Difficulty Breathing, 8 Fever, 1 Loss of Appetite, 4 Respiratory Distress, 2 Severe Malaria - Other, 2 Severe Malaria - Prostration / Very weak, unable to stand, 1 Tired, 2 Unconsciousness/Drowsiness, 4 Vomiting, and 14 Other. Broken down by SR/placebo, there were 14 occurrences in 4 placebo clusters and 31 occurrences in 8 SR clusters.

**I.4** Regarding the prevalence of the symptoms associated with the SAEs at the cluster level (% percentage of subjects with at least one SAE occurrence in a cluster), the largest is 1.87% associated with fever (Table I8).

<u>Interim Period</u>

There was a brief interim period (1/15/2022~ 3/20/2022) between the baseline and intervention periods. During the interim period, there was one SAE occurrence in the SR and PBO clusters, respectively and the PBO clusters had more AE occurrences (98) than the SR clusters (78) (Table 2).

<u>Tables and Figures</u>

**Table 1: SAE summary over study phases**

| Study Phase | Treatment | Status/Category | # of subjects | Total # of subjects |
|---|---|---|---|---|
| Baseline | Placebo | Death | 1 | 5 |
| | | Hospitalization, Event required an intervention in order to prevent a permanent incapacity. | 1 | |
| | | Hospitalization, Life threatening | 3 | |
| | SR | Hospitalization | 1 | 8 |
| | | Hospitalization, Life threatening | 6 | |
| | | Life threatening | 1 | |
| Intervention | Placebo | Death | 3 | 4 |
| | | Hospitalization, Life threatening | 1 | |
| | SR | Death | 6 | 9 |
| | | Event required an intervention in order to prevent a permanent incapacity. | 1 | |
| | | Hospitalization, Death | 1 | |
| | | Life threatening | 1 | |

Figure 1: Total number of AE symptom episodes during the baseline

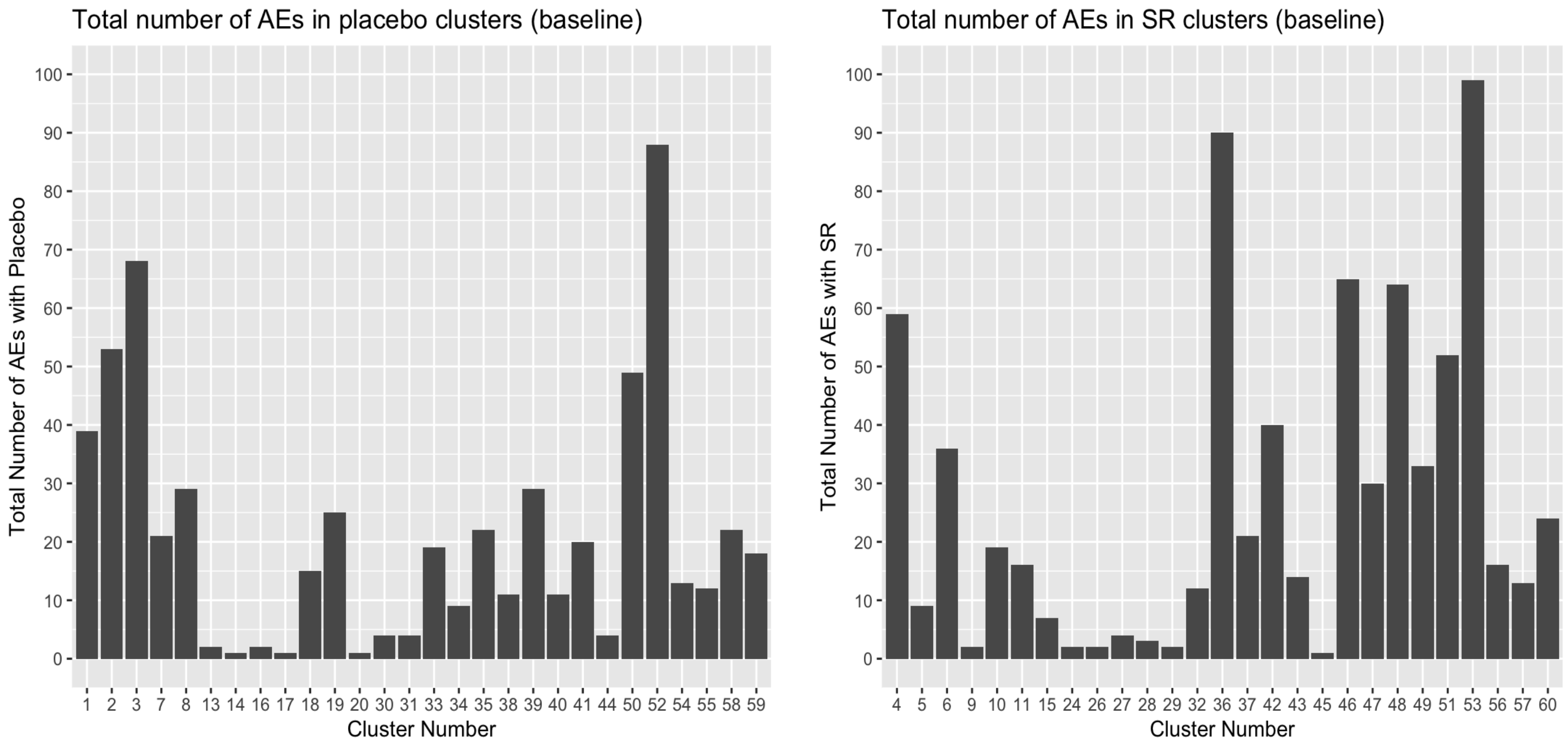

Figure 2: Total number of AE symptom episodes during the intervention

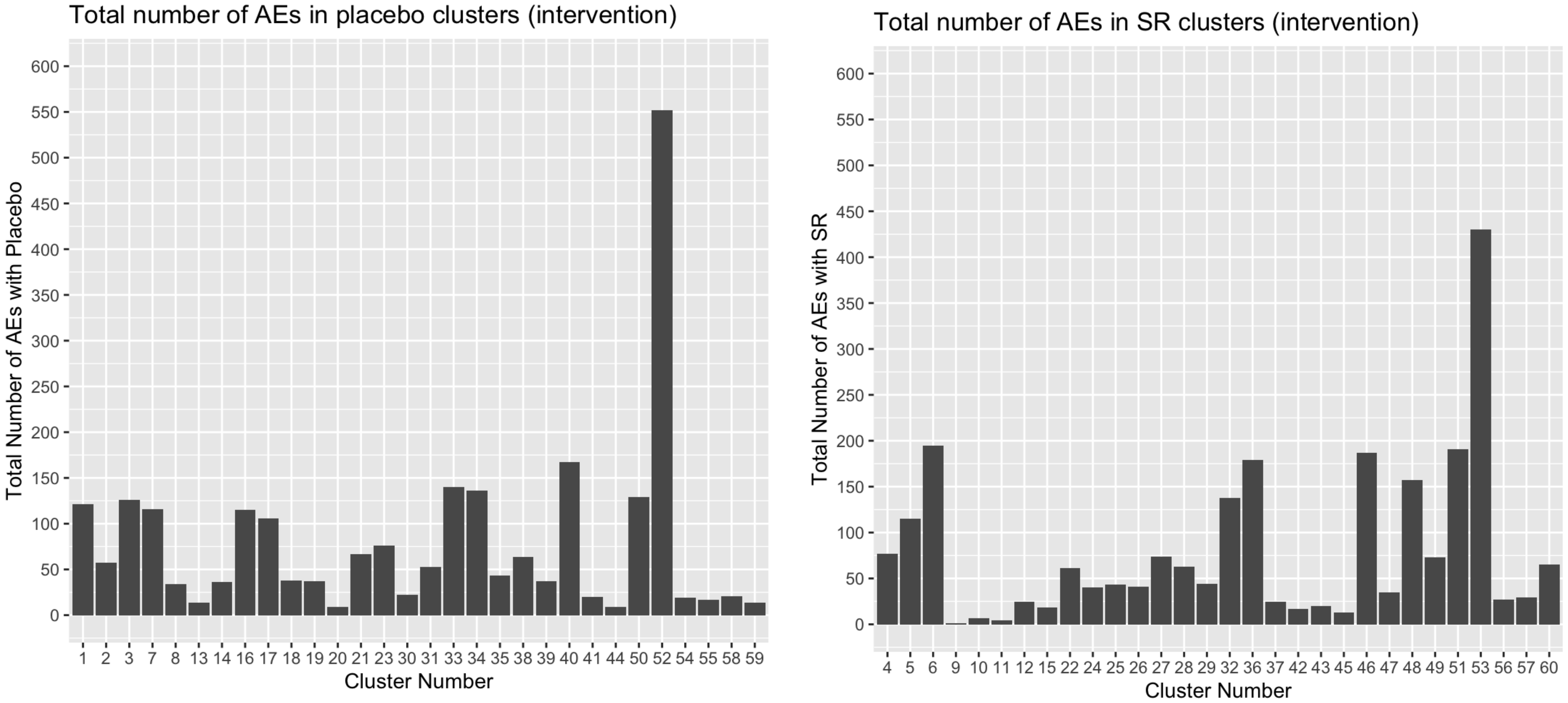

The AE and SAE summary results in the baseline and intervention periods are provided in the following Tables.

| | | **Table #** | **Contents** |
|---|---|---|---|
| **Baseline** | **AE** | B1 | number of AEs by symptom per cluster |
| | | B2 | number of AEs by symptom per cluster with SR |
| | | B3 | number of AEs by symptom per cluster with Placebo |
| | | B4 | frequency and percentage of subjects per cluster having at least one AE episode by symptom |
| | **SAE** | B5 | number of SAEs by symptom per cluster |
| | | B6 | number of SAEs by symptom per cluster with SR |
| | | B7 | number of SAEs by symptom per cluster with Placebo |
| | | B8 | frequency and percentage of subjects per cluster having at least one SAE episode by symptom |
| | | **Table #** | Contents |
| **Intervention** | **AE** | I1 | number of AEs by symptom per cluster |
| | | I2 | number of AEs by symptom per cluster with SR |
| | | I3 | number of AEs by symptom per cluster with Placebo |
| | | I4 | frequency and percentage of subjects per cluster having at least one AE episode by symptom |
| | **SAE** | I5 | number of SAEs by symptom per cluster |
| | | I6 | number of SAEs by symptom per cluster with SR |
| | | I7 | number of SAEs by symptom per cluster with Placebo |
| | | I8 | frequency and percentage of subjects per cluster having at least one SAE episode by symptom |

#### Table B1: number of AEs by symptom per cluster at the baseline (cluster numbers highlighted in red are SR clusters)

| Cluster | # of unique subjects with at least one AE | Abdominal Discomfort | Abdominal Pain | Aches | Anorexia | Chills | Convulsions | Cough | Decreased abduction of the left upper limb | Diarrhea | Difficulty Breathing | Dizziness | Eye Irritation | Fever | Headache | Loss of appetite | Nausea / Vomiting | Runny Nose | Severe Malaria - Convulsions / Seizures | Severe Malaria - Dark Urine | Severe Malaria - Other | Severe Malaria - Prostration / Very weak, unable to stand | Severe Malaria - Respiratory Distress | Severe Malaria - Severe Pallor | Severe Malnutrition | Skin Irritation / Rash | Tired | Unable to eat, drink or suckle | Vomiting | Weak and unable to stand / move | Other |
|---|---|---|---|---|---|---|---|---|---|---|---|---|---|---|---|---|---|---|---|---|---|---|---|---|---|---|---|---|---|---|---|
| 1 | 17 | 1 | 2 | 0 | 0 | 1 | 0 | 5 | 0 | 4 | 0 | 0 | 0 | 14 | 0 | 4 | 0 | 0 | 0 | 0 | 0 | 0 | 0 | 0 | 0 | 1 | 0 | 0 | 6 | 0 | 1 |
| 2 | 18 | 1 | 0 | 0 | 0 | 1 | 0 | 5 | 0 | 6 | 0 | 0 | 0 | 21 | 1 | 5 | 2 | 0 | 0 | 0 | 0 | 0 | 0 | 0 | 0 | 1 | 1 | 0 | 8 | 0 | 1 |
| 3 | 26 | 0 | 2 | 0 | 0 | 1 | 0 | 2 | 0 | 0 | 0 | 0 | 0 | 31 | 2 | 1 | 2 | 0 | 0 | 0 | 0 | 0 | 0 | 0 | 0 | 0 | 0 | 0 | 24 | 0 | 3 |
| 4 | 16 | 0 | 3 | 0 | 0 | 0 | 0 | 7 | 0 | 1 | 0 | 0 | 0 | 25 | 3 | 3 | 2 | 0 | 0 | 0 | 0 | 0 | 0 | 0 | 0 | 0 | 0 | 0 | 10 | 0 | 5 |
| 5 | 8 | 0 | 0 | 0 | 0 | 0 | 0 | 3 | 0 | 0 | 0 | 0 | 0 | 3 | 0 | 0 | 1 | 0 | 0 | 0 | 0 | 0 | 0 | 0 | 0 | 0 | 0 | 0 | 0 | 0 | 2 |
| 6 | 18 | 1 | 0 | 0 | 0 | 0 | 0 | 8 | 0 | 1 | 1 | 0 | 1 | 10 | 1 | 1 | 0 | 1 | 0 | 1 | 1 | 0 | 0 | 0 | 0 | 0 | 0 | 0 | 6 | 0 | 3 |
| 7 | 13 | 0 | 0 | 0 | 0 | 0 | 0 | 0 | 0 | 2 | 1 | 0 | 0 | 6 | 0 | 0 | 1 | 0 | 0 | 0 | 0 | 0 | 0 | 0 | 0 | 0 | 0 | 0 | 6 | 0 | 5 |
| 8 | 18 | 2 | 1 | 0 | 0 | 1 | 0 | 3 | 0 | 2 | 0 | 0 | 0 | 2 | 3 | 0 | 1 | 0 | 0 | 0 | 0 | 0 | 0 | 0 | 0 | 0 | 1 | 0 | 13 | 0 | 0 |
| 9 | 1 | 0 | 0 | 0 | 0 | 0 | 0 | 0 | 0 | 0 | 0 | 0 | 0 | 1 | 1 | 0 | 0 | 0 | 0 | 0 | 0 | 0 | 0 | 0 | 0 | 0 | 0 | 0 | 0 | 0 | 0 |
| 10 | 11 | 1 | 2 | 0 | 0 | 0 | 0 | 1 | 0 | 2 | 0 | 0 | 0 | 2 | 0 | 0 | 0 | 0 | 0 | 0 | 3 | 0 | 0 | 0 | 0 | 0 | 2 | 0 | 6 | 0 | 0 |
| 11 | 10 | 0 | 0 | 0 | 0 | 1 | 0 | 2 | 0 | 0 | 0 | 0 | 0 | 2 | 0 | 0 | 0 | 0 | 0 | 0 | 3 | 0 | 0 | 0 | 0 | 0 | 4 | 0 | 2 | 0 | 2 |
| 13 | 1 | 0 | 0 | 0 | 0 | 0 | 0 | 1 | 0 | 0 | 0 | 0 | 0 | 0 | 0 | 0 | 1 | 0 | 0 | 0 | 0 | 0 | 0 | 0 | 0 | 0 | 0 | 0 | 0 | 0 | 0 |
| 14 | 1 | 0 | 0 | 0 | 0 | 0 | 0 | 0 | 0 | 0 | 0 | 0 | 1 | 0 | 0 | 0 | 0 | 0 | 0 | 0 | 0 | 0 | 0 | 0 | 0 | 0 | 0 | 0 | 0 | 0 | 0 |
| 15 | 4 | 0 | 0 | 0 | 0 | 0 | 0 | 2 | 0 | 0 | 0 | 0 | 0 | 3 | 0 | 0 | 1 | 0 | 0 | 0 | 0 | 0 | 0 | 0 | 0 | 0 | 0 | 0 | 1 | 0 | 0 |
| 16 | 2 | 0 | 0 | 0 | 0 | 0 | 0 | 0 | 0 | 1 | 0 | 0 | 0 | 0 | 0 | 0 | 0 | 0 | 0 | 0 | 1 | 0 | 0 | 0 | 0 | 0 | 0 | 0 | 0 | 0 | 0 |
| 17 | 1 | 0 | 0 | 0 | 0 | 0 | 0 | 0 | 0 | 0 | 0 | 0 | 0 | 0 | 0 | 0 | 0 | 0 | 0 | 0 | 1 | 0 | 0 | 0 | 0 | 0 | 0 | 0 | 0 | 0 | 0 |
| 18 | 8 | 0 | 0 | 0 | 1 | 0 | 0 | 0 | 0 | 0 | 0 | 0 | 0 | 7 | 0 | 0 | 0 | 0 | 0 | 0 | 0 | 0 | 0 | 0 | 0 | 0 | 0 | 0 | 7 | 0 | 0 |
| 19 | 14 | 1 | 0 | 0 | 0 | 0 | 0 | 3 | 0 | 3 | 0 | 0 | 0 | 6 | 1 | 0 | 0 | 0 | 0 | 0 | 0 | 0 | 0 | 0 | 0 | 1 | 0 | 0 | 8 | 0 | 2 |
| 20 | 1 | 0 | 0 | 0 | 0 | 0 | 0 | 0 | 0 | 0 | 0 | 0 | 0 | 1 | 0 | 0 | 0 | 0 | 0 | 0 | 0 | 0 | 0 | 0 | 0 | 0 | 0 | 0 | 0 | 0 | 0 |
| 24 | 2 | 0 | 0 | 0 | 0 | 0 | 0 | 0 | 0 | 0 | 0 | 0 | 0 | 1 | 0 | 0 | 0 | 0 | 0 | 0 | 0 | 0 | 0 | 0 | 0 | 0 | 0 | 0 | 0 | 0 | 1 |
| 26 | 2 | 0 | 0 | 0 | 0 | 0 | 0 | 0 | 0 | 0 | 0 | 0 | 0 | 0 | 0 | 0 | 0 | 0 | 0 | 0 | 1 | 1 | 0 | 0 | 0 | 0 | 0 | 0 | 0 | 0 | 0 |
| 27 | 3 | 0 | 0 | 0 | 0 | 0 | 0 | 0 | 0 | 0 | 0 | 0 | 0 | 0 | 2 | 0 | 0 | 0 | 0 | 0 | 1 | 0 | 0 | 0 | 0 | 0 | 0 | 0 | 0 | 0 | 1 |
| 28 | 3 | 0 | 0 | 0 | 0 | 0 | 0 | 0 | 0 | 0 | 0 | 0 | 0 | 1 | 0 | 0 | 0 | 0 | 0 | 0 | 0 | 0 | 0 | 0 | 0 | 0 | 0 | 0 | 2 | 0 | 0 |
| 29 | 2 | 0 | 0 | 0 | 0 | 0 | 0 | 0 | 0 | 0 | 0 | 0 | 0 | 2 | 0 | 0 | 0 | 0 | 0 | 0 | 0 | 0 | 0 | 0 | 0 | 0 | 0 | 0 | 0 | 0 | 0 |
| 30 | 2 | 0 | 0 | 0 | 0 | 0 | 1 | 0 | 0 | 0 | 0 | 0 | 0 | 0 | 0 | 0 | 0 | 0 | 0 | 0 | 0 | 0 | 0 | 0 | 0 | 0 | 0 | 0 | 0 | 1 | 2 |
| 31 | 3 | 0 | 0 | 0 | 0 | 0 | 0 | 0 | 0 | 0 | 0 | 0 | 0 | 2 | 0 | 1 | 0 | 0 | 0 | 0 | 0 | 0 | 0 | 0 | 0 | 0 | 0 | 0 | 0 | 0 | 1 |
| 32 | 6 | 0 | 1 | 0 | 0 | 2 | 0 | 1 | 0 | 1 | 0 | 0 | 0 | 3 | 1 | 1 | 0 | 0 | 0 | 0 | 0 | 0 | 0 | 0 | 0 | 0 | 1 | 0 | 1 | 0 | 0 |
| 33 | 10 | 0 | 0 | 0 | 1 | 0 | 0 | 5 | 0 | 0 | 0 | 0 | 0 | 8 | 3 | 1 | 0 | 0 | 0 | 0 | 0 | 0 | 0 | 0 | 0 | 0 | 0 | 0 | 1 | 0 | 0 |
| 34 | 6 | 0 | 0 | 0 | 0 | 0 | 0 | 3 | 0 | 0 | 0 | 0 | 0 | 3 | 1 | 1 | 0 | 0 | 0 | 0 | 0 | 0 | 0 | 0 | 0 | 0 | 0 | 0 | 0 | 0 | 1 |
| 35 | 9 | 0 | 0 | 0 | 0 | 2 | 0 | 4 | 0 | 1 | 0 | 0 | 0 | 5 | 3 | 0 | 1 | 0 | 0 | 0 | 0 | 0 | 0 | 0 | 0 | 0 | 0 | 0 | 5 | 0 | 1 |
| 36 | 25 | 2 | 2 | 0 | 0 | 1 | 0 | 16 | 0 | 7 | 0 | 0 | 0 | 28 | 2 | 5 | 2 | 0 | 0 | 4 | 6 | 0 | 0 | 0 | 0 | 0 | 0 | 0 | 14 | 0 | 1 |
| 37 | 9 | 0 | 0 | 0 | 0 | 1 | 0 | 6 | 0 | 1 | 0 | 0 | 0 | 4 | 5 | 0 | 0 | 0 | 0 | 0 | 0 | 0 | 0 | 0 | 0 | 0 | 0 | 0 | 4 | 0 | 0 |
| 38 | 8 | 0 | 0 | 0 | 0 | 0 | 0 | 2 | 0 | 0 | 0 | 0 | 0 | 2 | 1 | 0 | 1 | 0 | 0 | 0 | 0 | 0 | 0 | 0 | 0 | 0 | 0 | 0 | 5 | 0 | 0 |
| 39 | 17 | 2 | 0 | 0 | 0 | 0 | 0 | 7 | 0 | 0 | 0 | 0 | 0 | 9 | 5 | 0 | 0 | 0 | 0 | 0 | 3 | 0 | 0 | 0 | 0 | 0 | 0 | 0 | 2 | 0 | 1 |
| 40 | 6 | 0 | 0 | 1 | 0 | 0 | 0 | 0 | 1 | 0 | 0 | 0 | 0 | 1 | 0 | 1 | 0 | 0 | 0 | 0 | 0 | 0 | 1 | 1 | 0 | 0 | 1 | 0 | 1 | 0 | 3 |
| 41 | 14 | 0 | 0 | 0 | 0 | 0 | 0 | 0 | 0 | 0 | 0 | 0 | 0 | 4 | 3 | 0 | 0 | 0 | 0 | 0 | 10 | 0 | 0 | 0 | 0 | 0 | 0 | 0 | 3 | 0 | 0 |
| 42 | 17 | 0 | 0 | 0 | 0 | 0 | 0 | 2 | 0 | 0 | 0 | 0 | 0 | 5 | 4 | 1 | 1 | 0 | 0 | 0 | 12 | 0 | 0 | 0 | 0 | 0 | 0 | 0 | 7 | 0 | 8 |
| 43 | 7 | 0 | 0 | 0 | 0 | 0 | 0 | 2 | 0 | 0 | 0 | 0 | 0 | 3 | 1 | 0 | 0 | 0 | 0 | 0 | 4 | 0 | 0 | 0 | 0 | 0 | 0 | 0 | 3 | 0 | 1 |
| 44 | 3 | 0 | 0 | 0 | 0 | 0 | 0 | 1 | 0 | 3 | 0 | 0 | 0 | 0 | 0 | 0 | 0 | 0 | 0 | 0 | 0 | 0 | 0 | 0 | 0 | 0 | 0 | 0 | 0 | 0 | 0 |
| 45 | 1 | 0 | 0 | 0 | 0 | 0 | 0 | 0 | 0 | 0 | 0 | 0 | 0 | 1 | 0 | 0 | 0 | 0 | 0 | 0 | 0 | 0 | 0 | 0 | 0 | 0 | 0 | 0 | 0 | 0 | 0 |
| 46 | 21 | 0 | 3 | 0 | 0 | 1 | 0 | 6 | 0 | 5 | 0 | 0 | 0 | 15 | 6 | 4 | 3 | 0 | 0 | 1 | 8 | 2 | 0 | 0 | 0 | 0 | 1 | 0 | 7 | 0 | 3 |
| 47 | 11 | 0 | 1 | 0 | 0 | 1 | 0 | 3 | 0 | 1 | 0 | 0 | 0 | 5 | 6 | 3 | 0 | 0 | 0 | 0 | 1 | 0 | 0 | 1 | 0 | 0 | 0 | 1 | 5 | 1 | 1 |
| 48 | 19 | 0 | 2 | 0 | 2 | 2 | 0 | 16 | 0 | 3 | 0 | 0 | 0 | 14 | 4 | 3 | 5 | 0 | 0 | 1 | 0 | 0 | 0 | 0 | 0 | 1 | 0 | 0 | 11 | 0 | 0 |
| 49 | 9 | 0 | 3 | 0 | 0 | 0 | 0 | 2 | 0 | 0 | 0 | 1 | 0 | 5 | 7 | 4 | 0 | 2 | 0 | 0 | 0 | 0 | 0 | 1 | 1 | 0 | 0 | 0 | 5 | 0 | 2 |
| 50 | 20 | 2 | 1 | 0 | 0 | 1 | 0 | 7 | 0 | 3 | 0 | 0 | 0 | 14 | 4 | 0 | 5 | 2 | 2 | 0 | 2 | 0 | 0 | 0 | 0 | 0 | 0 | 0 | 5 | 0 | 1 |
| 51 | 19 | 0 | 3 | 0 | 0 | 1 | 0 | 8 | 0 | 5 | 0 | 0 | 0 | 11 | 2 | 1 | 2 | 0 | 1 | 1 | 6 | 0 | 0 | 0 | 0 | 0 | 0 | 0 | 10 | 0 | 1 |
| 52 | 22 | 1 | 1 | 2 | 2 | 3 | 0 | 22 | 0 | 5 | 0 | 0 | 0 | 16 | 8 | 11 | 7 | 0 | 0 | 0 | 0 | 1 | 0 | 1 | 0 | 0 | 0 | 0 | 4 | 0 | 4 |
| 53 | 26 | 0 | 2 | 0 | 2 | 0 | 0 | 19 | 0 | 7 | 0 | 0 | 0 | 29 | 9 | 10 | 4 | 0 | 0 | 0 | 0 | 0 | 0 | 0 | 0 | 1 | 0 | 0 | 15 | 0 | 1 |
| 54 | 8 | 2 | 0 | 0 | 0 | 0 | 0 | 2 | 0 | 0 | 0 | 0 | 0 | 3 | 0 | 0 | 1 | 0 | 0 | 0 | 0 | 0 | 0 | 0 | 0 | 0 | 0 | 0 | 5 | 0 | 0 |
| 55 | 6 | 0 | 0 | 0 | 0 | 0 | 0 | 0 | 0 | 0 | 0 | 0 | 0 | 5 | 0 | 1 | 0 | 0 | 0 | 0 | 0 | 0 | 0 | 0 | 0 | 0 | 0 | 0 | 6 | 0 | 0 |
| 56 | 13 | 1 | 0 | 0 | 0 | 0 | 0 | 1 | 0 | 0 | 0 | 0 | 0 | 4 | 2 | 1 | 1 | 0 | 0 | 0 | 0 | 0 | 0 | 0 | 0 | 0 | 0 | 0 | 6 | 0 | 0 |
| 57 | 8 | 0 | 1 | 0 | 0 | 0 | 0 | 1 | 0 | 0 | 0 | 0 | 0 | 3 | 1 | 0 | 0 | 0 | 0 | 0 | 0 | 0 | 0 | 0 | 0 | 0 | 0 | 0 | 7 | 0 | 0 |
| 58 | 12 | 0 | 0 | 0 | 0 | 0 | 0 | 2 | 0 | 0 | 0 | 0 | 0 | 6 | 2 | 0 | 1 | 0 | 0 | 0 | 0 | 0 | 0 | 0 | 0 | 0 | 0 | 0 | 11 | 0 | 0 |
| 59 | 11 | 0 | 0 | 0 | 0 | 0 | 0 | 0 | 0 | 0 | 0 | 0 | 0 | 9 | 1 | 0 | 0 | 0 | 0 | 0 | 0 | 0 | 0 | 0 | 0 | 0 | 0 | 0 | 8 | 0 | 0 |
| 60 | 11 | 0 | 1 | 0 | 0 | 1 | 0 | 2 | 0 | 0 | 1 | 0 | 0 | 2 | 5 | 1 | 1 | 0 | 0 | 0 | 0 | 0 | 0 | 1 | 0 | 0 | 0 | 0 | 5 | 0 | 4 |
| | # of unique subjects with at least one AE | Abdominal Discomfort | Abdominal Pain | Aches | Anorexia | Chills | Convulsions | Cough | Decreased abduction of the left upper limb | Diarrhea | Difficulty Breathing | Dizziness | Eye Irritation | Fever | Headache | Loss of appetite | Nausea / Vomiting | Runny Nose | Severe Malaria - Convulsions / Seizures | Severe Malaria - Dark Urine | Severe Malaria - Other | Severe Malaria - Prostration / Very weak, unable to stand | Severe Malaria - Respiratory Distress | Severe Malaria - Severe Pallor | Severe Malnutrition | Skin Irritation / Rash | Tired | Unable to eat, drink or suckle | Vomiting | Weak and unable to stand / move | Other |
| **min** | 1 | 0 | 0 | 0 | 0 | 0 | 0 | 0 | 0 | 0 | 0 | 0 | 0 | 0 | 0 | 0 | 0 | 0 | 0 | 0 | 0 | 0 | 0 | 0 | 0 | 0 | 0 | 0 | 0 | 0 | 0 |
| **max** | 26 | 2 | 3 | 2 | 2 | 3 | 1 | 22 | 1 | 7 | 1 | 1 | 1 | 31 | 9 | 11 | 7 | 2 | 2 | 4 | 12 | 2 | 1 | 1 | 1 | 1 | 4 | 1 | 24 | 1 | 8 |
| **median** | 9 | 0 | 0 | 0 | 0 | 0 | 0 | 2 | 0 | 0 | 0 | 0 | 0 | 3 | 1 | 0 | 0 | 0 | 0 | 0 | 0 | 0 | 0 | 0 | 0 | 0 | 0 | 0 | 5 | 0 | 1 |
| **mean** | 10.16 | 0.31 | 0.56 | 0.05 | 0.15 | 0.38 | 0.02 | 3.31 | 0.02 | 1.16 | 0.05 | 0.02 | 0.04 | 6.49 | 1.82 | 1.16 | 0.84 | 0.09 | 0.05 | 0.15 | 1.15 | 0.07 | 0.02 | 0.09 | 0.02 | 0.09 | 0.2 | 0.02 | 4.64 | 0.04 | 1.13 |
| **sd** | 7.24 | 0.63 | 0.96 | 0.3 | 0.49 | 0.68 | 0.13 | 4.88 | 0.13 | 1.95 | 0.23 | 0.13 | 0.19 | 7.79 | 2.3 | 2.29 | 1.48 | 0.4 | 0.3 | 0.59 | 2.58 | 0.33 | 0.13 | 0.29 | 0.13 | 0.29 | 0.65 | 0.13 | 4.85 | 0.19 | 1.64 |
| **total** | 559 | 17 | 31 | 3 | 8 | 21 | 1 | 182 | 1 | 64 | 3 | 1 | 2 | 357 | 100 | 64 | 46 | 5 | 3 | 8 | 63 | 4 | 1 | 5 | 1 | 5 | 11 | 1 | 255 | 2 | 62 |

Note: not every cluster had AE occurrences; Cluster 12 (SR), 21 (PBO), 22 (SR), 23 (PBO), 25 (SR) have zero AE occurrences at the baseline.

#### Table B2: number of AEs by symptom in the SR clusters at the baseline

| Cluster | # of unique subjects with at least one AE | Abdominal Discomfort | Abdominal Pain | Anorexia | Chills | Cough | Diarrhea | Difficulty Breathing | Dizziness | Eye Irritation | Fever | Headache | Loss of appetite | Nausea / Vomiting | Runny Nose | Severe Malaria - Convulsions / Seizures | Severe Malaria - Dark Urine | Severe Malaria - Other | Severe Malaria - Prostration / Very weak, unable to stand | Severe Malaria - Severe Pallor | Severe Malnutrition | Skin Irritation / Rash | Tired | Unable to eat, drink or suckle | Vomiting | Weak and unable to stand / move | Other |
|---|---|---|---|---|---|---|---|---|---|---|---|---|---|---|---|---|---|---|---|---|---|---|---|---|---|---|---|
| 4 | 16 | 0 | 3 | 0 | 0 | 7 | 1 | 0 | 0 | 0 | 25 | 3 | 3 | 2 | 0 | 0 | 0 | 0 | 0 | 0 | 0 | 0 | 0 | 0 | 10 | 0 | 5 |
| 5 | 8 | 0 | 0 | 0 | 0 | 3 | 0 | 0 | 0 | 0 | 3 | 0 | 0 | 1 | 0 | 0 | 0 | 0 | 0 | 0 | 0 | 0 | 0 | 0 | 0 | 0 | 2 |
| 6 | 18 | 1 | 0 | 0 | 0 | 8 | 1 | 1 | 0 | 1 | 10 | 1 | 1 | 0 | 1 | 0 | 1 | 1 | 0 | 0 | 0 | 0 | 0 | 0 | 6 | 0 | 3 |
| 9 | 1 | 0 | 0 | 0 | 0 | 0 | 0 | 0 | 0 | 0 | 1 | 1 | 0 | 0 | 0 | 0 | 0 | 0 | 0 | 0 | 0 | 0 | 0 | 0 | 0 | 0 | 0 |
| 10 | 11 | 1 | 2 | 0 | 0 | 1 | 2 | 0 | 0 | 0 | 2 | 0 | 0 | 0 | 0 | 0 | 0 | 3 | 0 | 0 | 0 | 0 | 2 | 0 | 6 | 0 | 0 |
| 11 | 10 | 0 | 0 | 0 | 1 | 2 | 0 | 0 | 0 | 0 | 2 | 0 | 0 | 0 | 0 | 0 | 0 | 3 | 0 | 0 | 0 | 0 | 4 | 0 | 2 | 0 | 2 |
| 15 | 4 | 0 | 0 | 0 | 0 | 2 | 0 | 0 | 0 | 0 | 3 | 0 | 0 | 1 | 0 | 0 | 0 | 0 | 0 | 0 | 0 | 0 | 0 | 0 | 1 | 0 | 0 |
| 24 | 2 | 0 | 0 | 0 | 0 | 0 | 0 | 0 | 0 | 0 | 1 | 0 | 0 | 0 | 0 | 0 | 0 | 0 | 0 | 0 | 0 | 0 | 0 | 0 | 0 | 0 | 1 |
| 26 | 2 | 0 | 0 | 0 | 0 | 0 | 0 | 0 | 0 | 0 | 0 | 0 | 0 | 0 | 0 | 0 | 0 | 1 | 1 | 0 | 0 | 0 | 0 | 0 | 0 | 0 | 0 |
| 27 | 3 | 0 | 0 | 0 | 0 | 0 | 0 | 0 | 0 | 0 | 0 | 2 | 0 | 0 | 0 | 0 | 0 | 1 | 0 | 0 | 0 | 0 | 0 | 0 | 0 | 0 | 1 |
| 28 | 3 | 0 | 0 | 0 | 0 | 0 | 0 | 0 | 0 | 0 | 1 | 0 | 0 | 0 | 0 | 0 | 0 | 0 | 0 | 0 | 0 | 0 | 0 | 0 | 2 | 0 | 0 |
| 29 | 2 | 0 | 0 | 0 | 0 | 0 | 0 | 0 | 0 | 0 | 2 | 0 | 0 | 0 | 0 | 0 | 0 | 0 | 0 | 0 | 0 | 0 | 0 | 0 | 0 | 0 | 0 |
| 32 | 6 | 0 | 1 | 0 | 2 | 1 | 1 | 0 | 0 | 0 | 3 | 1 | 1 | 0 | 0 | 0 | 0 | 0 | 0 | 0 | 0 | 0 | 1 | 0 | 1 | 0 | 0 |
| 36 | 25 | 2 | 2 | 0 | 1 | 16 | 7 | 0 | 0 | 0 | 28 | 2 | 5 | 2 | 0 | 0 | 4 | 6 | 0 | 0 | 0 | 0 | 0 | 0 | 14 | 0 | 1 |
| 37 | 9 | 0 | 0 | 0 | 1 | 6 | 1 | 0 | 0 | 0 | 4 | 5 | 0 | 0 | 0 | 0 | 0 | 0 | 0 | 0 | 0 | 0 | 0 | 0 | 4 | 0 | 0 |
| 42 | 17 | 0 | 0 | 0 | 0 | 2 | 0 | 0 | 0 | 0 | 5 | 4 | 1 | 1 | 0 | 0 | 0 | 12 | 0 | 0 | 0 | 0 | 0 | 0 | 7 | 0 | 8 |
| 43 | 7 | 0 | 0 | 0 | 0 | 2 | 0 | 0 | 0 | 0 | 3 | 1 | 0 | 0 | 0 | 0 | 0 | 4 | 0 | 0 | 0 | 0 | 0 | 0 | 3 | 0 | 1 |
| 45 | 1 | 0 | 0 | 0 | 0 | 0 | 0 | 0 | 0 | 0 | 1 | 0 | 0 | 0 | 0 | 0 | 0 | 0 | 0 | 0 | 0 | 0 | 0 | 0 | 0 | 0 | 0 |
| 46 | 21 | 0 | 3 | 0 | 1 | 6 | 5 | 0 | 0 | 0 | 15 | 6 | 4 | 3 | 0 | 0 | 1 | 8 | 2 | 0 | 0 | 0 | 1 | 0 | 7 | 0 | 3 |
| 47 | 11 | 0 | 1 | 0 | 1 | 3 | 1 | 0 | 0 | 0 | 5 | 6 | 3 | 0 | 0 | 0 | 0 | 1 | 0 | 1 | 0 | 0 | 0 | 1 | 5 | 1 | 1 |
| 48 | 19 | 0 | 2 | 2 | 2 | 16 | 3 | 0 | 0 | 0 | 14 | 4 | 3 | 5 | 0 | 0 | 1 | 0 | 0 | 0 | 0 | 1 | 0 | 0 | 11 | 0 | 0 |
| 49 | 9 | 0 | 3 | 0 | 0 | 2 | 0 | 0 | 1 | 0 | 5 | 7 | 4 | 0 | 2 | 0 | 0 | 0 | 0 | 1 | 1 | 0 | 0 | 0 | 5 | 0 | 2 |
| 51 | 19 | 0 | 3 | 0 | 1 | 8 | 5 | 0 | 0 | 0 | 11 | 2 | 1 | 2 | 0 | 1 | 1 | 6 | 0 | 0 | 0 | 0 | 0 | 0 | 10 | 0 | 1 |
| 53 | 26 | 0 | 2 | 2 | 0 | 19 | 7 | 0 | 0 | 0 | 29 | 9 | 10 | 4 | 0 | 0 | 0 | 0 | 0 | 0 | 0 | 1 | 0 | 0 | 15 | 0 | 1 |
| 56 | 13 | 1 | 0 | 0 | 0 | 1 | 0 | 0 | 0 | 0 | 4 | 2 | 1 | 1 | 0 | 0 | 0 | 0 | 0 | 0 | 0 | 0 | 0 | 0 | 6 | 0 | 0 |
| 57 | 8 | 0 | 1 | 0 | 0 | 1 | 0 | 0 | 0 | 0 | 3 | 1 | 0 | 0 | 0 | 0 | 0 | 0 | 0 | 0 | 0 | 0 | 0 | 0 | 7 | 0 | 0 |
| 60 | 11 | 0 | 1 | 0 | 1 | 2 | 0 | 1 | 0 | 0 | 2 | 5 | 1 | 1 | 0 | 0 | 0 | 0 | 0 | 1 | 0 | 0 | 0 | 0 | 5 | 0 | 4 |
| | # of unique subjects with at least one AE | Abdominal Discomfort | Abdominal Pain | Anorexia | Chills | Cough | Diarrhea | Difficulty Breathing | Dizziness | Eye Irritation | Fever | Headache | Loss of appetite | Nausea / Vomiting | Runny Nose | Severe Malaria - Convulsions / Seizures | Severe Malaria - Dark Urine | Severe Malaria - Other | Severe Malaria - Prostration / Very weak, unable to stand | Severe Malaria - Severe Pallor | Severe Malnutrition | Skin Irritation / Rash | Tired | Unable to eat, drink or suckle | Vomiting | Weak and unable to stand / move | Other |
| min | 1 | 0 | 0 | 0 | 0 | 0 | 0 | 0 | 0 | 0 | 0 | 0 | 0 | 0 | 0 | 0 | 0 | 0 | 0 | 0 | 0 | 0 | 0 | 0 | 0 | 0 | 0 |
| max | 26 | 2 | 3 | 2 | 2 | 19 | 7 | 1 | 1 | 1 | 29 | 9 | 10 | 5 | 2 | 1 | 4 | 12 | 2 | 1 | 1 | 1 | 4 | 1 | 15 | 1 | 8 |
| median | 9 | 0 | 0 | 0 | 0 | 2 | 0 | 0 | 0 | 0 | 3 | 1 | 0 | 0 | 0 | 0 | 0 | 0 | 0 | 0 | 0 | 0 | 0 | 0 | 5 | 0 | 1 |
| mean | 10.44 | 0.19 | 0.89 | 0.15 | 0.41 | 4 | 1.26 | 0.07 | 0.04 | 0.04 | 6.74 | 2.3 | 1.41 | 0.85 | 0.11 | 0.04 | 0.3 | 1.7 | 0.11 | 0.11 | 0.04 | 0.07 | 0.3 | 0.04 | 4.7 | 0.04 | 1.33 |
| sd | 7.45 | 0.48 | 1.15 | 0.53 | 0.64 | 5.32 | 2.18 | 0.27 | 0.19 | 0.19 | 8.41 | 2.57 | 2.29 | 1.35 | 0.42 | 0.19 | 0.82 | 3.04 | 0.42 | 0.32 | 0.19 | 0.27 | 0.87 | 0.19 | 4.43 | 0.19 | 1.9 |
| total | 282 | 5 | 24 | 4 | 11 | 108 | 34 | 2 | 1 | 1 | 182 | 62 | 38 | 23 | 3 | 1 | 8 | 46 | 3 | 3 | 1 | 2 | 8 | 1 | 127 | 1 | 36 |

Note: not every cluster had AE occurrences. The SR clusters at the baseline did not have any AE occurrences include Cluster 12 (SR), 22 (SR), 25 (SR).

#### Table B3: number of AEs by symptom in the placebo clusters at the baseline

| Cluster | # of unique subjects with at least one AE | Abdominal Discomfort | Abdominal Pain | Aches | Anorexia | Chills | Convulsions | Cough | Decreased abduction of the left upper limb | Diarrhea | Difficulty Breathing | Eye Irritation | Fever | Headache | Loss of appetite | Nausea / Vomiting | Runny Nose | Severe Malaria - Convulsions / Seizures | Severe Malaria - Other | Severe Malaria - Prostration / Very weak, unable to stand | Severe Malaria - Respiratory Distress | Severe Malaria - Severe Pallor | Skin Irritation / Rash | Tired | Vomiting | Weak and unable to stand / move | Other |
|---|---|---|---|---|---|---|---|---|---|---|---|---|---|---|---|---|---|---|---|---|---|---|---|---|---|---|---|
| 1 | 17 | 1 | 2 | 0 | 0 | 1 | 0 | 5 | 0 | 4 | 0 | 0 | 14 | 0 | 4 | 0 | 0 | 0 | 0 | 0 | 0 | 0 | 1 | 0 | 6 | 0 | 1 |
| 2 | 18 | 1 | 0 | 0 | 0 | 1 | 0 | 5 | 0 | 6 | 0 | 0 | 21 | 1 | 5 | 2 | 0 | 0 | 0 | 0 | 0 | 0 | 1 | 1 | 8 | 0 | 1 |
| 3 | 26 | 0 | 2 | 0 | 0 | 1 | 0 | 2 | 0 | 0 | 0 | 0 | 31 | 2 | 1 | 2 | 0 | 0 | 0 | 0 | 0 | 0 | 0 | 0 | 24 | 0 | 3 |
| 7 | 13 | 0 | 0 | 0 | 0 | 0 | 0 | 0 | 0 | 2 | 1 | 0 | 6 | 0 | 0 | 1 | 0 | 0 | 0 | 0 | 0 | 0 | 0 | 0 | 6 | 0 | 5 |
| 8 | 18 | 2 | 1 | 0 | 0 | 1 | 0 | 3 | 0 | 2 | 0 | 0 | 2 | 3 | 0 | 1 | 0 | 0 | 0 | 0 | 0 | 0 | 0 | 1 | 13 | 0 | 0 |
| 13 | 1 | 0 | 0 | 0 | 0 | 0 | 0 | 1 | 0 | 0 | 0 | 0 | 0 | 0 | 0 | 1 | 0 | 0 | 0 | 0 | 0 | 0 | 0 | 0 | 0 | 0 | 0 |
| 14 | 1 | 0 | 0 | 0 | 0 | 0 | 0 | 0 | 0 | 0 | 0 | 1 | 0 | 0 | 0 | 0 | 0 | 0 | 0 | 0 | 0 | 0 | 0 | 0 | 0 | 0 | 0 |
| 16 | 2 | 0 | 0 | 0 | 0 | 0 | 0 | 0 | 0 | 1 | 0 | 0 | 0 | 0 | 0 | 0 | 0 | 0 | 1 | 0 | 0 | 0 | 0 | 0 | 0 | 0 | 0 |
| 17 | 1 | 0 | 0 | 0 | 0 | 0 | 0 | 0 | 0 | 0 | 0 | 0 | 0 | 0 | 0 | 0 | 0 | 0 | 1 | 0 | 0 | 0 | 0 | 0 | 0 | 0 | 0 |
| 18 | 8 | 0 | 0 | 0 | 1 | 0 | 0 | 0 | 0 | 0 | 0 | 0 | 7 | 0 | 0 | 0 | 0 | 0 | 0 | 0 | 0 | 0 | 0 | 0 | 7 | 0 | 0 |
| 19 | 14 | 1 | 0 | 0 | 0 | 0 | 0 | 3 | 0 | 3 | 0 | 0 | 6 | 1 | 0 | 0 | 0 | 0 | 0 | 0 | 0 | 0 | 1 | 0 | 8 | 0 | 2 |
| 20 | 1 | 0 | 0 | 0 | 0 | 0 | 0 | 0 | 0 | 0 | 0 | 0 | 1 | 0 | 0 | 0 | 0 | 0 | 0 | 0 | 0 | 0 | 0 | 0 | 0 | 0 | 0 |
| 30 | 2 | 0 | 0 | 0 | 0 | 0 | 1 | 0 | 0 | 0 | 0 | 0 | 0 | 0 | 0 | 0 | 0 | 0 | 0 | 0 | 0 | 0 | 0 | 0 | 0 | 1 | 2 |
| 31 | 3 | 0 | 0 | 0 | 0 | 0 | 0 | 0 | 0 | 0 | 0 | 0 | 2 | 0 | 1 | 0 | 0 | 0 | 0 | 0 | 0 | 0 | 0 | 0 | 0 | 0 | 1 |
| 33 | 10 | 0 | 0 | 0 | 1 | 0 | 0 | 5 | 0 | 0 | 0 | 0 | 8 | 3 | 1 | 0 | 0 | 0 | 0 | 0 | 0 | 0 | 0 | 0 | 1 | 0 | 0 |
| 34 | 6 | 0 | 0 | 0 | 0 | 0 | 0 | 3 | 0 | 0 | 0 | 0 | 3 | 1 | 1 | 0 | 0 | 0 | 0 | 0 | 0 | 0 | 0 | 0 | 0 | 0 | 1 |
| 35 | 9 | 0 | 0 | 0 | 0 | 2 | 0 | 4 | 0 | 1 | 0 | 0 | 5 | 3 | 0 | 1 | 0 | 0 | 0 | 0 | 0 | 0 | 0 | 0 | 5 | 0 | 1 |
| 38 | 8 | 0 | 0 | 0 | 0 | 0 | 0 | 2 | 0 | 0 | 0 | 0 | 2 | 1 | 0 | 1 | 0 | 0 | 0 | 0 | 0 | 0 | 0 | 0 | 5 | 0 | 0 |
| 39 | 17 | 2 | 0 | 0 | 0 | 0 | 0 | 7 | 0 | 0 | 0 | 0 | 9 | 5 | 0 | 0 | 0 | 0 | 3 | 0 | 0 | 0 | 0 | 0 | 2 | 0 | 1 |
| 40 | 6 | 0 | 0 | 1 | 0 | 0 | 0 | 0 | 1 | 0 | 0 | 0 | 1 | 0 | 1 | 0 | 0 | 0 | 0 | 0 | 1 | 1 | 0 | 1 | 1 | 0 | 3 |
| 41 | 14 | 0 | 0 | 0 | 0 | 0 | 0 | 0 | 0 | 0 | 0 | 0 | 4 | 3 | 0 | 0 | 0 | 0 | 10 | 0 | 0 | 0 | 0 | 0 | 3 | 0 | 0 |
| 44 | 3 | 0 | 0 | 0 | 0 | 0 | 0 | 1 | 0 | 3 | 0 | 0 | 0 | 0 | 0 | 0 | 0 | 0 | 0 | 0 | 0 | 0 | 0 | 0 | 0 | 0 | 0 |
| 50 | 20 | 2 | 1 | 0 | 0 | 1 | 0 | 7 | 0 | 3 | 0 | 0 | 14 | 4 | 0 | 5 | 2 | 2 | 2 | 0 | 0 | 0 | 0 | 0 | 5 | 0 | 1 |
| 52 | 22 | 1 | 1 | 2 | 2 | 3 | 0 | 22 | 0 | 5 | 0 | 0 | 16 | 8 | 11 | 7 | 0 | 0 | 0 | 1 | 0 | 1 | 0 | 0 | 4 | 0 | 4 |
| 54 | 8 | 2 | 0 | 0 | 0 | 0 | 0 | 2 | 0 | 0 | 0 | 0 | 3 | 0 | 0 | 1 | 0 | 0 | 0 | 0 | 0 | 0 | 0 | 0 | 5 | 0 | 0 |
| 55 | 6 | 0 | 0 | 0 | 0 | 0 | 0 | 0 | 0 | 0 | 0 | 0 | 5 | 0 | 1 | 0 | 0 | 0 | 0 | 0 | 0 | 0 | 0 | 0 | 6 | 0 | 0 |
| 58 | 12 | 0 | 0 | 0 | 0 | 0 | 0 | 2 | 0 | 0 | 0 | 0 | 6 | 2 | 0 | 1 | 0 | 0 | 0 | 0 | 0 | 0 | 0 | 0 | 11 | 0 | 0 |
| 59 | 11 | 0 | 0 | 0 | 0 | 0 | 0 | 0 | 0 | 0 | 0 | 0 | 9 | 1 | 0 | 0 | 0 | 0 | 0 | 0 | 0 | 0 | 0 | 0 | 8 | 0 | 0 |
| | # of unique subjects with at least one AE | Abdominal Discomfort | Abdominal Pain | Aches | Anorexia | Chills | Convulsions | Cough | Decreased abduction of the left upper limb | Diarrhea | Difficulty Breathing | Eye Irritation | Fever | Headache | Loss of appetite | Nausea / Vomiting | Runny Nose | Severe Malaria - Convulsions / Seizures | Severe Malaria - Other | Severe Malaria - Prostration / Very weak, unable to stand | Severe Malaria - Respiratory Distress | Severe Malaria - Severe Pallor | Skin Irritation / Rash | Tired | Vomiting | Weak and unable to stand / move | Other |
| **min** | 1 | 0 | 0 | 0 | 0 | 0 | 0 | 0 | 0 | 0 | 0 | 0 | 0 | 0 | 0 | 0 | 0 | 0 | 0 | 0 | 0 | 0 | 0 | 0 | 0 | 0 | 0 |
| **max** | 26 | 2 | 2 | 2 | 2 | 3 | 1 | 22 | 1 | 6 | 1 | 1 | 31 | 8 | 11 | 7 | 2 | 2 | 10 | 1 | 1 | 1 | 1 | 1 | 24 | 1 | 5 |
| **median** | 8.5 | 0 | 0 | 0 | 0 | 0 | 0 | 1.5 | 0 | 0 | 0 | 0 | 4.5 | 0.5 | 0 | 0 | 0 | 0 | 0 | 0 | 0 | 0 | 0 | 0 | 4.5 | 0 | 0 |
| **mean** | 9.89 | 0.43 | 0.25 | 0.11 | 0.14 | 0.36 | 0.04 | 2.64 | 0.04 | 1.07 | 0.04 | 0.04 | 6.25 | 1.36 | 0.93 | 0.82 | 0.07 | 0.07 | 0.61 | 0.04 | 0.04 | 0.07 | 0.11 | 0.11 | 4.57 | 0.04 | 0.93 |
| **sd** | 7.16 | 0.74 | 0.59 | 0.42 | 0.45 | 0.73 | 0.19 | 4.4 | 0.19 | 1.74 | 0.19 | 0.19 | 7.3 | 1.95 | 2.31 | 1.61 | 0.38 | 0.38 | 1.97 | 0.19 | 0.19 | 0.26 | 0.31 | 0.31 | 5.31 | 0.19 | 1.36 |
| **total** | 277 | 12 | 7 | 3 | 4 | 10 | 1 | 74 | 1 | 30 | 1 | 1 | 175 | 38 | 26 | 23 | 2 | 2 | 17 | 1 | 1 | 2 | 3 | 3 | 128 | 1 | 26 |

Note: not every cluster had AE occurrences. The PBO clusters at the baseline did not have any AE occurrences include Cluster 21 (PBO), 23 (PBO).

**Table B4: frequency and percentage of subjects having at least one AE episode by symptom at the baseline (cluster numbers highlighted in red are SR clusters)**

| Cluster | Total # of subjects | Abdominal Discomfort | | Abdominal Pain | | Aches | | Anorexia | | Chills | | Convulsions | | Cough | | Decreased abduction of the left upper limb | | Diarrhea | | Difficulty Breathing | | Dizziness | | Eye Irritation | | Fever | | Headache | | Loss of appetite | |
|---|---|---|---|---|---|---|---|---|---|---|---|---|---|---|---|---|---|---|---|---|---|---|---|---|---|---|---|---|---|---|---|
| | | # | % | # | % | # | % | # | % | # | % | # | % | # | % | # | % | # | % | # | % | # | % | # | % | # | % | # | % | # | % |
| 1 | 32 | 1 | 3.1 | 2 | 6.2 | 0 | 0 | 0 | 0 | 1 | 3.1 | 0 | 0 | 4 | 12.5 | 0 | 0 | 4 | 12.5 | 0 | 0 | 0 | 0 | 0 | 0 | 12 | 37.5 | 0 | 0 | 4 | 12.5 |
| 2 | 32 | 1 | 3.1 | 0 | 0 | 0 | 0 | 0 | 0 | 1 | 3.1 | 0 | 0 | 3 | 9.4 | 0 | 0 | 5 | 15.6 | 0 | 0 | 0 | 0 | 0 | 0 | 15 | 46.9 | 1 | 3.1 | 5 | 15.6 |
| 3 | 32 | 0 | 0 | 2 | 6.2 | 0 | 0 | 0 | 0 | 1 | 3.1 | 0 | 0 | 2 | 6.2 | 0 | 0 | 0 | 0 | 0 | 0 | 0 | 0 | 0 | 0 | 20 | 62.5 | 2 | 6.2 | 1 | 3.1 |
| 4 | 32 | 0 | 0 | 2 | 6.2 | 0 | 0 | 0 | 0 | 0 | 0 | 0 | 0 | 5 | 15.6 | 0 | 0 | 1 | 3.1 | 0 | 0 | 0 | 0 | 0 | 0 | 16 | 50 | 3 | 9.4 | 3 | 9.4 |
| 5 | 32 | 0 | 0 | 0 | 0 | 0 | 0 | 0 | 0 | 0 | 0 | 0 | 0 | 3 | 9.4 | 0 | 0 | 0 | 0 | 0 | 0 | 0 | 0 | 0 | 0 | 3 | 9.4 | 0 | 0 | 0 | 0 |
| 6 | 32 | 1 | 3.1 | 0 | 0 | 0 | 0 | 0 | 0 | 0 | 0 | 0 | 0 | 7 | 21.9 | 0 | 0 | 1 | 3.1 | 1 | 3.1 | 0 | 0 | 1 | 3.1 | 9 | 28.1 | 1 | 3.1 | 1 | 3.1 |
| 7 | 32 | 0 | 0 | 0 | 0 | 0 | 0 | 0 | 0 | 0 | 0 | 0 | 0 | 0 | 0 | 0 | 0 | 2 | 6.2 | 1 | 3.1 | 0 | 0 | 0 | 0 | 6 | 18.8 | 0 | 0 | 0 | 0 |
| 8 | 31 | 2 | 6.5 | 1 | 3.2 | 0 | 0 | 0 | 0 | 1 | 3.2 | 0 | 0 | 3 | 9.7 | 0 | 0 | 2 | 6.5 | 0 | 0 | 0 | 0 | 0 | 0 | 2 | 6.5 | 3 | 9.7 | 0 | 0 |
| 9 | 26 | 0 | 0 | 0 | 0 | 0 | 0 | 0 | 0 | 0 | 0 | 0 | 0 | 0 | 0 | 0 | 0 | 0 | 0 | 0 | 0 | 0 | 0 | 0 | 0 | 1 | 3.8 | 1 | 3.8 | 0 | 0 |
| 10 | 31 | 1 | 3.2 | 2 | 6.5 | 0 | 0 | 0 | 0 | 0 | 0 | 0 | 0 | 1 | 3.2 | 0 | 0 | 2 | 6.5 | 0 | 0 | 0 | 0 | 0 | 0 | 2 | 6.5 | 0 | 0 | 0 | 0 |
| 11 | 32 | 0 | 0 | 0 | 0 | 0 | 0 | 0 | 0 | 1 | 3.1 | 0 | 0 | 2 | 6.2 | 0 | 0 | 0 | 0 | 0 | 0 | 0 | 0 | 0 | 0 | 2 | 6.2 | 0 | 0 | 0 | 0 |
| 13 | 32 | 0 | 0 | 0 | 0 | 0 | 0 | 0 | 0 | 0 | 0 | 0 | 0 | 1 | 3.1 | 0 | 0 | 0 | 0 | 0 | 0 | 0 | 0 | 0 | 0 | 0 | 0 | 0 | 0 | 0 | 0 |
| 14 | 32 | 0 | 0 | 0 | 0 | 0 | 0 | 0 | 0 | 0 | 0 | 0 | 0 | 0 | 0 | 0 | 0 | 0 | 0 | 0 | 0 | 0 | 0 | 1 | 3.1 | 0 | 0 | 0 | 0 | 0 | 0 |
| 15 | 32 | 0 | 0 | 0 | 0 | 0 | 0 | 0 | 0 | 0 | 0 | 0 | 0 | 2 | 6.2 | 0 | 0 | 0 | 0 | 0 | 0 | 0 | 0 | 0 | 0 | 3 | 9.4 | 0 | 0 | 0 | 0 |
| 16 | 32 | 0 | 0 | 0 | 0 | 0 | 0 | 0 | 0 | 0 | 0 | 0 | 0 | 0 | 0 | 0 | 0 | 1 | 3.1 | 0 | 0 | 0 | 0 | 0 | 0 | 0 | 0 | 0 | 0 | 0 | 0 |
| 17 | 32 | 0 | 0 | 0 | 0 | 0 | 0 | 0 | 0 | 0 | 0 | 0 | 0 | 0 | 0 | 0 | 0 | 0 | 0 | 0 | 0 | 0 | 0 | 0 | 0 | 0 | 0 | 0 | 0 | 0 | 0 |
| 18 | 32 | 0 | 0 | 0 | 0 | 0 | 0 | 1 | 3.1 | 0 | 0 | 0 | 0 | 0 | 0 | 0 | 0 | 0 | 0 | 0 | 0 | 0 | 0 | 0 | 0 | 5 | 15.6 | 0 | 0 | 0 | 0 |
| 19 | 32 | 1 | 3.1 | 0 | 0 | 0 | 0 | 0 | 0 | 0 | 0 | 0 | 0 | 3 | 9.4 | 0 | 0 | 2 | 6.2 | 0 | 0 | 0 | 0 | 0 | 0 | 6 | 18.8 | 1 | 3.1 | 0 | 0 |
| 20 | 32 | 0 | 0 | 0 | 0 | 0 | 0 | 0 | 0 | 0 | 0 | 0 | 0 | 0 | 0 | 0 | 0 | 0 | 0 | 0 | 0 | 0 | 0 | 0 | 0 | 1 | 3.1 | 0 | 0 | 0 | 0 |
| 24 | 30 | 0 | 0 | 0 | 0 | 0 | 0 | 0 | 0 | 0 | 0 | 0 | 0 | 0 | 0 | 0 | 0 | 0 | 0 | 0 | 0 | 0 | 0 | 0 | 0 | 1 | 3.3 | 0 | 0 | 0 | 0 |
| 26 | 32 | 0 | 0 | 0 | 0 | 0 | 0 | 0 | 0 | 0 | 0 | 0 | 0 | 0 | 0 | 0 | 0 | 0 | 0 | 0 | 0 | 0 | 0 | 0 | 0 | 0 | 0 | 0 | 0 | 0 | 0 |
| 27 | 32 | 0 | 0 | 0 | 0 | 0 | 0 | 0 | 0 | 0 | 0 | 0 | 0 | 0 | 0 | 0 | 0 | 0 | 0 | 0 | 0 | 0 | 0 | 0 | 0 | 0 | 0 | 2 | 6.2 | 0 | 0 |
| 28 | 31 | 0 | 0 | 0 | 0 | 0 | 0 | 0 | 0 | 0 | 0 | 0 | 0 | 0 | 0 | 0 | 0 | 0 | 0 | 0 | 0 | 0 | 0 | 0 | 0 | 1 | 3.2 | 0 | 0 | 0 | 0 |
| 29 | 32 | 0 | 0 | 0 | 0 | 0 | 0 | 0 | 0 | 0 | 0 | 0 | 0 | 0 | 0 | 0 | 0 | 0 | 0 | 0 | 0 | 0 | 0 | 0 | 0 | 2 | 6.2 | 0 | 0 | 0 | 0 |
| 30 | 31 | 0 | 0 | 0 | 0 | 0 | 0 | 0 | 0 | 0 | 0 | 1 | 3.2 | 0 | 0 | 0 | 0 | 0 | 0 | 0 | 0 | 0 | 0 | 0 | 0 | 0 | 0 | 0 | 0 | 0 | 0 |
| 31 | 24 | 0 | 0 | 0 | 0 | 0 | 0 | 0 | 0 | 0 | 0 | 0 | 0 | 0 | 0 | 0 | 0 | 0 | 0 | 0 | 0 | 0 | 0 | 0 | 0 | 2 | 8.3 | 0 | 0 | 1 | 4.2 |
| 32 | 31 | 0 | 0 | 1 | 3.2 | 0 | 0 | 0 | 0 | 2 | 6.5 | 0 | 0 | 1 | 3.2 | 0 | 0 | 1 | 3.2 | 0 | 0 | 0 | 0 | 0 | 0 | 3 | 9.7 | 1 | 3.2 | 1 | 3.2 |
| 33 | 32 | 0 | 0 | 0 | 0 | 0 | 0 | 1 | 3.1 | 0 | 0 | 0 | 0 | 4 | 12.5 | 0 | 0 | 0 | 0 | 0 | 0 | 0 | 0 | 0 | 0 | 8 | 25 | 3 | 9.4 | 1 | 3.1 |
| 34 | 32 | 0 | 0 | 0 | 0 | 0 | 0 | 0 | 0 | 0 | 0 | 0 | 0 | 3 | 9.4 | 0 | 0 | 0 | 0 | 0 | 0 | 0 | 0 | 0 | 0 | 3 | 9.4 | 1 | 3.1 | 1 | 3.1 |
| 35 | 32 | 0 | 0 | 0 | 0 | 0 | 0 | 0 | 0 | 2 | 6.2 | 0 | 0 | 3 | 9.4 | 0 | 0 | 1 | 3.1 | 0 | 0 | 0 | 0 | 0 | 0 | 3 | 9.4 | 3 | 9.4 | 0 | 0 |
| 36 | 32 | 2 | 6.2 | 2 | 6.2 | 0 | 0 | 0 | 0 | 1 | 3.1 | 0 | 0 | 10 | 31.2 | 0 | 0 | 5 | 15.6 | 0 | 0 | 0 | 0 | 0 | 0 | 18 | 56.2 | 1 | 3.1 | 4 | 12.5 |
| 37 | 32 | 0 | 0 | 0 | 0 | 0 | 0 | 0 | 0 | 1 | 3.1 | 0 | 0 | 6 | 18.8 | 0 | 0 | 1 | 3.1 | 0 | 0 | 0 | 0 | 0 | 0 | 3 | 9.4 | 5 | 15.6 | 0 | 0 |
| 38 | 32 | 0 | 0 | 0 | 0 | 0 | 0 | 0 | 0 | 0 | 0 | 0 | 0 | 2 | 6.2 | 0 | 0 | 0 | 0 | 0 | 0 | 0 | 0 | 0 | 0 | 2 | 6.2 | 1 | 3.1 | 0 | 0 |
| 39 | 32 | 2 | 6.2 | 0 | 0 | 0 | 0 | 0 | 0 | 0 | 0 | 0 | 0 | 7 | 21.9 | 0 | 0 | 0 | 0 | 0 | 0 | 0 | 0 | 0 | 0 | 9 | 28.1 | 4 | 12.5 | 0 | 0 |
| 40 | 33 | 0 | 0 | 0 | 0 | 1 | 3 | 0 | 0 | 0 | 0 | 0 | 0 | 0 | 0 | 1 | 3 | 0 | 0 | 0 | 0 | 0 | 0 | 0 | 0 | 1 | 3 | 0 | 0 | 1 | 3 |
| 41 | 32 | 0 | 0 | 0 | 0 | 0 | 0 | 0 | 0 | 0 | 0 | 0 | 0 | 0 | 0 | 0 | 0 | 0 | 0 | 0 | 0 | 0 | 0 | 0 | 0 | 4 | 12.5 | 2 | 6.2 | 0 | 0 |
| 42 | 32 | 0 | 0 | 0 | 0 | 0 | 0 | 0 | 0 | 0 | 0 | 0 | 0 | 2 | 6.2 | 0 | 0 | 0 | 0 | 0 | 0 | 0 | 0 | 0 | 0 | 5 | 15.6 | 4 | 12.5 | 1 | 3.1 |
| 43 | 32 | 0 | 0 | 0 | 0 | 0 | 0 | 0 | 0 | 0 | 0 | 0 | 0 | 1 | 3.1 | 0 | 0 | 0 | 0 | 0 | 0 | 0 | 0 | 0 | 0 | 2 | 6.2 | 1 | 3.1 | 0 | 0 |
| 44 | 32 | 0 | 0 | 0 | 0 | 0 | 0 | 0 | 0 | 0 | 0 | 0 | 0 | 1 | 3.1 | 0 | 0 | 2 | 6.2 | 0 | 0 | 0 | 0 | 0 | 0 | 0 | 0 | 0 | 0 | 0 | 0 |
| 45 | 32 | 0 | 0 | 0 | 0 | 0 | 0 | 0 | 0 | 0 | 0 | 0 | 0 | 0 | 0 | 0 | 0 | 0 | 0 | 0 | 0 | 0 | 0 | 0 | 0 | 1 | 3.1 | 0 | 0 | 0 | 0 |
| 46 | 32 | 0 | 0 | 3 | 9.4 | 0 | 0 | 0 | 0 | 1 | 3.1 | 0 | 0 | 6 | 18.8 | 0 | 0 | 5 | 15.6 | 0 | 0 | 0 | 0 | 0 | 0 | 13 | 40.6 | 5 | 15.6 | 3 | 9.4 |
| 47 | 32 | 0 | 0 | 1 | 3.1 | 0 | 0 | 0 | 0 | 1 | 3.1 | 0 | 0 | 3 | 9.4 | 0 | 0 | 1 | 3.1 | 0 | 0 | 0 | 0 | 0 | 0 | 4 | 12.5 | 6 | 18.8 | 2 | 6.2 |
| 48 | 32 | 0 | 0 | 2 | 6.2 | 0 | 0 | 2 | 6.2 | 2 | 6.2 | 0 | 0 | 11 | 34.4 | 0 | 0 | 3 | 9.4 | 0 | 0 | 0 | 0 | 0 | 0 | 11 | 34.4 | 4 | 12.5 | 3 | 9.4 |
| 49 | 32 | 0 | 0 | 3 | 9.4 | 0 | 0 | 0 | 0 | 0 | 0 | 0 | 0 | 2 | 6.2 | 0 | 0 | 0 | 0 | 0 | 0 | 1 | 3.1 | 0 | 0 | 5 | 15.6 | 6 | 18.8 | 4 | 12.5 |
| 50 | 32 | 2 | 6.2 | 1 | 3.1 | 0 | 0 | 0 | 0 | 1 | 3.1 | 0 | 0 | 7 | 21.9 | 0 | 0 | 3 | 9.4 | 0 | 0 | 0 | 0 | 0 | 0 | 12 | 37.5 | 4 | 12.5 | 0 | 0 |
| 51 | 32 | 0 | 0 | 2 | 6.2 | 0 | 0 | 0 | 0 | 1 | 3.1 | 0 | 0 | 6 | 18.8 | 0 | 0 | 3 | 9.4 | 0 | 0 | 0 | 0 | 0 | 0 | 10 | 31.2 | 2 | 6.2 | 1 | 3.1 |
| 52 | 32 | 1 | 3.1 | 1 | 3.1 | 2 | 6.2 | 2 | 6.2 | 2 | 6.2 | 0 | 0 | 14 | 43.8 | 0 | 0 | 5 | 15.6 | 0 | 0 | 0 | 0 | 0 | 0 | 14 | 43.8 | 6 | 18.8 | 10 | 31.2 |
| 53 | 32 | 0 | 0 | 2 | 6.2 | 0 | 0 | 2 | 6.2 | 0 | 0 | 0 | 0 | 13 | 40.6 | 0 | 0 | 6 | 18.8 | 0 | 0 | 0 | 0 | 0 | 0 | 20 | 62.5 | 8 | 25 | 9 | 28.1 |
| 54 | 32 | 2 | 6.2 | 0 | 0 | 0 | 0 | 0 | 0 | 0 | 0 | 0 | 0 | 2 | 6.2 | 0 | 0 | 0 | 0 | 0 | 0 | 0 | 0 | 0 | 0 | 3 | 9.4 | 0 | 0 | 0 | 0 |
| 55 | 32 | 0 | 0 | 0 | 0 | 0 | 0 | 0 | 0 | 0 | 0 | 0 | 0 | 0 | 0 | 0 | 0 | 0 | 0 | 0 | 0 | 0 | 0 | 0 | 0 | 3 | 9.4 | 0 | 0 | 1 | 3.1 |
| 56 | 32 | 1 | 3.1 | 0 | 0 | 0 | 0 | 0 | 0 | 0 | 0 | 0 | 0 | 1 | 3.1 | 0 | 0 | 0 | 0 | 0 | 0 | 0 | 0 | 0 | 0 | 4 | 12.5 | 2 | 6.2 | 1 | 3.1 |
| 57 | 32 | 0 | 0 | 1 | 3.1 | 0 | 0 | 0 | 0 | 0 | 0 | 0 | 0 | 1 | 3.1 | 0 | 0 | 0 | 0 | 0 | 0 | 0 | 0 | 0 | 0 | 3 | 9.4 | 1 | 3.1 | 0 | 0 |
| 58 | 32 | 0 | 0 | 0 | 0 | 0 | 0 | 0 | 0 | 0 | 0 | 0 | 0 | 2 | 6.2 | 0 | 0 | 0 | 0 | 0 | 0 | 0 | 0 | 0 | 0 | 6 | 18.8 | 2 | 6.2 | 0 | 0 |
| 59 | 32 | 0 | 0 | 0 | 0 | 0 | 0 | 0 | 0 | 0 | 0 | 0 | 0 | 0 | 0 | 0 | 0 | 0 | 0 | 0 | 0 | 0 | 0 | 0 | 0 | 9 | 28.1 | 1 | 3.1 | 0 | 0 |
| 60 | 32 | 0 | 0 | 1 | 3.1 | 0 | 0 | 0 | 0 | 1 | 3.1 | 0 | 0 | 2 | 6.2 | 0 | 0 | 0 | 0 | 1 | 3.1 | 0 | 0 | 0 | 0 | 2 | 6.2 | 5 | 15.6 | 1 | 3.1 |
| | Total # of subjects | Abdominal Discomfort | | Abdominal Pain | | Aches | | Anorexia | | Chills | | Convulsions | | Cough | | Decreased abduction of the left upper limb | | Diarrhea | | Difficulty Breathing | | Dizziness | | Eye Irritation | | Fever | | Headache | | Loss of appetite | |
| | | # | % | # | % | # | % | # | % | # | % | # | % | # | % | # | % | # | % | # | % | # | % | # | % | # | % | # | % | # | % |
| min | 24 | 0 | 0 | 0 | 0 | 0 | 0 | 0 | 0 | 0 | 0 | 0 | 0 | 0 | 0 | 0 | 0 | 0 | 0 | 0 | 0 | 0 | 0 | 0 | 0 | 0 | 0 | 0 | 0 | 0 | 0 |

| Cluster | Nausea / Vomiting | | Runny Nose | | Severe Malaria - Convulsions / Seizures | | Severe Malaria - Dark Urine | | Severe Malaria - Other | | Severe malaria: prostration / very weak, unable to stand | | Severe Malaria - Respiratory Distress | | Severe Malaria - Severe Pallor | | Severe Malnutrition | | Skin Irritation / Rash | | Tired | | Unable to eat, drink or suckle | | Vomiting | | Weak and unable to stand / move | | Other | |
|---|---|---|---|---|---|---|---|---|---|---|---|---|---|---|---|---|---|---|---|---|---|---|---|---|---|---|---|---|---|---|
| | # | % | # | % | # | % | # | % | # | % | # | % | # | % | # | % | # | % | # | % | # | % | # | % | # | % | # | % | # | % |
| 1 | 0 | 0 | 0 | 0 | 0 | 0 | 0 | 0 | 0 | 0 | 0 | 0 | 0 | 0 | 0 | 0 | 0 | 0 | 1 | 3.1 | 0 | 0 | 0 | 0 | 5 | 15.6 | 0 | 0 | 1 | 3.1 |
| 2 | 2 | 6.2 | 0 | 0 | 0 | 0 | 0 | 0 | 0 | 0 | 0 | 0 | 0 | 0 | 0 | 0 | 0 | 0 | 1 | 3.1 | 1 | 3.1 | 0 | 0 | 7 | 21.9 | 0 | 0 | 1 | 3.1 |
| 3 | 2 | 6.2 | 0 | 0 | 0 | 0 | 0 | 0 | 0 | 0 | 0 | 0 | 0 | 0 | 0 | 0 | 0 | 0 | 0 | 0 | 0 | 0 | 0 | 0 | 17 | 53.1 | 0 | 0 | 2 | 6.2 |
| 4 | 2 | 6.2 | 0 | 0 | 0 | 0 | 0 | 0 | 0 | 0 | 0 | 0 | 0 | 0 | 0 | 0 | 0 | 0 | 0 | 0 | 0 | 0 | 0 | 0 | 8 | 25 | 0 | 0 | 3 | 9.4 |
| 5 | 1 | 3.1 | 0 | 0 | 0 | 0 | 0 | 0 | 0 | 0 | 0 | 0 | 0 | 0 | 0 | 0 | 0 | 0 | 0 | 0 | 0 | 0 | 0 | 0 | 0 | 0 | 0 | 0 | 2 | 6.2 |
| 6 | 0 | 0 | 1 | 3.1 | 0 | 0 | 1 | 3.1 | 1 | 3.1 | 0 | 0 | 0 | 0 | 0 | 0 | 0 | 0 | 0 | 0 | 0 | 0 | 0 | 0 | 6 | 18.8 | 0 | 0 | 3 | 9.4 |
| 7 | 1 | 3.1 | 0 | 0 | 0 | 0 | 0 | 0 | 0 | 0 | 0 | 0 | 0 | 0 | 0 | 0 | 0 | 0 | 0 | 0 | 0 | 0 | 0 | 0 | 4 | 12.5 | 0 | 0 | 3 | 9.4 |
| 8 | 1 | 3.2 | 0 | 0 | 0 | 0 | 0 | 0 | 0 | 0 | 0 | 0 | 0 | 0 | 0 | 0 | 0 | 0 | 0 | 0 | 1 | 3.2 | 0 | 0 | 10 | 32.3 | 0 | 0 | 0 | 0 |
| 9 | 0 | 0 | 0 | 0 | 0 | 0 | 0 | 0 | 0 | 0 | 0 | 0 | 0 | 0 | 0 | 0 | 0 | 0 | 0 | 0 | 0 | 0 | 0 | 0 | 0 | 0 | 0 | 0 | 0 | 0 |
| 10 | 0 | 0 | 0 | 0 | 0 | 0 | 0 | 0 | 3 | 9.7 | 0 | 0 | 0 | 0 | 0 | 0 | 0 | 0 | 0 | 0 | 1 | 3.2 | 0 | 0 | 5 | 16.1 | 0 | 0 | 0 | 0 |
| 11 | 0 | 0 | 0 | 0 | 0 | 0 | 0 | 0 | 3 | 9.4 | 0 | 0 | 0 | 0 | 0 | 0 | 0 | 0 | 0 | 0 | 4 | 12.5 | 0 | 0 | 2 | 6.2 | 0 | 0 | 2 | 6.2 |
| 13 | 1 | 3.1 | 0 | 0 | 0 | 0 | 0 | 0 | 0 | 0 | 0 | 0 | 0 | 0 | 0 | 0 | 0 | 0 | 0 | 0 | 0 | 0 | 0 | 0 | 0 | 0 | 0 | 0 | 0 | 0 |
| 14 | 0 | 0 | 0 | 0 | 0 | 0 | 0 | 0 | 0 | 0 | 0 | 0 | 0 | 0 | 0 | 0 | 0 | 0 | 0 | 0 | 0 | 0 | 0 | 0 | 0 | 0 | 0 | 0 | 0 | 0 |
| 15 | 1 | 3.1 | 0 | 0 | 0 | 0 | 0 | 0 | 0 | 0 | 0 | 0 | 0 | 0 | 0 | 0 | 0 | 0 | 0 | 0 | 0 | 0 | 0 | 0 | 1 | 3.1 | 0 | 0 | 0 | 0 |
| 16 | 0 | 0 | 0 | 0 | 0 | 0 | 0 | 0 | 1 | 3.1 | 0 | 0 | 0 | 0 | 0 | 0 | 0 | 0 | 0 | 0 | 0 | 0 | 0 | 0 | 0 | 0 | 0 | 0 | 0 | 0 |
| 17 | 0 | 0 | 0 | 0 | 0 | 0 | 0 | 0 | 1 | 3.1 | 0 | 0 | 0 | 0 | 0 | 0 | 0 | 0 | 0 | 0 | 0 | 0 | 0 | 0 | 0 | 0 | 0 | 0 | 0 | 0 |
| 18 | 0 | 0 | 0 | 0 | 0 | 0 | 0 | 0 | 0 | 0 | 0 | 0 | 0 | 0 | 0 | 0 | 0 | 0 | 0 | 0 | 0 | 0 | 0 | 0 | 7 | 21.9 | 0 | 0 | 0 | 0 |
| 19 | 0 | 0 | 0 | 0 | 0 | 0 | 0 | 0 | 0 | 0 | 0 | 0 | 0 | 0 | 0 | 0 | 0 | 0 | 1 | 3.1 | 0 | 0 | 0 | 0 | 7 | 21.9 | 0 | 0 | 2 | 6.2 |
| 20 | 0 | 0 | 0 | 0 | 0 | 0 | 0 | 0 | 0 | 0 | 0 | 0 | 0 | 0 | 0 | 0 | 0 | 0 | 0 | 0 | 0 | 0 | 0 | 0 | 0 | 0 | 0 | 0 | 0 | 0 |
| 24 | 0 | 0 | 0 | 0 | 0 | 0 | 0 | 0 | 0 | 0 | 0 | 0 | 0 | 0 | 0 | 0 | 0 | 0 | 0 | 0 | 0 | 0 | 0 | 0 | 0 | 0 | 0 | 0 | 1 | 3.3 |
| 26 | 0 | 0 | 0 | 0 | 0 | 0 | 0 | 0 | 1 | 3.1 | 1 | 3.1 | 0 | 0 | 0 | 0 | 0 | 0 | 0 | 0 | 0 | 0 | 0 | 0 | 0 | 0 | 0 | 0 | 0 | 0 |
| 27 | 0 | 0 | 0 | 0 | 0 | 0 | 0 | 0 | 1 | 3.1 | 0 | 0 | 0 | 0 | 0 | 0 | 0 | 0 | 0 | 0 | 0 | 0 | 0 | 0 | 0 | 0 | 0 | 0 | 1 | 3.1 |
| 28 | 0 | 0 | 0 | 0 | 0 | 0 | 0 | 0 | 0 | 0 | 0 | 0 | 0 | 0 | 0 | 0 | 0 | 0 | 0 | 0 | 0 | 0 | 0 | 0 | 2 | 6.5 | 0 | 0 | 0 | 0 |
| 29 | 0 | 0 | 0 | 0 | 0 | 0 | 0 | 0 | 0 | 0 | 0 | 0 | 0 | 0 | 0 | 0 | 0 | 0 | 0 | 0 | 0 | 0 | 0 | 0 | 0 | 0 | 0 | 0 | 0 | 0 |
| 30 | 0 | 0 | 0 | 0 | 0 | 0 | 0 | 0 | 0 | 0 | 0 | 0 | 0 | 0 | 0 | 0 | 0 | 0 | 0 | 0 | 0 | 0 | 0 | 0 | 0 | 0 | 1 | 3.2 | 2 | 6.5 |
| 31 | 0 | 0 | 0 | 0 | 0 | 0 | 0 | 0 | 0 | 0 | 0 | 0 | 0 | 0 | 0 | 0 | 0 | 0 | 0 | 0 | 0 | 0 | 0 | 0 | 0 | 0 | 0 | 0 | 1 | 4.2 |
| 32 | 0 | 0 | 0 | 0 | 0 | 0 | 0 | 0 | 0 | 0 | 0 | 0 | 0 | 0 | 0 | 0 | 0 | 0 | 0 | 0 | 1 | 3.2 | 0 | 0 | 1 | 3.2 | 0 | 0 | 0 | 0 |
| 33 | 0 | 0 | 0 | 0 | 0 | 0 | 0 | 0 | 0 | 0 | 0 | 0 | 0 | 0 | 0 | 0 | 0 | 0 | 0 | 0 | 0 | 0 | 0 | 0 | 1 | 3.1 | 0 | 0 | 0 | 0 |
| 34 | 0 | 0 | 0 | 0 | 0 | 0 | 0 | 0 | 0 | 0 | 0 | 0 | 0 | 0 | 0 | 0 | 0 | 0 | 0 | 0 | 0 | 0 | 0 | 0 | 0 | 0 | 0 | 0 | 1 | 3.1 |
| 35 | 1 | 3.1 | 0 | 0 | 0 | 0 | 0 | 0 | 0 | 0 | 0 | 0 | 0 | 0 | 0 | 0 | 0 | 0 | 0 | 0 | 0 | 0 | 0 | 0 | 3 | 9.4 | 0 | 0 | 1 | 3.1 |
| 36 | 2 | 6.2 | 0 | 0 | 0 | 0 | 2 | 6.2 | 6 | 18.8 | 0 | 0 | 0 | 0 | 0 | 0 | 0 | 0 | 0 | 0 | 0 | 0 | 0 | 0 | 11 | 34.4 | 0 | 0 | 1 | 3.1 |
| 37 | 0 | 0 | 0 | 0 | 0 | 0 | 0 | 0 | 0 | 0 | 0 | 0 | 0 | 0 | 0 | 0 | 0 | 0 | 0 | 0 | 0 | 0 | 0 | 0 | 3 | 9.4 | 0 | 0 | 0 | 0 |
| 38 | 1 | 3.1 | 0 | 0 | 0 | 0 | 0 | 0 | 0 | 0 | 0 | 0 | 0 | 0 | 0 | 0 | 0 | 0 | 0 | 0 | 0 | 0 | 0 | 0 | 5 | 15.6 | 0 | 0 | 0 | 0 |
| 39 | 0 | 0 | 0 | 0 | 0 | 0 | 0 | 0 | 2 | 6.2 | 0 | 0 | 0 | 0 | 0 | 0 | 0 | 0 | 0 | 0 | 0 | 0 | 0 | 0 | 2 | 6.2 | 0 | 0 | 1 | 3.1 |
| 40 | 0 | 0 | 0 | 0 | 0 | 0 | 0 | 0 | 0 | 0 | 0 | 0 | 1 | 3 | 1 | 3 | 0 | 0 | 0 | 0 | 1 | 3 | 0 | 0 | 1 | 3 | 0 | 0 | 3 | 9.1 |
| 41 | 0 | 0 | 0 | 0 | 0 | 0 | 0 | 0 | 9 | 28.1 | 0 | 0 | 0 | 0 | 0 | 0 | 0 | 0 | 0 | 0 | 0 | 0 | 0 | 0 | 3 | 9.4 | 0 | 0 | 0 | 0 |
| 42 | 1 | 3.1 | 0 | 0 | 0 | 0 | 0 | 0 | 10 | 31.2 | 0 | 0 | 0 | 0 | 0 | 0 | 0 | 0 | 0 | 0 | 0 | 0 | 0 | 0 | 5 | 15.6 | 0 | 0 | 4 | 12.5 |
| 43 | 0 | 0 | 0 | 0 | 0 | 0 | 0 | 0 | 3 | 9.4 | 0 | 0 | 0 | 0 | 0 | 0 | 0 | 0 | 0 | 0 | 0 | 0 | 0 | 0 | 2 | 6.2 | 0 | 0 | 1 | 3.1 |
| 44 | 0 | 0 | 0 | 0 | 0 | 0 | 0 | 0 | 0 | 0 | 0 | 0 | 0 | 0 | 0 | 0 | 0 | 0 | 0 | 0 | 0 | 0 | 0 | 0 | 0 | 0 | 0 | 0 | 0 | 0 |
| 45 | 0 | 0 | 0 | 0 | 0 | 0 | 0 | 0 | 0 | 0 | 0 | 0 | 0 | 0 | 0 | 0 | 0 | 0 | 0 | 0 | 0 | 0 | 0 | 0 | 0 | 0 | 0 | 0 | 0 | 0 |
| 46 | 2 | 6.2 | 0 | 0 | 0 | 0 | 1 | 3.1 | 7 | 21.9 | 1 | 3.1 | 0 | 0 | 0 | 0 | 0 | 0 | 0 | 0 | 1 | 3.1 | 0 | 0 | 6 | 18.8 | 0 | 0 | 2 | 6.2 |
| 47 | 0 | 0 | 0 | 0 | 0 | 0 | 0 | 0 | 1 | 3.1 | 0 | 0 | 0 | 0 | 1 | 3.1 | 0 | 0 | 0 | 0 | 0 | 0 | 1 | 3.1 | 5 | 15.6 | 1 | 3.1 | 1 | 3.1 |
| 48 | 5 | 15.6 | 0 | 0 | 0 | 0 | 1 | 3.1 | 0 | 0 | 0 | 0 | 0 | 0 | 0 | 0 | 0 | 0 | 1 | 3.1 | 0 | 0 | 0 | 0 | 8 | 25 | 0 | 0 | 0 | 0 |
| 49 | 0 | 0 | 2 | 6.2 | 0 | 0 | 0 | 0 | 0 | 0 | 0 | 0 | 0 | 0 | 1 | 3.1 | 1 | 3.1 | 0 | 0 | 0 | 0 | 0 | 0 | 5 | 15.6 | 0 | 0 | 1 | 3.1 |
| 50 | 4 | 12.5 | 2 | 6.2 | 1 | 3.1 | 0 | 0 | 2 | 6.2 | 0 | 0 | 0 | 0 | 0 | 0 | 0 | 0 | 0 | 0 | 0 | 0 | 0 | 0 | 5 | 15.6 | 0 | 0 | 1 | 3.1 |
| 51 | 2 | 6.2 | 0 | 0 | 1 | 3.1 | 1 | 3.1 | 5 | 15.6 | 0 | 0 | 0 | 0 | 0 | 0 | 0 | 0 | 0 | 0 | 0 | 0 | 0 | 0 | 9 | 28.1 | 0 | 0 | 1 | 3.1 |
| 52 | 5 | 15.6 | 0 | 0 | 0 | 0 | 0 | 0 | 0 | 0 | 1 | 3.1 | 0 | 0 | 1 | 3.1 | 0 | 0 | 0 | 0 | 0 | 0 | 0 | 0 | 4 | 12.5 | 0 | 0 | 2 | 6.2 |
| 53 | 4 | 12.5 | 0 | 0 | 0 | 0 | 0 | 0 | 0 | 0 | 0 | 0 | 0 | 0 | 0 | 0 | 0 | 0 | 1 | 3.1 | 0 | 0 | 0 | 0 | 11 | 34.4 | 0 | 0 | 1 | 3.1 |
| 54 | 1 | 3.1 | 0 | 0 | 0 | 0 | 0 | 0 | 0 | 0 | 0 | 0 | 0 | 0 | 0 | 0 | 0 | 0 | 0 | 0 | 0 | 0 | 0 | 0 | 5 | 15.6 | 0 | 0 | 0 | 0 |
| 55 | 0 | 0 | 0 | 0 | 0 | 0 | 0 | 0 | 0 | 0 | 0 | 0 | 0 | 0 | 0 | 0 | 0 | 0 | 0 | 0 | 0 | 0 | 0 | 0 | 5 | 15.6 | 0 | 0 | 0 | 0 |
| 56 | 1 | 3.1 | 0 | 0 | 0 | 0 | 0 | 0 | 0 | 0 | 0 | 0 | 0 | 0 | 0 | 0 | 0 | 0 | 0 | 0 | 0 | 0 | 0 | 0 | 6 | 18.8 | 0 | 0 | 0 | 0 |
| 57 | 0 | 0 | 0 | 0 | 0 | 0 | 0 | 0 | 0 | 0 | 0 | 0 | 0 | 0 | 0 | 0 | 0 | 0 | 0 | 0 | 0 | 0 | 0 | 0 | 7 | 21.9 | 0 | 0 | 0 | 0 |
| 58 | 1 | 3.1 | 0 | 0 | 0 | 0 | 0 | 0 | 0 | 0 | 0 | 0 | 0 | 0 | 0 | 0 | 0 | 0 | 0 | 0 | 0 | 0 | 0 | 0 | 11 | 34.4 | 0 | 0 | 0 | 0 |
| 59 | 0 | 0 | 0 | 0 | 0 | 0 | 0 | 0 | 0 | 0 | 0 | 0 | 0 | 0 | 0 | 0 | 0 | 0 | 0 | 0 | 0 | 0 | 0 | 0 | 8 | 25 | 0 | 0 | 0 | 0 |
| 60 | 1 | 3.1 | 0 | 0 | 0 | 0 | 0 | 0 | 0 | 0 | 0 | 0 | 0 | 0 | 1 | 3.1 | 0 | 0 | 0 | 0 | 0 | 0 | 0 | 0 | 5 | 15.6 | 0 | 0 | 3 | 9.4 |
| | Nausea / Vomiting | | Runny Nose | | Severe Malaria - Convulsions / Seizures | | Severe Malaria - Dark Urine | | Severe Malaria - Other | | Severe Malaria - Prostration / Very weak, unable to stand | | Severe Malaria - Respiratory Distress | | Severe Malaria - Severe Pallor | | Severe Malnutrition | | Skin Irritation / Rash | | Tired | | Unable to eat, drink or suckle | | Vomiting | | Weak and unable to stand / move | | Other | |
| | # | % | # | % | # | % | # | % | # | % | # | % | # | % | # | % | # | % | # | % | # | % | # | % | # | % | # | % | # | % |
| min | 0 | 0 | 0 | 0 | 0 | 0 | 0 | 0 | 0 | 0 | 0 | 0 | 0 | 0 | 0 | 0 | 0 | 0 | 0 | 0 | 0 | 0 | 0 | 0 | 0 | 0 | 0 | 0 | 0 | 0 |

| | | | | | | | | | | | | | | | | | | | | | | | | | | | | | | | | | | | | | | | | | | | | | | | | | | | | | | | | | | | | |
|---|---|---|---|---|---|---|---|---|---|---|---|---|---|---|---|---|---|---|---|---|---|---|---|---|---|---|---|---|---|---|---|---|---|---|---|---|---|---|---|---|---|---|---|---|---|---|---|---|---|---|---|---|---|---|---|---|---|---|---|---|---|
| **max** | 33 | 2 | 6.5 | 3 | 9.4 | 2 | 6.2 | 2 | 6.2 | 2 | 6.5 | 1 | 3.2 | 14 | 43.8 | 1 | 3 | 6 | 18.8 | 1 | 3.1 | 1 | 3.1 | 1 | 3.1 | 20 | 62.5 | 8 | 25 | 10 | 31.2 | 5 | 15.6 | 2 | 6.2 | 1 | 3.1 | 2 | 6.2 | 10 | 31.2 | 1 | 3.1 | 1 | 3 | 1 | 3.1 | 1 | 3.1 | 1 | 3.1 | 4 | 12.5 | 1 | 3.1 | 17 | 53.1 | 1 | 3.2 | 4 | 12.5 |
| **median** | 32 | 0 | 0 | 0 | 0 | 0 | 0 | 0 | 0 | 0 | 0 | 0 | 0 | 2 | 6.2 | 0 | 0 | 0 | 0 | 0 | 0 | 0 | 0 | 0 | 0 | 3 | 9.4 | 1 | 3.1 | 0 | 0 | 0 | 0 | 0 | 0 | 0 | 0 | 0 | 0 | 0 | 0 | 0 | 0 | 0 | 0 | 0 | 0 | 0 | 0 | 0 | 0 | 0 | 0 | 0 | 0 | 4 | 12.5 | 0 | 0 | 1 | 3.1 |
| **mean** | 31.64 | 0.31 | 0.97 | 0.53 | 1.65 | 0.05 | 0.17 | 0.15 | 0.45 | 0.36 | 1.13 | 0.02 | 0.06 | 2.65 | 8.3 | 0.02 | 0.05 | 1.02 | 3.19 | 0.05 | 0.17 | 0.02 | 0.06 | 0.04 | 0.11 | 5.27 | 16.54 | 1.67 | 5.24 | 1.07 | 3.37 | 0.76 | 2.38 | 0.09 | 0.28 | 0.04 | 0.11 | 0.11 | 0.34 | 1.02 | 3.18 | 0.05 | 0.17 | 0.02 | 0.05 | 0.09 | 0.28 | 0.02 | 0.06 | 0.09 | 0.28 | 0.18 | 0.57 | 0.02 | 0.06 | 3.96 | 12.42 | 0.04 | 0.11 | 0.87 | 2.74 |
| **sd** | 1.38 | 0.63 | 1.98 | 0.88 | 2.75 | 0.3 | 0.92 | 0.49 | 1.51 | 0.62 | 1.94 | 0.13 | 0.43 | 3.38 | 10.57 | 0.13 | 0.4 | 1.66 | 5.19 | 0.23 | 0.71 | 0.13 | 0.42 | 0.19 | 0.59 | 5.46 | 17.01 | 2.06 | 6.43 | 2.08 | 6.5 | 1.26 | 3.93 | 0.4 | 1.23 | 0.19 | 0.59 | 0.37 | 1.14 | 2.26 | 7.05 | 0.23 | 0.71 | 0.13 | 0.4 | 0.29 | 0.89 | 0.13 | 0.42 | 0.29 | 0.9 | 0.61 | 1.91 | 0.13 | 0.42 | 3.83 | 12.02 | 0.19 | 0.6 | 1.07 | 3.35 |

Note: not every cluster had AE occurrences; Cluster 12 (SR), 21 (PBO), 22 (SR), 23 (PBO), 25 (SR) have zero AE occurrences at the baseline.

**Table B5: number of SAEs by symptom in clusters that had at least one SAE occurrences at the baseline (cluster numbers highlighted in red are SR clusters)**

| Cluster | # of unique subjects with at least one SAE | Abdominal Pain | Anorexia | Chills | Convulsions | Fever | Loss of appetite | Scratch on face | Severe Malaria - Prostration / Very weak, unable to stand | Severe Malaria - Severe Pallor | Tired | Unable to eat, drink or suckle | Vomiting | Weak and unable to stand / move | Other | category |
|---|---|---|---|---|---|---|---|---|---|---|---|---|---|---|---|---|
| 1 | 1 | 0 | 0 | 0 | 0 | 0 | 0 | 0 | 0 | 0 | 0 | 0 | 0 | 0 | 1 | Death |
| 22 | 1 | 0 | 0 | 0 | 0 | 0 | 0 | 0 | 0 | 0 | 0 | 0 | 0 | 0 | 1 | Life threatening |
| 26 | 1 | 0 | 0 | 0 | 0 | 0 | 0 | 0 | 0 | 0 | 0 | 0 | 0 | 0 | 1 | Hospitalization, Life threatening |
| 28 | 1 | 0 | 0 | 0 | 1 | 0 | 0 | 0 | 0 | 0 | 0 | 0 | 0 | 0 | 0 | Hospitalization, Life threatening |
| 29 | 1 | 0 | 1 | 1 | 0 | 1 | 0 | 0 | 0 | 0 | 1 | 0 | 1 | 0 | 0 | Hospitalization, Life threatening |
| 34 | 1 | 0 | 0 | 0 | 0 | 0 | 0 | 0 | 0 | 0 | 0 | 0 | 0 | 0 | 1 | Hospitalization, Event required an intervention in order to prevent a permanent incapacity. |
| 40 | 1 | 0 | 0 | 0 | 0 | 1 | 1 | 0 | 1 | 1 | 1 | 0 | 1 | 0 | 1 | Hospitalization, Life threatening |
| 47 | 1 | 0 | 0 | 0 | 0 | 1 | 0 | 0 | 0 | 0 | 0 | 1 | 0 | 1 | 1 | Hospitalization, Life threatening |
| 49 | 1 | 0 | 0 | 0 | 0 | 0 | 0 | 0 | 0 | 0 | 0 | 0 | 0 | 0 | 2 | Hospitalization |
| 51 | 1 | 0 | 0 | 0 | 0 | 0 | 0 | 1 | 0 | 0 | 0 | 0 | 0 | 0 | 0 | Hospitalization, Life threatening |
| 52 | 2 | 1 | 1 | 0 | 0 | 0 | 0 | 0 | 0 | 0 | 1 | 0 | 1 | 0 | 0 | Hospitalization, Life threatening |
| 60 | 1 | 1 | 0 | 0 | 0 | 1 | 1 | 0 | 0 | 0 | 0 | 0 | 1 | 0 | 1 | Hospitalization, Life threatening |

**Table B6: number of SAEs by symptom in the SR clusters that had at least one SAE occurrences at the baseline**

| Cluster | # of unique subjects with at least one SAE | Abdominal Pain | Anorexia | Chills | Convulsions | Fever | Loss of appetite | Scratch on face | Tired | Unable to eat, drink or suckle | Vomiting | Weak and unable to stand / move | Other | category |
|---|---|---|---|---|---|---|---|---|---|---|---|---|---|---|
| 22 | 1 | 0 | 0 | 0 | 0 | 0 | 0 | 0 | 0 | 0 | 0 | 0 | 1 | Life threatening |
| 26 | 1 | 0 | 0 | 0 | 0 | 0 | 0 | 0 | 0 | 0 | 0 | 0 | 1 | Hospitalization, Life threatening |
| 28 | 1 | 0 | 0 | 0 | 1 | 0 | 0 | 0 | 0 | 0 | 0 | 0 | 0 | Hospitalization, Life threatening |
| 29 | 1 | 0 | 1 | 1 | 0 | 1 | 0 | 0 | 1 | 0 | 1 | 0 | 0 | Hospitalization, Life threatening |
| 47 | 1 | 0 | 0 | 0 | 0 | 1 | 0 | 0 | 0 | 1 | 0 | 1 | 1 | Hospitalization, Life threatening |
| 49 | 1 | 0 | 0 | 0 | 0 | 0 | 0 | 0 | 0 | 0 | 0 | 0 | 2 | Hospitalization |
| 51 | 1 | 0 | 0 | 0 | 0 | 0 | 0 | 1 | 0 | 0 | 0 | 0 | 0 | Hospitalization, Life threatening |

| | | | | | | | | | | | | | | |
|---|---|---|---|---|---|---|---|---|---|---|---|---|---|---|
| 60 | 1 | 1 | 0 | 0 | 0 | 1 | 1 | 0 | 0 | 0 | 1 | 0 | 1 | Hospitalization, Life threatening |

**Table B7: number of SAEs by symptom in the placebo clusters that had at least one SAE occurrences at the baseline**

| Cluster | # of unique subjects with at least one SAE | Abdominal Pain | Anorexia | Fever | Loss of appetite | Severe Malaria - Prostration / Very weak, unable to stand | Severe Malaria - Severe Pallor | Tired | Vomiting | Other | category |
|---|---|---|---|---|---|---|---|---|---|---|---|
| 1 | 1 | 0 | 0 | 0 | 0 | 0 | 0 | 0 | 0 | 1 | Death |
| 34 | 1 | 0 | 0 | 0 | 0 | 0 | 0 | 0 | 0 | 1 | Hospitalization, Event required an intervention in order to prevent a permanent incapacity. |
| 40 | 1 | 0 | 0 | 1 | 1 | 1 | 1 | 1 | 1 | 1 | Hospitalization, Life threatening |
| 52 | 2 | 1 | 1 | 0 | 0 | 0 | 0 | 1 | 1 | 0 | Hospitalization, Life threatening |

**Table B8: frequency and percentage of subjects having at least one SAE occurrence by symptom in each cluster at the baseline (clusters highlighted in red are SR clusters)**

| Cluster | Total # of subjects | Abdominal Pain | | Anorexia | | Chills | | Convulsions | | Fever | | Loss of appetite | | Scratch on face | | Severe Malaria - Prostration / Very weak, unable to stand | | Severe Malaria - Severe Pallor | | Tired | | Unable to eat, drink or suckle | | Vomiting | | Weak and unable to stand / move | | Other | |
|---|---|---|---|---|---|---|---|---|---|---|---|---|---|---|---|---|---|---|---|---|---|---|---|---|---|---|---|---|---|
| | | # | % | # | % | # | % | # | % | # | % | # | % | # | % | # | % | # | % | # | % | # | % | # | % | # | % | # | % |
| 1 | 32 | 0 | 0 | 0 | 0 | 0 | 0 | 0 | 0 | 0 | 0 | 0 | 0 | 0 | 0 | 0 | 0 | 0 | 0 | 0 | 0 | 0 | 0 | 0 | 0 | 0 | 0 | 1 | 3.1 |
| 22 | 32 | 0 | 0 | 0 | 0 | 0 | 0 | 0 | 0 | 0 | 0 | 0 | 0 | 0 | 0 | 0 | 0 | 0 | 0 | 0 | 0 | 0 | 0 | 0 | 0 | 0 | 0 | 1 | 3.1 |
| 26 | 32 | 0 | 0 | 0 | 0 | 0 | 0 | 0 | 0 | 0 | 0 | 0 | 0 | 0 | 0 | 0 | 0 | 0 | 0 | 0 | 0 | 0 | 0 | 0 | 0 | 0 | 0 | 1 | 3.1 |
| 28 | 31 | 0 | 0 | 0 | 0 | 0 | 0 | 1 | 3.2 | 0 | 0 | 0 | 0 | 0 | 0 | 0 | 0 | 0 | 0 | 0 | 0 | 0 | 0 | 0 | 0 | 0 | 0 | 0 | 0 |
| 29 | 32 | 0 | 0 | 1 | 3.1 | 1 | 3.1 | 0 | 0 | 1 | 3.1 | 0 | 0 | 0 | 0 | 0 | 0 | 0 | 0 | 1 | 3.1 | 0 | 0 | 1 | 3.1 | 0 | 0 | 0 | 0 |
| 34 | 32 | 0 | 0 | 0 | 0 | 0 | 0 | 0 | 0 | 0 | 0 | 0 | 0 | 0 | 0 | 0 | 0 | 0 | 0 | 0 | 0 | 0 | 0 | 0 | 0 | 0 | 0 | 1 | 3.1 |
| 40 | 33 | 0 | 0 | 0 | 0 | 0 | 0 | 0 | 0 | 1 | 3 | 1 | 3 | 0 | 0 | 1 | 3 | 1 | 3 | 1 | 3 | 0 | 0 | 1 | 3 | 0 | 0 | 1 | 3 |
| 47 | 32 | 0 | 0 | 0 | 0 | 0 | 0 | 0 | 0 | 1 | 3.1 | 0 | 0 | 0 | 0 | 0 | 0 | 0 | 0 | 0 | 0 | 1 | 3.1 | 0 | 0 | 1 | 3.1 | 1 | 3.1 |
| 49 | 32 | 0 | 0 | 0 | 0 | 0 | 0 | 0 | 0 | 0 | 0 | 0 | 0 | 0 | 0 | 0 | 0 | 0 | 0 | 0 | 0 | 0 | 0 | 0 | 0 | 0 | 0 | 1 | 3.1 |
| 51 | 32 | 0 | 0 | 0 | 0 | 0 | 0 | 0 | 0 | 0 | 0 | 0 | 0 | 1 | 3.1 | 0 | 0 | 0 | 0 | 0 | 0 | 0 | 0 | 0 | 0 | 0 | 0 | 0 | 0 |
| 52 | 32 | 1 | 3.1 | 1 | 3.1 | 0 | 0 | 0 | 0 | 0 | 0 | 0 | 0 | 0 | 0 | 0 | 0 | 0 | 0 | 1 | 3.1 | 0 | 0 | 1 | 3.1 | 0 | 0 | 0 | 0 |
| 60 | 32 | 1 | 3.1 | 0 | 0 | 0 | 0 | 0 | 0 | 1 | 3.1 | 1 | 3.1 | 0 | 0 | 0 | 0 | 0 | 0 | 0 | 0 | 0 | 0 | 1 | 3.1 | 0 | 0 | 1 | 3.1 |

#### Table I1: number of AEs by symptom per cluster during the intervention (cluster numbers highlighted in red are SR clusters)

| Cluster | # of unique subjects with at least one AE | Abdominal Pain | Aches | Anorexia | Chills | Convulsions | Cough | Diarrhea | Dizziness | Eye Irritation | Fever | Headache | Jaundice | Loss of appetite | Nausea / Vomiting | Respiratory distress | Runny Nose | Severe Malaria - Convulsions / Seizures | Severe Malaria - Dark Urine | Severe Malaria - Other | Severe Malaria - Prostration / Very weak, unable to stand | Severe Malaria - Severe Pallor | Severe Malaria - Unconsciousness / Drowsiness | Skin Irritation / Rash | Tired | Unconsciousness / Drowsiness | Vomiting | Weak and unable to stand / move | Other |
|---|---|---|---|---|---|---|---|---|---|---|---|---|---|---|---|---|---|---|---|---|---|---|---|---|---|---|---|---|---|
| 1 | 27 | 3 | 0 | 0 | 1 | 1 | 26 | 0 | 0 | 0 | 53 | 14 | 0 | 5 | 0 | 0 | 0 | 0 | 0 | 0 | 0 | 0 | 0 | 1 | 0 | 0 | 13 | 0 | 4 |
| 2 | 21 | 2 | 0 | 0 | 2 | 0 | 10 | 0 | 0 | 0 | 29 | 2 | 0 | 2 | 0 | 0 | 0 | 0 | 0 | 0 | 0 | 0 | 0 | 0 | 0 | 0 | 6 | 0 | 4 |
| 3 | 30 | 9 | 0 | 0 | 0 | 0 | 25 | 0 | 0 | 0 | 50 | 21 | 0 | 2 | 0 | 0 | 0 | 0 | 0 | 0 | 0 | 0 | 0 | 0 | 0 | 0 | 9 | 0 | 10 |
| 4 | 22 | 7 | 0 | 0 | 0 | 0 | 22 | 0 | 0 | 0 | 28 | 3 | 0 | 4 | 0 | 0 | 0 | 0 | 0 | 0 | 0 | 0 | 0 | 0 | 0 | 0 | 8 | 0 | 5 |
| 5 | 25 | 8 | 0 | 0 | 1 | 0 | 14 | 0 | 0 | 0 | 50 | 14 | 0 | 0 | 0 | 0 | 0 | 0 | 0 | 0 | 0 | 0 | 0 | 0 | 0 | 0 | 17 | 0 | 11 |
| 6 | 30 | 6 | 0 | 0 | 4 | 0 | 32 | 1 | 0 | 0 | 79 | 26 | 0 | 1 | 0 | 0 | 0 | 0 | 0 | 3 | 0 | 0 | 0 | 0 | 0 | 0 | 27 | 0 | 16 |
| 7 | 27 | 3 | 0 | 0 | 2 | 0 | 14 | 2 | 0 | 0 | 47 | 13 | 0 | 0 | 0 | 0 | 0 | 0 | 0 | 1 | 0 | 0 | 0 | 0 | 0 | 0 | 23 | 0 | 11 |
| 8 | 11 | 0 | 0 | 0 | 1 | 0 | 5 | 1 | 0 | 0 | 2 | 4 | 0 | 0 | 0 | 0 | 0 | 0 | 0 | 14 | 0 | 0 | 0 | 0 | 4 | 0 | 2 | 0 | 1 |
| 9 | 1 | 0 | 0 | 0 | 0 | 0 | 0 | 0 | 0 | 0 | 0 | 0 | 0 | 0 | 0 | 0 | 0 | 0 | 0 | 1 | 0 | 0 | 0 | 0 | 0 | 0 | 0 | 0 | 0 |
| 10 | 4 | 1 | 0 | 0 | 0 | 0 | 1 | 0 | 0 | 0 | 0 | 2 | 0 | 0 | 0 | 0 | 0 | 0 | 0 | 3 | 0 | 0 | 0 | 0 | 0 | 0 | 0 | 0 | 0 |
| 11 | 4 | 0 | 0 | 0 | 0 | 0 | 0 | 0 | 0 | 0 | 0 | 0 | 0 | 0 | 0 | 0 | 0 | 0 | 0 | 4 | 0 | 0 | 0 | 0 | 0 | 0 | 0 | 0 | 0 |
| 12 | 15 | 2 | 0 | 0 | 0 | 0 | 3 | 1 | 0 | 0 | 18 | 1 | 0 | 0 | 0 | 0 | 0 | 0 | 0 | 0 | 0 | 0 | 0 | 0 | 0 | 0 | 0 | 0 | 0 |
| 13 | 9 | 0 | 0 | 0 | 0 | 0 | 1 | 1 | 0 | 0 | 8 | 2 | 0 | 0 | 0 | 0 | 0 | 0 | 0 | 0 | 0 | 0 | 0 | 0 | 0 | 0 | 1 | 0 | 1 |
| 14 | 18 | 0 | 0 | 0 | 0 | 0 | 5 | 0 | 0 | 0 | 27 | 3 | 0 | 0 | 0 | 0 | 0 | 0 | 0 | 0 | 0 | 0 | 0 | 0 | 0 | 0 | 1 | 0 | 0 |
| 15 | 11 | 0 | 0 | 0 | 0 | 0 | 2 | 2 | 0 | 0 | 8 | 0 | 0 | 1 | 0 | 0 | 0 | 0 | 0 | 0 | 0 | 0 | 0 | 0 | 0 | 0 | 3 | 0 | 2 |
| 16 | 23 | 1 | 0 | 0 | 3 | 0 | 14 | 5 | 0 | 1 | 26 | 14 | 0 | 0 | 0 | 0 | 0 | 0 | 0 | 19 | 0 | 4 | 0 | 0 | 4 | 0 | 24 | 0 | 0 |
| 17 | 23 | 7 | 3 | 0 | 1 | 0 | 7 | 2 | 2 | 0 | 31 | 12 | 0 | 2 | 0 | 0 | 0 | 0 | 0 | 12 | 0 | 5 | 0 | 0 | 2 | 0 | 16 | 0 | 4 |
| 18 | 17 | 2 | 0 | 0 | 0 | 0 | 11 | 1 | 0 | 0 | 15 | 2 | 0 | 0 | 0 | 0 | 0 | 0 | 0 | 0 | 0 | 0 | 0 | 0 | 0 | 0 | 5 | 0 | 2 |
| 19 | 21 | 0 | 0 | 0 | 0 | 0 | 6 | 1 | 0 | 0 | 10 | 4 | 0 | 0 | 0 | 0 | 0 | 0 | 0 | 0 | 0 | 0 | 0 | 3 | 0 | 0 | 6 | 0 | 7 |
| 20 | 7 | 0 | 0 | 0 | 0 | 0 | 0 | 0 | 0 | 0 | 0 | 0 | 0 | 0 | 0 | 0 | 0 | 0 | 0 | 9 | 0 | 0 | 0 | 0 | 0 | 0 | 0 | 0 | 0 |
| 21 | 21 | 6 | 1 | 0 | 0 | 0 | 5 | 2 | 0 | 0 | 20 | 13 | 0 | 1 | 0 | 0 | 0 | 0 | 0 | 0 | 0 | 0 | 1 | 0 | 0 | 1 | 13 | 0 | 4 |
| 22 | 13 | 6 | 1 | 0 | 2 | 0 | 7 | 1 | 0 | 0 | 17 | 11 | 0 | 2 | 0 | 0 | 0 | 0 | 0 | 1 | 0 | 0 | 0 | 0 | 0 | 0 | 13 | 0 | 0 |
| 23 | 19 | 11 | 0 | 0 | 0 | 0 | 5 | 1 | 0 | 0 | 24 | 12 | 0 | 3 | 0 | 0 | 0 | 0 | 0 | 0 | 0 | 0 | 0 | 3 | 0 | 0 | 10 | 0 | 7 |
| 24 | 14 | 5 | 1 | 0 | 0 | 0 | 5 | 0 | 0 | 0 | 15 | 5 | 1 | 0 | 0 | 0 | 0 | 0 | 0 | 0 | 0 | 0 | 0 | 1 | 0 | 0 | 5 | 0 | 2 |
| 25 | 21 | 3 | 0 | 0 | 0 | 0 | 6 | 0 | 0 | 0 | 21 | 3 | 0 | 1 | 0 | 0 | 0 | 0 | 0 | 0 | 0 | 0 | 0 | 1 | 0 | 0 | 5 | 0 | 3 |
| 26 | 17 | 3 | 1 | 1 | 0 | 0 | 4 | 2 | 1 | 0 | 15 | 6 | 0 | 4 | 0 | 0 | 0 | 0 | 0 | 0 | 0 | 0 | 0 | 0 | 0 | 0 | 3 | 0 | 1 |
| 27 | 18 | 2 | 0 | 0 | 0 | 0 | 7 | 0 | 0 | 1 | 30 | 8 | 0 | 1 | 0 | 0 | 0 | 0 | 0 | 0 | 4 | 2 | 2 | 0 | 0 | 2 | 6 | 2 | 7 |
| 28 | 22 | 6 | 0 | 0 | 0 | 1 | 5 | 0 | 0 | 0 | 24 | 13 | 0 | 0 | 0 | 0 | 0 | 0 | 0 | 0 | 0 | 0 | 0 | 0 | 0 | 0 | 9 | 0 | 5 |
| 29 | 18 | 1 | 1 | 0 | 0 | 0 | 2 | 0 | 1 | 0 | 17 | 7 | 0 | 4 | 0 | 0 | 0 | 0 | 0 | 0 | 0 | 0 | 0 | 0 | 0 | 0 | 10 | 0 | 1 |
| 30 | 12 | 1 | 0 | 0 | 0 | 0 | 1 | 0 | 0 | 0 | 17 | 3 | 0 | 0 | 0 | 0 | 0 | 0 | 0 | 0 | 0 | 0 | 0 | 0 | 0 | 0 | 0 | 0 | 0 |
| 31 | 15 | 2 | 0 | 0 | 1 | 0 | 1 | 0 | 2 | 0 | 22 | 9 | 0 | 0 | 0 | 0 | 0 | 2 | 0 | 0 | 0 | 0 | 0 | 0 | 1 | 0 | 10 | 0 | 3 |
| 32 | 26 | 7 | 0 | 2 | 0 | 0 | 26 | 5 | 0 | 0 | 49 | 5 | 0 | 8 | 0 | 0 | 0 | 0 | 0 | 0 | 0 | 0 | 0 | 0 | 0 | 0 | 20 | 0 | 16 |
| 33 | 28 | 8 | 0 | 0 | 2 | 0 | 15 | 2 | 1 | 0 | 54 | 15 | 0 | 3 | 0 | 0 | 0 | 0 | 0 | 0 | 0 | 0 | 0 | 1 | 0 | 0 | 26 | 0 | 13 |
| 34 | 28 | 6 | 0 | 2 | 3 | 0 | 13 | 2 | 0 | 0 | 49 | 17 | 0 | 6 | 0 | 0 | 0 | 0 | 0 | 0 | 0 | 0 | 0 | 2 | 0 | 0 | 21 | 0 | 15 |
| 35 | 18 | 2 | 0 | 0 | 0 | 0 | 6 | 2 | 0 | 0 | 1 | 12 | 0 | 0 | 0 | 0 | 0 | 0 | 0 | 1 | 0 | 0 | 0 | 1 | 0 | 0 | 14 | 0 | 4 |
| 36 | 30 | 7 | 3 | 0 | 0 | 0 | 14 | 2 | 0 | 0 | 71 | 21 | 0 | 6 | 0 | 0 | 1 | 0 | 1 | 0 | 0 | 0 | 0 | 2 | 0 | 0 | 30 | 0 | 21 |
| 37 | 14 | 0 | 0 | 0 | 1 | 0 | 5 | 1 | 0 | 0 | 1 | 6 | 0 | 0 | 0 | 0 | 0 | 0 | 0 | 0 | 0 | 0 | 0 | 0 | 0 | 0 | 8 | 0 | 3 |
| 38 | 22 | 0 | 0 | 0 | 1 | 0 | 7 | 5 | 2 | 0 | 3 | 26 | 0 | 0 | 0 | 0 | 0 | 0 | 0 | 0 | 0 | 0 | 0 | 0 | 0 | 0 | 17 | 0 | 3 |
| 39 | 20 | 0 | 0 | 0 | 2 | 0 | 4 | 1 | 1 | 0 | 4 | 11 | 0 | 0 | 0 | 0 | 0 | 0 | 0 | 0 | 0 | 0 | 0 | 0 | 0 | 0 | 12 | 0 | 2 |
| 40 | 29 | 5 | 0 | 2 | 0 | 0 | 18 | 4 | 0 | 0 | 68 | 20 | 0 | 10 | 0 | 0 | 0 | 0 | 0 | 0 | 0 | 1 | 0 | 0 | 1 | 0 | 22 | 0 | 16 |
| 41 | 12 | 1 | 0 | 0 | 0 | 0 | 0 | 0 | 0 | 0 | 7 | 4 | 0 | 0 | 0 | 0 | 0 | 0 | 0 | 7 | 0 | 0 | 0 | 0 | 0 | 0 | 1 | 0 | 0 |
| 42 | 8 | 0 | 0 | 0 | 0 | 0 | 1 | 0 | 0 | 0 | 6 | 3 | 0 | 0 | 0 | 0 | 0 | 0 | 0 | 6 | 0 | 0 | 0 | 0 | 0 | 0 | 0 | 0 | 1 |
| 43 | 11 | 0 | 0 | 0 | 0 | 0 | 0 | 0 | 0 | 0 | 7 | 2 | 0 | 0 | 0 | 0 | 0 | 0 | 0 | 10 | 0 | 0 | 0 | 0 | 0 | 0 | 1 | 0 | 0 |
| 44 | 5 | 0 | 0 | 0 | 0 | 0 | 1 | 0 | 0 | 0 | 5 | 0 | 0 | 1 | 0 | 0 | 0 | 0 | 0 | 0 | 0 | 0 | 0 | 0 | 0 | 0 | 2 | 0 | 0 |
| 45 | 5 | 1 | 0 | 0 | 0 | 0 | 0 | 0 | 0 | 0 | 6 | 3 | 0 | 0 | 0 | 0 | 0 | 0 | 0 | 0 | 0 | 0 | 0 | 0 | 0 | 0 | 2 | 0 | 1 |
| 46 | 31 | 12 | 2 | 0 | 1 | 0 | 15 | 7 | 1 | 0 | 58 | 41 | 0 | 5 | 0 | 0 | 0 | 0 | 0 | 1 | 1 | 0 | 0 | 0 | 0 | 0 | 20 | 0 | 23 |
| 47 | 11 | 3 | 0 | 0 | 0 | 0 | 4 | 3 | 0 | 0 | 9 | 4 | 0 | 1 | 0 | 1 | 0 | 0 | 0 | 0 | 0 | 0 | 0 | 0 | 0 | 0 | 8 | 0 | 2 |
| 48 | 28 | 7 | 3 | 0 | 4 | 0 | 15 | 3 | 0 | 0 | 64 | 19 | 0 | 7 | 0 | 0 | 0 | 0 | 0 | 0 | 2 | 0 | 0 | 1 | 0 | 0 | 29 | 0 | 3 |
| 49 | 16 | 1 | 0 | 0 | 2 | 0 | 10 | 8 | 0 | 0 | 14 | 7 | 0 | 1 | 0 | 1 | 0 | 0 | 0 | 0 | 0 | 0 | 0 | 0 | 2 | 0 | 14 | 0 | 13 |
| 50 | 26 | 10 | 2 | 0 | 0 | 0 | 16 | 6 | 0 | 0 | 45 | 18 | 0 | 7 | 0 | 0 | 0 | 0 | 1 | 1 | 0 | 0 | 0 | 0 | 0 | 0 | 17 | 0 | 6 |
| 51 | 29 | 7 | 3 | 0 | 4 | 0 | 18 | 2 | 1 | 0 | 62 | 35 | 1 | 7 | 1 | 0 | 0 | 0 | 1 | 0 | 2 | 0 | 0 | 0 | 0 | 0 | 33 | 0 | 14 |
| 52 | 32 | 16 | 7 | 1 | 12 | 0 | 103 | 14 | 1 | 0 | 236 | 15 | 0 | 47 | 0 | 0 | 2 | 0 | 0 | 0 | 0 | 0 | 0 | 0 | 3 | 0 | 64 | 0 | 31 |
| 53 | 32 | 20 | 8 | 0 | 8 | 0 | 69 | 5 | 1 | 0 | 186 | 19 | 0 | 45 | 0 | 0 | 0 | 0 | 0 | 0 | 0 | 0 | 0 | 4 | 0 | 0 | 44 | 0 | 21 |
| 54 | 14 | 0 | 0 | 0 | 0 | 0 | 0 | 0 | 0 | 0 | 6 | 1 | 0 | 0 | 0 | 0 | 0 | 0 | 0 | 0 | 0 | 0 | 0 | 0 | 1 | 0 | 11 | 0 | 0 |
| 55 | 12 | 0 | 0 | 0 | 0 | 0 | 0 | 0 | 0 | 0 | 1 | 5 | 0 | 0 | 0 | 0 | 0 | 0 | 0 | 0 | 0 | 0 | 0 | 0 | 0 | 0 | 10 | 0 | 1 |
| 56 | 20 | 0 | 0 | 0 | 0 | 0 | 1 | 0 | 0 | 0 | 3 | 2 | 0 | 0 | 0 | 0 | 0 | 0 | 0 | 0 | 0 | 0 | 0 | 0 | 0 | 0 | 20 | 0 | 1 |
| 57 | 21 | 0 | 0 | 0 | 0 | 0 | 1 | 0 | 0 | 0 | 9 | 4 | 0 | 0 | 0 | 0 | 0 | 0 | 0 | 0 | 0 | 0 | 0 | 0 | 0 | 0 | 15 | 0 | 0 |
| 58 | 14 | 0 | 0 | 0 | 0 | 0 | 0 | 0 | 0 | 0 | 5 | 2 | 0 | 0 | 0 | 0 | 0 | 0 | 0 | 0 | 0 | 0 | 0 | 1 | 0 | 0 | 12 | 0 | 1 |

| | | | | | | | | | | | | | | | | | | | | | | | | | | | | | |
|---|---|---|---|---|---|---|---|---|---|---|---|---|---|---|---|---|---|---|---|---|---|---|---|---|---|---|---|---|---|
| 59 | 13 | 0 | 0 | 0 | 0 | 0 | 0 | 0 | 0 | 0 | 5 | 4 | 0 | 0 | 0 | 0 | 0 | 0 | 0 | 0 | 0 | 0 | 0 | 0 | 0 | 0 | 5 | 0 | 0 |
| 60 | 16 | 3 | 0 | 1 | 0 | 0 | 14 | 0 | 0 | 0 | 17 | 3 | 2 | 1 | 0 | 0 | 2 | 0 | 0 | 0 | 1 | 0 | 0 | 1 | 0 | 0 | 14 | 0 | 6 |
| | # of unique subjects with at least one AE | **Abdominal Pain** | **Aches** | **Anorexia** | **Chills** | **Convulsions** | **Cough** | **Diarrhea** | **Dizziness** | **Eye Irritation** | **Fever** | **Headache** | **Jaundice** | **Loss of appetite** | **Nausea / Vomiting** | **Respiratory distress** | **Runny Nose** | **Severe Malaria - Convulsions / Seizures** | **Severe Malaria - Dark Urine** | **Severe Malaria - Other** | **Severe Malaria - Prostration / Very weak, unable to stand** | **Severe Malaria - Severe Pallor** | **Severe Malaria - Unconsciousness / Drowsiness** | **Skin Irritation / Rash** | **Tired** | **Unconsciousness / Drowsiness** | **Vomiting** | **Weak and unable to stand / move** | **Other** |
| **min** | 1 | 0 | 0 | 0 | 0 | 0 | 0 | 0 | 0 | 0 | 0 | 0 | 0 | 0 | 0 | 0 | 0 | 0 | 0 | 0 | 0 | 0 | 0 | 0 | 0 | 0 | 0 | 0 | 0 |
| **max** | 32 | 20 | 8 | 2 | 12 | 1 | 103 | 14 | 2 | 1 | 236 | 41 | 2 | 47 | 1 | 1 | 2 | 2 | 1 | 19 | 4 | 5 | 2 | 4 | 4 | 2 | 64 | 2 | 31 |
| **median** | 18 | 2 | 0 | 0 | 0 | 0 | 5 | 1 | 0 | 0 | 17 | 5.5 | 0 | 0 | 0 | 0 | 0 | 0 | 0 | 0 | 0 | 0 | 0 | 0 | 0 | 0 | 10 | 0 | 3 |
| **mean** | 18.45 | 3.55 | 0.6 | 0.15 | 0.97 | 0.03 | 10.37 | 1.58 | 0.23 | 0.03 | 29.23 | 9.18 | 0.07 | 3.13 | 0.02 | 0.03 | 0.08 | 0.03 | 0.05 | 1.55 | 0.17 | 0.2 | 0.05 | 0.37 | 0.3 | 0.05 | 12.28 | 0.03 | 5.47 |
| **sd** | 8.01 | 4.28 | 1.54 | 0.48 | 2.06 | 0.18 | 16.47 | 2.53 | 0.53 | 0.18 | 40.43 | 8.84 | 0.31 | 8.41 | 0.13 | 0.18 | 0.38 | 0.26 | 0.22 | 3.82 | 0.64 | 0.86 | 0.29 | 0.84 | 0.89 | 0.29 | 11.86 | 0.26 | 6.98 |
| **total** | 1107 | 213 | 36 | 9 | 58 | 2 | 622 | 95 | 14 | 2 | 1754 | 551 | 4 | 188 | 1 | 2 | 5 | 2 | 3 | 93 | 10 | 12 | 3 | 22 | 18 | 3 | 737 | 2 | 328 |

#### Table I2: number of AEs by symptom in the SR clusters during the intervention

| Cluster | # of unique subjects with at least one AE | Abdominal Pain | Aches | Anorexia | Chills | Convulsions | Cough | Diarrhea | Dizziness | Eye Irritation | Fever | Headache | Jaundice | Loss of appetite | Nausea / Vomiting | Respiratory distress | Runny Nose | Severe Malaria - Dark Urine | Severe Malaria - Other | Severe Malaria - Prostration / Very weak, unable to stand | Severe Malaria - Severe Pallor | Severe Malaria - Unconsciousness / Drowsiness | Skin Irritation / Rash | Tired | Unconsciousness / Drowsiness | Vomiting | Weak and unable to stand / move | Other |
|---|---|---|---|---|---|---|---|---|---|---|---|---|---|---|---|---|---|---|---|---|---|---|---|---|---|---|---|---|
| 4 | 22 | 7 | 0 | 0 | 0 | 0 | 22 | 0 | 0 | 0 | 28 | 3 | 0 | 4 | 0 | 0 | 0 | 0 | 0 | 0 | 0 | 0 | 0 | 0 | 0 | 8 | 0 | 5 |
| 5 | 25 | 8 | 0 | 0 | 1 | 0 | 14 | 0 | 0 | 0 | 50 | 14 | 0 | 0 | 0 | 0 | 0 | 0 | 0 | 0 | 0 | 0 | 0 | 0 | 0 | 17 | 0 | 11 |
| 6 | 30 | 6 | 0 | 0 | 4 | 0 | 32 | 1 | 0 | 0 | 79 | 26 | 0 | 1 | 0 | 0 | 0 | 0 | 3 | 0 | 0 | 0 | 0 | 0 | 0 | 27 | 0 | 16 |
| 9 | 1 | 0 | 0 | 0 | 0 | 0 | 0 | 0 | 0 | 0 | 0 | 0 | 0 | 0 | 0 | 0 | 0 | 0 | 1 | 0 | 0 | 0 | 0 | 0 | 0 | 0 | 0 | 0 |
| 10 | 4 | 1 | 0 | 0 | 0 | 0 | 1 | 0 | 0 | 0 | 0 | 2 | 0 | 0 | 0 | 0 | 0 | 0 | 3 | 0 | 0 | 0 | 0 | 0 | 0 | 0 | 0 | 0 |
| 11 | 4 | 0 | 0 | 0 | 0 | 0 | 0 | 0 | 0 | 0 | 0 | 0 | 0 | 0 | 0 | 0 | 0 | 0 | 4 | 0 | 0 | 0 | 0 | 0 | 0 | 0 | 0 | 0 |
| 12 | 15 | 2 | 0 | 0 | 0 | 0 | 3 | 1 | 0 | 0 | 18 | 1 | 0 | 0 | 0 | 0 | 0 | 0 | 0 | 0 | 0 | 0 | 0 | 0 | 0 | 0 | 0 | 0 |
| 15 | 11 | 0 | 0 | 0 | 0 | 0 | 2 | 2 | 0 | 0 | 8 | 0 | 0 | 1 | 0 | 0 | 0 | 0 | 0 | 0 | 0 | 0 | 0 | 0 | 0 | 3 | 0 | 2 |
| 22 | 13 | 6 | 1 | 0 | 2 | 0 | 7 | 1 | 0 | 0 | 17 | 11 | 0 | 2 | 0 | 0 | 0 | 0 | 1 | 0 | 0 | 0 | 0 | 0 | 0 | 13 | 0 | 0 |
| 24 | 14 | 5 | 1 | 0 | 0 | 0 | 5 | 0 | 0 | 0 | 15 | 5 | 1 | 0 | 0 | 0 | 0 | 0 | 0 | 0 | 0 | 0 | 1 | 0 | 0 | 5 | 0 | 2 |
| 25 | 21 | 3 | 0 | 0 | 0 | 0 | 6 | 0 | 0 | 0 | 21 | 3 | 0 | 1 | 0 | 0 | 0 | 0 | 0 | 0 | 0 | 0 | 1 | 0 | 0 | 5 | 0 | 3 |
| 26 | 17 | 3 | 1 | 1 | 0 | 0 | 4 | 2 | 1 | 0 | 15 | 6 | 0 | 4 | 0 | 0 | 0 | 0 | 0 | 0 | 0 | 0 | 0 | 0 | 0 | 3 | 0 | 1 |
| 27 | 18 | 2 | 0 | 0 | 0 | 0 | 7 | 0 | 0 | 1 | 30 | 8 | 0 | 1 | 0 | 0 | 0 | 0 | 0 | 4 | 2 | 2 | 0 | 0 | 2 | 6 | 2 | 7 |
| 28 | 22 | 6 | 0 | 0 | 0 | 1 | 5 | 0 | 0 | 0 | 24 | 13 | 0 | 0 | 0 | 0 | 0 | 0 | 0 | 0 | 0 | 0 | 0 | 0 | 0 | 9 | 0 | 5 |
| 29 | 18 | 1 | 1 | 0 | 0 | 0 | 2 | 0 | 1 | 0 | 17 | 7 | 0 | 4 | 0 | 0 | 0 | 0 | 0 | 0 | 0 | 0 | 0 | 0 | 0 | 10 | 0 | 1 |
| 32 | 26 | 7 | 0 | 2 | 0 | 0 | 26 | 5 | 0 | 0 | 49 | 5 | 0 | 8 | 0 | 0 | 0 | 0 | 0 | 0 | 0 | 0 | 0 | 0 | 0 | 20 | 0 | 16 |
| 36 | 30 | 7 | 3 | 0 | 0 | 0 | 14 | 2 | 0 | 0 | 71 | 21 | 0 | 6 | 0 | 0 | 1 | 1 | 0 | 0 | 0 | 0 | 2 | 0 | 0 | 30 | 0 | 21 |
| 37 | 14 | 0 | 0 | 0 | 1 | 0 | 5 | 1 | 0 | 0 | 1 | 6 | 0 | 0 | 0 | 0 | 0 | 0 | 0 | 0 | 0 | 0 | 0 | 0 | 0 | 8 | 0 | 3 |
| 42 | 8 | 0 | 0 | 0 | 0 | 0 | 1 | 0 | 0 | 0 | 6 | 3 | 0 | 0 | 0 | 0 | 0 | 0 | 6 | 0 | 0 | 0 | 0 | 0 | 0 | 0 | 0 | 1 |
| 43 | 11 | 0 | 0 | 0 | 0 | 0 | 0 | 0 | 0 | 0 | 7 | 2 | 0 | 0 | 0 | 0 | 0 | 0 | 10 | 0 | 0 | 0 | 0 | 0 | 0 | 1 | 0 | 0 |
| 45 | 5 | 1 | 0 | 0 | 0 | 0 | 0 | 0 | 0 | 0 | 6 | 3 | 0 | 0 | 0 | 0 | 0 | 0 | 0 | 0 | 0 | 0 | 0 | 0 | 0 | 2 | 0 | 1 |
| 46 | 31 | 12 | 2 | 0 | 1 | 0 | 15 | 7 | 1 | 0 | 58 | 41 | 0 | 5 | 0 | 0 | 0 | 0 | 1 | 1 | 0 | 0 | 0 | 0 | 0 | 20 | 0 | 23 |
| 47 | 11 | 3 | 0 | 0 | 0 | 0 | 4 | 3 | 0 | 0 | 9 | 4 | 0 | 1 | 0 | 1 | 0 | 0 | 0 | 0 | 0 | 0 | 0 | 0 | 0 | 8 | 0 | 2 |
| 48 | 28 | 7 | 3 | 0 | 4 | 0 | 15 | 3 | 0 | 0 | 64 | 19 | 0 | 7 | 0 | 0 | 0 | 0 | 0 | 2 | 0 | 0 | 1 | 0 | 0 | 29 | 0 | 3 |
| 49 | 16 | 1 | 0 | 0 | 2 | 0 | 10 | 8 | 0 | 0 | 14 | 7 | 0 | 1 | 0 | 1 | 0 | 0 | 0 | 0 | 0 | 0 | 0 | 2 | 0 | 14 | 0 | 13 |
| 51 | 29 | 7 | 3 | 0 | 4 | 0 | 18 | 2 | 1 | 0 | 62 | 35 | 1 | 7 | 1 | 0 | 0 | 1 | 0 | 2 | 0 | 0 | 0 | 0 | 0 | 33 | 0 | 14 |
| 53 | 32 | 20 | 8 | 0 | 8 | 0 | 69 | 5 | 1 | 0 | 186 | 19 | 0 | 45 | 0 | 0 | 0 | 0 | 0 | 0 | 0 | 0 | 4 | 0 | 0 | 44 | 0 | 21 |
| 56 | 20 | 0 | 0 | 0 | 0 | 0 | 1 | 0 | 0 | 0 | 3 | 2 | 0 | 0 | 0 | 0 | 0 | 0 | 0 | 0 | 0 | 0 | 0 | 0 | 0 | 20 | 0 | 1 |
| 57 | 21 | 0 | 0 | 0 | 0 | 0 | 1 | 0 | 0 | 0 | 9 | 4 | 0 | 0 | 0 | 0 | 0 | 0 | 0 | 0 | 0 | 0 | 0 | 0 | 0 | 15 | 0 | 0 |
| 60 | 16 | 3 | 0 | 1 | 0 | 0 | 14 | 0 | 0 | 0 | 17 | 3 | 2 | 1 | 0 | 0 | 2 | 0 | 0 | 1 | 0 | 0 | 1 | 0 | 0 | 14 | 0 | 6 |
| | # of unique subjects with at least one AE | Abdominal Pain | Aches | Anorexia | Chills | Convulsions | Cough | Diarrhea | Dizziness | Eye Irritation | Fever | Headache | Jaundice | Loss of appetite | Nausea / Vomiting | Respiratory distress | Runny Nose | Severe Malaria - Dark Urine | Severe Malaria - Other | Severe Malaria - Prostration / Very weak, unable to stand | Severe Malaria - Severe Pallor | Severe Malaria - Unconsciousness / Drowsiness | Skin Irritation / Rash | Tired | Unconsciousness / Drowsiness | Vomiting | Weak and unable to stand / move | Other |
| **min** | 1 | 0 | 0 | 0 | 0 | 0 | 0 | 0 | 0 | 0 | 0 | 0 | 0 | 0 | 0 | 0 | 0 | 0 | 0 | 0 | 0 | 0 | 0 | 0 | 0 | 0 | 0 | 0 |
| **max** | 32 | 20 | 8 | 2 | 8 | 1 | 69 | 8 | 1 | 1 | 186 | 41 | 2 | 45 | 1 | 1 | 2 | 1 | 10 | 4 | 2 | 2 | 4 | 2 | 2 | 44 | 2 | 23 |
| **median** | 17.5 | 3 | 0 | 0 | 0 | 0 | 5 | 0 | 0 | 0 | 17 | 5 | 0 | 1 | 0 | 0 | 0 | 0 | 0 | 0 | 0 | 0 | 0 | 0 | 0 | 8.5 | 0 | 2.5 |
| **mean** | 17.77 | 3.93 | 0.77 | 0.13 | 0.9 | 0.03 | 10.1 | 1.43 | 0.17 | 0.03 | 29.47 | 9.1 | 0.13 | 3.3 | 0.03 | 0.07 | 0.1 | 0.07 | 0.97 | 0.33 | 0.07 | 0.07 | 0.33 | 0.07 | 0.07 | 12.13 | 0.07 | 5.93 |
| **sd** | 8.73 | 4.43 | 1.68 | 0.43 | 1.84 | 0.18 | 13.91 | 2.19 | 0.38 | 0.18 | 37.57 | 10.37 | 0.43 | 8.26 | 0.18 | 0.25 | 0.4 | 0.25 | 2.24 | 0.88 | 0.37 | 0.37 | 0.84 | 0.37 | 0.37 | 11.5 | 0.37 | 7.27 |
| **total** | 533 | 118 | 23 | 4 | 27 | 1 | 303 | 43 | 5 | 1 | 884 | 273 | 4 | 99 | 1 | 2 | 3 | 2 | 29 | 10 | 2 | 2 | 10 | 2 | 2 | 364 | 2 | 178 |

### Table I3: number of AEs by symptom in the placebo clusters during the intervention

| Cluster | Total # of subjects | Abdominal Pain | Aches | Anorexia | Chills | Convulsions | Cough | Diarrhea | Dizziness | Eye Irritation | Fever | Headache | Loss of appetite | Runny Nose | Severe Malaria - Convulsions / Seizures | Severe Malaria - Dark Urine | Severe Malaria - Other | Severe Malaria - Severe Pallor | Severe Malaria - Unconsciousness / Drowsiness | Skin Irritation / Rash | Tired | Unconsciousness / Drowsiness | Vomiting | Other |
|---|---|---|---|---|---|---|---|---|---|---|---|---|---|---|---|---|---|---|---|---|---|---|---|---|
| 1 | 27 | 3 | 0 | 0 | 1 | 1 | 26 | 0 | 0 | 0 | 53 | 14 | 5 | 0 | 0 | 0 | 0 | 0 | 0 | 1 | 0 | 0 | 13 | 4 |
| 2 | 21 | 2 | 0 | 0 | 2 | 0 | 10 | 0 | 0 | 0 | 29 | 2 | 2 | 0 | 0 | 0 | 0 | 0 | 0 | 0 | 0 | 0 | 6 | 4 |
| 3 | 30 | 9 | 0 | 0 | 0 | 0 | 25 | 0 | 0 | 0 | 50 | 21 | 2 | 0 | 0 | 0 | 0 | 0 | 0 | 0 | 0 | 0 | 9 | 10 |
| 7 | 27 | 3 | 0 | 0 | 2 | 0 | 14 | 2 | 0 | 0 | 47 | 13 | 0 | 0 | 0 | 0 | 1 | 0 | 0 | 0 | 0 | 0 | 23 | 11 |
| 8 | 11 | 0 | 0 | 0 | 1 | 0 | 5 | 1 | 0 | 0 | 2 | 4 | 0 | 0 | 0 | 0 | 14 | 0 | 0 | 0 | 4 | 0 | 2 | 1 |
| 13 | 9 | 0 | 0 | 0 | 0 | 0 | 1 | 1 | 0 | 0 | 8 | 2 | 0 | 0 | 0 | 0 | 0 | 0 | 0 | 0 | 0 | 0 | 1 | 1 |
| 14 | 18 | 0 | 0 | 0 | 0 | 0 | 5 | 0 | 0 | 0 | 27 | 3 | 0 | 0 | 0 | 0 | 0 | 0 | 0 | 0 | 0 | 0 | 1 | 0 |
| 16 | 23 | 1 | 0 | 0 | 3 | 0 | 14 | 5 | 0 | 1 | 26 | 14 | 0 | 0 | 0 | 0 | 19 | 4 | 0 | 0 | 4 | 0 | 24 | 0 |
| 17 | 23 | 7 | 3 | 0 | 1 | 0 | 7 | 2 | 2 | 0 | 31 | 12 | 2 | 0 | 0 | 0 | 12 | 5 | 0 | 0 | 2 | 0 | 16 | 4 |
| 18 | 17 | 2 | 0 | 0 | 0 | 0 | 11 | 1 | 0 | 0 | 15 | 2 | 0 | 0 | 0 | 0 | 0 | 0 | 0 | 0 | 0 | 0 | 5 | 2 |
| 19 | 21 | 0 | 0 | 0 | 0 | 0 | 6 | 1 | 0 | 0 | 10 | 4 | 0 | 0 | 0 | 0 | 0 | 0 | 0 | 3 | 0 | 0 | 6 | 7 |
| 20 | 7 | 0 | 0 | 0 | 0 | 0 | 0 | 0 | 0 | 0 | 0 | 0 | 0 | 0 | 0 | 0 | 9 | 0 | 0 | 0 | 0 | 0 | 0 | 0 |
| 21 | 21 | 6 | 1 | 0 | 0 | 0 | 5 | 2 | 0 | 0 | 20 | 13 | 1 | 0 | 0 | 0 | 0 | 0 | 1 | 0 | 0 | 1 | 13 | 4 |
| 23 | 19 | 11 | 0 | 0 | 0 | 0 | 5 | 1 | 0 | 0 | 24 | 12 | 3 | 0 | 0 | 0 | 0 | 0 | 0 | 3 | 0 | 0 | 10 | 7 |
| 30 | 12 | 1 | 0 | 0 | 0 | 0 | 1 | 0 | 0 | 0 | 17 | 3 | 0 | 0 | 0 | 0 | 0 | 0 | 0 | 0 | 0 | 0 | 0 | 0 |
| 31 | 15 | 2 | 0 | 0 | 1 | 0 | 1 | 0 | 2 | 0 | 22 | 9 | 0 | 0 | 2 | 0 | 0 | 0 | 0 | 0 | 1 | 0 | 10 | 3 |
| 33 | 28 | 8 | 0 | 0 | 2 | 0 | 15 | 2 | 1 | 0 | 54 | 15 | 3 | 0 | 0 | 0 | 0 | 0 | 0 | 1 | 0 | 0 | 26 | 13 |
| 34 | 28 | 6 | 0 | 2 | 3 | 0 | 13 | 2 | 0 | 0 | 49 | 17 | 6 | 0 | 0 | 0 | 0 | 0 | 0 | 2 | 0 | 0 | 21 | 15 |
| 35 | 18 | 2 | 0 | 0 | 0 | 0 | 6 | 2 | 0 | 0 | 1 | 12 | 0 | 0 | 0 | 0 | 1 | 0 | 0 | 1 | 0 | 0 | 14 | 4 |
| 38 | 22 | 0 | 0 | 0 | 1 | 0 | 7 | 5 | 2 | 0 | 3 | 26 | 0 | 0 | 0 | 0 | 0 | 0 | 0 | 0 | 0 | 0 | 17 | 3 |
| 39 | 20 | 0 | 0 | 0 | 2 | 0 | 4 | 1 | 1 | 0 | 4 | 11 | 0 | 0 | 0 | 0 | 0 | 0 | 0 | 0 | 0 | 0 | 12 | 2 |
| 40 | 29 | 5 | 0 | 2 | 0 | 0 | 18 | 4 | 0 | 0 | 68 | 20 | 10 | 0 | 0 | 0 | 0 | 1 | 0 | 0 | 1 | 0 | 22 | 16 |
| 41 | 12 | 1 | 0 | 0 | 0 | 0 | 0 | 0 | 0 | 0 | 7 | 4 | 0 | 0 | 0 | 0 | 7 | 0 | 0 | 0 | 0 | 0 | 1 | 0 |
| 44 | 5 | 0 | 0 | 0 | 0 | 0 | 1 | 0 | 0 | 0 | 5 | 0 | 1 | 0 | 0 | 0 | 0 | 0 | 0 | 0 | 0 | 0 | 2 | 0 |
| 50 | 26 | 10 | 2 | 0 | 0 | 0 | 16 | 6 | 0 | 0 | 45 | 18 | 7 | 0 | 0 | 1 | 1 | 0 | 0 | 0 | 0 | 0 | 17 | 6 |
| 52 | 32 | 16 | 7 | 1 | 12 | 0 | 103 | 14 | 1 | 0 | 236 | 15 | 47 | 2 | 0 | 0 | 0 | 0 | 0 | 0 | 3 | 0 | 64 | 31 |
| 54 | 14 | 0 | 0 | 0 | 0 | 0 | 0 | 0 | 0 | 0 | 6 | 1 | 0 | 0 | 0 | 0 | 0 | 0 | 0 | 0 | 1 | 0 | 11 | 0 |
| 55 | 12 | 0 | 0 | 0 | 0 | 0 | 0 | 0 | 0 | 0 | 1 | 5 | 0 | 0 | 0 | 0 | 0 | 0 | 0 | 0 | 0 | 0 | 10 | 1 |
| 58 | 14 | 0 | 0 | 0 | 0 | 0 | 0 | 0 | 0 | 0 | 5 | 2 | 0 | 0 | 0 | 0 | 0 | 0 | 0 | 1 | 0 | 0 | 12 | 1 |
| 59 | 13 | 0 | 0 | 0 | 0 | 0 | 0 | 0 | 0 | 0 | 5 | 4 | 0 | 0 | 0 | 0 | 0 | 0 | 0 | 0 | 0 | 0 | 5 | 0 |
| | Total # of subjects | Abdominal Pain | Aches | Anorexia | Chills | Convulsions | Cough | Diarrhea | Dizziness | Eye Irritation | Fever | Headache | Loss of appetite | Runny Nose | Severe Malaria - Convulsions / Seizures | Severe Malaria - Dark Urine | Severe Malaria - Other | Severe Malaria - Severe Pallor | Severe Malaria - Unconsciousness / Drowsiness | Skin Irritation / Rash | Tired | Unconsciousness / Drowsiness | Vomiting | Other |
| **min** | 5 | 0 | 0 | 0 | 0 | 0 | 0 | 0 | 0 | 0 | 0 | 0 | 0 | 0 | 0 | 0 | 0 | 0 | 0 | 0 | 0 | 0 | 0 | 0 |
| **max** | 32 | 16 | 7 | 2 | 12 | 1 | 103 | 14 | 2 | 1 | 236 | 26 | 47 | 2 | 2 | 1 | 19 | 5 | 1 | 3 | 4 | 1 | 64 | 31 |
| **median** | 19.5 | 1.5 | 0 | 0 | 0 | 0 | 5.5 | 1 | 0 | 0 | 18.5 | 10 | 0 | 0 | 0 | 0 | 0 | 0 | 0 | 0 | 0 | 0 | 10.5 | 3 |
| **mean** | 19.13 | 3.17 | 0.43 | 0.17 | 1.03 | 0.03 | 10.63 | 1.73 | 0.3 | 0.03 | 29 | 9.27 | 2.97 | 0.07 | 0.07 | 0.03 | 2.13 | 0.33 | 0.03 | 0.4 | 0.53 | 0.03 | 12.43 | 5 |
| **sd** | 7.29 | 4.17 | 1.41 | 0.53 | 2.28 | 0.18 | 18.93 | 2.86 | 0.65 | 0.18 | 43.74 | 7.17 | 8.68 | 0.37 | 0.37 | 0.18 | 4.9 | 1.15 | 0.18 | 0.86 | 1.17 | 0.18 | 12.39 | 6.76 |
| **total** | 574 | 95 | 13 | 5 | 31 | 1 | 319 | 52 | 9 | 1 | 870 | 278 | 89 | 2 | 2 | 1 | 64 | 10 | 1 | 12 | 16 | 1 | 373 | 150 |

**Table I4: frequency and percentage of subjects having at least one AE episode by symptom in each cluster during the intervention (clusters highlighted in red are SR clusters)**

| Cluster | Total # of subjects | Abdominal Pain | | Aches | | Anorexia | | Chills | | Convulsions | | Cough | | Diarrhea | | Dizziness | | Eye Irritation | | Fever | | Headache | | Jaundice | | Loss of appetite | | Nausea / Vomiting | | Respiratory distress | | Runny Nose | | Severe Malaria - Convulsions / Seizures | | Severe Malaria - Dark Urine | | Severe Malaria - Other | | Severe Malaria - Prostration / Very weak, unable to stand | | Severe Malaria - Severe Pallor | | Severe Malaria - Unconsciousness / Drowsiness | | Skin Irritation / Rash | | Tired | | Unconsciousness / Drowsiness | | Vomiting | | Weak and unable to stand / move | | Other | |
|---|---|---|---|---|---|---|---|---|---|---|---|---|---|---|---|---|---|---|---|---|---|---|---|---|---|---|---|---|---|---|---|---|---|---|---|---|---|---|---|---|---|---|---|---|---|---|---|---|---|---|---|---|---|---|---|---|---|
| | | # | % | # | % | # | % | # | % | # | % | # | % | # | % | # | % | # | % | # | % | # | % | # | % | # | % | # | % | # | % | # | % | # | % | # | % | # | % | # | % | # | % | # | % | # | % | # | % | # | % | # | % | # | % | # | % |
| 1 | 31 | 3 | 9.7 | 0 | 0 | 0 | 0 | 1 | 3.2 | 1 | 3.2 | 16 | 51.6 | 0 | 0 | 0 | 0 | 0 | 0 | 24 | 77.4 | 12 | 38.7 | 0 | 0 | 5 | 16.1 | 0 | 0 | 0 | 0 | 0 | 0 | 0 | 0 | 0 | 0 | 0 | 0 | 0 | 0 | 0 | 0 | 0 | 0 | 1 | 3.2 | 0 | 0 | 0 | 0 | 9 | 29 | 0 | 0 | 2 | 6.5 |
| 2 | 32 | 1 | 3.1 | 0 | 0 | 0 | 0 | 2 | 6.2 | 0 | 0 | 9 | 28.1 | 0 | 0 | 0 | 0 | 0 | 0 | 17 | 53.1 | 2 | 6.2 | 0 | 0 | 2 | 6.2 | 0 | 0 | 0 | 0 | 0 | 0 | 0 | 0 | 0 | 0 | 0 | 0 | 0 | 0 | 0 | 0 | 0 | 0 | 0 | 0 | 0 | 0 | 0 | 0 | 6 | 18.8 | 0 | 0 | 3 | 9.4 |
| 3 | 32 | 8 | 25 | 0 | 0 | 0 | 0 | 0 | 0 | 0 | 0 | 14 | 43.8 | 0 | 0 | 0 | 0 | 0 | 0 | 27 | 84.4 | 13 | 40.6 | 0 | 0 | 2 | 6.2 | 0 | 0 | 0 | 0 | 0 | 0 | 0 | 0 | 0 | 0 | 0 | 0 | 0 | 0 | 0 | 0 | 0 | 0 | 0 | 0 | 0 | 0 | 0 | 0 | 8 | 25 | 0 | 0 | 10 | 31.2 |
| 4 | 32 | 5 | 15.6 | 0 | 0 | 0 | 0 | 0 | 0 | 0 | 0 | 14 | 43.8 | 0 | 0 | 0 | 0 | 0 | 0 | 17 | 53.1 | 3 | 9.4 | 0 | 0 | 4 | 12.5 | 0 | 0 | 0 | 0 | 0 | 0 | 0 | 0 | 0 | 0 | 0 | 0 | 0 | 0 | 0 | 0 | 0 | 0 | 0 | 0 | 0 | 0 | 0 | 0 | 6 | 18.8 | 0 | 0 | 4 | 12.5 |
| 5 | 32 | 6 | 18.8 | 0 | 0 | 0 | 0 | 1 | 3.1 | 0 | 0 | 9 | 28.1 | 0 | 0 | 0 | 0 | 0 | 0 | 23 | 71.9 | 10 | 31.2 | 0 | 0 | 0 | 0 | 0 | 0 | 0 | 0 | 0 | 0 | 0 | 0 | 0 | 0 | 0 | 0 | 0 | 0 | 0 | 0 | 0 | 0 | 0 | 0 | 0 | 0 | 0 | 0 | 11 | 34.4 | 0 | 0 | 9 | 28.1 |
| 6 | 32 | 4 | 12.5 | 0 | 0 | 0 | 0 | 4 | 12.5 | 0 | 0 | 15 | 46.9 | 1 | 3.1 | 0 | 0 | 0 | 0 | 28 | 87.5 | 16 | 50 | 0 | 0 | 1 | 3.1 | 0 | 0 | 0 | 0 | 0 | 0 | 0 | 0 | 0 | 0 | 3 | 9.4 | 0 | 0 | 0 | 0 | 0 | 0 | 0 | 0 | 0 | 0 | 0 | 0 | 18 | 56.2 | 0 | 0 | 13 | 40.6 |
| 7 | 32 | 3 | 9.4 | 0 | 0 | 0 | 0 | 2 | 6.2 | 0 | 0 | 11 | 34.4 | 2 | 6.2 | 0 | 0 | 0 | 0 | 21 | 65.6 | 10 | 31.2 | 0 | 0 | 0 | 0 | 0 | 0 | 0 | 0 | 0 | 0 | 0 | 0 | 0 | 0 | 1 | 3.1 | 0 | 0 | 0 | 0 | 0 | 0 | 0 | 0 | 0 | 0 | 0 | 0 | 15 | 46.9 | 0 | 0 | 11 | 34.4 |
| 8 | 32 | 0 | 0 | 0 | 0 | 0 | 0 | 1 | 3.1 | 0 | 0 | 4 | 12.5 | 1 | 3.1 | 0 | 0 | 0 | 0 | 2 | 6.2 | 3 | 9.4 | 0 | 0 | 0 | 0 | 0 | 0 | 0 | 0 | 0 | 0 | 0 | 0 | 0 | 0 | 9 | 28.1 | 0 | 0 | 0 | 0 | 0 | 0 | 0 | 0 | 3 | 9.4 | 0 | 0 | 2 | 6.2 | 0 | 0 | 1 | 3.1 |
| 9 | 30 | 0 | 0 | 0 | 0 | 0 | 0 | 0 | 0 | 0 | 0 | 0 | 0 | 0 | 0 | 0 | 0 | 0 | 0 | 0 | 0 | 0 | 0 | 0 | 0 | 0 | 0 | 0 | 0 | 0 | 0 | 0 | 0 | 0 | 0 | 0 | 0 | 1 | 3.3 | 0 | 0 | 0 | 0 | 0 | 0 | 0 | 0 | 0 | 0 | 0 | 0 | 0 | 0 | 0 | 0 | 0 | 0 |
| 10 | 32 | 1 | 3.1 | 0 | 0 | 0 | 0 | 0 | 0 | 0 | 0 | 1 | 3.1 | 0 | 0 | 0 | 0 | 0 | 0 | 0 | 0 | 2 | 6.2 | 0 | 0 | 0 | 0 | 0 | 0 | 0 | 0 | 0 | 0 | 0 | 0 | 0 | 0 | 2 | 6.2 | 0 | 0 | 0 | 0 | 0 | 0 | 0 | 0 | 0 | 0 | 0 | 0 | 0 | 0 | 0 | 0 | 0 | 0 |
| 11 | 32 | 0 | 0 | 0 | 0 | 0 | 0 | 0 | 0 | 0 | 0 | 0 | 0 | 0 | 0 | 0 | 0 | 0 | 0 | 0 | 0 | 0 | 0 | 0 | 0 | 0 | 0 | 0 | 0 | 0 | 0 | 0 | 0 | 0 | 0 | 0 | 0 | 4 | 12.5 | 0 | 0 | 0 | 0 | 0 | 0 | 0 | 0 | 0 | 0 | 0 | 0 | 0 | 0 | 0 | 0 | 0 | 0 |
| 12 | 32 | 2 | 6.2 | 0 | 0 | 0 | 0 | 0 | 0 | 0 | 0 | 3 | 9.4 | 1 | 3.1 | 0 | 0 | 0 | 0 | 14 | 43.8 | 1 | 3.1 | 0 | 0 | 0 | 0 | 0 | 0 | 0 | 0 | 0 | 0 | 0 | 0 | 0 | 0 | 0 | 0 | 0 | 0 | 0 | 0 | 0 | 0 | 0 | 0 | 0 | 0 | 0 | 0 | 0 | 0 | 0 | 0 | 0 | 0 |
| 13 | 31 | 0 | 0 | 0 | 0 | 0 | 0 | 0 | 0 | 0 | 0 | 1 | 3.2 | 1 | 3.2 | 0 | 0 | 0 | 0 | 8 | 25.8 | 2 | 6.5 | 0 | 0 | 0 | 0 | 0 | 0 | 0 | 0 | 0 | 0 | 0 | 0 | 0 | 0 | 0 | 0 | 0 | 0 | 0 | 0 | 0 | 0 | 0 | 0 | 0 | 0 | 0 | 0 | 1 | 3.2 | 0 | 0 | 1 | 3.2 |
| 14 | 31 | 0 | 0 | 0 | 0 | 0 | 0 | 0 | 0 | 0 | 0 | 5 | 16.1 | 0 | 0 | 0 | 0 | 0 | 0 | 17 | 54.8 | 3 | 9.7 | 0 | 0 | 0 | 0 | 0 | 0 | 0 | 0 | 0 | 0 | 0 | 0 | 0 | 0 | 0 | 0 | 0 | 0 | 0 | 0 | 0 | 0 | 0 | 0 | 0 | 0 | 0 | 0 | 1 | 3.2 | 0 | 0 | 0 | 0 |
| 15 | 32 | 0 | 0 | 0 | 0 | 0 | 0 | 0 | 0 | 0 | 0 | 2 | 6.2 | 2 | 6.2 | 0 | 0 | 0 | 0 | 7 | 21.9 | 0 | 0 | 0 | 0 | 1 | 3.1 | 0 | 0 | 0 | 0 | 0 | 0 | 0 | 0 | 0 | 0 | 0 | 0 | 0 | 0 | 0 | 0 | 0 | 0 | 0 | 0 | 0 | 0 | 0 | 0 | 3 | 9.4 | 0 | 0 | 2 | 6.2 |
| 16 | 31 | 1 | 3.2 | 0 | 0 | 0 | 0 | 2 | 6.5 | 0 | 0 | 8 | 25.8 | 3 | 9.7 | 0 | 0 | 1 | 3.2 | 16 | 51.6 | 9 | 29 | 0 | 0 | 0 | 0 | 0 | 0 | 0 | 0 | 0 | 0 | 0 | 0 | 0 | 0 | 10 | 32.3 | 0 | 0 | 4 | 12.9 | 0 | 0 | 0 | 0 | 3 | 9.7 | 0 | 0 | 12 | 38.7 | 0 | 0 | 0 | 0 |
| 17 | 31 | 6 | 19.4 | 3 | 9.7 | 0 | 0 | 1 | 3.2 | 0 | 0 | 4 | 12.9 | 2 | 6.5 | 1 | 3.2 | 0 | 0 | 18 | 58.1 | 8 | 25.8 | 0 | 0 | 2 | 6.5 | 0 | 0 | 0 | 0 | 0 | 0 | 0 | 0 | 0 | 0 | 8 | 25.8 | 0 | 0 | 3 | 9.7 | 0 | 0 | 0 | 0 | 1 | 3.2 | 0 | 0 | 11 | 35.5 | 0 | 0 | 4 | 12.9 |
| 18 | 32 | 1 | 3.1 | 0 | 0 | 0 | 0 | 0 | 0 | 0 | 0 | 7 | 21.9 | 1 | 3.1 | 0 | 0 | 0 | 0 | 12 | 37.5 | 2 | 6.2 | 0 | 0 | 0 | 0 | 0 | 0 | 0 | 0 | 0 | 0 | 0 | 0 | 0 | 0 | 0 | 0 | 0 | 0 | 0 | 0 | 0 | 0 | 0 | 0 | 0 | 0 | 0 | 0 | 5 | 15.6 | 0 | 0 | 2 | 6.2 |
| 19 | 32 | 0 | 0 | 0 | 0 | 0 | 0 | 0 | 0 | 0 | 0 | 6 | 18.8 | 1 | 3.1 | 0 | 0 | 0 | 0 | 10 | 31.2 | 3 | 9.4 | 0 | 0 | 0 | 0 | 0 | 0 | 0 | 0 | 0 | 0 | 0 | 0 | 0 | 0 | 0 | 0 | 0 | 0 | 0 | 0 | 0 | 0 | 3 | 9.4 | 0 | 0 | 0 | 0 | 6 | 18.8 | 0 | 0 | 6 | 18.8 |
| 20 | 32 | 0 | 0 | 0 | 0 | 0 | 0 | 0 | 0 | 0 | 0 | 0 | 0 | 0 | 0 | 0 | 0 | 0 | 0 | 0 | 0 | 0 | 0 | 0 | 0 | 0 | 0 | 0 | 0 | 0 | 0 | 0 | 0 | 0 | 0 | 0 | 0 | 7 | 21.9 | 0 | 0 | 0 | 0 | 0 | 0 | 0 | 0 | 0 | 0 | 0 | 0 | 0 | 0 | 0 | 0 | 0 | 0 |
| 21 | 32 | 4 | 12.5 | 1 | 3.1 | 0 | 0 | 0 | 0 | 0 | 0 | 4 | 12.5 | 2 | 6.2 | 0 | 0 | 0 | 0 | 16 | 50 | 9 | 28.1 | 0 | 0 | 1 | 3.1 | 0 | 0 | 0 | 0 | 0 | 0 | 0 | 0 | 0 | 0 | 0 | 0 | 0 | 0 | 0 | 0 | 1 | 3.1 | 0 | 0 | 0 | 0 | 1 | 3.1 | 10 | 31.2 | 0 | 0 | 4 | 12.5 |
| 22 | 31 | 3 | 9.7 | 1 | 3.2 | 0 | 0 | 1 | 3.2 | 0 | 0 | 5 | 16.1 | 1 | 3.2 | 0 | 0 | 0 | 0 | 10 | 32.3 | 9 | 29 | 0 | 0 | 2 | 6.5 | 0 | 0 | 0 | 0 | 0 | 0 | 0 | 0 | 0 | 0 | 1 | 3.2 | 0 | 0 | 0 | 0 | 0 | 0 | 0 | 0 | 0 | 0 | 0 | 0 | 7 | 22.6 | 0 | 0 | 0 | 0 |
| 23 | 31 | 8 | 25.8 | 0 | 0 | 0 | 0 | 0 | 0 | 0 | 0 | 5 | 16.1 | 1 | 3.2 | 0 | 0 | 0 | 0 | 12 | 38.7 | 9 | 29 | 0 | 0 | 3 | 9.7 | 0 | 0 | 0 | 0 | 0 | 0 | 0 | 0 | 0 | 0 | 0 | 0 | 0 | 0 | 0 | 0 | 0 | 0 | 3 | 9.7 | 0 | 0 | 0 | 0 | 8 | 25.8 | 0 | 0 | 6 | 19.4 |
| 24 | 28 | 3 | 10.7 | 1 | 3.6 | 0 | 0 | 0 | 0 | 0 | 0 | 4 | 14.3 | 0 | 0 | 0 | 0 | 0 | 0 | 10 | 35.7 | 3 | 10.7 | 1 | 3.6 | 0 | 0 | 0 | 0 | 0 | 0 | 0 | 0 | 0 | 0 | 0 | 0 | 0 | 0 | 0 | 0 | 0 | 0 | 0 | 0 | 1 | 3.6 | 0 | 0 | 0 | 0 | 5 | 17.9 | 0 | 0 | 2 | 7.1 |
| 25 | 32 | 3 | 9.4 | 0 | 0 | 0 | 0 | 0 | 0 | 0 | 0 | 5 | 15.6 | 0 | 0 | 0 | 0 | 0 | 0 | 14 | 43.8 | 2 | 6.2 | 0 | 0 | 1 | 3.1 | 0 | 0 | 0 | 0 | 0 | 0 | 0 | 0 | 0 | 0 | 0 | 0 | 0 | 0 | 0 | 0 | 0 | 0 | 1 | 3.1 | 0 | 0 | 0 | 0 | 5 | 15.6 | 0 | 0 | 2 | 6.2 |
| 26 | 32 | 3 | 9.4 | 1 | 3.1 | 1 | 3.1 | 0 | 0 | 0 | 0 | 4 | 12.5 | 2 | 6.2 | 1 | 3.1 | 0 | 0 | 11 | 34.4 | 6 | 18.8 | 0 | 0 | 4 | 12.5 | 0 | 0 | 0 | 0 | 0 | 0 | 0 | 0 | 0 | 0 | 0 | 0 | 0 | 0 | 0 | 0 | 0 | 0 | 0 | 0 | 0 | 0 | 0 | 0 | 3 | 9.4 | 0 | 0 | 1 | 3.1 |
| 27 | 32 | 2 | 6.2 | 0 | 0 | 0 | 0 | 0 | 0 | 0 | 0 | 6 | 18.8 | 0 | 0 | 0 | 0 | 1 | 3.1 | 16 | 50 | 6 | 18.8 | 0 | 0 | 1 | 3.1 | 0 | 0 | 0 | 0 | 0 | 0 | 0 | 0 | 0 | 0 | 0 | 0 | 2 | 6.2 | 1 | 3.1 | 1 | 3.1 | 0 | 0 | 0 | 0 | 1 | 3.1 | 4 | 12.5 | 1 | 3.1 | 5 | 15.6 |
| 28 | 32 | 4 | 12.5 | 0 | 0 | 0 | 0 | 0 | 0 | 1 | 3.1 | 5 | 15.6 | 0 | 0 | 0 | 0 | 0 | 0 | 18 | 56.2 | 7 | 21.9 | 0 | 0 | 0 | 0 | 0 | 0 | 0 | 0 | 0 | 0 | 0 | 0 | 0 | 0 | 0 | 0 | 0 | 0 | 0 | 0 | 0 | 0 | 0 | 0 | 0 | 0 | 0 | 0 | 9 | 28.1 | 0 | 0 | 5 | 15.6 |
| 29 | 31 | 1 | 3.2 | 1 | 3.2 | 0 | 0 | 0 | 0 | 0 | 0 | 2 | 6.5 | 0 | 0 | 1 | 3.2 | 0 | 0 | 11 | 35.5 | 5 | 16.1 | 0 | 0 | 4 | 12.9 | 0 | 0 | 0 | 0 | 0 | 0 | 0 | 0 | 0 | 0 | 0 | 0 | 0 | 0 | 0 | 0 | 0 | 0 | 0 | 0 | 0 | 0 | 0 | 0 | 9 | 29 | 0 | 0 | 1 | 3.2 |
| 30 | 32 | 1 | 3.1 | 0 | 0 | 0 | 0 | 0 | 0 | 0 | 0 | 1 | 3.1 | 0 | 0 | 0 | 0 | 0 | 0 | 11 | 34.4 | 3 | 9.4 | 0 | 0 | 0 | 0 | 0 | 0 | 0 | 0 | 0 | 0 | 0 | 0 | 0 | 0 | 0 | 0 | 0 | 0 | 0 | 0 | 0 | 0 | 0 | 0 | 0 | 0 | 0 | 0 | 0 | 0 | 0 | 0 | 0 | 0 |
| 31 | 25 | 2 | 8 | 0 | 0 | 0 | 0 | 1 | 4 | 0 | 0 | 1 | 4 | 0 | 0 | 2 | 8 | 0 | 0 | 11 | 44 | 7 | 28 | 0 | 0 | 0 | 0 | 0 | 0 | 0 | 0 | 0 | 0 | 1 | 4 | 0 | 0 | 0 | 0 | 0 | 0 | 0 | 0 | 0 | 0 | 0 | 0 | 1 | 4 | 0 | 0 | 8 | 32 | 0 | 0 | 3 | 12 |
| 32 | 32 | 5 | 15.6 | 0 | 0 | 2 | 6.2 | 0 | 0 | 0 | 0 | 17 | 53.1 | 5 | 15.6 | 0 | 0 | 0 | 0 | 24 | 75 | 5 | 15.6 | 0 | 0 | 7 | 21.9 | 0 | 0 | 0 | 0 | 0 | 0 | 0 | 0 | 0 | 0 | 0 | 0 | 0 | 0 | 0 | 0 | 0 | 0 | 0 | 0 | 0 | 0 | 0 | 0 | 14 | 43.8 | 0 | 0 | 11 | 34.4 |
| 33 | 32 | 8 | 25 | 0 | 0 | 0 | 0 | 2 | 6.2 | 0 | 0 | 11 | 34.4 | 1 | 3.1 | 1 | 3.1 | 0 | 0 | 26 | 81.2 | 11 | 34.4 | 0 | 0 | 3 | 9.4 | 0 | 0 | 0 | 0 | 0 | 0 | 0 | 0 | 0 | 0 | 0 | 0 | 0 | 0 | 0 | 0 | 0 | 0 | 1 | 3.1 | 0 | 0 | 0 | 0 | 20 | 62.5 | 0 | 0 | 8 | 25 |
| 34 | 32 | 6 | 18.8 | 0 | 0 | 2 | 6.2 | 3 | 9.4 | 0 | 0 | 9 | 28.1 | 2 | 6.2 | 0 | 0 | 0 | 0 | 25 | 78.1 | 14 | 43.8 | 0 | 0 | 5 | 15.6 | 0 | 0 | 0 | 0 | 0 | 0 | 0 | 0 | 0 | 0 | 0 | 0 | 0 | 0 | 0 | 0 | 0 | 0 | 2 | 6.2 | 0 | 0 | 0 | 0 | 15 | 46.9 | 0 | 0 | 10 | 31.2 |
| 35 | 32 | 2 | 6.2 | 0 | 0 | 0 | 0 | 0 | 0 | 0 | 0 | 5 | 15.6 | 2 | 6.2 | 0 | 0 | 0 | 0 | 1 | 3.1 | 8 | 25 | 0 | 0 | 0 | 0 | 0 | 0 | 0 | 0 | 0 | 0 | 0 | 0 | 0 | 0 | 1 | 3.1 | 0 | 0 | 0 | 0 | 0 | 0 | 1 | 3.1 | 0 | 0 | 0 | 0 | 7 | 21.9 | 0 | 0 | 4 | 12.5 |
| 36 | 32 | 5 | 15.6 | 3 | 9.4 | 0 | 0 | 0 | 0 | 0 | 0 | 11 | 34.4 | 2 | 6.2 | 0 | 0 | 0 | 0 | 22 | 68.8 | 14 | 43.8 | 0 | 0 | 5 | 15.6 | 0 | 0 | 0 | 0 | 1 | 3.1 | 0 | 0 | 1 | 3.1 | 0 | 0 | 0 | 0 | 0 | 0 | 0 | 0 | 2 | 6.2 | 0 | 0 | 0 | 0 | 15 | 46.9 | 0 | 0 | 11 | 34.4 |
| 37 | 31 | 0 | 0 | 0 | 0 | 0 | 0 | 1 | 3.2 | 0 | 0 | 5 | 16.1 | 1 | 3.2 | 0 | 0 | 0 | 0 | 1 | 3.2 | 6 | 19.4 | 0 | 0 | 0 | 0 | 0 | 0 | 0 | 0 | 0 | 0 | 0 | 0 | 0 | 0 | 0 | 0 | 0 | 0 | 0 | 0 | 0 | 0 | 0 | 0 | 0 | 0 | 0 | 0 | 8 | 25.8 | 0 | 0 | 3 | 9.7 |
| 38 | 32 | 0 | 0 | 0 | 0 | 0 | 0 | 1 | 3.1 | 0 | 0 | 6 | 18.8 | 3 | 9.4 | 2 | 6.2 | 0 | 0 | 3 | 9.4 | 18 | 56.2 | 0 | 0 | 0 | 0 | 0 | 0 | 0 | 0 | 0 | 0 | 0 | 0 | 0 | 0 | 0 | 0 | 0 | 0 | 0 | 0 | 0 | 0 | 0 | 0 | 0 | 0 | 0 | 0 | 13 | 40.6 | 0 | 0 | 3 | 9.4 |
| 39 | 32 | 0 | 0 | 0 | 0 | 0 | 0 | 2 | 6.2 | 0 | 0 | 3 | 9.4 | 1 | 3.1 | 1 | 3.1 | 0 | 0 | 4 | 12.5 | 10 | 31.2 | 0 | 0 | 0 | 0 | 0 | 0 | 0 | 0 | 0 | 0 | 0 | 0 | 0 | 0 | 0 | 0 | 0 | 0 | 0 | 0 | 0 | 0 | 0 | 0 | 0 | 0 | 0 | 0 | 11 | 34.4 | 0 | 0 | 1 | 3.1 |
| 40 | 32 | 5 | 15.6 | 0 | 0 | 2 | 6.2 | 0 | 0 | 0 | 0 | 14 | 43.8 | 3 | 9.4 | 0 | 0 | 0 | 0 | 26 | 81.2 | 18 | 56.2 | 0 | 0 | 8 | 25 | 0 | 0 | 0 | 0 | 0 | 0 | 0 | 0 | 0 | 0 | 0 | 0 | 0 | 0 | 1 | 3.1 | 0 | 0 | 0 | 0 | 1 | 3.1 | 0 | 0 | 17 | 53.1 | 0 | 0 | 12 | 37.5 |
| 41 | 32 | 1 | 3.1 | 0 | 0 | 0 | 0 | 0 | 0 | 0 | 0 | 0 | 0 | 0 | 0 | 0 | 0 | 0 | 0 | 6 | 18.8 | 4 | 12.5 | 0 | 0 | 0 | 0 | 0 | 0 | 0 | 0 | 0 | 0 | 0 | 0 | 0 | 0 | 6 | 18.8 | 0 | 0 | 0 | 0 | 0 | 0 | 0 | 0 | 0 | 0 | 0 | 0 | 1 | 3.1 | 0 | 0 | 0 | 0 |
| 42 | 32 | 0 | 0 | 0 | 0 | 0 | 0 | 0 | 0 | 0 | 0 | 1 | 3.1 | 0 | 0 | 0 | 0 | 0 | 0 | 6 | 18.8 | 2 | 6.2 | 0 | 0 | 0 | 0 | 0 | 0 | 0 | 0 | 0 | 0 | 0 | 0 | 0 | 0 | 4 | 12.5 | 0 | 0 | 0 | 0 | 0 | 0 | 0 | 0 | 0 | 0 | 0 | 0 | 0 | 0 | 0 | 0 | 1 | 3.1 |
| 43 | 29 | 0 | 0 | 0 | 0 | 0 | 0 | 0 | 0 | 0 | 0 | 0 | 0 | 0 | 0 | 0 | 0 | 0 | 0 | 5 | 17.2 | 2 | 6.9 | 0 | 0 | 0 | 0 | 0 | 0 | 0 | 0 | 0 | 0 | 0 | 0 | 0 | 0 | 8 | 27.6 | 0 | 0 | 0 | 0 | 0 | 0 | 0 | 0 | 0 | 0 | 0 | 0 | 1 | 3.4 | 0 | 0 | 0 | 0 |
| 44 | 31 | 0 | 0 | 0 | 0 | 0 | 0 | 0 | 0 | 0 | 0 | 1 | 3.2 | 0 | 0 | 0 | 0 | 0 | 0 | 5 | 16.1 | 0 | 0 | 0 | 0 | 1 | 3.2 | 0 | 0 | 0 | 0 | 0 | 0 | 0 | 0 | 0 | 0 | 0 | 0 | 0 | 0 | 0 | 0 | 0 | 0 | 0 | 0 | 0 | 0 | 0 | 0 | 2 | 6.5 | 0 | 0 | 0 | 0 |
| 45 | 32 | 1 | 3.1 | 0 | 0 | 0 | 0 | 0 | 0 | 0 | 0 | 0 | 0 | 0 | 0 | 0 | 0 | 0 | 0 | 5 | 15.6 | 3 | 9.4 | 0 | 0 | 0 | 0 | 0 | 0 | 0 | 0 | 0 | 0 | 0 | 0 | 0 | 0 | 0 | 0 | 0 | 0 | 0 | 0 | 0 | 0 | 0 | 0 | 0 | 0 | 0 | 0 | 2 | 6.2 | 0 | 0 | 1 | 3.1 |
| 46 | 32 | 9 | 28.1 | 2 | 6.2 | 0 | 0 | 1 | 3.1 | 0 | 0 | 11 | 34.4 | 4 | 12.5 | 1 | 3.1 | 0 | 0 | 28 | 87.5 | 22 | 68.8 | 0 | 0 | 5 | 15.6 | 0 | 0 | 0 | 0 | 0 | 0 | 0 | 0 | 0 | 0 | 1 | 3.1 | 1 | 3.1 | 0 | 0 | 0 | 0 | 0 | 0 | 0 | 0 | 0 | 0 | 14 | 43.8 | 0 | 0 | 18 | 56.2 |
| 47 | 32 | 3 | 9.4 | 0 | 0 | 0 | 0 | 0 | 0 | 0 | 0 | 4 | 12.5 | 2 | 6.2 | 0 | 0 | 0 | 0 | 6 | 18.8 | 4 | 12.5 | 0 | 0 | 1 | 3.1 | 0 | 0 | 1 | 3.1 | 0 | 0 | 0 | 0 | 0 | 0 | 0 | 0 | 0 | 0 | 0 | 0 | 0 | 0 | 0 | 0 | 0 | 0 | 0 | 0 | 6 | 18.8 | 0 | 0 | 2 | 6.2 |
| 48 | 32 | 7 | 21.9 | 3 | 9.4 | 0 | 0 | 3 | 9.4 | 0 | 0 | 11 | 34.4 | 2 | 6.2 | 0 | 0 | 0 | 0 | 28 | 87.5 | 13 | 40.6 | 0 | 0 | 5 | 15.6 | 0 | 0 | 0 | 0 | 0 | 0 | 0 | 0 | 0 | 0 | 0 | 0 | 1 | 3.1 | 0 | 0 | 0 | 0 | 1 | 3.1 | 0 | 0 | 0 | 0 | 19 | 59.4 | 0 | 0 | 3 | 9.4 |
| 49 | 32 | 1 | 3.1 | 0 | 0 | 0 | 0 | 2 | 6.2 | 0 | 0 | 7 | 21.9 | 7 | 21.9 | 0 | 0 | 0 | 0 | 11 | 34.4 | 6 | 18.8 | 0 | 0 | 1 | 3.1 | 0 | 0 | 1 | 3.1 | 0 | 0 | 0 | 0 | 0 | 0 | 0 | 0 | 0 | 0 | 0 | 0 | 0 | 0 | 0 | 0 | 1 | 3.1 | 0 | 0 | 10 | 31.2 | 0 | 0 | 8 | 25 |
| 50 | 31 | 8 | 25.8 | 2 | 6.5 | 0 | 0 | 0 | 0 | 0 | 0 | 12 | 38.7 | 3 | 9.7 | 0 | 0 | 0 | 0 | 25 | 80.6 | 14 | 45.2 | 0 | 0 | 6 | 19.4 | 0 | 0 | 0 | 0 | 0 | 0 | 0 | 0 | 1 | 3.2 | 1 | 3.2 | 0 | 0 | 0 | 0 | 0 | 0 | 0 | 0 | 0 | 0 | 0 | 0 | 15 | 48.4 | 0 | 0 | 6 | 19.4 |
| 51 | 32 | 7 | 21.9 | 3 | 9.4 | 0 | 0 | 4 | 12.5 | 0 | 0 | 12 | 37.5 | 2 | 6.2 | 1 | 3.1 | 0 | 0 | 24 | 75 | 21 | 65.6 | 1 | 3.1 | 6 | 18.8 | 1 | 3.1 | 0 | 0 | 0 | 0 | 0 | 0 | 1 | 3.1 | 0 | 0 | 1 | 3.1 | 0 | 0 | 0 | 0 | 0 | 0 | 0 | 0 | 0 | 0 | 15 | 46.9 | 0 | 0 | 9 | 28.1 |
| 52 | 32 | 9 | 28.1 | 6 | 18.8 | 1 | 3.1 | 8 | 25 | 0 | 0 | 30 | 93.8 | 11 | 34.4 | 1 | 3.1 | 0 | 0 | 32 | 100 | 8 | 25 | 0 | 0 | 25 | 78.1 | 0 | 0 | 0 | 0 | 2 | 6.2 | 0 | 0 | 0 | 0 | 0 | 0 | 0 | 0 | 0 | 0 | 0 | 0 | 0 | 0 | 2 | 6.2 | 0 | 0 | 28 | 87.5 | 0 | 0 | 19 | 59.4 |
| 53 | 32 | 11 | 34.4 | 7 | 21.9 | 0 | 0 | 6 | 18.8 | 0 | 0 | 26 | 81.2 | 4 | 12.5 | 1 | 3.1 | 0 | 0 | 32 | 100 | 14 | 43.8 | 0 | 0 | 26 | 81.2 | 0 | 0 | 0 | 0 | 0 | 0 | 0 | 0 | 0 | 0 | 0 | 0 | 0 | 0 | 0 | 0 | 0 | 0 | 4 | 12.5 | 0 | 0 | 0 | 0 | 23 | 71.9 | 0 | 0 | 14 | 43.8 |
| 54 | 32 | 0 | 0 | 0 | 0 | 0 | 0 | 0 | 0 | 0 | 0 | 0 | 0 | 0 | 0 | 0 | 0 | 0 | 0 | 5 | 15.6 | 1 | 3.1 | 0 | 0 | 0 | 0 | 0 | 0 | 0 | 0 | 0 | 0 | 0 | 0 | 0 | 0 | 0 | 0 | 0 | 0 | 0 | 0 | 0 | 0 | 0 | 0 | 1 | 3.1 | 0 | 0 | 9 | 28.1 | 0 | 0 | 0 | 0 |
| 55 | 31 | 0 | 0 | 0 | 0 | 0 | 0 | 0 | 0 | 0 | 0 | 0 | 0 | 0 | 0 | 0 | 0 | 0 | 0 | 1 | 3.2 | 5 | 16.1 | 0 | 0 | 0 | 0 | 0 | 0 | 0 | 0 | 0 | 0 | 0 | 0 | 0 | 0 | 0 | 0 | 0 | 0 | 0 | 0 | 0 | 0 | 0 | 0 | 0 | 0 | 0 | 0 | 7 | 22.6 | 0 | 0 | 1 | 3.2 |
| 56 | 32 | 0 | 0 | 0 | 0 | 0 | 0 | 0 | 0 | 0 | 0 | 1 | 3.1 | 0 | 0 | 0 | 0 | 0 | 0 | 3 | 9.4 | 2 | 6.2 | 0 | 0 | 0 | 0 | 0 | 0 | 0 | 0 | 0 | 0 | 0 | 0 | 0 | 0 | 0 | 0 | 0 | 0 | 0 | 0 | 0 | 0 | 0 | 0 | 0 | 0 | 0 | 0 | 15 | 46.9 | 0 | 0 | 1 | 3.1 |
| 57 | 32 | 0 | 0 | 0 | 0 | 0 | 0 | 0 | 0 | 0 | 0 | 1 | 3.1 | 0 | 0 | 0 | 0 | 0 | 0 | 8 | 25 | 4 | 12.5 | 0 | 0 | 0 | 0 | 0 | 0 | 0 | 0 | 0 | 0 | 0 | 0 | 0 | 0 | 0 | 0 | 0 | 0 | 0 | 0 | 0 | 0 | 0 | 0 | 0 | 0 | 0 | 0 | 13 | 40.6 | 0 | 0 | 0 | 0 |
| 58 | 32 | 0 | 0 | 0 | 0 | 0 | 0 | 0 | 0 | 0 | 0 | 0 | 0 | 0 | 0 | 0 | 0 | 0 | 0 | 5 | 15.6 | 2 | 6.2 | 0 | 0 | 0 | 0 | 0 | 0 | 0 | 0 | 0 | 0 | 0 | 0 | 0 | 0 | 0 | 0 | 0 | 0 | 0 | 0 | 0 | 0 | 1 | 3.1 | 0 | 0 | 0 | 0 | 11 | 34.4 | 0 | 0 | 1 | 3.1 |
| 59 | 32 | 0 | 0 | 0 | 0 | 0 | 0 | 0 | 0 | 0 | 0 | 0 | 0 | 0 | 0 | 0 | 0 | 0 | 0 | 5 | 15.6 | 3 | 9.4 | 0 | 0 | 0 | 0 | 0 | 0 | 0 | 0 | 0 | 0 | 0 | 0 | 0 | 0 | 0 | 0 | 0 | 0 | 0 | 0 | 0 | 0 | 0 | 0 | 0 | 0 | 0 | 0 | 5 | 15.6 | 0 | 0 | 0 | 0 |
| 60 | 32 | 2 | 6.2 | 0 | 0 | 1 | 3.1 | 0 | 0 | 0 | 0 | 13 | 40.6 | 0 | 0 | 0 | 0 | 0 | 0 | 12 | 37.5 | 3 | 9.4 | 1 | 3.1 | 1 | 3.1 | 0 | 0 | 0 | 0 | 1 | 3.1 | 0 | 0 | 0 | 0 | 0 | 0 | 1 | 3.1 | 0 | 0 | 0 | 0 | 1 | 3.1 | 0 | 0 | 0 | 0 | 11 | 34.4 | 0 | 0 | 4 | 12.5 |
| | Total # of subjects | Abdominal Pain | | Aches | | Anorexia | | Chills | | Convulsions | | Cough | | Diarrhea | | Dizziness | | Eye Irritation | | Fever | | Headache | | Jaundice | | Loss of appetite | | Nausea / Vomiting | | Respiratory distress | | Runny Nose | | Severe Malaria - Convulsions / Seizures | | Severe Malaria - Dark Urine | | Severe Malaria - Other | | severe malaria Prostration / very weak, unable to stand | | Severe Malaria - Severe Pallor | | Severe Malaria - Unconsciousness / Drowsiness | | Skin Irritation / Rash | | Tired | | Unconsciousness / Drowsiness | | Vomiting | | Weak and unable to stand / move | | Other | |
| | | # | % | # | % | # | % | # | % | # | % | # | % | # | % | # | % | # | % | # | % | # | % | # | % | # | % | # | % | # | % | # | % | # | % | # | % | # | % | # | % | # | % | # | % | # | % | # | % | # | % | # | % | # | % | # | % |
| min | 25 | 0 | 0 | 0 | 0 | 0 | 0 | 0 | 0 | 0 | 0 | 0 | 0 | 0 | 0 | 0 | 0 | 0 | 0 | 0 | 0 | 0 | 0 | 0 | 0 | 0 | 0 | 0 | 0 | 0 | 0 | 0 | 0 | 0 | 0 | 0 | 0 | 0 | 0 | 0 | 0 | 0 | 0 | 0 | 0 | 0 | 0 | 0 | 0 | 0 | 0 | 0 | 0 | 0 | 0 | 0 | 0 |
| max | 32 | 11 | 34.4 | 7 | 21.9 | 2 | 6.2 | 8 | 25 | 1 | 3.2 | 30 | 93.8 | 11 | 34.4 | 2 | 8 | 1 | 3.2 | 32 | 100 | 22 | 68.8 | 1 | 3.6 | 26 | 81.2 | 1 | 3.1 | 1 | 3.1 | 2 | 6.2 | 1 | 4 | 1 | 3.2 | 10 | 32.3 | 2 | 6.2 | 4 | 12.9 | 1 | 3.1 | 4 | 12.5 | 3 | 9.7 | 1 | 3.1 | 28 | 87.5 | 1 | 3.1 | 19 | 59.4 |
| median | 32 | 2 | 6.2 | 0 | 0 | 0 | 0 | 0 | 0 | 0 | 0 | 5 | 15.6 | 1 | 3.1 | 0 | 0 | 0 | 0 | 11 | 36.6 | 5 | 16.1 | 0 | 0 | 0 | 0 | 0 | 0 | 0 | 0 | 0 | 0 | 0 | 0 | 0 | 0 | 0 | 0 | 0 | 0 | 0 | 0 | 0 | 0 | 0 | 0 | 0 | 0 | 0 | 0 | 8 | 25.8 | 0 | 0 | 2 | 6.8 |
| mean | 31.53 | 2.75 | 8.69 | 0.57 | 1.79 | 0.15 | 0.47 | 0.82 | 2.57 | 0.03 | 0.11 | 6.37 | 20.05 | 1.27 | 3.97 | 0.22 | 0.71 | 0.03 | 0.11 | 13.08 | 41.39 | 6.7 | 21.21 | 0.05 | 0.16 | 2.3 | 7.22 | 0.02 | 0.05 | 0.03 | 0.1 | 0.07 | 0.21 | 0.02 | 0.07 | 0.05 | 0.16 | 1.12 | 3.57 | 0.1 | 0.31 | 0.15 | 0.48 | 0.03 | 0.1 | 0.37 | 1.16 | 0.22 | 0.7 | 0.03 | 0.1 | 8.48 | 26.82 | 0.02 | 0.05 | 4.13 | 13.01 |
| sd | 1.14 | 3 | 9.44 | 1.39 | 4.38 | 0.48 | 1.49 | 1.56 | 4.87 | 0.18 | 0.57 | 6.35 | 19.89 | 1.94 | 6.07 | 0.49 | 1.65 | 0.18 | 0.57 | 9.33 | 29.2 | 5.56 | 17.43 | 0.22 | 0.72 | 4.84 | 15.15 | 0.13 | 0.4 | 0.18 | 0.56 | 0.31 | 0.97 | 0.13 | 0.52 | 0.22 | 0.69 | 2.5 | 8.06 | 0.35 | 1.1 | 0.66 | 2.12 | 0.18 | 0.56 | 0.84 | 2.66 | 0.64 | 2.05 | 0.18 | 0.56 | 6.4 | 20.03 | 0.13 | 0.4 | 4.76 | 14.88 |

**Table I5: number of SAEs by symptom in clusters with one at least one SAE occurrence during the intervention (clusters highlighted in red are SR clusters)**

| Cluster | # of unique subjects with at least one SAE | Abdominal Pain | Convulsions | Cough | Diarrhea | Difficulty Breathing | Fever | Loss of appetite | Respiratory distress | Severe Malaria - Other | Severe Malaria - Prostration / Very weak, unable to stand | Tired | Unconsciousness / Drowsiness | Vomiting | Other | category |
|---|---|---|---|---|---|---|---|---|---|---|---|---|---|---|---|---|
| 2 | 1 | 0 | 0 | 0 | 0 | 0 | 2 | 1 | 0 | 0 | 0 | 0 | 0 | 0 | 3 | Hospitalization, Life threatening |
| 4 | 1 | 0 | 0 | 0 | 0 | 0 | 0 | 0 | 0 | 0 | 0 | 0 | 0 | 0 | 2 | Life threatening |
| 15 | 1 | 0 | 0 | 0 | 2 | 0 | 1 | 0 | 0 | 2 | 2 | 0 | 2 | 2 | 0 | Hospitalization, Death |
| 17 | 1 | 0 | 0 | 0 | 0 | 0 | 1 | 0 | 1 | 0 | 0 | 0 | 0 | 0 | 1 | Death |
| 24 | 1 | 1 | 0 | 0 | 0 | 0 | 0 | 0 | 0 | 0 | 0 | 0 | 0 | 0 | 2 | Death |
| 29 | 1 | 0 | 0 | 0 | 0 | 1 | 0 | 0 | 1 | 0 | 0 | 0 | 0 | 0 | 0 | Death |
| 34 | 1 | 0 | 0 | 1 | 0 | 0 | 0 | 0 | 1 | 0 | 0 | 1 | 0 | 0 | 0 | Death |
| 43 | 1 | 0 | 0 | 0 | 1 | 0 | 0 | 0 | 0 | 0 | 0 | 0 | 0 | 1 | 2 | Death |
| 47 | 1 | 0 | 1 | 0 | 0 | 0 | 1 | 0 | 0 | 0 | 0 | 0 | 0 | 0 | 0 | Death |
| 49 | 2 | 0 | 0 | 0 | 0 | 0 | 2 | 0 | 1 | 0 | 0 | 0 | 0 | 0 | 1 | Death |
| 50 | 1 | 0 | 0 | 0 | 0 | 0 | 1 | 0 | 0 | 0 | 0 | 0 | 0 | 1 | 0 | Death |
| 51 | 1 | 0 | 0 | 0 | 0 | 0 | 0 | 0 | 0 | 0 | 0 | 0 | 0 | 0 | 3 | Event required an intervention in order to prevent a permanent incapacity. |

**Table I6: number of SAEs by symptom in the SR clusters with one at least one SAE occurrence during the intervention**

| Cluster | # of unique subjects with at least one SAE | Abdominal Pain | Convulsions | Diarrhea | Difficulty Breathing | Fever | Respiratory distress | Severe Malaria - Other | Severe Malaria - Prostration / Very weak, unable to stand | Unconsciousness / Drowsiness | Vomiting | Other | category |
|---|---|---|---|---|---|---|---|---|---|---|---|---|---|
| 4 | 1 | 0 | 0 | 0 | 0 | 0 | 0 | 0 | 0 | 0 | 0 | 2 | Life threatening |
| 15 | 1 | 0 | 0 | 2 | 0 | 1 | 0 | 2 | 2 | 2 | 2 | 0 | Hospitalization, Death |
| 24 | 1 | 1 | 0 | 0 | 0 | 0 | 0 | 0 | 0 | 0 | 0 | 2 | Death |
| 29 | 1 | 0 | 0 | 0 | 1 | 0 | 1 | 0 | 0 | 0 | 0 | 0 | Death |
| 43 | 1 | 0 | 0 | 1 | 0 | 0 | 0 | 0 | 0 | 0 | 1 | 2 | Death |
| 47 | 1 | 0 | 1 | 0 | 0 | 1 | 0 | 0 | 0 | 0 | 0 | 0 | Death |
| 49 | 2 | 0 | 0 | 0 | 0 | 2 | 1 | 0 | 0 | 0 | 0 | 1 | Death |
| 51 | 1 | 0 | 0 | 0 | 0 | 0 | 0 | 0 | 0 | 0 | 0 | 3 | Event required an intervention in order to prevent a permanent incapacity. |

**Table I7: number of SAEs by symptom in the placebo clusters with one at least one SAE occurrence during the intervention**

| Cluster | # of unique subjects with at least one SAE | Cough | Fever | Loss of appetite | Respiratory distress | Tired | Vomiting | Other | category |
|---|---|---|---|---|---|---|---|---|---|
| 2 | 1 | 0 | 2 | 1 | 0 | 0 | 0 | 3 | Hospitalization, Life threatening |
| 17 | 1 | 0 | 1 | 0 | 1 | 0 | 0 | 1 | Death |
| 34 | 1 | 1 | 0 | 0 | 1 | 1 | 0 | 0 | Death |
| 50 | 1 | 0 | 1 | 0 | 0 | 0 | 1 | 0 | Death |

**Table I8: frequency and percentage of subjects in clusters with at least one SAE occurrence by symptom during the intervention (clusters highlighted in red are SR clusters)**

| Cluster | Total # of subjects | Abdominal Pain | | Convulsions | | Cough | | Diarrhea | | Difficulty Breathing | | Fever | | Loss of appetite | | Respiratory distress | | Severe Malaria - Other | | Severe Malaria - Prostration / Very weak, unable to stand | | Tired | | Unconsciousness / Drowsiness | | Vomiting | | Other | |
|---|---|---|---|---|---|---|---|---|---|---|---|---|---|---|---|---|---|---|---|---|---|---|---|---|---|---|---|---|---|
| | | # | % | # | % | # | % | # | % | # | % | # | % | # | % | # | % | # | % | # | % | # | % | # | % | # | % | # | % |
| 2 | 32 | 0 | 0 | 0 | 0 | 0 | 0 | 0 | 0 | 0 | 0 | 1 | 3.1 | 1 | 3.1 | 0 | 0 | 0 | 0 | 0 | 0 | 0 | 0 | 0 | 0 | 0 | 0 | 1 | 3.1 |
| 4 | 32 | 0 | 0 | 0 | 0 | 0 | 0 | 0 | 0 | 0 | 0 | 0 | 0 | 0 | 0 | 0 | 0 | 0 | 0 | 0 | 0 | 0 | 0 | 0 | 0 | 0 | 0 | 1 | 3.1 |
| 15 | 32 | 0 | 0 | 0 | 0 | 0 | 0 | 1 | 3.1 | 0 | 0 | 1 | 3.1 | 0 | 0 | 0 | 0 | 1 | 3.1 | 1 | 3.1 | 0 | 0 | 1 | 3.1 | 1 | 3.1 | 0 | 0 |
| 17 | 31 | 0 | 0 | 0 | 0 | 0 | 0 | 0 | 0 | 0 | 0 | 1 | 3.2 | 0 | 0 | 1 | 3.2 | 0 | 0 | 0 | 0 | 0 | 0 | 0 | 0 | 0 | 0 | 1 | 3.2 |
| 24 | 28 | 1 | 3.6 | 0 | 0 | 0 | 0 | 0 | 0 | 0 | 0 | 0 | 0 | 0 | 0 | 0 | 0 | 0 | 0 | 0 | 0 | 0 | 0 | 0 | 0 | 0 | 0 | 1 | 3.6 |
| 29 | 31 | 0 | 0 | 0 | 0 | 0 | 0 | 0 | 0 | 1 | 3.2 | 0 | 0 | 0 | 0 | 1 | 3.2 | 0 | 0 | 0 | 0 | 0 | 0 | 0 | 0 | 0 | 0 | 0 | 0 |
| 34 | 32 | 0 | 0 | 0 | 0 | 1 | 3.1 | 0 | 0 | 0 | 0 | 0 | 0 | 0 | 0 | 1 | 3.1 | 0 | 0 | 0 | 0 | 1 | 3.1 | 0 | 0 | 0 | 0 | 0 | 0 |
| 43 | 29 | 0 | 0 | 0 | 0 | 0 | 0 | 1 | 3.4 | 0 | 0 | 0 | 0 | 0 | 0 | 0 | 0 | 0 | 0 | 0 | 0 | 0 | 0 | 0 | 0 | 1 | 3.4 | 1 | 3.4 |
| 47 | 32 | 0 | 0 | 1 | 3.1 | 0 | 0 | 0 | 0 | 0 | 0 | 1 | 3.1 | 0 | 0 | 0 | 0 | 0 | 0 | 0 | 0 | 0 | 0 | 0 | 0 | 0 | 0 | 0 | 0 |
| 49 | 32 | 0 | 0 | 0 | 0 | 0 | 0 | 0 | 0 | 0 | 0 | 2 | 6.2 | 0 | 0 | 1 | 3.1 | 0 | 0 | 0 | 0 | 0 | 0 | 0 | 0 | 0 | 0 | 1 | 3.1 |
| 50 | 31 | 0 | 0 | 0 | 0 | 0 | 0 | 0 | 0 | 0 | 0 | 1 | 3.2 | 0 | 0 | 0 | 0 | 0 | 0 | 0 | 0 | 0 | 0 | 0 | 0 | 1 | 3.2 | 0 | 0 |
| 51 | 32 | 0 | 0 | 0 | 0 | 0 | 0 | 0 | 0 | 0 | 0 | 0 | 0 | 0 | 0 | 0 | 0 | 0 | 0 | 0 | 0 | 0 | 0 | 0 | 0 | 0 | 0 | 1 | 3.1 |

**Table 2: AE/SAE listing for the interim period between the baseline and intervention periods (1/15/2022~ 3/20/2022)**

| Treatment | Symptom | Frequency | total |
|---|---|---|---|
| Placebo | Chills | 2 | 98 |
| | Cough | 14 | |
| | Diarrhea | 4 | |
| | Fever | 41 | |
| | Headache | 6 | |
| | Loss of appetite | 6 | |
| | Other | 5 | |
| | Severe Malaria - Other | 1 | |
| | Skin Irritation / Rash | 2 | |
| | Vomiting | 17 | |
| | **Death (SAE)** | 1 | 1 |
| SR | Abdominal Pain | 2 | 78 |
| | Chills | 3 | |
| | Cough | 11 | |
| | Diarrhea | 5 | |
| | Fever | 27 | |
| | Headache | 5 | |
| | Loss of appetite | 3 | |
| | Other | 4 | |
| | Vomiting | 18 | |
| | **Hospitalization (SAE)** | 1 | 1 |

Note: There were 100 unique subjects involved in AEs/SAEs altogether, with 98 experiencing AEs, 1 SAE, and 1 both.
